# GWAS defines pathogenic signaling pathways and prioritizes drug targets for IgA nephropathy

**DOI:** 10.1101/2021.11.19.21265383

**Authors:** Krzysztof Kiryluk, Elena Sanchez-Rodriguez, Xu-jie Zhou, Francesca Zanoni, Lili Liu, Nikol Mladkova, Atlas Khan, Maddalena Marasa, Jun Y. Zhang, Olivia Balderes, Simone Sanna-Cherchi, Andrew S. Bomback, Pietro A. Canetta, Gerald B. Appel, Jai Radhakrishnan, Hernan Trimarchi, Ben Sprangers, Daniel C. Cattran, Heather Reich, York Pei, Pietro Ravani, Kresimir Galesic, Dita Maixnerova, Vladimir Tesar, Benedicte Stengel, Marie Metzger, Guillaume Canaud, Nicolas Maillard, Francois Berthoux, Laureline Berthelot, Evangeline Pillebout, Renato Monteiro, Raoul Nelson, Robert Wyatt, William Smoyer, John Mahan, Al-Akash Samhar, Guillermo Hidalgo, Alejandro Quiroga, Patricia Weng, Raji Sreedharan, David Selewski, Keefe Davis, Mahmoud Kallash, Tetyana L. Vasylyeva, Michelle Rheault, Aftab Chishti, Daniel Ranch, Scott E. Wenderfer, Dmitry Samsonov, Donna J. Claes, Akchurin Oleh, Dimitrios Goumenos, Maria Stangou, Judit Nagy, Tibor Kovacs, Enrico Fiaccadori, Antonio Amoroso, Cristina Barlassina, Daniele Cusi, Lucia Del Vecchio, Giovanni Giorgio Battaglia, Monica Bodria, Emanuela Boer, Luisa Bono, Giuliano Boscutti, Gianluca Caridi, Francesca Lugani, GianMarco Ghiggeri, Rosanna Coppo, Licia Peruzzi, Vittoria Esposito, Ciro Esposito, Sandro Feriozzi, Rosaria Polci, Giovanni Frasca, Marco Galliani, Maurizio Garozzo, Adele Mitrotti, Loreto Gesualdo, Simona Granata, Gianluigi Zaza, Francesco Londrino, Riccardo Magistroni, Isabella Pisani, Andrea Magnano, Carmelita Marcantoni, Piergiorgio Messa, Renzo Mignani, Antonello Pani, Claudio Ponticelli, Dario Roccatello, Maurizio Salvadori, Erica Salvi, Domenico Santoro, Guido Gembillo, Silvana Savoldi, Donatella Spotti, Pasquale Zamboli, Claudia Izzi, Federico Alberici, Elisa Delbarba, Michał Florczak, Natalia Krata, Krzysztof Mucha, Leszek Pączek, Stanisław Niemczyk, Barbara Moszczuk, Malgorzata Pańczyk-Tomaszewska, Malgorzata Mizerska-Wasiak, Agnieszka Perkowska-Ptasińska, Teresa Bączkowska, Magdalena Durlik, Krzysztof Pawlaczyk, Przemyslaw Sikora, Marcin Zaniew, Dorota Kaminska, Magdalena Krajewska, Izabella Kuzmiuk-Glembin, Zbigniew Heleniak, Barbara Bullo-Piontecka, Tomasz Liberek, Alicja Dębska-Slizien, Tomasz Hryszko, Anna Materna-Kiryluk, Monika Miklaszewska, Maria Szczepańska, Katarzyna Dyga, Edyta Machura, Katarzyna Siniewicz-Luzeńczyk, Monika Pawlak-Bratkowska, Marcin Tkaczyk, Dariusz Runowski, Norbert Kwella, Dorota Drożdż, Ireneusz Habura, Florian Kronenberg, Larisa Prikhodina, David van Heel, Bertrand Fontaine, Chris Cotsapas, Cisca Wijmenga, Andre Franke, Vito Annese, Peter K. Gregersen, Sreeja Parameswaran, Matthew Weirauch, Leah Kottyan, John B Harley, Hitoshi Suzuki, Ichiei Narita, Shin Goto, Hajeong Lee, Dong Ki Kim, Yon Su Kim, Jin-Ho Park, BeLong Cho, Murim Choi, Ans Van Wijk, Ana Huerta, Elisabet Ars, Jose Ballarin, Sigrid Lundberg, Bruno Vogt, Laila-Yasmin Mani, Yasar Caliskan, Jonathan Barratt, Thilini Abeygunaratne, Philip A. Kalra, Daniel P. Gale, Ulf Panzer, Thomas Rauen, Jürgen Floege, Pascal Schlosser, Arif B. Ekici, Kai-Uwe Eckardt, Nan Chen, Jingyuan Xie, Richard P. Lifton, Ruth J. F. Loos, Eimear E. Kenny, Iuliana Ionita-Laza, Anna Köttgen, Bruce Julian, Jan Novak, Francesco Scolari, Hong Zhang, Ali G. Gharavi

## Abstract

IgA nephropathy (IgAN) is a progressive form of kidney disease defined by glomerular deposition of IgA. We performed a genome-wide association study involving 10,146 kidney biopsy-diagnosed IgAN cases and 28,751 matched controls across 17 international cohorts. We defined 30 independent genome-wide significant risk loci jointly explaining 11% of disease risk. A total of 16 loci were novel, including *TNFSF4, REL, CD28, CXCL8/PF4V1, LY86, LYN, ANXA3, TNFSF8/15, REEP3, ZMIZ1, RELA, ETS1, IGH, IRF8, TNFRSF13B* and *FCAR*. The SNP-based heritability of IgAN was estimated at 23%. We observed a positive genetic correlation between IgAN and total serum IgA levels, allergy, tonsillectomy, and several infections, and a negative correlation with inflammatory bowel disease. All significant non-HLA loci shared with serum IgA levels had a concordant effect on the risk of IgAN. Moreover, IgAN loci were globally enriched in gene orthologs causing abnormal IgA levels when genetically manipulated in mice. The explained heritability was enriched in the regulatory elements of cells from the immune and hematopoietic systems and intestinal mucosa, providing support for the pathogenic role of extra-renal tissues. The polygenic risk of IgAN was associated with early disease onset, increased lifetime risk of kidney failure, as well as hematuria and several other traits in a phenome-wide association study of 590,515 individuals. In the comprehensive functional annotation analysis of candidate causal genes across genome-wide significant loci, we observed the convergence of biological candidates on a common set of inflammatory signaling pathways and cytokine ligand-receptor pairs, prioritizing potential new drug targets.

## INTRODUCTION

IgA nephropathy (IgAN) is a common form of immune-mediated glomerulonephritis characterized by glomerular deposition of IgA-containing immune complexes and manifesting with hematuria, proteinuria, and often kidney failure. Examination of kidney-biopsy tissue and demonstration of glomerular IgA deposits is required to establish the diagnosis. No approved targeted therapies presently exist for IgAN, and a large fraction of cases progress to kidney failure requiring kidney transplantation or dialysis. There are no validated molecular predictors of progression to kidney failure.

As the diagnosis requires a kidney biopsy, genetic discoveries have been hindered by small sample sizes of the existing genetic cohorts. Approximately 15 independent loci have been previously identified in association with IgAN, implicating defects in the complement pathway, intestinal network of IgA production, and innate immunity against mucosal pathogens^1–7^. These findings have already led to the reformulation of the existing IgAN pathogenesis model, with most candidate mechanisms mapping to the immune system rather than the kidney^8, 9^. Nevertheless, prior GWAS had a two-stage design, with sample size of the discovery stage often limiting the power to discover new loci.

Herein, we report a well-powered GWAS discovery study for IgAN involving 38,897 individuals (10,146 kidney biopsy-defined cases and 28,751 controls) recruited across 17 international cohorts. With the discovery of 16 novel non-HLA GWAS loci, we provide strong support for a highly polygenic architecture of IgAN. We assess functional consequences of the risk alleles, define causal cell types and signaling pathways, and explore genetic correlations and pleiotropic associations of the risk loci. We demonstrate that IgAN polygenic score predicts kidney disease outcomes. Importantly, we report convergence of multiple risk loci on the set of common signaling pathways and ligand-receptor pairs involved in the regulation of IgA production, prioritizing plausible new molecular drug targets.

## RESULTS

### Study Design

We performed a standardized GWAS and meta-analysis of 17 independent international case-control cohorts (12 newly genotyped and five previously published GWAS cohorts) comprising a total of 38,897 individuals (10,146 biopsy-proven cases and 28,751 controls). Of the 17 cohorts, 11 were of European ancestry with participants recruited from nephrology centers in Italy, Poland, Germany, France, Belgium, Czech Republic, Hungary, Croatia, Turkey, Spain, Sweden, U.K., U.S., Canada, and Argentina, and six cohorts were of East-Asian ancestry with participants recruited from nephrology centers in China, Japan, and Korea. A total of 14 cohorts (8,139 cases and 17,713 controls) were genotyped with high-density SNP arrays, ancestrally matched using principal component-based methods, and imputed using whole genome sequence reference panels specific for each ethnicity, and three additional cohorts (2,007 cases and 11,038 controls) were genotyped with the Immunochip platform (**Supplementary Table 1**).

Detailed description of each cohort, genotyping platform, quality control analysis, and ancestry and imputation analyses are provided as **Supplementary Notes**. Our primary discovery involved the combined analysis of all 17 cohorts under a log-additive genetic model. Additional exploratory analyses were conducted to identify any potential ancestry-specific and sex-specific locus, including under alternative (dominant and recessive) genetic models.

### Genome-wide significant loci

The results of combined meta-analyses across all cohorts are summarized in **Figure 1, Tables 2 and Supplementary Tables 2-4**. We confirmed multiple independently genome-wide significant (p<5×10^−8^) signals in the HLA region, with an overall λ=1.048 and 1.042 before and after excluding the HLA region (**Extended Data Figure 1**). In addition, we detected 24 independently associated non-HLA loci at a genome-wide significance, including eight known non-HLA loci (*CFH, DEFA1/4, CARD9, ITGAM/ITGAX*, *TNFSF13*, *LIF/OSM, FCRL3, IRF4/DUSP22*), 16 novel loci (*TNFSF4, CD28, REL, PF4V1/CXCL1, LY86/RREB1, LYN, ANXA3, TNFSF8/15, REEP3, ZMIZ1/PPIF, OVOL1/RELA, ETS1, IGH, IRF8, TNFRSF13B* and *FCAR*, see **Extended Data Figure 2)**, in addition to 48 independent suggestive non-HLA signals at *P*<1×10^−5^ (**Supplementary Tables 2 and 3**). Ethnicity-specific meta-analyses revealed another independent genome-wide significant association that was evident only in the East-Asian cohorts (*CCR6* locus, **Extended Data Figure 3a**) and 11 additional suggestive signals in this ancestral group (**Supplementary Table 4**). One of the suggestive signals in the combined meta-analysis (*PADI3/PADI4* locus) reached genome-wide significance in the East-Asian meta-analysis under a recessive model (**Extended Data Figure 3b**). The European-only meta-analysis showed a total of 19 suggestive signals, but no additional new genome-wide significant loci (**Supplementary Table 4**). No gender-specific loci were found in a gender-stratified meta-analysis, or in the analysis of sex chromosomal markers.

**Figure 1.**
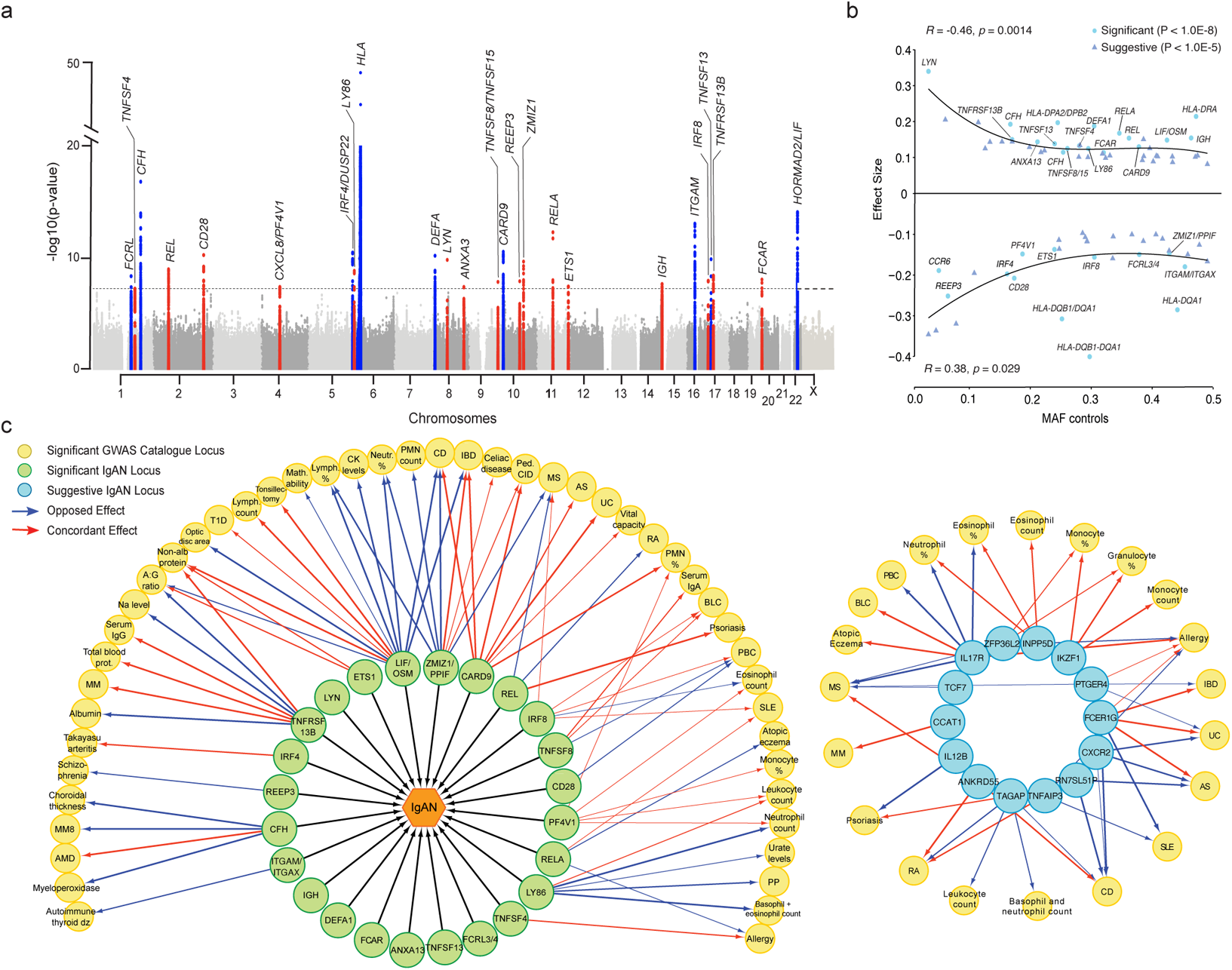
Transethnic GWAS for IgAN: **(a)** Manhattan plot for the combined meta-analysis across 38,897 individuals; the dotted horizontal line indicates a genome-wide significant p-value 5.0E-08; y-axis shows −log10 of p-values and is truncated to accommodate the HLA signal; x-axis shows genomic position for each chromosome (1-22 and X); red: novel genome-wide significant loci associated with IgAN; dark blue: previously known loci reaching genome-wide significance in this study; locus name based on the top candidate gene based on our biological prioritization strategy; **(b)** Effect size (beta, y-axis) as a function of minor allelic frequency (MAF, x-axis) for suggestive and significant GWAS loci; minor alleles with positive effect sizes (risk alleles) are represented at the top and negative effect sizes (protective alleles) are represented at the bottom; a 3-degree polynomial regression curve was fitted to illustrate positive and negative correlations; correlation coefficients (R) and their corresponding P-values (P) are also provided; light-blue circles represent genome-wide significant loci and are labeled using the most likely candidate gene per locus, blue triangles represent suggestive loci. **(c)** Pleiotropic effects of non-HLA GWAS loci for IgAN based on the NHGRI GWAS catalogue; only genome-wide significant associations in LD (r2 > 0.5) with IgAN top SNPs are included as edges; yellow are diseases and traits sharing at least one locus with IgAN; edge thickness is proportional to the LD between the IgAN top SNP and the lead association SNP for GWAS catalogue traits; concordant effects are indicated in red, opposed effects in blue; green nodes represent IgAN GWAS loci, light-blue nodes are IgAN suggestive loci; only 13 suggestive loci sharing at least one pleiotropic association with a genome-wide significant IgAN locus are depicted.

We next performed stepwise conditional analyses of the 24 genome-wide significant non-HLA loci, but only the *CFH* locus showed evidence of at least two independent genome-wide significant variants (**Supplementary Table 5 and Extended Data Figure 4a**). Stepwise conditional analyses of the *HLA* region revealed a complex pattern of association, with at least five independently genome-wide significant SNPs in the combined analysis of all cohorts (**Extended Data Figure 4b, Supplementary Table 6**). In addition, the patterns of association across the HLA region differed when stepwise conditioning was performed separately in East-Asian and European cohorts – a total of five independent signals were detected in the European cohorts, and four in the East-Asian cohorts. In the overall meta-analysis, we observed an inverse relationship between minor allelic frequency of the top independently associated variants and their effect sizes, consistent with the effects of purifying selection (**Figure 1b**).

Based on genome-wide summary statistics, we estimated the SNP-based heritability of IgAN at 0.23 (95%CI: 0.15-0.30). Excluding the MHC region reduced SNP-based heritability estimate to 0.12 (95%CI: 0.10-0.13), suggesting that HLA and non-HLA loci respectively contribute approximately 50% of the polygenic risk. Using the genetic risk score (expressed as the sum of the risk alleles weighted by their individual effect sizes) based on 30 independently genome-wide significant SNPs (30-SNP GRS) explained 11% of overall disease variance, a significant improvement compared to the 6% explained by the previous 15-SNP GRS^4^.

### Classical HLA alleles

To better understand the signal at the HLA locus, we imputed amino-acid sequences and classical HLA alleles at four-digit resolution at class II (*HLA-DQB1, -DQA1* and *-DRB1*) and class I (*HLA-A, -B*, and *-C*) genes (see **Methods**). We used ethnicity-specific reference panels for imputation, followed by association testing within each ancestral group separately. The analysis of imputed amino-acid sequences in class II genes pointed to *DRB1* as the gene with most strongly associated polymorphic positions in both ancestral groups (**Extended Data Figure 5**). In East Asian cohorts, stepwise conditioning of multiallelic sites demonstrated independently significant associations at DRβ1 positions 11 and 71, and the same positions were also strongly associated with the disease in Europeans. Specifically, Proline at position 11 (in LD with Arginine at position 71 and corresponding to DRB1*1501) conveyed significant protection in both ancestral groups (**Supplementary Table 7**). There was also an independently significant risk effect of Arginine at position 71 (in LD with Valine at position 11) with consistent direction of effect across both ancestries. In Europeans, we additionally observed significant effects of two substitutions at position 11, Glycine (protective) and Leucine (risk). These two substitutions are less frequent in East Asian populations.

The association patterns of classical HLA alleles were complex, but generally consistent with the analysis of amino acid sequences. In East Asians, we observed a protective effect of the *DRB1*1501*-*DQA1*0102-DQB1*0602* haplotype (DR15 serotype), and an independent risk effect of *DRB1*0405,* with no significant associations after conditioning for both *DRB1*1501* and *DRB1*0405* (**Supplementary Table 8**). In Europeans, we confirmed a strong protective association of the *DRB1*1501*-*DQA1*0102-DQB1*0602* haplotype. *DRB1*0405* had low allelic frequency in Europeans, thus this association was not replicated. Instead, we observed three additional independent European haplotypes (rare in East Asians), including two protective haplotypes, *DRB1*0301-DQA1*0501-DQB1*0201* (DR3 serotype) and *DRB1*0701-DQA1*0201-DQB1*0202/0203* (DR7 serotype), and one risk haplotype *DRB1*0101-DQA1*0101-DQB1*0501* (DR1 serotype, **Supplementary Table 9**). After conditioning for the four independently significant *DRB1* alleles residing on these haplotypes, we observed additional independent protective associations of *DQA1*0102* and *DPA1*0103* in Europeans. There were no independently significant associations for the class I genes. Collectively, these analyses point to MHC class II region, with *DRB1, DQA1*, and *DPA1* as the most likely candidate genes, but the extended LD across the region, combined with population differences in haplotype diversity and limitations related to imputation preclude further dissection of this complex signal.

### Pleiotropic associations of individual IgAN loci

To describe the full spectrum of pleiotropic associations of individual risk variants, we cross-annotated all non-HLA signals against all genome-wide association studies listed in the NHGRI GWAS Catalogue (**Supplementary Tables 10-11**). We identified both concordant and opposed associations for multiple autoimmune and inflammatory diseases, suggesting that these conditions may share pathogenic pathways with IgAN. Among the loci with the highest level of pleiotropy were *HORMAD2/LIF* and *ZMIZ1/PPIF*. Other loci with autoimmune pleiotropy were *CARD9, TNFSF8/15*, *REL, OVOL1/RELA*, *IRF4/DUSP22,* and *IRF8*. Additional novel loci, including *TNFRSF13B, PF4V1/CXCL1, LY86/RREB1,* and *ETS1*, showed concordant effects on blood levels of distinct immune cell types and immunoglobulins suggesting that these loci may act through stimulation of immune cell proliferation and immunoglobulin production. When we expanded this analysis to all suggestive loci, we found that 14 of the 47 suggestive loci were associated with the same autoimmune or blood immune cell traits as the genome-wide significant loci, prioritizing these 14 loci for future follow-up studies (**Figure 1c**).

### Shared genetic architecture with serum IgA levels and related traits

To interrogate shared susceptibility between IgAN and other common diseases, we explored genome-wide genetic correlations with selected immune, infectious, and cardio-metabolic traits using bivariate LD score regression (**Figure 2, Supplementary Table 12**)^10^. We found negative genetic correlations with primary sclerosing cholangitis (r_g_=-0.37, *P*=4.1×10^−3^), inflammatory bowel disease (r_g_=-0.16, *P*=9.9×10^−3^), and Crohn’s disease (r_g_=-0.17, *P*=1.0×10^−2^), and positive correlations with pneumonia (r_g_=0.26, *P*=9.0×10^−4^) and urinary tract infection (r_g_=0.25, *P*=2.1×10^−3^).^−^ After excluding HLA, we also observed a positive genetic correlation with serum IgA levels (r_g_=0.31, *P*=2.1×10^−3^), allergy (r_g_=0.18, *P*=5.2×10^−3^), and tonsillectomy (r_g_=0.17, *P*=0.036), a procedure performed for recurrent pharyngeal infections and also sometimes used to treat relapsing IgAN^11^. We next performed look-ups of all independent IgAN risk alleles against our latest GWAS for serum IgA levels (**Supplementary Table 13**). Of 25 non-HLA IgAN risk loci, 9 were nominally (P<0.05) associated with increased serum IgA levels, all with concordant effects. Conversely, of 31 significant loci for IgA levels 12 were nominally associated with the risk of IgAN, also with concordant effects. The intersection include four highly significant loci in both GWAS: *TNFSF13* (GWAS for IgA levels *P_IgA-level_*=9.4×10^−8^), *TNFSF8/15* (*P_IgA-level_*=3.2×10^−10^), *OVOL1/RELA* (*P_IgA-level_*=2.6×10^−22^), and *LIF/HORMAD2* (*P_IgA-level_*=6.7×10^−17^). At the same time, the allelic effects at the HLA locus were either opposed or not associated with serum IgA levels, consistent with our genetic correlation analyses in which positive genetic correlation with IgA levels is significant only after exclusion of the HLA region.

**Figure 2.**
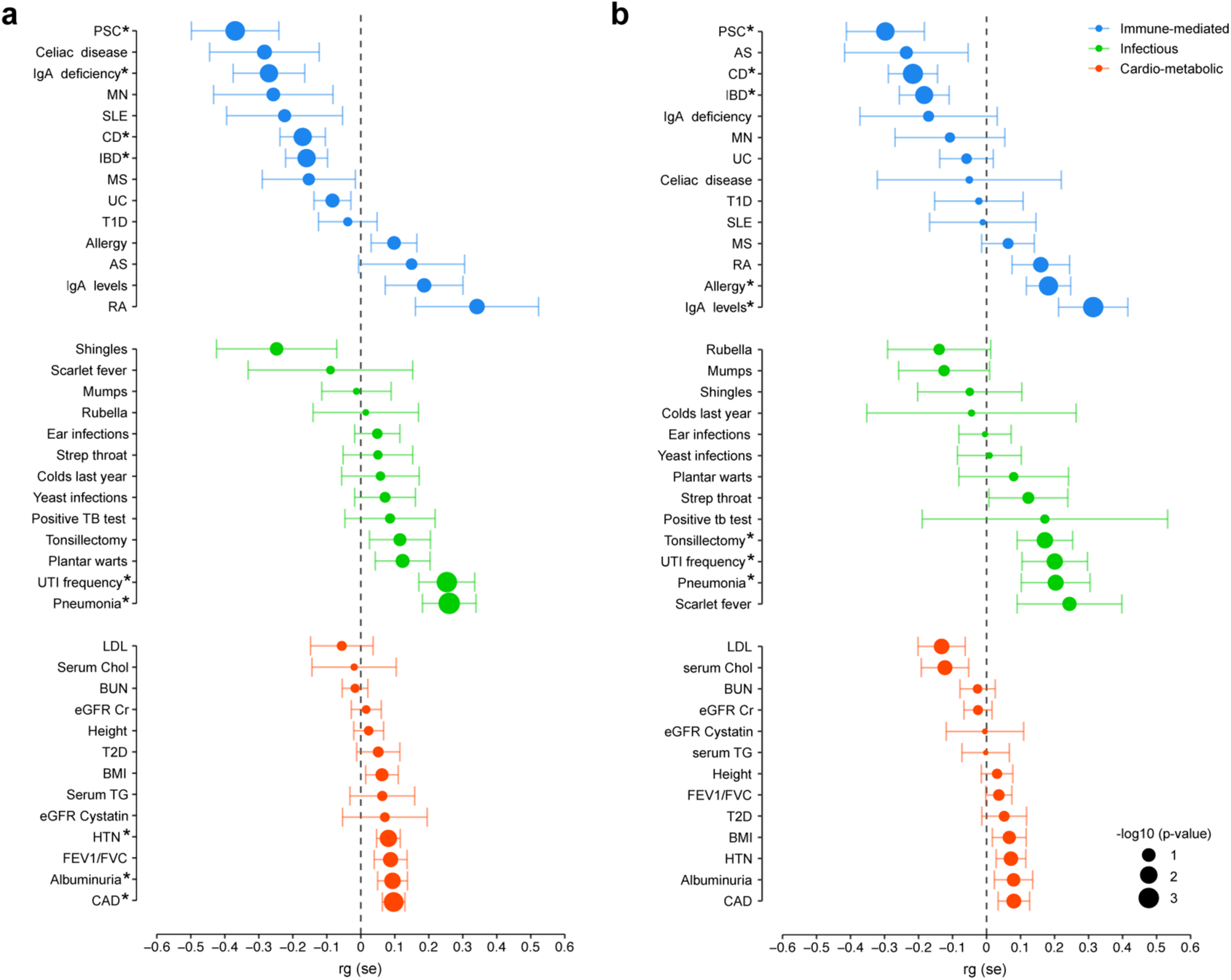
Genome-wide genetic correlation analysis between IgA nephropathy and other complex traits (a) including HLA region and (b) excluding HLA region. The traits are organized by immune-mediated (blue), infectious (green), and cardiometabolic (orange) categories and sorted based on the genetic correlation coefficient (rg); point estimates are provided with their 95% confidence intervals. PSC: primary sclerosing cholangitis, MN: membranous nephropathy, SLE: systemic lupus erythematosus, CD: Crohn’s disease, IBD: inflammatory bowel disease, MS: multiple sclerosis, UC: ulcerative colitis, T1D: type 1 diabetes, AS: ankylosing spondylitis, RA: rheumatoid arthritis, Chol: total serum cholesterol levels, TG: total serum triglycerides levels, LDL: low-density lipoprotein levels, BUN: blood urea nitrogen, eGFR Cr: estimated glomerular filtration rate using serum creatinine levels, T2D: type 2 diabetes, BMI: body mass index, eGFR Cystatin: estimated glomerular filtration rate using serum cystatin levels, HTN: essential hypertension, FEV1/FVC: forced expiration volume at 1 second over forced vital capacity, CAD: coronary artery disease. * phenotypic correlations at P<0.05.

### Mouse phenotypes support the role of dysregulated IgA production in IgAN

We tested the candidate gene set defined by our significant GWAS loci for overlap with human ortholog gene sets producing 27 phenotype categories when genetically manipulated in mice. We observed top-most significant enrichments in ‘*Immune system phenotype*’ (*P*=1.3×10^−12^) and ‘*Hematopoietic system phenotype*’ (*P*=3.2×10^−9^) (**Supplementary Table 14**). Within these categories, we observed significant enrichments in gene sets whose disruption in mice were associated with ‘*Abnormal IgA levels*’ (*P*=6.4×10^−6^) (**Extended Data Figure 6**), including *TNFSF13, TNFSF13B, ITGAM, RELA, REL, CD28,* and *LYN* genes. These observations corroborate our findings of overlapping GWAS loci between serum IgA levels and IgAN and further highlight the role of dysregulated IgA production in the disease pathogenesis. Moreover, this analysis strongly supports the named genes as causal at the corresponding loci and nominates appropriate animal models for experimental follow-up.

### Global pathway and tissue/cell type enrichment analyses

We next used several unbiased strategies to explore biological pathway and tissue enrichments using genome-wide approaches. Pathway-enrichment analysis using MAGMA^12^ revealed 24 enriched gene sets (**Extended Data Figure 7**). The most strongly enriched GO terms after excluding HLA region were ‘*Immune System Processes*’ (enrichment *P*=1.4×10^−9^) and ‘*Immune Response*’ (enrichment *P*=2.6×10^−9^). Examination of genome-wide significant non-HLA loci revealed significant enrichments in pathways involved in innate and adaptive immunity, with the most significant enrichment in the ‘*Cytokine-Cytokine Receptor Interactions*’ (enrichment *P*=4.0×10^−11^) (**Figure 3a**), suggesting several cytokine ligand-receptor interactions may drive disease pathogenesis.

**Figure 3.**
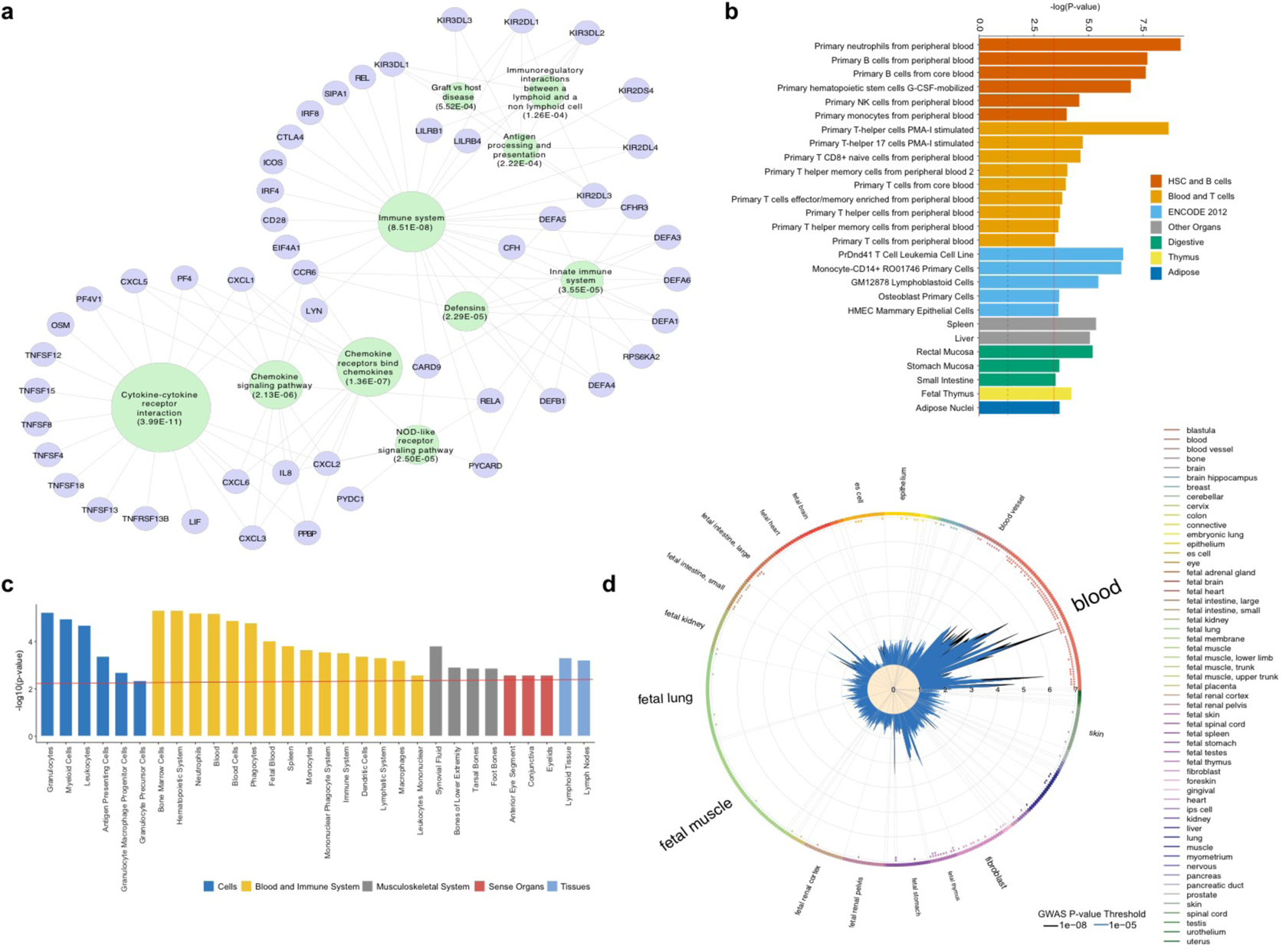
Global pathway, cell type, and tissue enrichment analyses: **(a)** KEGG, REACTOME and BIOCARTA pathway enrichment map based on the gene set defined by genome-wide significant IgA nephropathy loci after excluding HLA region; the top 10 most significantly enriched pathways and their intersecting GWAS genes are shown; a node size reflects −log10-transformed P-values of the multiple-testing adjusted hypergeometric enrichment test in GSEA. **(b)** Cell type-specific heritability enrichment for functional annotations based on FUN-LDA scoring system for all ENCODE and Roadmap Epigenomics cell types and tissues; the histogram shows the results of partitioned heritability by cell type-specific FUN-LDA functional annotations; only significant results grouped by the tissue type are depicted; a solid red line represents the Bonferroni-corrected −log10 of the p-value threshold of significance (*P*=3.9×10^−4^); a dotted black line represents the −log10 of the nominal p-value of 0.05; the most significant heritability enrichment were found in blood and immune cells and as well as gastrointestinal mucosal tissues. **(c)** Tissue and cell-type enrichment analysis with DEPICT; only cells and tissues with a false discovery rate < 0.05 are shown; Y-axis represents the −log10 of the p-value and x-axis shows the first level MeSH tissue and cell type annotations; the strongest enrichment is observed for blood and immune cells. **(d)** Global GWAS enrichment in DNase I–hypersensitive sites (DHS) using GARFIELD; radial lines show odds ratios at two GWAS p-values thresholds (T) for all DHS cells and tissues on the outer circle; dots in the inner ring of the outer circle denote significance of GARFIELD enrichment at *T*<1.0×10^−5^ (outermost) and *T*<1.0×10^−8^ (innermost) after multiple-testing correction for the number of effective annotations. The dots colored with respect to the tissue cell type tested (the font size of tissue labels reflects the number of cell types from that tissue, only tissues). Similar to FUN-LDA, GWAS results are most enriched in DHS sites in blood and immune cells, and intestinal mucosal tissue.

To map the most likely causal tissues and cell types, we partitioned SNP-based heritability across the genome by tissue and cell-type-specific functional scores derived using the FUN-LDA method^13^. We found the most significant heritability enrichments in blood, immune, and gastrointestinal mucosa cells (**Figure 3b, Supplementary Table 15**). The top enriched cell-types were *Primary neutrophils from peripheral blood* (*P*=5.9×10^−10^), *PMA-I-stimulated primary T helper cells* (*P*=2.1×10^−9^) and *Primary B cells from peripheral blood* (*P*=2.0×10^−8^). Analogous analysis performed using experimental mouse datasets pointed to *Small intestine inflammatory cells under basal conditions and after Salmonella infection* as the top tissue (**Extended Data Figure 7**). Additional independent analytical methods (DEPICT^14^ and GARFIELD^15^) similarly prioritized extra-renal tissues as likely causal in IgAN, converging on hematopoietic, immune, and gastrointestinal tissues as the most likely tissues to harbor causal cell types (**Figures 3c and 3d, Supplementary Tables 16-17**).

### Transcription factor enrichment analysis

We tested for potential intersection of GWAS signals with a comprehensive database of transcription factor (TF) ChIP-seq datasets using the Regulatory Element Locus Intersection (RELI) algorithm^16^. In the analysis of genome-wide significant and suggestive loci, we detected significant intersection with binding sites for up 32 TFs in 52 immune cell types, with the most significant enrichments for RELA (corrected P=5.3×10^−13^) and NFKB1 (corrected P=1.9×10^−12^, **Figure 4d, Supplementary Table 18**). Nearly half of these transcription factors interact with Epstein-Barr-virus super-enhancers, which control B cell proliferation and have previously been found to intersect multiple autoimmune disease loci^16, 17^. Moreover, some of the prioritized TFs, such RUNX^18^ and SMAD^19^ family, are well known to regulate IgA levels, and RUNX3, RUNX2, and RELA loci are significantly associated with IgA levels (*see accompanying manuscript*), further suggesting perturbations in IgA homeostasis as a primary pathogenetic factor IgAN.

**Figure 4.**
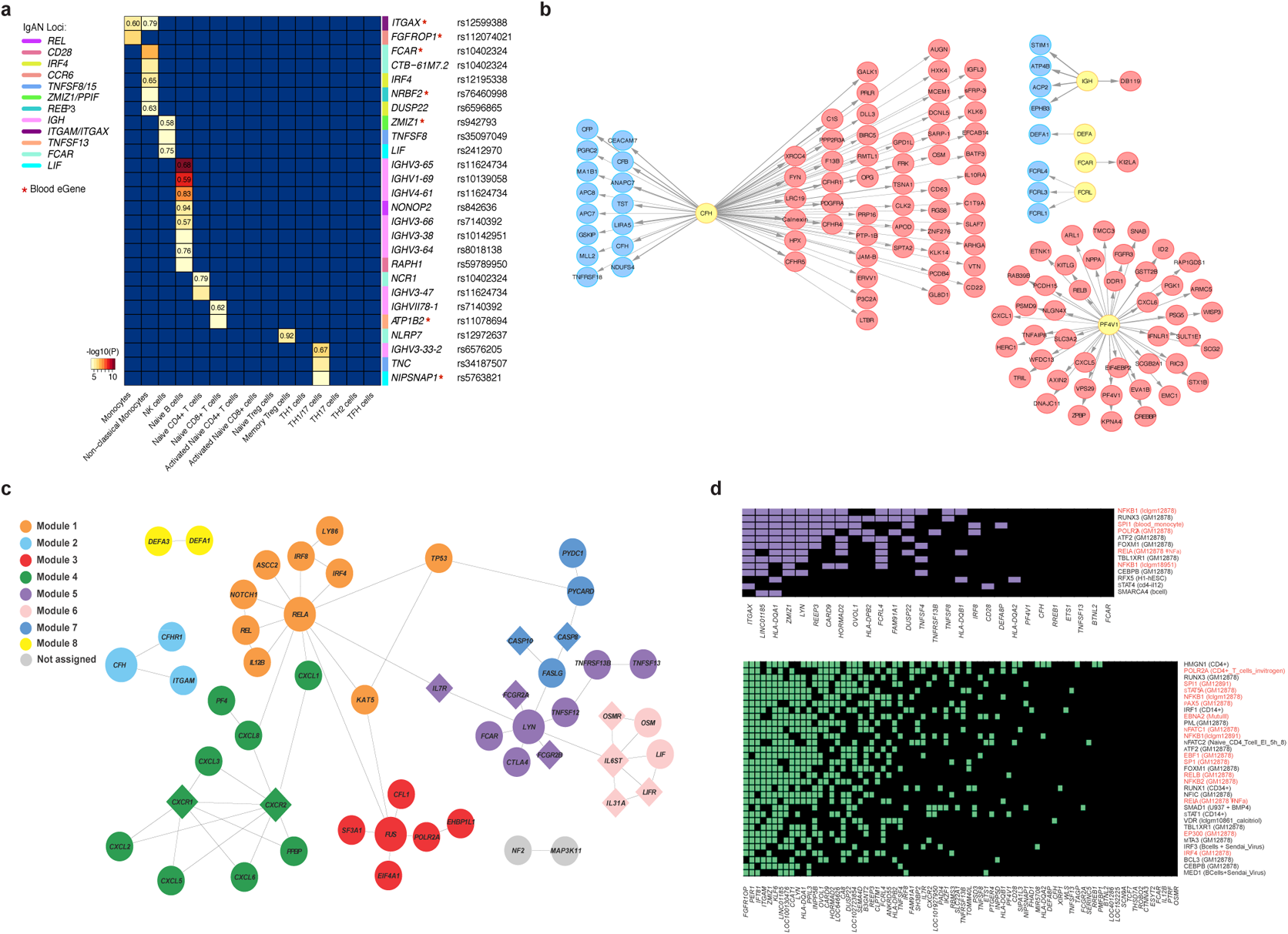
Summary of candidate causal gene prioritizations across IgAN GWAS loci excluding HLA region: **(a)** cis-eQTL effects in primary immune cells: x-axis: 13 immune cell types of the Database of Immune Cell expression QTL epigenomics (DICE) project; posterior probability for a shared causal variant (PP4) is shown for the e-QTL effects that co-localize at PP4>0.50; y-axis: significant eGene-eSNP pair with shared GWAS loci depicted by a color bar; *eGenes with detectable blood cis-eQTLs. **(b)** pQTL effects in blood: IgAN risk alleles or their proxies (yellow nodes) with significant blood pQTLs depicted by blue (reduced protein levels) and red (increased protein levels) nodes; edge thickness corresponds to the LD between the lead pQTL SNP and the top IgAN GWAS SNP at a given locus. **(c)** Protein-protein interaction (PPI) network for candidate genes at GWAS loci based on InWeb_IM: modules represent genes that are more connected to one another than they are to other genes; each module exhibits a functional enrichment network based on Gene Ontology (GO) biological processes: module 1 represents response to stress and defense response networks (orange), module 2 represents regulation of inflammatory response network (light blue), module 3 represents mRNA splicing via spliceosome network (red), module 4 represents chemokine-mediated signaling pathway network (green), module 5 represents immune response network (purple), module 6 represents cytokine-mediated signaling pathway network (pink), module 7 represents regulation of I-Kappa B kinase/NF-kappaB signaling and apoptotic signaling pathway networks (dark blue) and module 8 represents innate immune response in mucosa and antibacterial humoral response networks (yellow); the gray module has no functional enrichment. Overall, this network has more connectivity than expected by chance (permutation *P*<2e-03). **(d)** Intersection with transcription factor (TF) ChIP-seq peaks with the significant (top) and suggestive (bottom) IgAN risk loci; x-axis: IgAN risk loci; y-axis: top significant TFs ranked by the number of intersecting loci; a colored box at the intersection indicates that at a given locus has at least one IgAN-associated variant located within a ChiP-seq peak for the given TF; Datasets were considered significant if their RELI corrected p-values were less than 1E-04; TFs binding to EBNA2 super-enhancers colored in red; ChiP-seq dataset cell type indicated in parentheses; related cell lines for a given TF (e.g., GM12878 and GM12891) were merged for clarity.

### Protein-protein interactions and ligand-receptor pairs

We next tested whether candidate genes within our significant loci encode proteins that are likely to have physical interactions. Using a refined database of high-confidence PPIs, we constructed a network with 76 candidate proteins defined by GWAS using InWeb_IM^20^ and GeneMANIA^21^. The final network composed of a total of 53 nodes and 63 edges exhibited an excess of direct physical interactions compared to null expectation (*P*<1.0×10^−16^, **Figure 4c**). Gene set enrichment analyses of individual modules in this network (**Supplementary Table 19**) identified strong enrichments in stress and defense responses (module 1), chemokine signaling pathways (module 4), immune responses (module 5), cytokine-mediated signaling (module 6), and regulation of NF-κB signaling (module 7). Consistent with the observed enrichments in chemokine and cytokine pathways and global cytokine-receptor interactions, we identified enrichment in soluble ligand-receptor pairs, attributable to 16 ligand-receptor pairs spanning 12 independent significant or suggestive loci (enrichment *P*=0.01, **Supplementary Table 20**). This included APRIL and its receptor TACI encoded by two independent genome-wide significant loci (*TNFSF13* and *TNFRSF13B*, respectively), both implicated in IgA homeostasis. Several IL6-related cytokine-receptor pairs were also identified (IL6-IL6ST, LIF-LIFR/IL6ST, OSM-OSMR/LIFR/IL6ST), with *OSM/LIF* being encoded by a single genome-wide significant locus, and related receptors being encoded by two independent suggestive loci, *OSMR/LIFR* and *IL6ST*. Notably, APRIL is known to alter the glycosylation of IgA^22^, IL6, LIF and OSM are involved in mucosal immunity, and IL6 and LIF leads to enhanced production of galactose-deficient IgA1^23–25^. These ligand-receptor pairs nominate candidate genes within corresponding loci, and delineate potentially targetable pathogenetic pathways in IgAN.

### Functional annotations of individual GWAS loci

We intersected our association loci with tissue and cell-type specific functional scores and assessed their co-localization with expression quantitative trait loci (eQTL) in primary immune cells, whole blood, and other tissues (see **Methods**)^26^. We also performed cross-annotation with blood proteome and metabolome data. The majority of top signals mapped to non-coding regions, with the exception of two risk loci (rs4077515 *CARD9* p.(Ser12Ile), and rs3803800 *TNFSF13* p.(Asn96Ser)). The *CARD9* risk allele (rs4077515-T), a nonsynonymous S12N substitution in exon 2 of *CARD9*, is associated with increased blood transcript level of *CARD9* and a significant splice QTL in GTEx (**Extended Data Figure 8**). The protective allele is associated with a truncation of the functional CARD domain, while the risk allele is associated with higher levels of the intact, active isoform, affecting both expression and splicing of *CARD9*.

For top signals mapping to non-coding regions, we found 79 significant *cis*-eQTL effects with 17 IgAN co-localizations at 20 independent non-HLA risk loci (**Supplementary Tables 21-22 and** **Figure 4a**). Twelve loci had 27 significant *cis*-eQTL effects across 13 primary immune cell types, and 17 of the 27 *cis*-eQTLs co-localized with IgAN with PP4>0.5 (**Supplementary Table 21)**. In GTEx, we further found 19 *cis*-eQTL effects for eight IgAN loci across the 28 available tissues and cell types. As an example, two loci (*ITGAM/ITGAX* and *IRF4*/*DUSP22)* mapped specifically to monocytes, an understudied cell type in IgAN. The top signals at these loci intersect monocyte-specific functional elements by FUN-LDA, and co-localize with monocyte-specific eQTLs, with the risk alleles associated with up- and down regulation of *ITGAX* and *IRF4/DUSP22,* respectively. As another example of cell type specificity, the *ZMIZ1/PPIF* locus co-localized with eQTL in NK cells, with the risk allele associated with lower expression of *ZMIZ1*, which encodes an inhibitor of JAK/STAT signaling and is also involved in TGF-β signaling and intestinal inflammation^27, 28^. In whole blood, notable eQTL co-localizations included the *FCRL3* risk locus, where the risk allele was associated with reduced transcript levels of *FCRL3* and *FCRL5,* and with lower levels of circulating FCRL3 protein (**Supplementary Table 23**). As FCRL3 is a specific receptor for secretory IgA^29, 30^, we prioritized *FCRL3* is the most likely causal gene at this locus.

Three independent IgAN risk loci with colocalizing cis-eQTLs also exhibited trans-eQTL effects, suggesting that these loci induce a more global transcriptional perturbation in blood cells (**Supplementary Table 24**), For example, the *CARD9* locus was associated with 12 trans-eQTL effects, nine of which involve genes in the ‘*Type I interferon signaling pathway*’ (enrichment *P*=9.5×10^−18^). The *TNFSF8/15* locus was associated with eight trans-eQTL effects with three representing ‘*Cytokines involved in lymphocyte differentiation*’ (enrichment *P*=4.3×10^−3^). Interestingly, *ITGAX* locus had only one trans-eQTL association, lowering mRNA level of *IGHG4,* encoded by an independent IgAN risk locus on chr.14.

Finally, other loci were associated with perturbations in blood proteome or metabolome. The *PF4V1* locus colocalized with *PF4V1* cis-eQTL and exhibited multiple pQTL associations with blood protein levels (**Supplementary Table 23**), including 4 cis and 40 trans-pQTLs proteins. These proteins were most enriched in the GO process of ‘*Positive Regulation of Neutrophil Chemotaxis*’ (enrichment P=1.3×10^−3^), providing further evidence as *PF4V1* as the most likely causal gene for IgAN^31^. Similarly, the *CFH* locus, where a protective allele is known to tag a common deletion of the *CFHR1* and *CFHR3* genes^2^, was associated with reduced expression of *CFHR1* and *CFHR3* in the liver and other tissues including the kidney (**Supplementary Tables 25-26**). This allele was also associated with reduced levels of circulating FHR1 (encoded by *CFHR1*) and higher levels of Factor H in blood (**Supplementary Table 23**). Moreover, this locus exhibited a widespread proteomic and metabolomic signature in blood, with 64 additional trans-pQTL associations including seven proteins involved in the ‘*Regulation of complement cascade*’ (enrichment *P*=2.1×10^−10^, **Figure 4b**), and altered blood levels of multiple inflammation-related metabolites (**Supplementary Table 27**)^32, 33^.

### Integrative prioritization of biological candidate genes

To systematically prioritize the 311 candidate genes encoded within the 25 significant non-MHC risk loci, we scored for convergence of *in silico* annotation methods by assigning one point for each of the following criteria: 1) genes most proximal to the top SNP at the locus; 2) genes with a non-synonymous coding variant tagged (r2≥0.8) by the top SNP; 3) genes with a 3D chromatin interaction predicted by the Activity-by-Contact (ABC) model^34^ or 4) GeneHancer^35^, with enhancers that are intersected by variants tagged (r2≥0.8) by the top SNP or contained within a 95% credible set for the locus; 5) e-genes controlled by at least one eQTL (any GTEx tissue) tagged by the top SNP; 6) e-genes co-localizing with the risk locus in peripheral blood or 7) primary immune cells at PP4>0.5; 8) p-genes encoding blood proteins controlled by at least one cis-pQTL tagged by the top SNP; 9) genes prioritized by PPI network connectivity analysis at *P*<0.05; 10) genes with shared mouse knockout phenotypes; 11) genes within shared MAGMA pathways; 12) genes prioritized by DEPICT, and 13) genes prioritized by manual review of the literature as related to IgAN, IgA production, or mucosal immunity. Using this approach, we prioritized 27 ‘biological candidate genes’, 20 (74%) of which were also most proximal genes to the top SNP (**Figure 5**).

**Figure 5.**
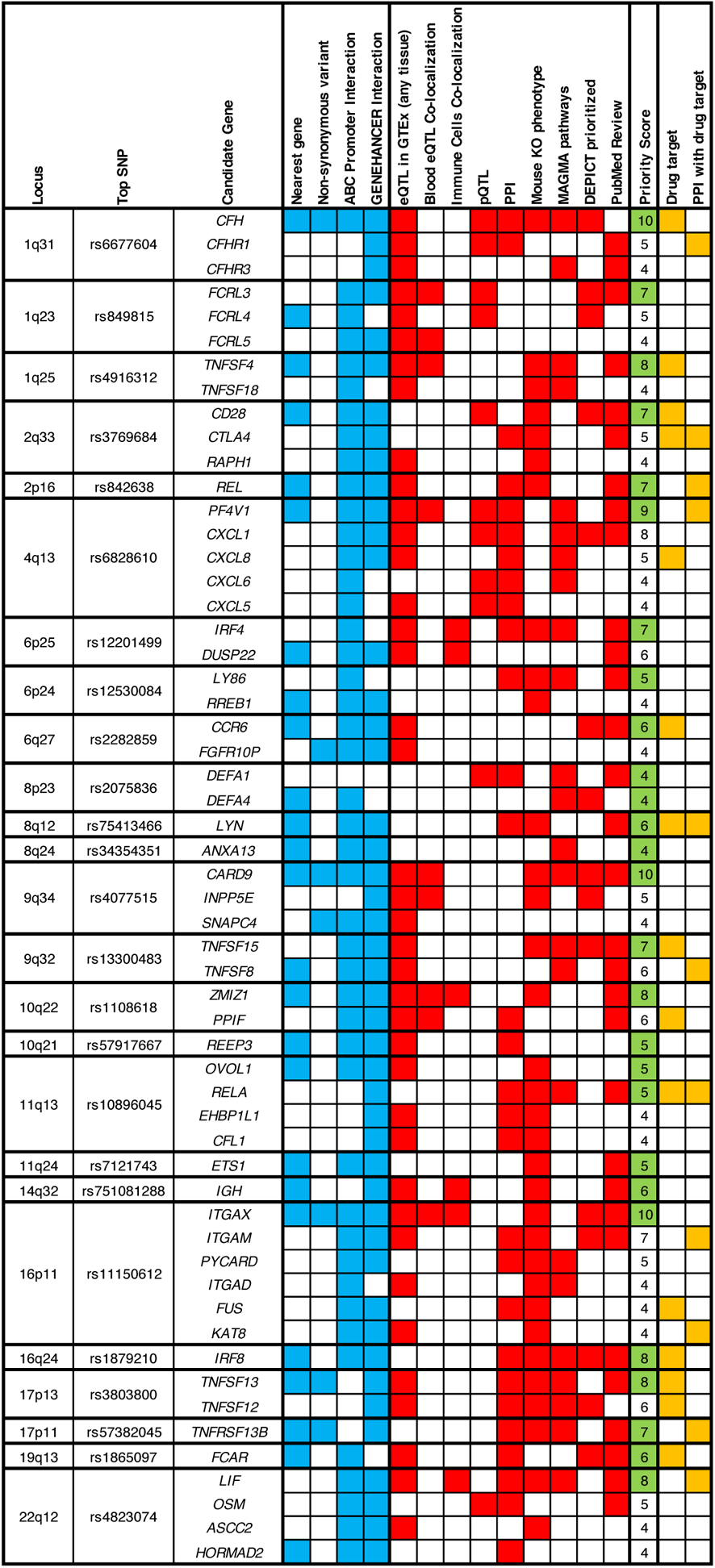
Prioritization of candidate genes at non-HLA loci: blue boxes indicate prioritization criteria based on genomic coordinates (the nearest gene to the index SNP, exonic variant in LD with the top SNP, or top signal intersecting chromatin interaction site with the gene promoter); red boxes indicate the presence of additional functional criteria (any GTEx eQTL effect, blood and immune cell eQTL co-localization, pQTL effects, PPI network connectivity, shared mouse KO phenotype, shared pathways by MAGMA, prioritized by DEPICT, and prioritized by manual PubMed review). The priority score represents a sum of the 13 scoring criteria depicted in blue and red. The genes with the maximum score at each locus (green boxes) were defined as ‘biological candidate genes’. Additional annotation indicates drug target genes (yellow boxes). Only 56 of 311 positional candidate genes with a score >3 are depicted.

### Prioritization of plausible drug targets

To facilitate drug repurposing and to prioritize new targets with GWAS support, we evaluated whether any of the 311 genes contained within significant loci encoded a protein or directly interacted with a protein that was a pharmacologically active drug target either approved or in development for any human disease. In total 13 GWAS loci (52%) encoded 17 proteins that were already targeted by existing drugs, and 11 loci (44%) encoded 14 proteins with a direct PPI drug target (**Supplementary Table 28 and** **Figure 6**). Among the top 27 high priority ‘biological candidates’ defined by our scoring system, 11 (40%) were targeted directly or indirectly by the existing drugs. This included the following drug categories: (1) new inhibitors of the alternative complement pathway, such as APL-2, AMY-101, and several others^36^ that are currently in clinical trials for C3 glomerulopathies and age-related macular degeneration; (2) drugs that block the activation of B cells by inhibiting APRIL or TACI interactions such as Atacicept and related drugs that are already in clinal trials for IgAN; (3) drugs that inhibit T cell activation by targeting ligands of the T-cell stimulatory CD28 protein, such as Belatacept (approved for kidney allograft rejection) or Abatacept (approved for rheumatoid arthritis); (4) drugs that inhibit IL8 (ABX-IL8) or IL8 receptor (Clotrimazole); and (5) drugs that inhibit NF-κB pathway, such as Bardoxolone that is already in clinical trials for glomerular disorders. We also note that some of our top prioritized causal genes with expression increased by the risk alleles, such as CARD9, ITGAX, PF4V1, CFHR1, or FCAR do not yet have effective drug inhibitors. Other loci encode secreted proteins that appear protective, such as FCRL3 and TNFSF4, suggesting that targeting their upregulation may present a rational therapeutic strategy. Our data additionally implies that activation of transcriptional programs controlled by ZMIZ1 and IRF4, but reduced activation of NF-κB, may also convey a protective effect.

**Figure 6:**
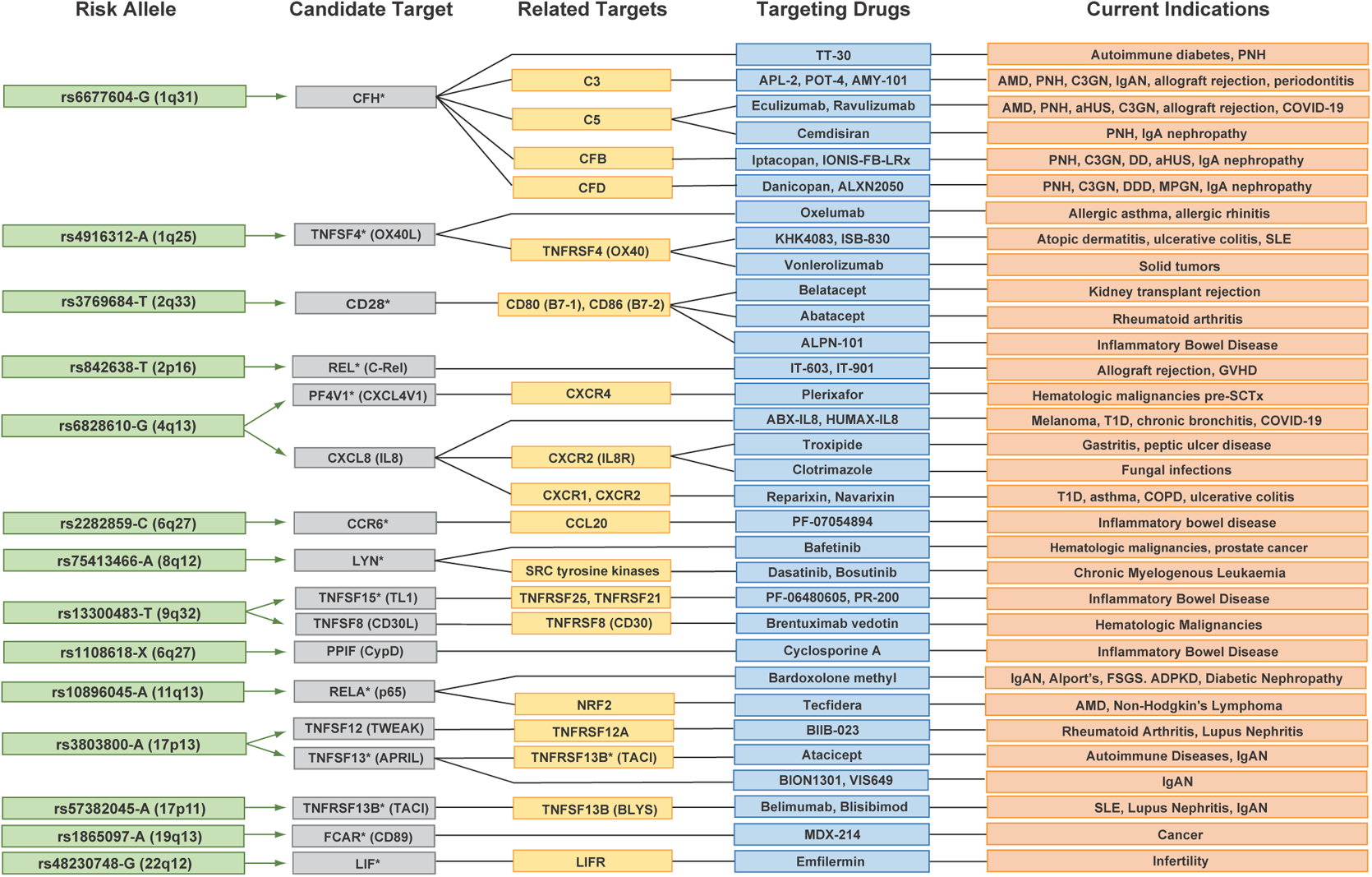
Drug targets among candidate causal genes. IgAN risk alleles (green), prioritized positional candidate genes (gray), related genes in PPI (e.g. ligands/receptors) or same pathway (yellow), targeting drugs approved or currently in clinical trials including agonists and antagonists (blue), and diseases targeted by these drugs (orange); high priority targets defined in Figure 5 are indicated by an asterisk; GWAS loci with candidate causal genes not targeted by the existing drugs are not depicted.

### Genome-wide polygenic risk and clinical correlations

Based on GWAS summary statistics after excluding Immunochip cohorts, we designed and optimized a genome-wide polygenic risk score (GPS) for IgAN. The best-performing GPS was based on LDPred method and assumed 1% causal variants genome-wide. When tested in the independent GCKD Study^37, 38^, the GPS explained approximately 7.3% of disease risk (*P*=3.1×10^−12^, C-statistic 0.65, 95%CI: 0.61-0.68). We hypothesized that higher polygenic risk captured by the GPS was associated with disease severity and faster progression among cases diagnosed with IgAN. To test this hypothesis, we performed a comprehensive analysis of clinical disease features in association with the GPS (**Supplementary Table 29**). Consistent with previous observations for the 15-SNP GRS^4^, the GPS was inversely associated with the age at diagnosis, with individuals in the top 20% tail of the GPS distribution diagnosed 2.2 years sooner 17 (95%CI: 1.3-3.1, *P*=2.5×10^−6^) compared to the rest of the cohort. The GPS was also significantly associated with faster progression to kidney failure among 2,879 IgAN cases with long-term follow-up data (HR=1.17 per standard deviation, 95%CI 1.09-1.24, *P*=3.3×10^−6^). For example, individuals in the top 20% tail of the GPS distribution had 34% increased risk of kidney failure (HR=1.34, 95%CI=1.15-1.56, *P*=2.0×10^−4^), while individuals in the top 10% tail had 48% increased risk of kidney failure (HR=1.48, 95%CI 1.22-1.79, *P=*6.6×10^−5^) compared to the rest of the cohort (**Figure 7a**).

**Figure 7.**
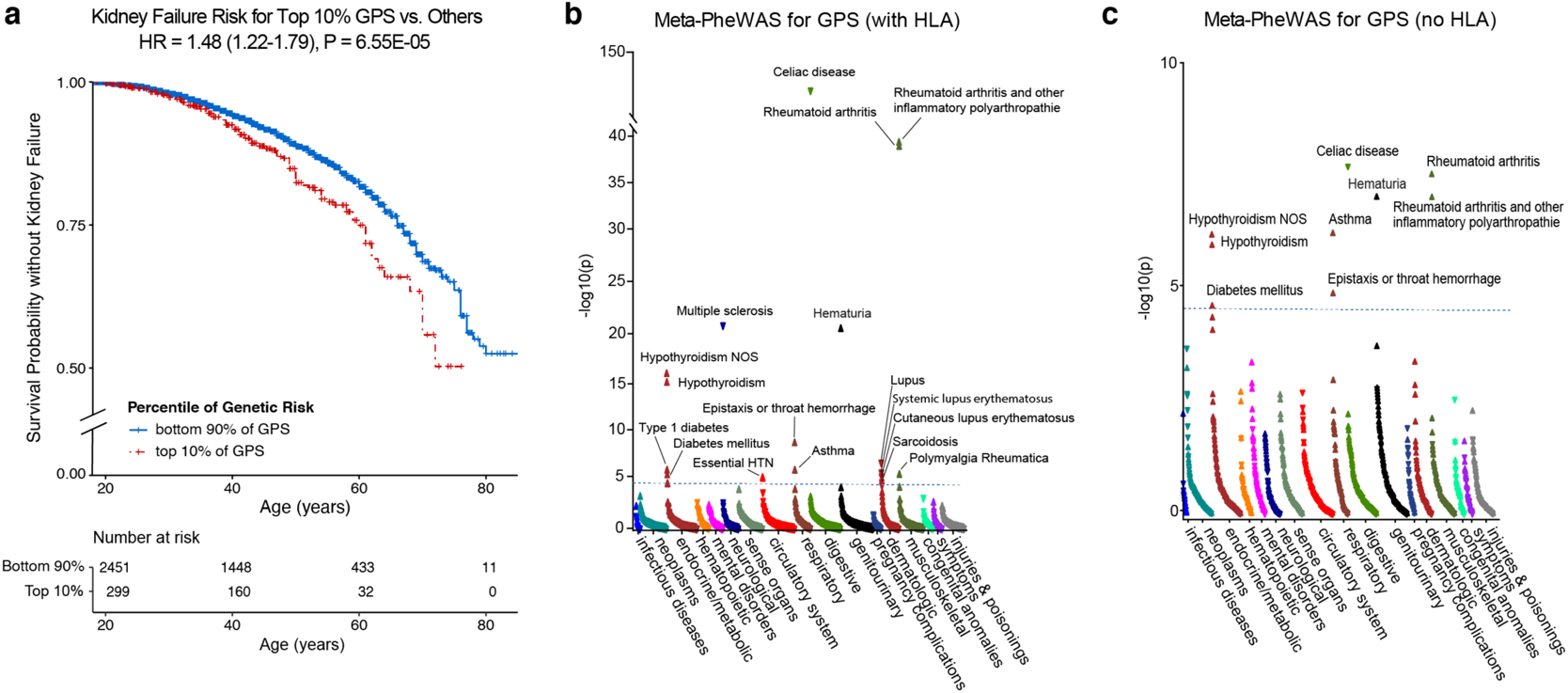
Clinical associations of the genome-wide polygenic risk score (GPS) for IgA nephropathy. **(a)** Survival analysis of a lifetime risk of kidney failure for IgAN cases in the top 90^th^ percentile of the GPS distribution (N=2,879 cases with follow up data). The x-axis shows age starting from 18 years, the y-axis shows survival probability without kidney failure with the number of participants at risk at each age cut-off of 20, 40, 60, and 80 years depicted below; HR: hazard ratio (95% confidence interval) of kidney failure adjusted for sex, site, and ancestry. **(b)** Phenome-wide association study (PheWAS) for the GPS, including **(c)** the GPS without the HLA region, based on joint meta-analysis of eMERGE-III (N=102,138) and UKBB (N=488,377) datasets. The x-axis indicates electronic health record phenotypes (phecodes) grouped by system and sorted by significance. The y-axis indicates the level of statistical significance expressed as −log(P-value). An upward triangle indicates a positive association (increased risk) and a downward triangle indicates a negative association (decreased risk) with increasing GPS. Dotted horizontal line represents the significance threshold after Bonferroni correction for the number of phenotypes; significant associations are labeled.

To explore additional clinical associations of the GPS, we performed meta-phenome-wide association study (meta-PheWAS) including a total of 590,515 participants with GWAS data liked to electronic health records, combining UK Biobank and Electronic Health Records and Genomics-III (eMERGE-III) consortium datasets (**Figure 7b**). We detected a significant positive correlation of the GPS with hematuria, the most common manifestation of IgAN (OR per standard deviation=1.06, P=7.3×10^−21^). Other notable associations included a protective association with celiac disease (OR_SD_=0.60, P=4.2×10^−148^) and several risk associations, including with rheumatoid arthritis (OR_SD_=1.15, P=1.1×10^−39^), hypothyroidism (OR_SD_=1.05, P=2.0×10^−15^), epistaxis or throat hemorrhage (OR_SD_=1.09, P=2.6×10^−9^), and asthma (OR_SD_=1.02, P=1.5×10^−6^). The above associations remained significant after removing the HLA region from the GPS (**Figure 7c, Supplemental Table 30**). Notably, the directions of effect were generally consistent with our genome-wide genetic correlation analyses of IgAN with related traits, providing an independent validation of the shared polygenic architecture for these traits.

## DISCUSSION

Our GWAS of 10,146 cases and 28,751 controls provided support for a highly polygenic architecture of IgAN with estimated SNP-based heritability of ~23%. We identified 16 novel susceptibility loci for IgAN, bringing the total number of known risk loci to over 30 and explaining ~11% of the variance in disease risk. The higher polygenic risk was associated with earlier disease onset and greater lifetime risk of kidney failure, suggesting that polygenic background is predictive of a more aggressive disease. Future studies are needed to test if our polygenic stratification is useful in the diagnosis, clinical risk assessment, or prediction of treatment responsiveness.

Our results reinforce the hypothesis that the genetic regulation of IgA production represents the key pathogenic pathway in IgAN. Significant risk loci were enriched in human orthologs of mouse genes that, when genetically modified, cause abnormal IgA levels. We also observed positive genetic correlation between IgAN and serum IgA levels. Moreover, 21 of 25 independent genome-wide significant non-HLA risk loci for IgAN appear to have concordant effect on serum IgA levels, and four of these 21 are also genome-wide significant in a GWAS for serum IgA levels, including loci encoding TNFSF8, APRIL, LIF, and RELA.

We observed positive genetic correlations with IgA levels, infections, and tonsillectomy indicating a genetic link between IgA system, common infections, and IgAN. The association with tonsillectomy is especially intriguing, because IgAN is often triggered by pharyngitis, and tonsillectomy has been employed as a treatment for IgAN^11^. In contrast, the observed negative genetic correlations with inflammatory bowel disease may be due to genetically increased production of secretory IgA that has known homeostatic anti-inflammatory and immunosuppressive effects at the level of the gut mucosa^39^. Moreover, our analyses of partitioned heritability clearly support extra-renal tissues as the most likely culprit, prioritizing cells of the immune and hematopoietic systems, and intestinal mucosal tissue, and this extra-renal mapping of causal tissues is fully consistent with the well-established clinical observation that IgAN commonly recurs in kidney allografts after transplantation^40^. In addition, our cell-type specific analyses point to the role of neutrophils, monocytes, and NK cells in IgAN. These cell types have not been generally considered as relevant to the IgAN pathogenesis based on prior evidence.

Our GWAS loci encoded proteins that were more likely to interact physically despite being encoded by distant genomic regions. This included several ligand-receptor pairs that are amenable to therapeutic targeting. IgAN currently lacks effective targeted therapies, and recent pharmaceutical database analyses indicate that drug targets with genetic support are more likely to advance in the development pipeline^41^. Based on our results, we prioritized several candidate genes whose products are targeted by drugs that are presently approved or in clinical development for another condition, and which could be repurposed for IgAN. Mechanistic studies are still needed to confirm our candidate target genes prioritized by our *in silico* annotations.

Our study has several limitations. In the meta-analysis, we pooled data across multiple heterogenous cohorts recruited across diverse clinical settings, ancestries, and nationalities. Nevertheless, we used stringent biopsy-based diagnostic criteria, standardized covariate definitions, genetic matching by platform and ancestry, and uniform statistical analysis for each cohort. It is worth pointing out that our dataset is dominated by European (66%) and East Asian (34%) ancestry cases, therefore our results may not be generalizable to other patient populations. Notably, IgAN is less frequent among individuals of African ancestry, including African Americans, suggesting that protective genetic effects may exist, but further studies are needed to address this hypothesis. We were also not able to evaluate the contribution of rare variants in this study, and sequencing studies are still needed to evaluate relative contributions of rare and common variants to the overall disease risk.

## ONLINE METHODS

### Study Cohorts, Genotyping, Genotype Quality Control and Imputation

The recruitment of patients, genotyping, imputation, and detailed quality control analyses are described by cohort in the **Supplemental Notes**.

### Association Analyses and Meta-analyses

We conducted genome-wide association analysis in each of the 17 cohorts using imputed genotype dosage data under a logistic regression additive model with adjustment for cohort-specific significant PCs in PLINK v1.9^42^. Subsequently, a fixed effects inverse-variance-weighted meta-analysis was performed to combine results from all cohorts using METAL version 2011-03-25^43^. Genome-wide distributions of p-values were examined visually using quantile-quantile (QQ) plots for each individual cohort as well as for the combined analysis. We also estimated the genomic inflation factors for each cohort. The final meta-analysis QQ plot showed no departure from the expected distribution of p-values, and the genomic inflation factor (λ) was estimated at 1.04 (**Extended Data Figure S1**). In addition, logistic regression association analysis assuming a dominant or a recessive genetic model was performed in PLINK using expected genotype counts, with no evidence of genomic inflation (λ=1.03 for dominant and 0.94 for recessive model) (**Extended Data Figure S3**). IgAN is more common in males; thus, we also performed sex-specific analyses, including chromosome X. Genome-wide logistic regression analyses were conducted separately in males and females within each cohort and subsequently meta-analyzed using METAL. A total of 21,236 males and 17,661 females were used in the meta-analysis with overall genomic inflation factors of 1.01 for males and 0.99 for females. A total of 1,990,322 high quality imputed chromosome X markers (r^2^>0.8 and MAF>0.01) were analyzed separately by sex, encoding genotypes as (0, 2) in males and (0, 1, 2) in females.

Significant PCs for each cohort were included as covariates in each model. We defined a locus as genome-wide significant if at least one SNP in the locus had p-value ≤ 5.0E-08 and it was successfully typed or imputed in ≥ 50% of analyzed cohorts. Signals with a p-value ≤ 1.0E-05 were considered as suggestive.

Summary of meta-analysis results are provided in **Table 1 and Supplementary Tables S2-4**.

**Table 1.** New and known genome-wide significant loci based on meta-analysis.

| Chr | Location (bp<sup>a</sup>) | SNP<sup>b</sup> | Locus | Risk Allele | Freq. European Controls | Freq. Asian Controls | OR | P-value | Q test | I<sup>2</sup> | Supporting Cohorts | New |
| --- | --- | --- | --- | --- | --- | --- | --- | --- | --- | --- | --- | --- |
| 1 | 157,542,162 | rs849815 | *FCRL* | A | 0.66 | 0.50 | 1.14 | 3.9E-09 | 0.57 | 0 | GWAS + IC | Known |
| 1 | 173,146,357 | rs4916312 | *TNFSF4* | A | 0.35 | 0.07 | 1.14 | 5.0E-08 | 0.68 | 0 | GWAS + IC | New |
| 1 | 196,686,918 | rs6677604 | *CFH* | G | 0.80 | 0.93 | 1.21 | 1.5E-17 | 0.97 | 0 | GWAS + IC | Known |
| 1 | 196,603,302 | rs12029571 | *CFH* | A | 0.22 | 0.35 | 1.12 | 2.5E-06 | 0.88 | 0 | GWAS | Known |
| 2 | 61,092,678 | rs842638 | *REL* | T | 0.44 | 0.15 | 1.17 | 9.6E-10 | 0.002 | 63.8 | GWAS | New |
| 2 | 204,584,759 | rs3769684 | *CD28* | T | 0.95 | 0.49 | 1.19 | 5.1E-11 | 0.93 | 0 | GWAS + IC | New |
| 4 | 74,725,320 | rs6828610 | *PF4V1/CXCL8* | G | 0.16 | 0.27 | 1.14 | 3.5E-08 | 0.98 | 0 | GWAS | New |
| 6 | 249,571 | rs12201499 | *IRF4/DUSP22* | C | 0.12 | 0.28 | 1.18 | 3.1E-11 | 0.39 | 5.1 | GWAS | Known |
| 6 | 7,214,676 | rs12530084 | *LY86* | C | 0.77 | 0.51 | 1.13 | 1.3E-09 | 0.49 | 0 | GWAS | New |
| 6 | 32,389,305 | rs9268557 | *HLA-DRA* | C | 0.51 | 0.57 | 1.24 | 4.5E-47 | 0.00 | 72.4 | GWAS + IC | Known |
| 6 | 32,667,829 | rs9275355 | *HLA-DQB/DQA* | C | 0.23 | 0.33 | 1.26 | 1.7E-34 | 0.21 | 24.8 | GWAS | Known |
| 6 | 32,599,999 | rs9272105 | *HLA-DQA* | A | 0.60 | 0.45 | 1.25 | 1.2E-28 | 0.001 | 73.2 | GWAS + IC | Known |
| 6 | 32,681,631 | rs9275596 | *HLA-DQB/DQA* | T | 0.66 | 0.81 | 1.33 | 3.2E-36 | 0.05 | 45.9 | GWAS + IC | Known |
| 6 | 33,074,288 | rs3128927 | *HLA-DPA/DPB* | C | 0.73 | 0.83 | 1.22 | 1.5E-25 | 0.38 | 6.1 | GWAS + IC | Known |
| 6 | 167,445,139 | rs2282859<sup>c</sup> | *CCR6* | C | 0.01 | 0.15 | 1.20 | 3.9E-08 | 0.06 | 43.8 | GWAS + IC | New |
| 8 | 6,808,722 | rs2075836 | *DEFA* | T | 0.31 | 0.30 | 1.21 | 5.8E-11 | 0.90 | 0 | GWAS | Known |
| 8 | 56,852,496 | rs75413466 | *LYN* | A | 0.02 | 0.06 | 1.40 | 1.4E-10 | 0.80 | 0 | GWAS + IC | New |
| 8 | 124,765,474 | rs34354351 | *ANXA3* | T | 0.17 | 0.32 | 1.15 | 3.5E-08 | 0.84 | 0 | GWAS | New |
| 9 | 117,643,362 | rs13300483 | *TNFSF8/15* | T | 0.24 | 0.31 | 1.13 | 1.3E-08 | 0.88 | 0 | GWAS + IC | New |
| 9 | 139,266,496 | rs4077515 | *CARD9* | T | 0.41 | 0.29 | 1.14 | 2.6E-11 | 0.41 | 1.8 | GWAS + IC | Known |
| 10 | 65,363,048 | rs57917667 | *REEP3* | G | 0.02 | 0.19 | 1.22 | 1.1E-08 | 0.91 | 0 | GWAS | New |
| 10 | 81,043,743 | rs1108618 | *ZMIZ1* | A | 0.60 | 0.49 | 1.14 | 1.9E-10 | 0.43 | 0.3 | GWAS + IC | New |
| 11 | 65,555,524 | rs10896045 | *RELA* | A | 0.30 | 0.48 | 1.18 | 4.7E-13 | 0.03 | 50.6 | GWAS | New |
| 11 | 128,487,069 | rs7121743 | *ETS1* | C | 0.16 | 0.47 | 1.13 | 3.4E-08 | 0.79 | 0 | GWAS | New |
| 14 | 107,222,014 | rs751081288 | *IGH* | A | 0.43 | 0.53 | 1.17 | 1.9E-08 | 0.74 | 0 | GWAS | New |
| 16 | 31,357,760 | rs11150612 | *ITGAM* | A | 0.64 | 0.27 | 1.16 | 8.4E-14 | 0.10 | 36.8 | GWAS+IC | Known |
| 16 | 86,017,715 | rs1879210 | *IRF8* | T | 0.64 | 0.86 | 1.14 | 9.9E-09 | 0.96 | 0 | GWAS+IC | New |
| 17 | 7,462,969 | rs3803800 | *TNFSF13* | A | 0.21 | 0.32 | 1.15 | 1.2E-10 | 0.19 | 27.1 | GWAS+IC | Known |
| 17 | 16,851,450 | rs57382045 | *TNFRSF13B* | A | 0.11 | 0.33 | 1.16 | 3.4E-09 | 0.84 | 0 | GWAS | New |
| 19 | 55,397,217 | rs1865097 | *FCAR* | A | 0.30 | 0.38 | 1.12 | 7.7E-09 | 0.49 | 0 | GWAS+IC | New |
| 22 | 30,512,478 | rs4823074 | *HORMAD2/LIF* | G | 0.54 | 0.67 | 1.16 | 7.8E-15 | 0.51 | 0 | GWAS | Known |

Chr., chromosome; freq., frequency; Q test, p-value for Cochran's Q statistic; I<sup>2</sup>, heterogeneity index; IC, Immunochip
<sup>a</sup> Genome Reference Consortium Human Build 37 (hg19).
<sup>b</sup> Only independent SNPs in each locus are included.
<sup>c</sup> East Asian genome-wide significant specific signal.

### Conditional Analyses

To detect multiple independent associations at individual genomic loci, we conducted a stepwise conditional analysis using a multi-SNP-based conditional and joint association analysis (COJO)^44^. This method approximates the variance-covariance matrix between association statistics with the LD information from an external reference panel and independent SNPs are selected in a stepwise manner using the GCTA tool version 1.92.0beta^44, 45^. Using our overall meta-analysis summary statistics, we conducted the GCTA conditional analysis with a threshold of *P*≤5.0E-08 and the LD reference composed of all European and East Asian cohorts from the 1000 Genomes Project Phase 3. Subsequent conditional analyses were performed for makers with a conditioned *P*≤5.0E-08 until no residual genome-wide significant associations were observed (**Supplementary Table S5**).

### HLA Imputation and Statistical Analysis

We used the SNP2HLA software to impute classical HLA alleles^46^. The Type 1 Diabetes Genetics Consortium (T1DGC) reference panel of 5,225 Europeans and 8,961 markers was used as a reference set for our European-ancestry cohorts^46^, and the Pan-Asian reference panel of 530 individuals and 8,245 markers was used for our East-Asian cohorts^47^. Only common and high-quality markers (MAF>0.01, R^2^>0.8) were used for association analysis. We analyzed each variant using a logistic regression model, assuming additive dosage effects and controlling for significant PCs of ancestry. For testing multi-allelic loci, we used the following logistic regression model:

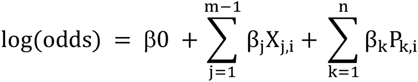

where *m* indicates a total number of alleles at a specific multi-allelic locus, *j* indicates a specific allele being tested, and *X_j,i_* is the imputed dosage for allele *j* for individual *i*; *β_0_* represents the intercept and *β_j_* represents the additive effect of an allele *j*; *P_k,i_* denotes the value for k^th^ PC of individual *i*, n is the total number of significant PCs in the dataset; *β_k_* is the effect size of principal component *k*. We compared log-likelihoods of two nested models: the full model containing the test locus (fitted model) and relevant covariates with the reduced model (null model) without the test locus, but with the same set of covariates. The deviance (*D*) was defined as – 2 x log likelihood ratio, which follows a χ^2^ distribution with *m-1* degrees of freedom, from which we calculated p-values:

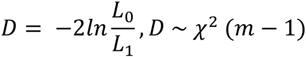

where *D* is the log-likelihood test value (deviance), *L_0_* is likelihood of the null model and *L_1_* the likelihood of the fitted model. To identify statistically independent effects, we first tested all bi-allelic and multi-allelic variants under the logistic regression model, as described above, and ranked them based on the p-value of the log likelihood test. Next, in a forward stepwise approach, we included in the logistic regression model the most statistically significant variant as a covariate, analyzed all remaining variants and ranked them based on the new p-value of the respective log likelihood test. We repeated the same steps until no variant or no HLA gene had P≤5.0E-08.

### HLA Peptide Sequence Analysis

HLA amino-acid polymorphisms have multiple possible residues at each peptide position. To test the effects of individual amino acid substitution sites we applied a conditional haplotype analysis using fully phased haplotypes across the HLA region. We tested each single amino acid position by first identifying the *m* possible amino-acid residues occurring at that position and then using *m-1* degrees of freedom test to derive p-values with a single amino-acid residue arbitrary selected as a reference. For conditioning on individual amino-acid sites, we used the following procedure: by adding a new amino-acid position to the model, a total of *k* additional unique haplotypes were generated and tested over the null model (without a new amino-acid position) using the likelihood ratio test with *k* degrees of freedom. If the new position was independently significant, we further updated the null model to include all unique haplotypes created by all amino-acid residues at both positions to identify another independent position. The procedure was repeated until no statistically significant (conditioned P≤5.0E-08) position was observed.

### Heritability and Genetic Correlations

SNP-based heritability was estimated using LD score regression (LDSC software)^10^ based on final meta-analysis summary statistics and LD scores estimated from 1000 Genomes phase 3 European and East Asian populations combined^48^. To assess the contribution of HLA region, we also estimated SNP heritability after excluding the entire MHC region (Chr.6: 28,000,000-33,000,000 bp). To investigate evidence for possible shared genetic effects between IgAN and other traits, we estimated genetic correlations using bivariate LD score regression^10^. For each phenotype, we used GWAS summary statistics from the largest GWAS available with a minimum coverage of 2 million SNPs. We excluded traits with estimated SNP-based heritability <1%. Genetic correlations were calculated with and without the HLA region. GWAS summary statistics for the relevant immune and cardio-metabolic traits were provided by corresponding consortia, or downloaded from the LD-hub or GWAS catalog; summary statistics for infection-related phenotypes were provided by 23andMe^49^.

### Pleiotropy Maps

All GWAS loci were cross-annotated against the studies listed in the GWAS catalogue (last update: January 31, 2019). For each locus, we selected all variants in strong LD (r^2^ ≥ 0.8) with the top SNP. We then queried the GWAS catalogue for genome-wide significant (p<5.0E-08) associations of the selected SNPs with other diseases and traits. The results were then manually verified by reviewing the original publications to confirm the direction of allelic effects. In cases where there were multiple GWAS for the same trait, we selected the SNP associations based on the largest sample size/lowest P-value and highest LD with our top SNP(s). To evaluate the overlap of pleiotropic effects between significant and suggestive IgAN loci, the traits associated with significant IgAN loci were queried themselves against GWAS catalogue for associations with any of the suggestive SNPs or their proxies. The results were represented as a shared susceptibility network map created in Cytoscape v3.7.0 software.

### Polygenic Risk Models

To assess the cumulative effect of independent significant and suggestive loci, we performed a genetic risk score (GRS) analysis. We first created two new GRS models based on the combined meta-analysis results: the 30-SNP model which comprises 30 independent genome-wide significant SNPs (25 non-HLA plus 5 HLA SNPs), and the 77-SNP GRS model which includes the same 30 SNPs plus 47 additional independent SNPs representative of the suggestive loci (p<1.0E-05). Each GRS was defined as the sum of the number of risk alleles weighted by their effect sizes. We required imputation R^2^>0.3 for including a SNP in the GRS calculation; individuals typed with Immunochip were excluded. Each GRS was standardized using a Z-score transformation with the mean and standard deviation of the control distribution. We evaluated the performance of each GRS by estimating two goodness of fit measures, Nagelkerke’s pseudo R^2^ and the area under the receiver operating characteristics curve (AUROC).

Genome-wide polygenic score (GPS) was calculated using LDpred^50^ and LD-pruning and p-value thresholding (P+T) methods, similar to the methods in recent studies^51, 52^. We used the combined meta-analysis including high quality imputed SNPs that overlapped across all cohorts (2,408,512 SNPs) but excluding Immunochip cohorts. In the LDPred method, the genetic architecture prior for variant effect sizes is a Gaussian distribution that has two parameters: heritability and a fraction of causal variants. Heritability was estimated using LD score regression, and the fraction of causal variants was used as a tuning parameter (ρ) across the following range: 1, 0.3, 0.1, 0.03, 0.01, 0.003, 0.001, 3.0E-04, 1.0E-04, 1.0E-05, 1.0E-06. Using LDPred, a genome-wide predictor was calculated for each value of ρ and the best performing score was selected. The P+T method prunes variants in LD and considers only variants with a p-value under a certain threshold. Using a range of different r^2^ (0.2, 0.4, 0.6, 0.8) and p-value thresholds (1, 0.3, 0.1, 0.03, 0.01, 0.003, 0.001, 3.0E-04, 1.0E-04, 3.0E-05, 1.0E-05, 1.0E-06, 1.0E-07, 5.0E-08, 1.0E-08), we again selected the best performing model. The performance of 30-SNP GRS, 77-SNP GRS, and best GPS were compared to the previously published 15-SNP GRS^4^. We additionally tested these models in the German Chronic Kidney Disease (GCKD) study^37, 38^, including 314 histologically confirmed IgAN cases versus 663 disease controls with a biopsy-diagnosed kidney disease of anther cause (see **Supplemental Note**). The analyses were implemented in R v.3.5.2 software.

### Gene Set and Pathway Enrichment Analyses

We defined each IgAN-associated genomic locus by first selecting all proxy SNPs in LD (r^2^≥0.5) with the lead SNP, then extending the genomic region 250 kb upstream and downstream of the first and last proxy SNP based on genomic position. Each region was then annotated using BiomaRt package, which retrieves Ensembl human gene annotations. Gene sets were created for all genome-wide significant and suggestive loci but excluding the HLA region. For gene set enrichment analysis, we used established gene sets from the Molecular Signatures Database (MSigDB), including GO, KEGG, BioCarta, REACTOME, chemical and genetic perturbations (CGP), and transcription factor targets (TFT). Statistical significance for enrichment was set at FDR q-value<0.05. We additionally applied a genome-wide gene set enrichment testing approach (excluding the HLA region) using Multi-marker Analysis of GenoMic Annotation (MAGMA) method with default parameters^12^. We also used DEPICT v1 release 194^14^ to perform pathway/gene set enrichment and tissue/cell-type analyses. For this analysis, we first used PLINK to identify independently associated SNPs setting P<5.0E-05 and r^2^<0.05 in a physical window of 500 kb. We then used DEPICT to prioritize genes, identified reconstituted gene sets that are enriched in genes from associated regions, and identified tissue and cell-type annotations in which genes from associated regions are highly expressed. Specifically, for each tissue, the DEPICT method performs a *t*-test comparing the tissue-specific expression of IgAN-associated genes versus all other genes. Next, for each tissue, empirical enrichment p-values are computed by repeatedly sampling random sets of loci from the entire genome to estimate the null distribution for the enrichment statistic as previously described^53, 54^.

### Prioritization of Causal Tissues and Cell Types

We estimated heritability enrichment of SNPs from GWAS summary statistics for functional categories in tissue and cell-type specific regulatory elements using stratified LD score regression. This method regresses the chi-squared statistics of SNPs from summary statistics on their LD scores^10^, and partitions SNP heritability by functional annotation^55^. We used summary statistics obtained from the meta-analysis of all case-control cohorts excluding the MHC region and excluding the cohorts typed with Immunochip. Heritability enrichment was defined as the proportion of SNP heritability in a specific category, divided by the proportion of SNPs that belong to that category. We first calculated heritability enrichment for a baseline model of 96 non-cell-type-specific functional categories. Along with the 1000 Genomes reference panel, we used this baseline model as a control to assess heritability enrichment in cell-type specific functional annotations. We used functional annotations from the ENCODE and Roadmap Epigenomics Consortium^56^, as well as mouse immune cell-specific functional categories from the Immunological Gene Project (ImmGen)^57^. Using the same method, we evaluated tissue- and cell-specific heritability enrichments based on the FUN-LDA functional scoring system^13^. As an alternative method, we also used GARFIELD v2^15^ to assess enrichment within the ENCODE and Roadmap-derived regulatory regions.

### Analysis of Relevant Phenotypes in Mice

As a complementary approach to prioritize genes within each locus, we used the Mouse Genome Informatics (MGI) database to identify potential genes the disruption of which causes relevant phenotypes in mice^58^. All phenotypes in MGI are categorized based on the Mammalian Phenotype (MP) ontology and emerge as a result of different genetic models, including targeted knockout animals and chemically induced (ENU) and spontaneous mutations. MGI includes a total of 17,101 mouse genes with Human orthologs^59^. We defined a gene set of 62 genes with mouse orthologs across the 24 non-HLA risk loci for testing against MGI phenotypes to define significantly enriched categories.

### Functional Annotations of Individual Loci

For the purpose of detailed functional annotation, we calculated 95% credible sets for each of the genome wide significant loci using CAVIAR software^60^. We added variants that were neither typed or imputed in our data, but in strong LD with the top SNP based on external reference (r2≥0.8 in 1000G European and East Asian populations). These SNP sets were annotated using ANNOVAR to first define any coding variants and their predicted effects. Using FUN-LDA method, we next calculated the posterior probability of a functional effect for each of the selected variants across 127 tissues and cell types as described previously^13^. These SNPs were also interrogated against the following datasets: (1) eQTLs for 13 human immune cell types from the Database of Immune Cell eQTLs (DICE) project^61^; (2) blood eQTLs from the eQTLGen Consortium^26^ (31,684 individuals), (3) GTEx tissue eQTLs^62^ (4) GTEx splicing QTLs (sQTLs); (5) kidney eQTLs (glomerular and tubular) from the Kidney eQTLs Atlas^63^, (6) blood mQTLs from KORA (N=2,820) and TwinsUK (N=7,824) studies^64, 65^, and (7) blood pQTLs from three recent well-powered studies^66–68^. We additionally performed co-localization analysis between IgAN and eQTL summary statistics for each GWAS locus using COLOC software^69^. We considered PP4>0.5 as supportive of a shared causal variant.

### Protein-Protein Interactions

Protein-protein interactions across genome wide significant and suggestive genes have been predicted using InWeb_InBioMap (InWeb_IM)^70^. InWeb_IM is a curated and computationally derived protein-protein network of 420,000 protein-protein interactions that has 2.8 times more interactions than other comparable resources. We used high confidence PPIs from InWeb_IM using the recommended cut off confidence score ≥ 0.1. All annotated genes within the 76 significant and suggestive IgAN loci were used to probe the PPI database; the final network contained a total of 53 nodes connected by 63 edges.

Enrichment P-value was computed using a hypergeometric test and corrected for multiple testing using the method of Benjamini and Hochberg. The network components were grouped into modules based on their pathway categorization; the GLay community clustering algorithm was implemented for module detection and modules were visualized in GeNets^71^. Subsequently, the Clustering with Overlapping Neighborhood Expansion (ClusterONE) algorithm^72^ implemented in Cytoscape^73^ was used to extract protein clusters using the default parameters and the InWeb_IM confidence score as edge weights.

Functional and pathway enrichments within the PPI networks were identified using STRING^74^ based on Gene Ontology (GO), KEGG, and Reactome databases. In addition, we used ToppGene Suite^75^ to calculate protein-protein interaction enrichment p-values. A Bonferroni-corrected P < 0.05 was used as enrichment significance cut-off.

### Transcription Factor-DNA Binding Interactions

To identify TF binding sites enriched across IgAN risk loci, we used the RELI (Regulatory Element Locus Intersection) algorithm^16^. RELI uses a set of genetic variants as input, expands the set using LD blocks (r^2^>0.8) and calculates the statistical intersection of the resulting loci with ChiP-seq datasets by counting the number of loci with one or more variants intersecting the TF ChiP-seq peaks. The LD blocks were calculated using 1000 Genomes Project East Asian and European populations combined. The null distribution was generated using 2,000 random repeats of the procedure and was used to calculate z-scores and empirical p-values for the observed intersection. The final reported p-values are Bonferroni-corrected (Pc) for the 1,544 TF datasets tested, as previously published^16^. As input, we used a set of 28 independent genome-wide significant loci (P≤5.0E-08) and a set of 76 loci including genome-wide significant and suggestive variants (P<1.0E-05). A Pc<1.0E-04 was used as a significance cut-off for each set.

### Ligand-Receptor Pairs

To identify the number of potential ligand-receptor pairs significantly associated with IgAN, we queried The Database of Ligand-Receptor Partners (DLRP)^76^. This database includes cytokines, chemokines, and growth, angiogenesis, and developmental factors, and contains 175 protein ligands, 131 protein receptors and 451 experimentally determined ligand-receptor pairings. To test for ligand-receptor enrichment in our dataset, we used a hypergeometric test for overlap between this dataset and the gene set defined by our significant and suggestive GWAS loci.

### Analysis of Drug Targets

We obtained drug target genes and corresponding drug information from DrugBank^77^, the Therapeutic Targets Database (TTD)^78^, the Open Targets Platform^79^, and GlobalData combined with manual literature searches. To search for potential drug targets, we extracted all genes in direct PPIs with IgAN risk genes by using the In_Web_IM database. We selected drug target genes that had pharmacological activities and human orthologues, and that were targeted by any of the drugs that are approved or currently in development (experimental or in clinical trials).

### Overall Prioritization of Biological Candidate Genes

Each of the positional candidate genes was scored adopting the following criteria and calculating the number of the satisfied criteria, including: (1) genes most proximal to the top SNP; (2) genes with coding variants in 95% credible sets and/or high LD (r^2^>0.8) with the index SNP; (3) genes with promoter chromatin interaction by Activity-by-Contact (ABC) model^34^ or (4) GeneHancer^35^ involving regions intersected by top SNP and its 95% credible sets/high LD proxies; (5) e-genes controlled by at least one eQTL (any tissue) tagged by the top SNP in any tissues (primary immune cells, whole blood, kidney, GTEx); (6) e-genes co-localized with the risk locus in blood or (7) primary immune cells with PP4>0.5; (8) p-genes controlled by at least one blood pQTL tagged by the top SNP; (9) genes prioritized by PPI network connectivity analysis at P<0.05; (10) genes that when knocked out in mice produce at least two phenotype labels: ‘immune system’, ‘haematopoietic system’, or ‘cellular phenotype’; (11) genes prioritized by MAGMA, (12) DEPICT with gene-based p<0.05, or (13) manual review of the literature as related to IgAN, IgA production, or mucosal immunity.

### Genotype–phenotype Correlations

Our polygenic risk models (15-SNP, 30-SNP, 77-SNP, and GPS) were tested for clinical correlations in the subset of cases with available clinical data. We fitted each standardized risk score in a regression model to predict clinical disease features at the time of diagnosis, including age at biopsy, estimated glomerular filtration rate (GFR), proteinuria, microhematuria, hypertension (HTN), and gross hematuria. The GFR was estimated using the CKD-EPI formula in adults^80^ and Schwartz formula in pediatric cases^81^. The estimated GFR was normalized with a natural logarithm transformation; proteinuria was normalized with a ln(P24+1) transformation; microhematuria was assessed with urinary dipstick and defined as positive if 1+ or greater; gross hematuria was defined by the reported history of red urine before a diagnostic biopsy; hypertension was defined as systolic blood pressure ≥ 140 mmHg and/or diastolic blood pressure ≥ 90 mmHg, or anti-hypertensive medication use. For the analysis of longitudinal outcomes, we performed survival analysis with the primary outcome of kidney failure defined as eGFR<15ml/min/1.73 m^2^ or initiation of kidney replacement therapy (dialysis or kidney transplantation). All analyses were adjusted for age, gender, site, and race/ethnicity. The analyses were implemented in R v3.5.2.

### Phenome-wide association studies (PheWAS)

We performed meta-analysis of PheWAS results (meta-PheWAS) across two large biobank-based datasets: Electronic Medical Records and Genomics-III (eMERGE-III) and the UK Biobank (UKBB). The eMERGE-III network provides access to EHR information linked to GWAS data for 102,138 individuals; detailed quality control analyses of genetic data have been described previously^82–84^. Briefly, GWAS datasets were imputed using the latest multiethnic Haplotype Reference Consortium (HRC) panel using Michigan Imputation Server^85^. The imputation was performed in 81 batches across the 12 contributing medical centers participating in eMERGE-I, II, and III. For post-imputation analyses, we included only markers with minor allele frequency (MAF) ≥ 0.01 and *R*^2^ ≥ 0.8 in ≥ 75% of batches. A total of 7,529,684 variants were retained for the GPS analysis. For principal component analysis (PCA), we used FlashPCA^86^ on a set of 48,509 common (MAF>0.01) and independent variants (pruned in PLINK with --indep-pairwise 500 50 0.05 command). The UKBB is a large prospective population-based cohort that enrolled individuals ages 40-69 for the purpose of genetic studies^87^. This cohort is comprised of 488,377 individuals recruited since 2006, genotyped with high-density SNP arrays, and linked to electronic health record data. All individuals underwent genome-wide genotyping with UK Biobank Axiom array from Affymetrix and UK BiLEVE Axiom arrays (~825,000 markers). Genotype imputation was carried out using a 1000 Genomes reference panel with IMPUTE4 software^88–90^. We then applied QC filters similar to eMERGE-III, retaining 9,233,643 common (MAF ≥ 0.01) variants imputed with high confidence (*R*^2^ ≥ 0.8). For principal component analysis by FlashPCA^86^, we used a set of 35,226 variants that were common (MAF>0.01) and pruned using the following command in PLINK --indep-pairwise 500 50 0.05. In order to test the GPS for associations phenome-wide across both eMERGE and UKBB datasets, we first harmonized the coded diagnoses data by converting all available ICD-10-CM codes to ICD-9-CM system. This approach was motivated by the fact that the great majority of data for eMERGE-III participants is already coded using ICD-9 system, and ICD-10 codes are more granular, thus a reverse conversion leads to mapping errors. After the conversion, the 102,138 genotyped eMERGE participants had a total of 20,783 ICD-9 codes that were then mapped to 1,817 distinct phecodes (disease-specific groupings of ICD codes). The 488,377 UKBB participants had a total of 10,221 ICD-9 codes that that were mapped to 1,817 phecodes. Phenome-wide associations were performed using the PheWAS R package^91^. The package uses pre-defined “control” groups for each phecode. The case definition requires a minimum of two ICD-9 codes from the “case” grouping of each phecode. In total, all 1,817 phecodes were tested using logistic regression with each phecode case-control status as an outcome and the polygenic score IgA nephropathy adjusted for age, sex, study site or batch, and 3 principal components of ancestry as a predictor. We then performed meta-PheWAS across both datasets combined using metagen with fixed effect in PheWAS R library^91^. To establish significant disease associations in PheWAS, we set the Bonferroni-corrected statistical significance threshold at 2.75E^−05^ (0.05/1,817) correcting for 1,817 independent phecodes tested.

### Web Resources

**COLOC:** https://cran.r-project.org/web/packages/coloc/

**DICE:** https://dice-database.org/

**EAGLE:** https://data.broadinstitute.org/alkesgroup/Eagle/

**eQTLGen:** https://www.eqtlgen.org/

**DEPICT:** https://data.broadinstitute.org/mpg/depict/index.html

**DLRP:** https://dip.doe-mbi.ucla.edu/dip/DLRP.cgi

**DrugBank:** https://www.drugbank.ca

**FUN-LDA:** http://www.columbia.edu/~ii2135/funlda.html

**GARFIELD:** https://www.ebi.ac.uk/birney-srv/GARFIELD/

**GCTA-COJO:** https://cnsgenomics.com/software/gcta/#COJO

**GeNets:** http://apps.broadinstitute.org/genets

**GlobalData**: https://www.globaldata.com/industries-we-cover/pharmaceutical

**GSEA:** http://software.broadinstitute.org/gsea/msigdb/

**GTEx:** https://gtexportal.org/home/

**GWAS catalog**: https://www.ebi.ac.uk/gwas

**InWeb:** http://apps.broadinstitute.org/genets#InWeb_InBiomap

**KING:** http://people.virginia.edu/~wc9c/KING/

**Kidney eQTL Atlas:** http://susztaklab.com/eqtl

**LD hub:** http://ldsc.broadinstitute.org/ldhub

**LDpred:** https://github.com/bvilhjal/ldpred

**LDSC:** https://github.com/bulik/ldsc

**Metabolomics GWAS Server:** http://metabolomics.helmholtz-muenchen.de/gwas/

**METAL:** http://csg.sph.umich.edu/abecasis/Metal/

**MGI:** http://www.informatics.jax.org

**MINIMAC3:** http://genome.sph.umich.edu/wiki/Minimac3

**MIP:** https://imputationserver.sph.umich.edu

**NEPTUNE eQTL Browser:** http://nephqtl.org/

**Open Targets:** https://www.targetvalidation.org

**PheWAS:** https://github.com/PheWAS/PheWAS

**PheWeb:** http://pheweb.sph.umich.edu/SAIGE-UKB/

**PLINK:** https://www.cog-genomics.org/plink/1.9/

**RELI:** https://github.com/WeirauchLab/RELI

**SNP2HLA:** http://software.broadinstitute.org/mpg/snp2hla/snp2hla_manual.html

**STRING:** https://string-db.org

**ToppGene:** https://toppgene.cchmc.org

**TTD:** https://db.idrblab.org/ttd

## Data Availability

All data produced in the present study are available upon reasonable request to the authors.

## Acknowledgments

We are grateful to all study participants across multiple nephrology centers worldwide for their contributions to this manuscript. This work was supported by the following institutions, grants, and funding agencies: Columbia University, Columbia Glomerular Center, IGA Nephropathy Foundation of America, and National Institute for Diabetes and Digestive Kidney Diseases (NIDDK) grants R01-DK105124 (KK, JN, BAJ) and RC2-DK116690 (KK), R01-DK082753 (AGG, JN, KK, FS, BAJ, RJW). Additional support was provided by R01LM013061 (KK), U01HG008680 (KK), U01AI152960 (KK), R01-DK078244 (JN, BAJ), R01-AI149431 (JN, BAJ), National Science Foundation of China (82022010, ZX; 82070733, ZH); Beijing Natural Science Foundation (Z190023,ZX); DFG Project-ID 192904750 – CRC 992 Medical Epigenetics (AK, PS), DFG Project-ID 431984000 – CRC 1453 (AK), and the EQUIP Program for Medical Scientists, Faculty of Medicine, University of Freiburg (PS). JBH, SP, KL, and WM were supported by R01HG010730, U01AI130830, R01NS099068, R01AI024717, R01AR073228, U01AI150748, R01AI148276, P01AI150585 and the US Department of Veterans Affairs Merit Award I01-BX001834 to JBH. DPG was supported by the St Peter’s Trust for Kidney, Bladder and Prostate Research. The UK cohort data was generated as a result of a grant from Kidney Research UK and the Medical Research Council (JB and DG). The German STOP-IgAN study was supported by the Deutsche Forschungsgemeinschaft (DFG, German Research Foundation) – CRU 5011 – Project-ID 445703531 (J.F., T.R.). The GCKD (German Chronic Kidney Disease) study was funded by grants from the German Ministry of Education and Research (BMBF, No. 01ER0804) and the KfH Foundation for Preventive Medicine. Unregistered grants to support the study were provided by Bayer, Fresenius Medical Care, and Amgen. Genotyping was supported by Bayer Pharma AG. The recruitment of Polish cases with IgA nephropathy was sponsored by the Polish Kidney Genetics Network (POLYGENES), a collaborative effort between Columbia University and Poznań University of Medical Sciences, Poland. The recruitment of Czech patients with IgA nephropathy was supported by the research project of General University Hospital in Prague (RVO-VFN64165). The recruitment of Russian cohort was supported by the Government Assignment of the Russian Ministry of Health, Assignment #200080056 of the Veltischev Research and Clinical Institute for Pediatrics of the Pirogov Russian National Research Medical University. The recruitment of the Swedish cohort was supported by the Nephrology and Rheumatology Departments at Karolinska University Hospital and Karolinska Institutet, Stockholm, and grants from the Swedish Society of. Medicine. We thank the Immunopathology Working Group of the Italian Society of Nephrology (ISN) for inviting their member sites to contribute to this study. We are also grateful to the Pediatric Nephrology Research Consortium (PNRC) for hosting and co-sponsoring the GIGA-kids Study that recruited pediatric patients with IgAN across PNRC sites. We are additionally grateful to Corinna Bowers for coordinating GIGA-kids and PMRC-based recruitment, Dr. Joann Narus for coordinating recruitment at the University of Utah, Dr. Ewa Elenberg and Dr. Shweta Shah for helping enroll subjects from Texas Children’s Hospital and Adisak Suwanichkul for sample shipping. The recruitment of the Korean cohort was supported by the Seoul National University Hospital Human Biobank, a member of the National Biobank of Korea, financed by the Ministry of Health and Welfare, Republic of Korea. We are grateful to the International Multiple Sclerosis Genetics Consortium (IMSGC), the Inflammatory Bowel Disease Genetics Consortium (IBDGC), the International Myositis Consortium (IMC), the Feinstein Institute for Medical Research, the French Biological Ressource Center for MS Genetics (REFGENSEP), Genethon, and INSERM for contributing Immunochip controls for the purpose of this study. We would also like to thank the Population Architecture Using Genomics and Epidemiology (PAGE) consortium, funded by the National Human Genome Research Institute (NHGRI) with co-funding from the NIMHD, for providing population controls genotyped with MEGA chip for this study. The funding sources were not involved in the study design, collection, analysis, and interpretation of data, writing of the report, or in the decision to submit the paper for publication.

## Authors Contributions

K. Kiryluk and A.G. Gharavi conceived the study, provided overall supervision of the project, and made the decision to publish the findings; E. Sanchez-Rodriguez and N. Mladkova performed quality control, imputation, and GWAS association analyses for GWAS and Immunochip discovery cohorts. E. Sanchez-Rodriguez performed final statistical analyses, meta-analyses, fine-mapping studies, functional annotations, and drug target analyses; F. Zanoni and A. Khan designed polygenic risk scores, tested for genome-wide genetic correlations, and performed clinical correlation analyses; L. Liu performed analysis of the HLA locus; J.B. Harley, M. Weirauch, S. Eswar, S. Parameswaran, and L. Kottyan performed RELI transcription factor analysis; I. Ionita-Laza consulted on the statistics and functional annotation of GWAS loci; K. Kiryluk, A.G. Gharavi, J. Novak provided biological interpretation of GWAS loci; X. Zhou and H. Zhang coordinated recruitment, genotyping, and analysis of the Beijing cohorts; F. Scolari coordinated recruitment activities across the Italian network of clinical recruitment sites; Y. Caliskan coordinated recruitment activities across the Turkish recruitment sites; H. Trimarchi coordinated recruitment across the Argentinian network of clinical recruitment sites; V.Tesar and D.Maixnerova coordinated the recruitment of the Czech cohort; J. Xie and N. Chan coordinated recruitment, genotyping, and analysis of the Shanghai cohorts; H. Suzuki recruited and clinically characterized the Japanese Discovery cohort. H. Lee, J. Park, BL Cho, Y.S. Kim, and D.K. Kim recruited and clinically characterized the Korean Discovery cohort. J. Floege and T. Rauen contributed DNA and clinical data for the German STOP-IgAN cohort; K. Kiryluk, R. Nelson, R. Wyatt, and B. Smoyer led the GIGA-kids study in collaboration with the Pediatric Nephrology Research Consortium (PNRC); M. Marasa, O. Balderes, J.Y. Zhang coordinated recruitment at Columbia University and managed clinical data and DNA samples; J. Barratt, and D. P. Gale provided summary statistics for the UK GWAS cohort; R. P. Lifton contributed previously published European and Beijing GWAS cohorts; A. B. Ekici, K-U. Eckardt, and A. Köttgen contributed German cases to the Immunochip discovery cohorts, and P. Schlosser performed polygenic risk score validation studies in the GCKD cohort; D. van Heel, B. Cotsapas, C. Wijmenga, A. Franke, V. Annese, and P.K. Gregersen contributed Illumina Immunochip idat files for population controls; R. Loss and E. Kenny contributed Illumina MEGA chip idat files for population controls. All other co-authors contributed to the recruitment and clinical characterization of IgAN patients and controls recruited from their respective clinical centers. All authors have read and approved the final version of manuscript.

## Ethics Statement

All subjects provided informed consent to participate in genetic studies, and the Institutional Review Board of Columbia University (protocol number IRB-AAAC7385) as well as local ethics review committees for each of the individual cohorts approved our study protocol.

## Data Availability

Primary genotype data for previously published cohorts is available through dbGAP under accession number phs000431.v2.p1. Genotype data for newly added cohorts will be available through dbGAP upon publication (accession number pending). Our IRB determined that the use of this dataset is restricted to genetic studies of kidney disease. The PAGE consortium control genotype data is available on dbGAP under accession number phs000356.v2.p1. The Electronic Medical Records and Genomics-III (eMERGE-III) imputed genotype and phenotype data are available through dbGAP, accession number: phs001584.v2.p2. The UK Biobank genotype and phenotype data are available through the UK Biobank web portal https://www.ukbiobank.ac.uk/. All data and summary statistics are also available from the corresponding authors upon reasonable request.

