## Supplemental information for "GWAS defines pathogenic signaling pathways and prioritizes drug targets for IgA nephropathy"

*Kiryluk K et al.*

### SUPPLEMENTAL INFORMATION:

#### SUPPLEMENTARY FIGURES:


**Extended Data Fig. 1.** ..... 3
**Extended Data Fig. 2.** ..... 4
**Extended Data Fig. 3.** ..... 5
**Extended Data Fig. 4.** ..... 6
**Extended Data Fig. 5.** ..... 7
**Extended Data Fig. 6.** ..... 8
**Extended Data Fig. 7.** ..... 9
**Extended Data Fig. 8.** ..... 10


#### SUPPLEMENTARY TABLES:


**Supplementary Table S1.** ..... 11
**Supplementary Table S2.** ..... 12
**Supplementary Table S3.** ..... 13
**Supplementary Table S4.** ..... 14
**Supplementary Table S5.** ..... 15
**Supplementary Table S6.** ..... 16
**Supplementary Table S7.** ..... 17
**Supplementary Table S8.** ..... 18
**Supplementary Table S9.** ..... 19
**Supplementary Table S10.** ..... 20
**Supplementary Table S11.** ..... 21
**Supplementary Table S12.** ..... 22
**Supplementary Table S13.** ..... 23
**Supplementary Table S14.** ..... 24
**Supplementary Table S15.** ..... 25
**Supplementary Table S16.** ..... 26
**Supplementary Table S17.** ..... 27
**Supplementary Table S18.** ..... 28
**Supplementary Table S19.** ..... 29
**Supplementary Table S20.** ..... 30
**Supplementary Table S21.** ..... 31
**Supplementary Table S22.** ..... 32
**Supplementary Table S23.** ..... 33
**Supplementary Table S24.** ..... 34
**Supplementary Table S25.** ..... 35
**Supplementary Table S26.** ..... 36
**Supplementary Table S27.** ..... 37
**Supplementary Table S28.** ..... 38
**Supplementary Table S29.** ..... 39
**Supplementary Table S30.** ..... 40


#### SUPPLEMENTARY NOTES:


**Discovery Cohorts** ..... 41
**Supplementary References** ..... 44

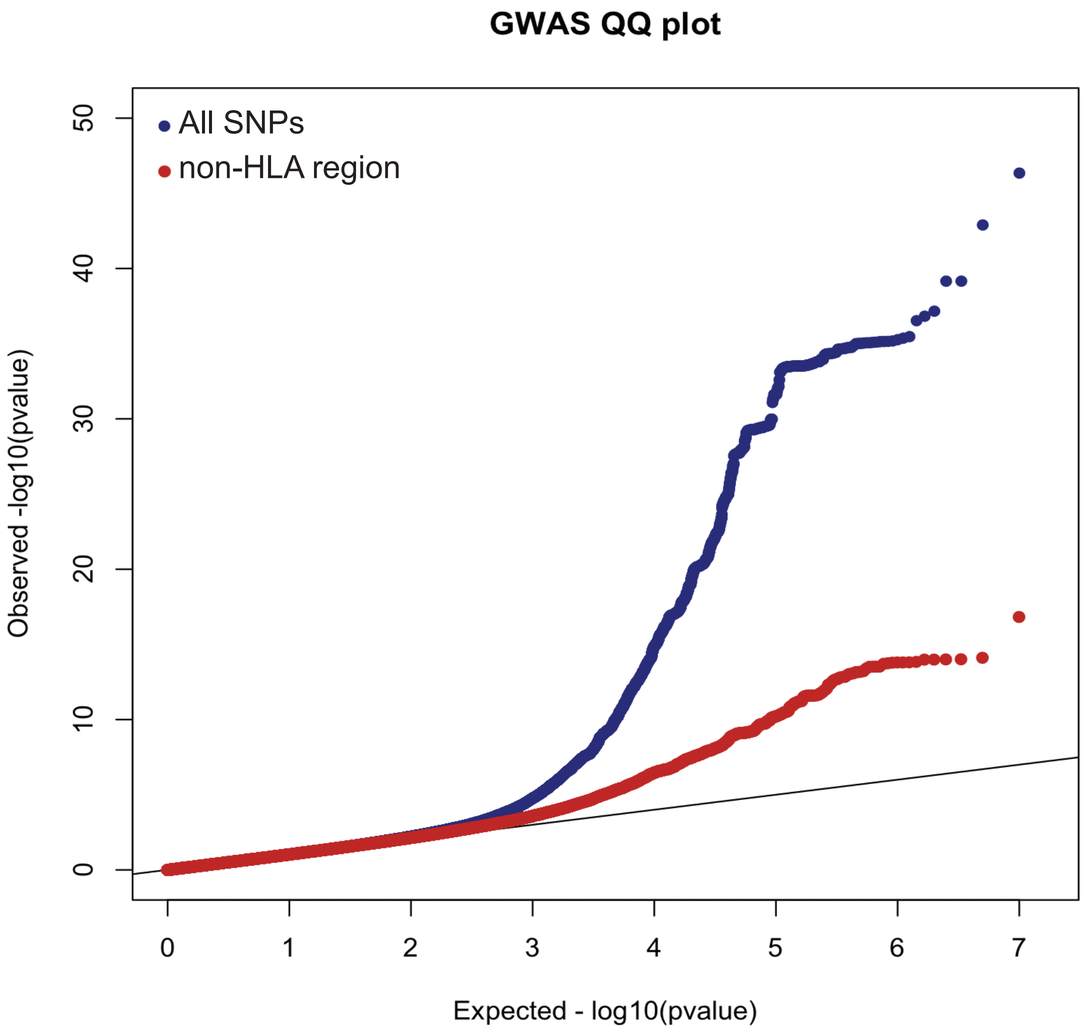


**Extended Data Fig. 1. Quantile-quantile (QQ) plot of the combined meta-analysis across 38,897 individuals.** Blue dots represent the QQ plot based on all SNPs in the meta-analysis and red dots represent the QQ plot after exclusion of SNPs within the MHC region. The overall genomic inflation factor ($\lambda$) was 1.048 with MHC, and 1.042 without MHC region.

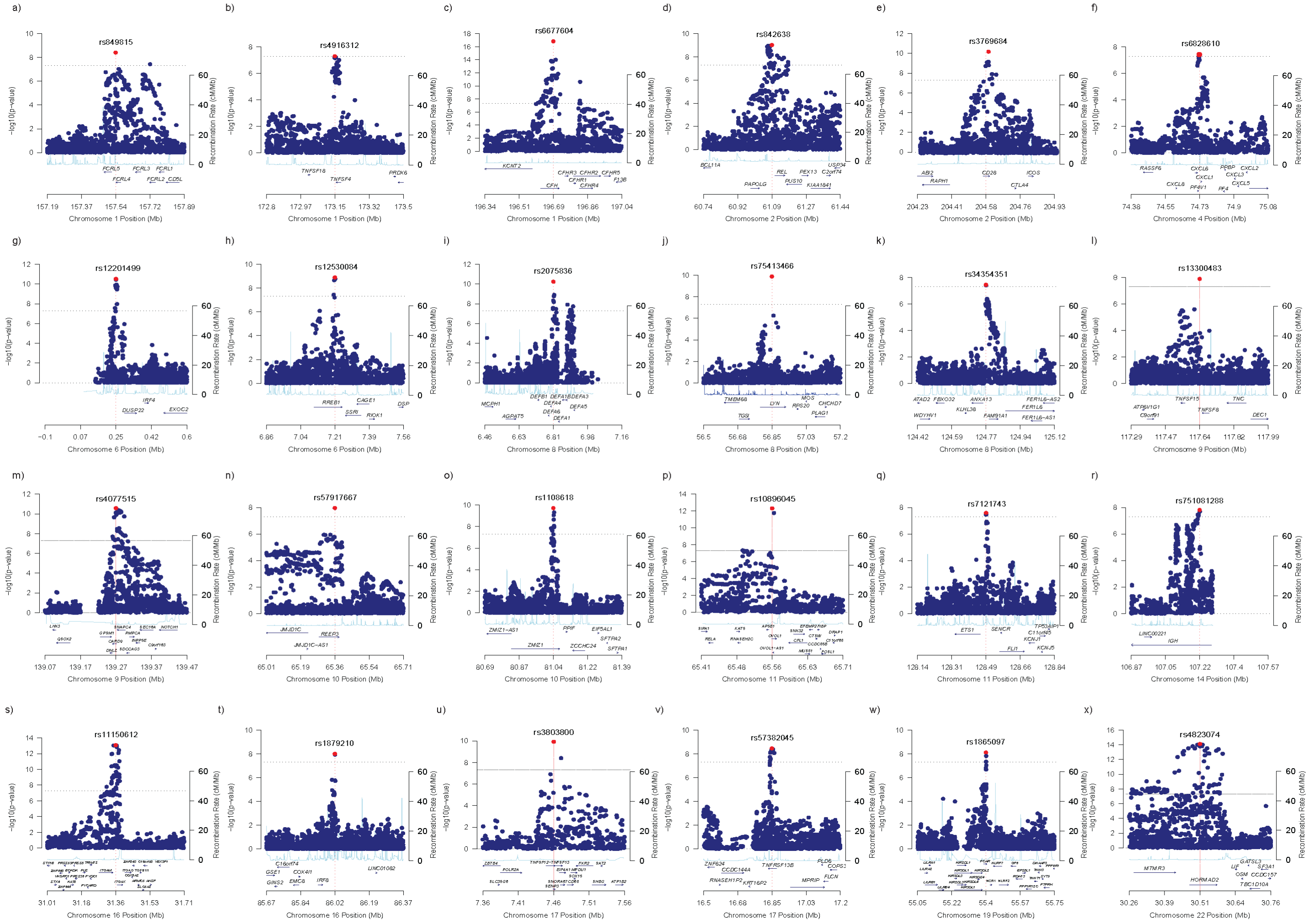


**Extended Data Fig. 2. Regional plots for non-HLA genome-wide significant loci: (a)** the *FCRL* locus, **(b)** the *TNFSF4* locus, **(c)** the *CFH* locus, **(d)** the *REL* locus, **(e)** the *CD28* locus, **(f)** the *PF4V1/CXCL8* locus, **(g)** the *IRF4* locus, **(h)** the *RREB1* locus, **(i)** the *DEFA* locus, **(j)** the *LYN* locus, **(k)** the *ANXA3* locus, **(l)** the *TNFSF8* locus, **(m)** the *CARD9* locus, **(n)** the *REEP3* locus, **(o)** the *ZMIZ1* locus, **(p)** the *RELA* locus, **(q)** the *ETS1* locus, **(r)** the *IGH* locus, **(s)** the *ITGAM* locus, **(t)** the *IRF8* locus, **(u)** the *TNFSF13* locus, **(v)** the *TNFRSF13B* locus, **(w)** the *FCAR* locus, **(x)** the *HORMAD2/LIF* locus. The x-axis shows the physical position in megabases (Mb, hg19 coordinates) and known genes; the left y-axis presents -log10 p-values for association statistics and the right y-axis shows the recombination rate across the region; the dotted horizontal line indicates a genome-wide significant threshold of 5.0E-08.

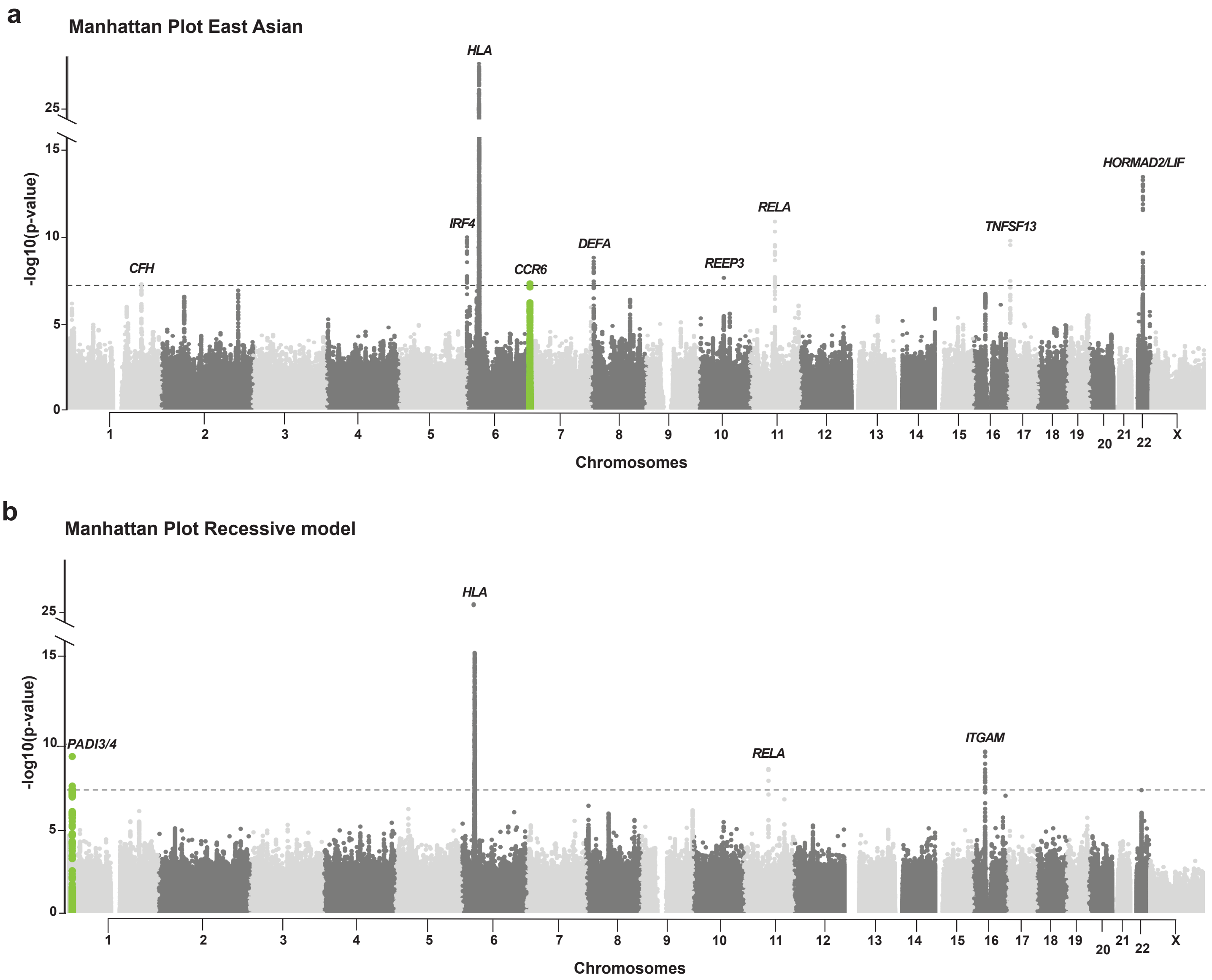


**Extended Data Fig. 3. Manhattan plots for East Asian GWAS subgroup analyses. (a)** East Asian meta-analysis under an additive genetic model (λ=1.040) revealed a new genome-wide significant locus on chr.6 (*CCR6,* green); **(b)** East Asian meta-analysis under a recessive model (λ=0.940) revealed a new genome-wide significant locus on chr.1 (encoding *PADI3* and *PADI4*, green); y-axis: -log10 of the p-value and is truncated to accommodate the HLA peak; x-axis: genomic position along each chromosome (1-22 and X); the dotted horizontal line indicates a genome-wide significant threshold of 5.0E-08.

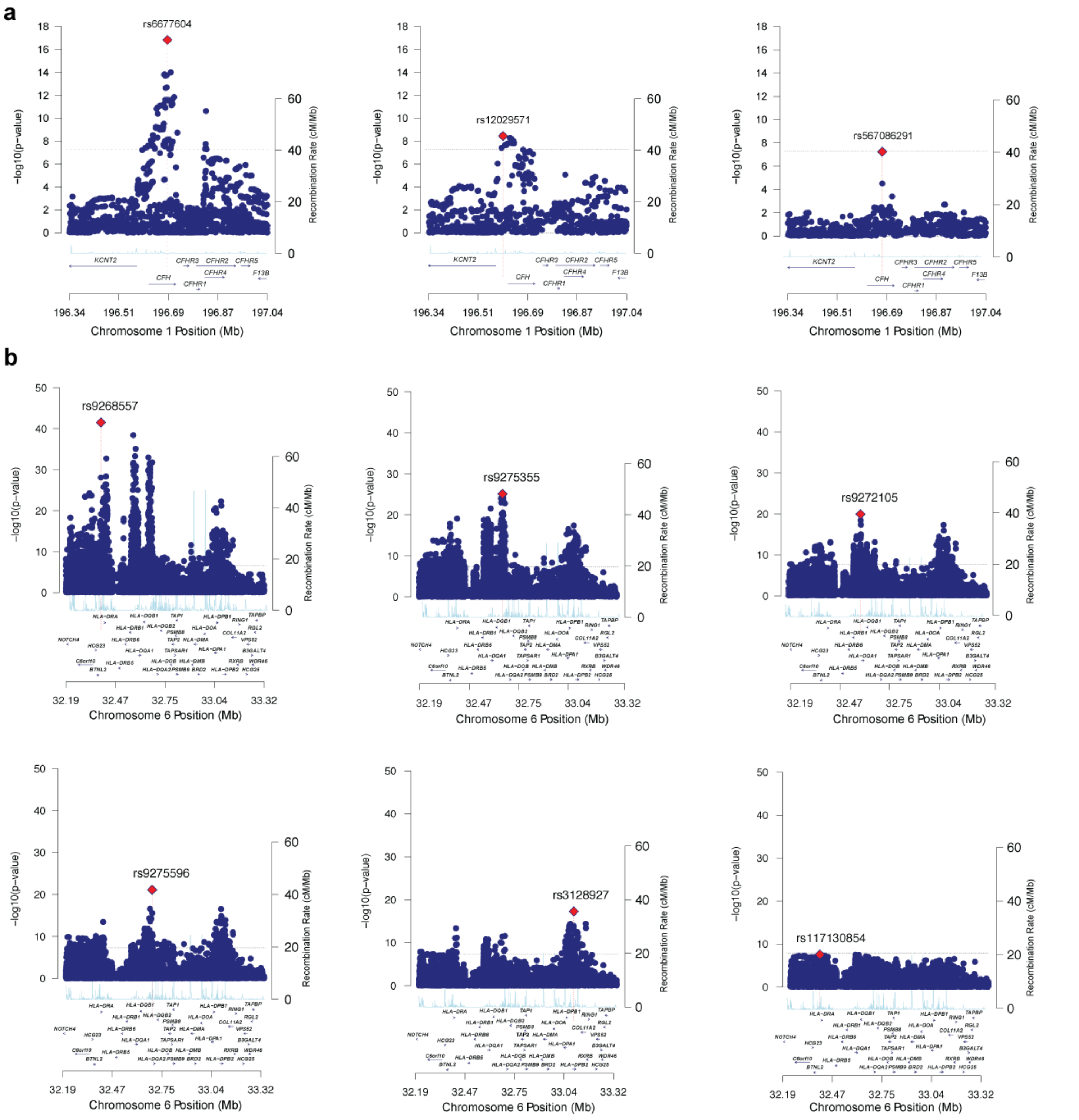


**Extended Data Fig. 4. Stepwise Conditional Analyses of the *CFH* and *HLA* loci. (a) *CFH* locus**: initial meta-analysis results without conditioning (top left); after conditioning for the top SNP rs6677604 (top middle); and after controlling for the two significant SNPs rs6677604 and rs12029571 (top right). **(b) *HLA* locus:** initial meta-analysis results without conditioning (middle left); after conditioning for the top SNP rs9268557 (middle), after controlling for rs9268557 and rs9275355 (middle right), after controlling for rs9268557, rs9275355 and rs9272105 (bottom left), after controlling for rs9268557, rs9275355 rs9272105 and rs9275596 (bottom middle), and after controlling for rs9268557, rs9275355 rs9272105, rs9275596 and rs3128927 (bottom right) with no additional significant signals. The x-axis shows genomic position in Mb (hg19 coordinates) and known genes; the left y-axis presents -log10 p-values for association statistics; the right y-axis (light-blue line) shows the averaged recombination rate across the region; the dotted horizontal line indicates a genome-wide significant threshold of 5.0E-08; the top SNP in each panel is marked by a red diamond.

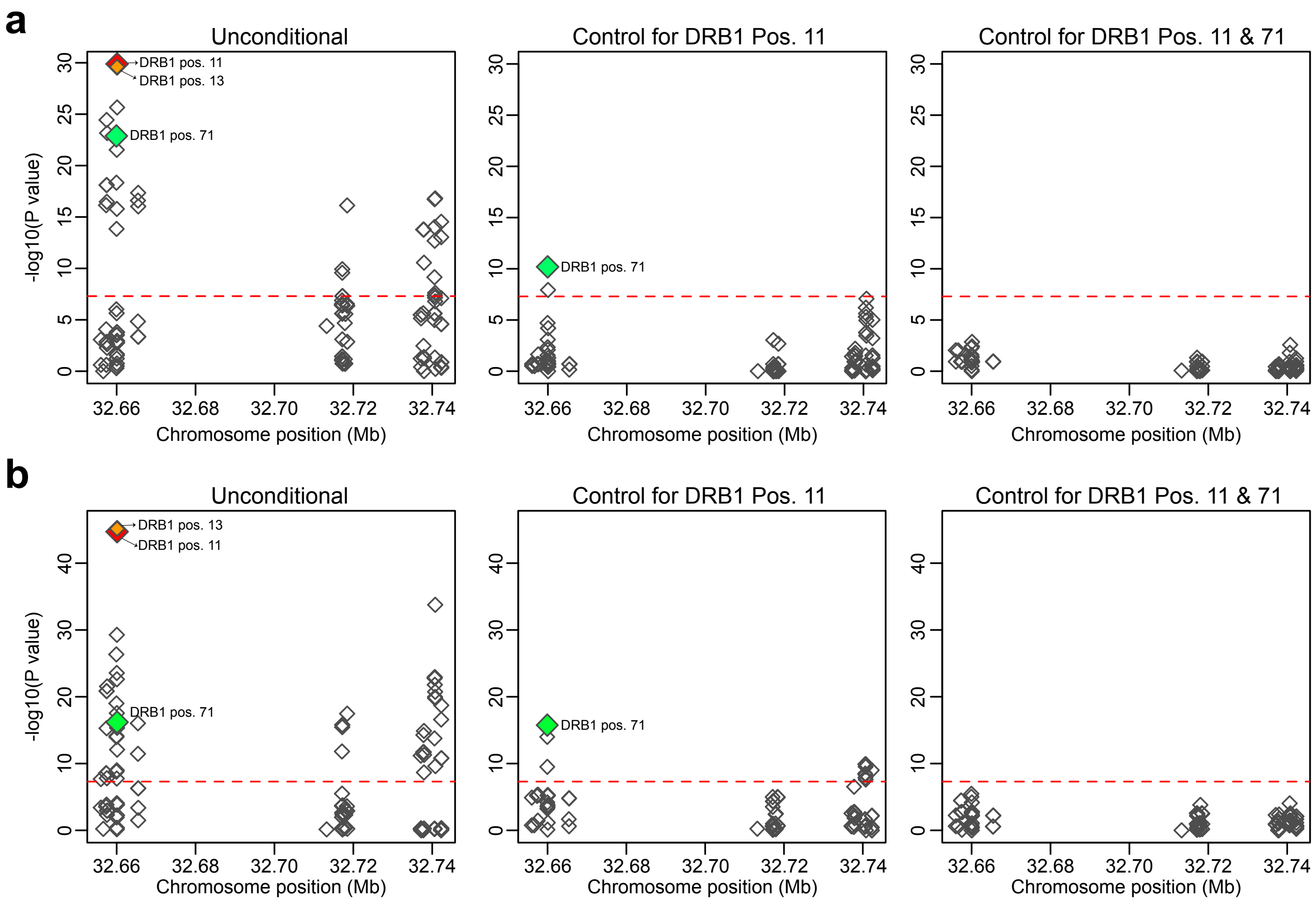


**Extended Data Fig. 5. Stepwise conditional analysis of imputed polymorphic amino-acid positions in DRβ1, DQβ1, and DQα1 peptides in (a) East Asian and (b) European cohorts.** Each symbol represents a polymorphic site tested for association with IgAN along the peptide sequence. Y-axis: genomic position of the sequence encoding each amino acid. X-axis: global statistical significance for each polymorphic site (multiallelic test with adjustment for cohort and principal components of ancestry).

**a**

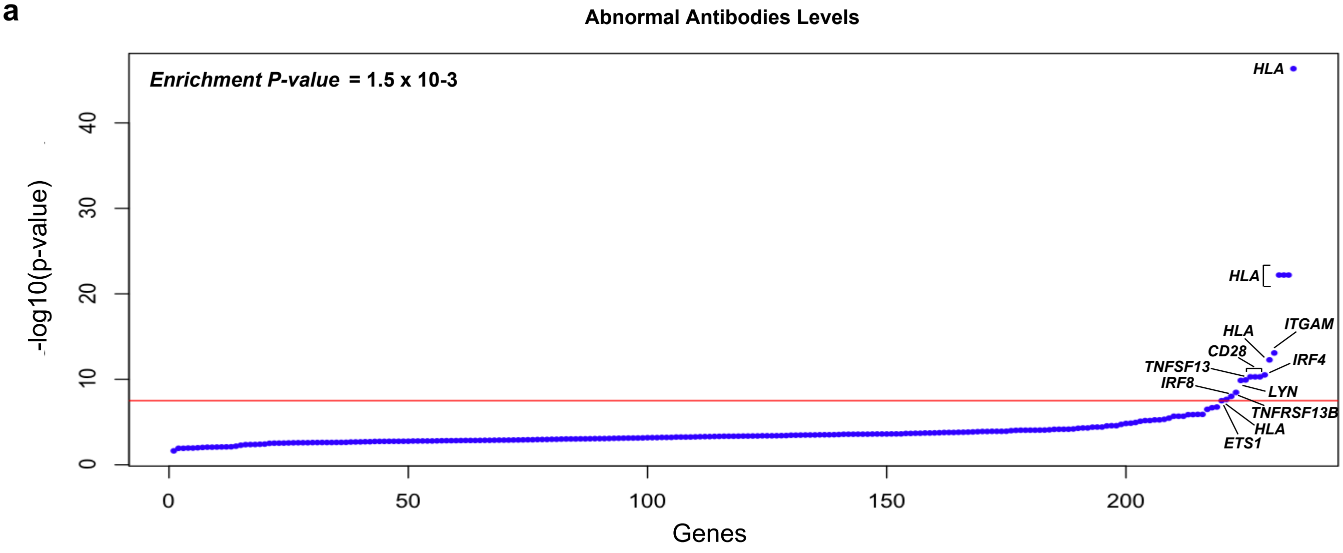


**b**

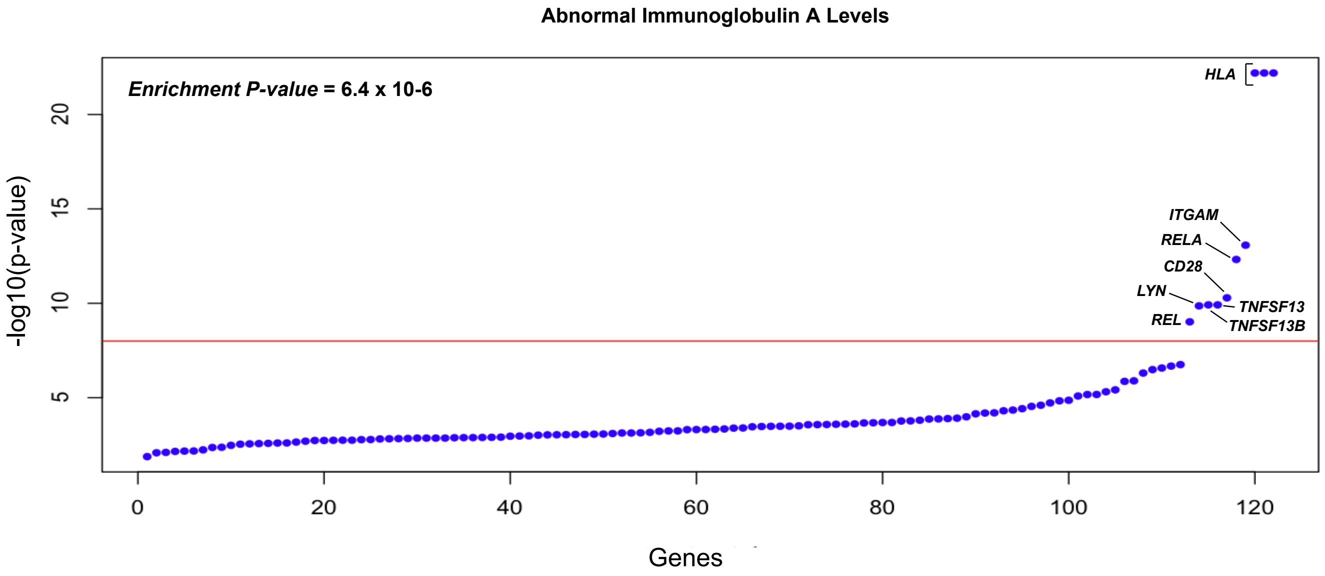


**Extended Data Fig. 6. Enrichment tests for the GWAS candidate gene set against human ortholog gene sets that when knocked out in mice result in (a)** “Abnormal Antibody Levels” and **(b)** “Abnormal Immunoglobulin A Levels”

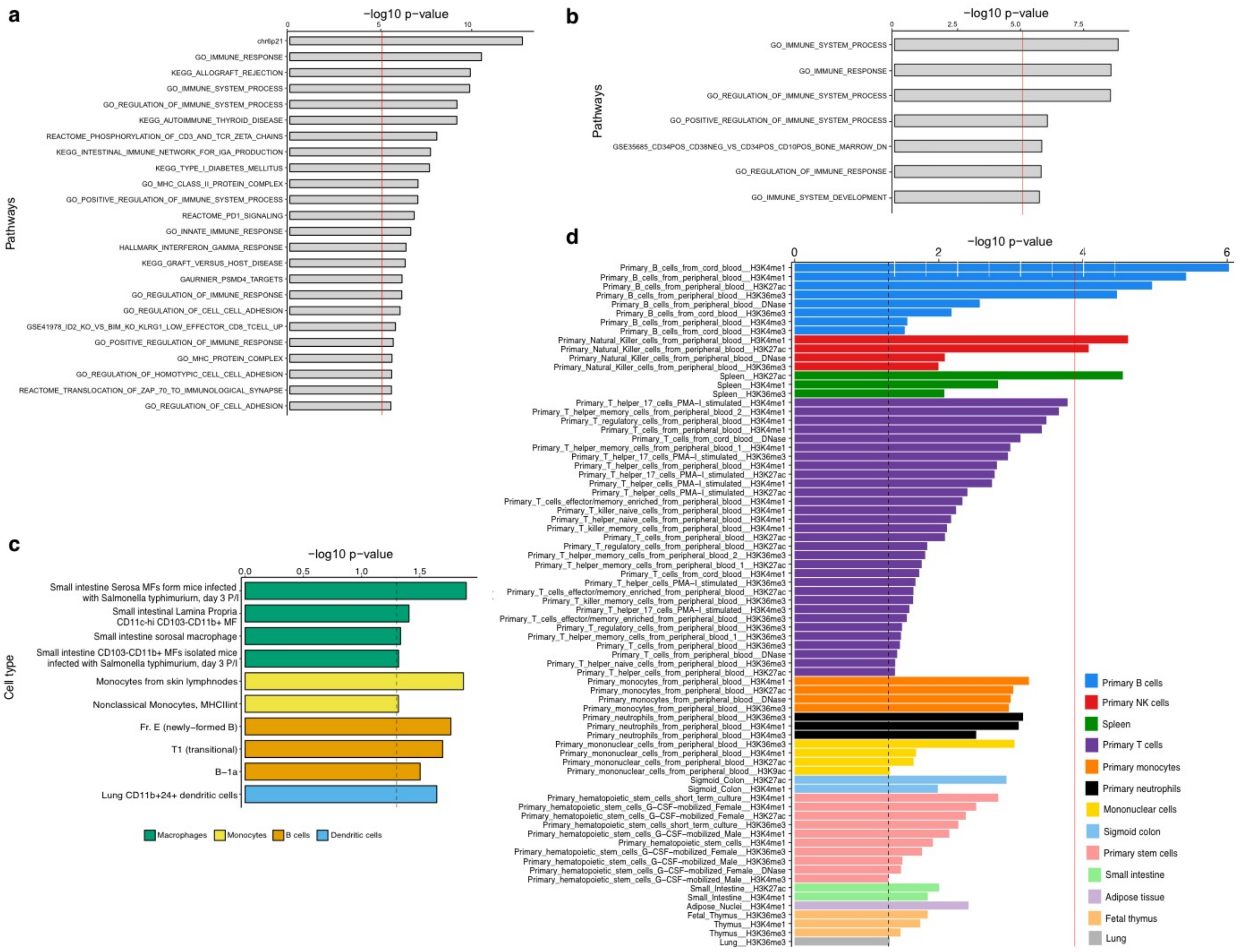


**Extended Data Fig. 7. Pathway, cell type and tissue enrichment analyses:** MAGMA pathway enrichment analysis based on GWAS summary statistics with **(a)** and without **(b)** the HLA region; -log10 P-value for the enrichment test are depicted along the x-axis, significant pathways are listed along the y-axis, red vertical lines indicate significance threshold corrected for multiple testing; **(c)** cell type-specific heritability enrichment for functional annotations in ImmGen dataset of mouse regulatory elements and expression data demonstrating the strongest enrichment in small intestine-derived macrophages profiled three days after Salmonella infection, and (**d**) Cell type-specific heritability enrichment for individual functional annotations generated by Roadmap Epigenomics demonstrates significant enrichment in immune cells, especially of B-cell lineage. All enrichments at nominal p-value<0.05 are displayed and grouped according to cell and tissue class. A solid red line represents a stringent Bonferroni-corrected -log10 of the p-value threshold of significance for the Roadmap dataset ($P$=1.3x10$^{-4}$). A dotted black line represents the -log10 of the nominal p-value of 0.05. GO: Gene Ontology; KEGG: Kyoto Encyclopedia of Genes and Genomes; MF: macrophages; P/I: post-injection.

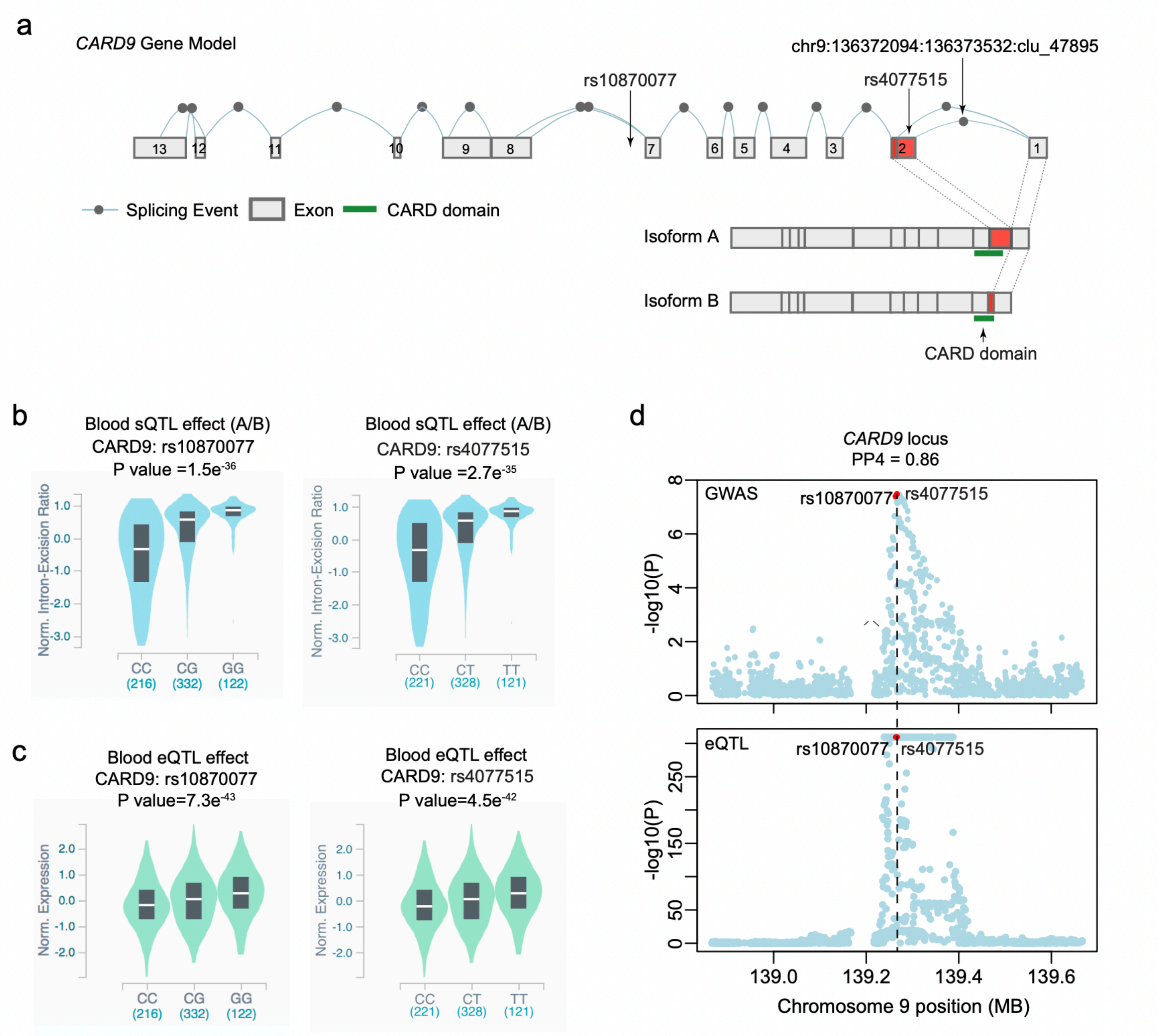


**Extended Data Fig. 8. Cis-regulatory effects at the *CARD9* locus**: **(a)** the *CARD9* gene model depicting 13 exons of *CARD9* and the splicing events from GTEx blood RNA-seq; the top IgAN risk allele (rs4077515-T) encodes S12N substitution in the second exon of *CARD9*; this SNP also exhibits a strong and statistically significant blood sQTL effect, wherein the risk allele is associated with higher rates of the chr9:136372094:136373532:clu_47895 splicing event, leading to the retention of exon 2 (red) in the coding sequence of CARD9 (isoform A), while the protective (rs4077515-C) allele is associated with the alternative splicing event that truncates exon 2 (isoform B); the functional CARD domain (green) maps to a portion of exon 2 that is intact in isoform A, but truncated in isoform B; **(b)** blood splice eQTL plots of normalized intron excision ratios (corresponding to the ratio of isoform A to isoform B) by the genotype of rs10870077 (top blood sQTL in GTEx) and rs4077515 (top SNP in GWAS for IgAN); these two SNPs are in near perfect linkage disequilibrium ($r^2$>0.98); **(c)** both rs10870077 and rs4077515 are also associated with a significant cis-eQTL effect on *CARD9* mRNA levels in GTEx blood; **(d)** the blood eQTL signal for CARD9 significantly co-localizes with the GWAS signal (PP4 of 0.86): top panel represents a regional plot for the GWAS signal (Immunochip data excluded); the bottom panel represents a blood cis-eQTL signal for CARD9 from the QTLGen Consortium meta-analysis (Y-axis truncated at P<1e-310); rs10870077 and rs4077515 are indicated in red.

**Table S1. Summary of study cohorts and genotyping platforms.**

| Cohort | Ethnicity | N Total | N Cases | N Controls | Genotyping Platform |
|---|---|---|---|---|---|
| French | European | 364 | 205 | 159 | Illumina 370-duo |
| Italian | European | 2385 | 1045 | 1340 | Illumina 1M-omni/1M-duo |
| North American | European | 1854 | 303 | 1551 | Illumina 550/660-quad |
| UK | European | 5243 | 464 | 4779 | Illumina HumanHap300/1M-duo |
| Turkish | European | 439 | 115 | 324 | Illumina Multi-Ethnic Global Array-8 |
| European I | European | 2456 | 920 | 1536 | Illumina Multi-Ethnic Global Array-8 |
| European II | European | 754 | 397 | 357 | Illumina Multi-Ethnic Global Array-8 |
| Argentinian | European | 194 | 100 | 94 | Illumina Multi-Ethnic Global Array-8 |
| Beijing I | East Asian | 2096 | 1194 | 902 | Illumina 610-quad |
| Shanghai | East Asian | 1345 | 656 | 689 | Illumina Multi-Ethnic Global Array-8 |
| Japanese | East Asian | 776 | 414 | 362 | Illumina Multi-Ethnic Global Array-8 |
| South Korean | East Asian | 1443 | 735 | 708 | Illumina Multi-Ethnic Global Array-8 |
| Beijing II | East Asian | 4151 | 479 | 3672 | Illumina Infinium OmniZhongHua-8 v1.3 |
| Beijing III | East Asian | 2352 | 1112 | 1240 | Illumina Infinium Global Screening Array-24 (GSA) |
| North Europe | European | 7624 | 527 | 7097 | Illumina HumanImmuno BeadChip (Immunochip) |
| Central-West Europe | European | 2202 | 966 | 1236 | Illumina HumanImmuno BeadChip (Immunochip) |
| South Europe | European | 3219 | 514 | 2705 | Illumina HumanImmuno BeadChip (Immunochip) |
| | Total | 38,897 | 10,146 | 28,751 | |

#### Table S2. Novel and known genome wide significant loci (p<5.0E-08)

| | | | | EUROPEAN | | | | EAST ASIAN | | | | COMBINED | | | | |
|---|---|---|---|---|---|---|---|---|---|---|---|---|---|---|---|---|
| **Chr** | **BP** | **SNP** | **Risk Allele** | **RAF Cases** | **RAF Controls** | **OR** | **P** | **RAF Cases** | **RAF Controls** | **OR** | **P** | **Meta OR** | **Meta P** | **Q test (P)** | **I2 (%)** | **Chip** |
| 1 | 196686918 | rs6677604 | G | 0.828 | 0.798 | 1.19 | 4.43E-12 | 0.956 | 0.934 | 1.28 | 1.77E-07 | 1.21 | 1.50E-17 | 0.966 | 0 | GWAS + IC |
| 1 | 157542162 | rs849815 | A | 0.690 | 0.663 | 1.13 | 3.36E-06 | 0.496 | 0.504 | 1.16 | 2.63E-04 | 1.14 | 3.91E-09 | 0.574 | 0 | GWAS + IC |
| 1 | 173146357 | rs4916312 | A | 0.371 | 0.355 | 1.15 | 5.14E-07 | 0.077 | 0.075 | 1.13 | 3.21E-02 | 1.14 | 5.00E-08 | 0.678 | 0 | GWAS + IC |
| 2 | 204584759 | rs3769684 | T | 0.951 | 0.946 | 1.26 | 2.69E-05 | 0.503 | 0.491 | 1.17 | 1.97E-07 | 1.19 | 5.14E-11 | 0.927 | 0 | GWAS + IC |
| 2 | 61092678 | rs842638 | T | 0.416 | 0.441 | 1.13 | 4.78E-05 | 0.120 | 0.149 | 1.23 | 1.44E-06 | 1.17 | 9.58E-10 | 0.002 | 63.8 | GWAS |
| 4 | 74725320 | rs6828610 | G | 0.169 | 0.156 | 1.26 | 3.62E-05 | 0.303 | 0.275 | 1.12 | 1.29E-04 | 1.14 | 3.54E-08 | 0.974 | 0 | GWAS |
| 6 | 249571 | rs12201499 | C | 0.116 | 0.119 | 1.12 | 2.11E-02 | 0.235 | 0.283 | 1.20 | 1.87E-10 | 1.18 | 3.07E-11 | 0.394 | 5.05 | GWAS |
| 6 | 7214676 | rs12530084 | C | 0.797 | 0.772 | 1.13 | 1.40E-04 | 0.535 | 0.513 | 1.13 | 2.29E-06 | 1.13 | 1.28E-09 | 0.488 | 0 | GWAS |
| 6 | 32389305 | rs9268557 | C | 0.6136 | 0.509 | 1.32 | 4.25E-38 | 0.626 | 0.566 | 1.21 | 2.72E-12 | 1.24 | 4.52E-47 | <0.0001 | 73.4 | GWAS + IC |
| 6 | 167445139 | rs2282859* | C | 0.017 | 0.014 | 1.02 | 8.33E-01 | 0.188 | 0.157 | 1.14 | 6.07E-07 | 1.17 | 2.68E-07 | 0.059 | 43.8 | GWAS+IC |
| 8 | 6808722 | rs2075836 | T | 0.284 | 0.311 | 1.17 | 4.04E-03 | 0.254 | 0.292 | 1.22 | 3.11E-09 | 1.21 | 5.85E-11 | 0.898 | 0 | GWAS |
| 8 | 56852496 | rs75413466 | A | 0.025 | 0.019 | 1.52 | 1.52E-06 | 0.069 | 0.059 | 1.34 | 1.06E-05 | 1.40 | 1.37E-10 | 0.797 | 0 | GWAS + IC |
| 8 | 124765474 | rs34354351 | T | 0.159 | 0.173 | 1.20 | 2.44E-05 | 0.301 | 0.319 | 1.13 | 1.90E-04 | 1.15 | 3.51E-08 | 0.838 | 0 | GWAS |
| 9 | 139266496 | rs4077515 | T | 0.437 | 0.413 | 1.17 | 2.46E-10 | 0.311 | 0.290 | 1.09 | 7.05E-03 | 1.14 | 2.65E-11 | 0.415 | 1.83 | GWAS + IC |
| 9 | 117643362 | rs13300483 | T | 0.262 | 0.245 | 1.14 | 9.74E-06 | 0.331 | 0.307 | 1.13 | 3.34E-04 | 1.13 | 1.27E-08 | 0.877 | 0 | GWAS + IC |
| 10 | 81043743 | rs1108618 | A | 0.643 | 0.599 | 1.13 | 7.22E-06 | 0.488 | 0.495 | 1.15 | 5.67E-06 | 1.14 | 1.93E-10 | 0.435 | 0.32 | GWAS + IC |
| 10 | 65363048 | rs57917667 | G | 0.018 | 0.020 | 1.18 | 2.83E-01 | 0.161 | 0.191 | 1.23 | 1.88E-08 | 1.22 | 1.09E-08 | 0.910 | 0 | GWAS |
| 11 | 65555524 | rs10896045 | A | 0.322 | 0.299 | 1.12 | 1.52E-03 | 0.473 | 0.485 | 1.23 | 1.16E-11 | 1.18 | 4.77E-13 | 0.033 | 50.6 | GWAS |
| 11 | 128487069 | rs7121743 | C | 0.172 | 0.156 | 1.11 | 1.27E-02 | 0.491 | 0.474 | 1.14 | 7.03E-07 | 1.13 | 3.40E-08 | 0.790 | 0 | GWAS |
| 14 | 107222014 | rs751081288 | A | 0.463 | 0.433 | 1.13 | 2.52E-03 | 0.601 | 0.565 | 1.20 | 1.31E-06 | 1.17 | 1.94E-08 | 0.743 | 0 | GWAS |
| 16 | 31357760 | rs11150612 | A | 0.599 | 0.645 | 1.15 | 4.00E-08 | 0.233 | 0.270 | 1.20 | 2.32E-07 | 1.16 | 8.37E-14 | 0.105 | 36.8 | GWAS+IC |
| 16 | 86017715 | rs1879210 | T | 0.668 | 0.637 | 1.12 | 2.51E-05 | 0.876 | 0.856 | 1.19 | 5.06E-05 | 1.14 | 9.92E-09 | 0.963 | 0 | GWAS + IC |
| 17 | 7462969 | rs3803800 | A | 0.221 | 0.210 | 1.09 | 4.94E-03 | 0.364 | 0.324 | 1.22 | 1.43E-10 | 1.15 | 1.21E-10 | 0.186 | 27.1 | GWAS + IC |
| 17 | 16851450 | rs57382045 | A | 0.146 | 0.113 | 1.24 | 1.93E-06 | 0.355 | 0.326 | 1.13 | 9.52E-05 | 1.16 | 3.45E-09 | 0.842 | 0 | GWAS |
| 19 | 55397217 | rs1865097 | A | 0.335 | 0.301 | 1.10 | 3.21E-04 | 0.412 | 0.380 | 1.15 | 2.65E-06 | 1.12 | 7.74E-09 | 0.493 | 0 | GWAS + IC |
| 22 | 30512478 | rs48230748 | G | 0.549 | 0.538 | 1.10 | 4.29E-04 | 0.709 | 0.669 | 1.22 | 3.18E-14 | 1.16 | 7.76E-15 | 0.506 | 0 | GWAS |

*** East Asian-specific locus**

#### Table S3. Suggestive loci in the combined trans-ethnic meta-analysis (p<5.0E-05)

| | | | | EUROPEAN | | | | EAST ASIAN | | | | COMBINED | | | | |
|---|---|---|---|---|---|---|---|---|---|---|---|---|---|---|---|---|
| CHR | BP | SNP | Risk Allele | RAF Cases | RAF Controls | OR | P | RAF Cases | RAF Controls | OR | P | Meta OR | Meta P | Q test (P) | I2 (%) | Chip |
| 1 | 15613946 | rs531901 | A | 0.492 | 0.459 | 1.16 | 1.14E-04 | 0.666 | 0.650 | 1.14 | 1.07E-02 | 1.153 | 3.76E-06 | 0.733 | 0 | GWAS |
| 1 | 161197244 | rs4233368 | A | 0.251 | 0.254 | 1.07 | 1.45E-02 | 0.446 | 0.412 | 1.15 | 7.21E-06 | 1.106 | 1.28E-06 | 0.147 | 31.54 | GWAS+IC |
| 1 | 161470505 | rs12139150 | G | 0.593 | 0.532 | 1.09 | 2.12E-04 | 0.789 | 0.773 | 1.10 | 1.75E-03 | 1.093 | 1.37E-06 | 0.818 | 0 | GWAS+IC |
| 1 | 17630193 | rs2501784 | A | 0.449 | 0.420 | 1.10 | 1.28E-04 | 0.650 | 0.628 | 1.12 | 2.36E-04 | 1.107 | 1.22E-07 | 0.019 | 53.11 | GWAS+IC |
| 1 | 38379770 | rs4653337 | G | 0.707 | 0.678 | 1.13 | 5.79E-04 | 0.722 | 0.686 | 1.11 | 3.67E-03 | 1.117 | 7.39E-06 | 0.856 | 0 | GWAS |
| 1 | 68720758 | rs10493441 | A | 0.095 | 0.090 | 1.13 | 2.22E-02 | 0.266 | 0.250 | 1.16 | 8.62E-06 | 1.154 | 6.29E-07 | 0.317 | 13.31 | GWAS |
| 1 | 85743741 | rs2735592 | G | 0.324 | 0.294 | 1.12 | 1.55E-05 | 0.289 | 0.291 | 1.09 | 2.22E-02 | 1.111 | 1.35E-06 | 0.863 | 0 | GWAS+IC |
| 2 | 167244822 | rs16852104 | A | 0.142 | 0.119 | 1.17 | 5.30E-03 | 0.269 | 0.245 | 1.15 | 2.25E-04 | 1.155 | 3.95E-06 | 0.326 | 13.64 | GWAS |
| 2 | 201739327 | rs4035021 | A | 0.389 | 0.360 | 1.13 | 5.40E-05 | 0.577 | 0.549 | 1.08 | 2.07E-02 | 1.110 | 5.68E-06 | 0.387 | 5.94 | GWAS |
| 2 | 218991005 | rs4674259 | G | 0.456 | 0.459 | 1.07 | 1.66E-03 | 0.653 | 0.634 | 1.10 | 4.79E-04 | 1.085 | 3.73E-06 | 0.036 | 48.25 | GWAS+IC |
| 2 | 234115739 | rs14243 | G | 0.245 | 0.210 | 1.16 | 6.53E-06 | 0.365 | 0.353 | 1.10 | 3.07E-03 | 1.127 | 1.70E-07 | 0.139 | 34.83 | GWAS |
| 2 | 43352888 | rs737013 | T | 0.402 | 0.373 | 1.13 | 3.26E-05 | 0.249 | 0.233 | 1.07 | 3.96E-02 | 1.100 | 8.86E-06 | 0.408 | 3.6 | GWAS |
| 2 | 62551472 | rs10865331 | A | 0.433 | 0.393 | 1.12 | 7.09E-06 | 0.382 | 0.368 | 1.07 | 3.19E-02 | 1.097 | 1.29E-06 | 0.458 | 0 | GWAS+IC |
| 3 | 101663386 | rs7625614 | T | 0.355 | 0.359 | 1.11 | 1.44E-03 | 0.667 | 0.653 | 1.11 | 1.22E-03 | 1.108 | 5.64E-06 | 0.913 | 0 | GWAS |
| 3 | 29361792 | rs55711830 | A | 0.375 | 0.347 | 1.16 | 4.65E-05 | 0.108 | 0.101 | 1.11 | 5.23E-02 | 1.143 | 7.77E-06 | 0.106 | 37.87 | GWAS |
| 3 | 39249753 | rs12497322 | G | 0.681 | 0.655 | 1.12 | 1.53E-02 | 0.524 | 0.489 | 1.11 | 1.82E-04 | 1.109 | 8.58E-06 | 0.008 | 79.18 | GWAS |
| 3 | 77447376 | rs28378063 | G | 0.355 | 0.336 | 1.11 | 2.36E-04 | 0.475 | 0.453 | 1.08 | 4.94E-03 | 1.096 | 5.20E-06 | 0.828 | 0 | GWAS |
| 4 | 2841240 | rs4690002 | T | 0.389 | 0.367 | 1.16 | 3.13E-03 | 0.484 | 0.448 | 1.16 | 1.99E-05 | 1.163 | 2.12E-07 | 0.834 | 0 | GWAS |
| 5 | 133455961 | rs151822 | G | 0.258 | 0.229 | 1.08 | 3.09E-02 | 0.512 | 0.473 | 1.14 | 2.61E-05 | 1.110 | 7.14E-06 | 0.417 | 2.06 | GWAS |
| 5 | 158748679 | rs3213097 | T | 0.249 | 0.228 | 1.10 | 1.24E-03 | 0.472 | 0.448 | 1.09 | 1.73E-03 | 1.092 | 6.88E-06 | 0.279 | 17.84 | GWAS+IC |
| 5 | 35857850 | rs10213865 | A | 0.767 | 0.741 | 1.11 | 2.60E-04 | 0.806 | 0.786 | 1.10 | 8.14E-03 | 1.108 | 6.57E-06 | 0.393 | 5.29 | GWAS+IC |
| 5 | 38867732 | rs395157 | T | 0.490 | 0.471 | 1.14 | 1.32E-06 | 0.253 | 0.247 | 1.06 | 9.95E-02 | 1.108 | 1.55E-06 | 0.527 | 0 | GWAS+IC |
| 5 | 40463739 | rs4957300 | C | 0.644 | 0.658 | 1.11 | 1.80E-05 | 0.306 | 0.298 | 1.07 | 2.89E-02 | 1.091 | 2.52E-06 | 0.629 | 0 | GWAS+IC |
| 5 | 55438851 | rs10065637 | C | 0.812 | 0.778 | 1.16 | 7.78E-08 | 0.967 | 0.978 | 0.53 | 2.26E-01 | 1.155 | 1.77E-07 | 0.198 | 30.14 | GWAS+IC |
| 5 | 79564517 | rs62364037 | A | 0.171 | 0.153 | 1.22 | 1.85E-06 | 0.000 | 0.000 | NA | NA | 1.219 | 1.85E-06 | 0.797 | 0 | GWAS |
| 6 | 138132123 | rs58905141 | G | 0.036 | 0.031 | 1.31 | 6.43E-07 | 0.048 | 0.039 | 1.20 | 1.44E-01 | 1.293 | 3.26E-07 | 0.954 | 0 | GWAS |
| 6 | 159499748 | rs2485360 | T | 0.297 | 0.286 | 1.11 | 5.20E-04 | 0.063 | 0.047 | 1.25 | 1.09E-03 | 1.126 | 7.77E-06 | 0.588 | 0 | GWAS+IC |
| 6 | 167445139 | rs2282859 | C | 0.017 | 0.014 | 1.02 | 8.33E-01 | 0.179 | 0.151 | 1.20 | 3.88E-08 | 1.172 | 2.68E-07 | 0.059 | 43.76 | GWAS+IC |
| 6 | 168048619 | rs111387965 | G | 0.050 | 0.040 | 1.21 | 5.46E-02 | 0.087 | 0.061 | 1.33 | 2.72E-05 | 1.286 | 7.67E-06 | 0.665 | 0 | GWAS |
| 6 | 5675845 | rs2142738 | T | 0.485 | 0.464 | 1.13 | 6.28E-05 | 0.710 | 0.686 | 1.10 | 1.38E-02 | 1.118 | 3.34E-06 | 0.039 | 47.55 | GWAS |
| 7 | 11625652 | rs17574506 | C | 0.391 | 0.439 | 1.13 | 4.60E-04 | 0.564 | 0.513 | 1.14 | 7.22E-05 | 1.139 | 1.24E-07 | 0.706 | 0 | GWAS |
| 7 | 158600277 | rs710423 | T | 0.175 | 0.163 | 1.09 | 4.43E-02 | 0.327 | 0.296 | 1.17 | 9.12E-07 | 1.137 | 3.07E-07 | 0.419 | 2.45 | GWAS |
| 7 | 50476778 | rs79441475 | A | 0.089 | 0.078 | 1.23 | 1.66E-06 | <0.001 | <0.001 | NA | NA | 1.228 | 1.66E-06 | 0.548 | 0 | GWAS+IC |
| 8 | 128226195 | rs6989575 | T | 0.320 | 0.287 | 1.12 | 8.63E-04 | 0.473 | 0.459 | 1.10 | 8.03E-04 | 1.111 | 2.40E-06 | 0.483 | 0 | GWAS |
| 8 | 18848034 | rs2410596 | C | 0.866 | 0.852 | 1.09 | 3.47E-02 | 0.328 | 0.349 | 1.11 | 8.76E-05 | 1.107 | 9.65E-06 | 0.148 | 31.44 | GWAS |
| 8 | 61022672 | rs2611364 | T | 0.628 | 0.610 | 1.14 | 2.21E-04 | 0.487 | 0.466 | 1.12 | 1.32E-04 | 1.131 | 1.17E-07 | 0.489 | 0 | GWAS |
| 8 | 81128822 | rs9774752 | C | 0.385 | 0.354 | 1.13 | 9.47E-03 | 0.437 | 0.474 | 1.10 | 3.03E-04 | 1.110 | 9.57E-06 | 0.008 | 79.21 | GWAS |
| 9 | 92078518 | rs4877094 | C | 0.194 | 0.174 | 1.08 | 4.69E-02 | 0.364 | 0.338 | 1.14 | 1.48E-05 | 1.121 | 3.51E-06 | 0.570 | 0 | GWAS |
| 10 | 3918174 | rs10904115 | T | 0.507 | 0.493 | 1.08 | 1.15E-02 | 0.471 | 0.438 | 1.14 | 8.97E-06 | 1.111 | 7.37E-07 | 0.441 | 0 | GWAS |
| 10 | 67783437 | rs10509258 | G | 0.331 | 0.307 | 1.10 | 1.24E-03 | 0.261 | 0.244 | 1.10 | 1.65E-03 | 1.102 | 6.52E-06 | 0.117 | 37.74 | GWAS |
| 11 | 64275670 | rs34429336 | C | 0.446 | 0.436 | 1.09 | 1.49E-03 | 0.348 | 0.323 | 1.10 | 1.16E-03 | 1.095 | 5.54E-06 | 0.081 | 40.13 | GWAS |
| 12 | 110666136 | rs56838584 | A | 0.913 | 0.900 | 1.27 | 9.24E-06 | 0.990 | 0.982 | 0.00 | 1.29E-01 | 1.274 | 4.68E-06 | 0.494 | 0 | GWAS |
| 16 | 72163694 | rs10492815 | A | 0.949 | 0.956 | 1.05 | 6.42E-01 | 0.756 | 0.712 | 1.20 | 6.41E-07 | 1.177 | 3.91E-06 | 0.814 | 0 | GWAS |
| 17 | 40565916 | rs72823055 | T | 0.106 | 0.091 | 1.19 | 4.26E-05 | 0.226 | 0.217 | 1.09 | 1.43E-02 | 1.133 | 6.79E-06 | 0.528 | 0 | GWAS+IC |
| 17 | 8038789 | rs9913189 | A | 0.625 | 0.588 | 1.16 | 4.12E-05 | 0.882 | 0.879 | 1.12 | 7.63E-02 | 1.147 | 8.82E-06 | 0.634 | 0 | GWAS |
| 19 | 38687076 | rs62123462 | A | 0.420 | 0.398 | 1.11 | 5.62E-03 | 0.476 | 0.401 | 1.17 | 1.63E-04 | 1.134 | 4.85E-06 | 0.057 | 48.89 | GWAS |
| 19 | 45487178 | rs204474 | C | 0.387 | 0.367 | 1.10 | 5.03E-03 | 0.579 | 0.557 | 1.11 | 1.21E-04 | 1.105 | 2.10E-06 | 0.668 | 0 | GWAS |
| 22 | 29957911 | rs469295 | T | 0.258 | 0.235 | 1.09 | 2.27E-03 | 0.603 | 0.577 | 1.13 | 4.55E-04 | 1.104 | 4.59E-06 | 0.736 | 0 | GWAS+IC |

**Table S4. Ancestry-specific suggestive loci (p<5.0E-05): (a) East Asians, (b) Europeans.**

| (a) | | | | EUROPEAN | | | | EAST ASIAN | | | | COMBINED | | | | |
|---|---|---|---|---|---|---|---|---|---|---|---|---|---|---|---|---|
| **CHR** | **BP** | **SNP** | **Risk Allele** | **RAF Cases** | **RAF Controls** | **OR** | **P** | **RAF Cases** | **RAF Controls** | **OR** | **P** | **OR** | **P** | **Q test (P)** | **I2 (%)** | **Chip** |
| 1 | 11937404 | rs7554327 | C | 0.513 | 0.528 | 1.03 | 2.05E-01 | 0.546 | 0.516 | 1.14 | 5.46E-07 | 1.04 | 2.60E-02 | 0.125 | 34.15 | GWAS+IC |
| 8 | 103543524 | rs10088534 | C | 0.627 | 0.631 | 1.02 | 5.97E-01 | 0.509 | 0.474 | 1.14 | 3.25E-07 | 1.08 | 4.18E-05 | 0.038 | 47.98 | GWAS |
| 9 | 37023483 | rs77134091 | C | 0.000 | 0.000 | NA | NA | 0.923 | 0.903 | 1.21 | 8.15E-06 | 1.21 | 8.15E-06 | 0.226 | 32.72 | GWAS EAS |
| 13 | 70019147 | rs79246816 | T | 0.897 | 0.902 | 1.06 | 2.60E-01 | 0.837 | 0.813 | 1.17 | 2.97E-06 | 1.13 | 1.25E-05 | 0.477 | 0 | GWAS |
| 14 | 21087181 | rs1652035 | A | 0.493 | 0.496 | 1.00 | 9.21E-01 | 0.677 | 0.645 | 1.18 | 5.37E-06 | 1.10 | 4.63E-04 | 0.009 | 65.17 | GWAS |
| 15 | 60913415 | rs339993 | A | NA | NA | NA | NA | 0.494 | 0.497 | 1.16 | 3.65E-06 | 1.16 | 3.65E-06 | 0.888 | 0 | GWAS EAS |
| 15 | 74869438 | rs8041357 | C | 0.953 | 0.961 | 1.04 | 5.73E-01 | 0.232 | 0.207 | 1.14 | 8.80E-06 | 1.13 | 1.92E-05 | 0.074 | 41.27 | GWAS |
| 18 | 72833866 | rs28694297 | T | 0.877 | 0.885 | 1.05 | 3.08E-01 | 0.793 | 0.444 | 1.14 | 9.33E-06 | 1.08 | 2.08E-03 | 0.077 | 40.75 | GWAS |
| 18 | 74760875 | rs7227084 | T | 0.753 | 0.739 | 1.03 | 3.88E-01 | 0.846 | 0.825 | 1.17 | 9.79E-06 | 1.10 | 2.21E-04 | 0.049 | 47.1 | GWAS |
| 19 | 52151778 | rs2902877 | A | 0.439 | 0.438 | 1.00 | 9.47E-01 | 0.686 | 0.663 | 1.16 | 3.59E-06 | 1.09 | 4.77E-04 | 0.163 | 30.78 | GWAS |
| 22 | 48968790 | rs910580 | C | 0.234 | 0.240 | 1.01 | 7.59E-01 | 0.448 | 0.401 | 1.18 | 1.61E-06 | 1.11 | 1.11E-04 | 0.032 | 52.37 | GWAS |

| (b) | | | | EUROPEAN | | | | EAST ASIAN | | | | COMBINED | | | | |
|---|---|---|---|---|---|---|---|---|---|---|---|---|---|---|---|---|
| **CHR** | **BP** | **SNP** | **Risk Allele** | **RAF Cases** | **RAF Controls** | **OR** | **P** | **RAF Cases** | **RAF Controls** | **OR** | **P** | **OR** | **P** | **Q test (P)** | **I2 (%)** | **Chip** |
| 1 | 106622787 | rs61814213 | A | 0.958 | 0.938 | 1.70 | 6.14E-06 | 0.723 | 0.718 | 1.03 | 4.56E-01 | 1.06 | 5.11E-02 | 0.840 | 0 | GWAS |
| 1 | 206656292 | rs10863389 | C | 0.650 | 0.620 | 1.11 | 2.43E-06 | 0.371 | 0.368 | 1.04 | 2.49E-01 | 1.05 | 4.42E-03 | 0.527 | 0 | GWAS+IC |
| 2 | 100801198 | rs11677053 | C | 0.423 | 0.403 | 1.11 | 7.29E-06 | 0.462 | 0.459 | 1.01 | 8.35E-01 | 1.07 | 2.98E-04 | 0.238 | 21.53 | GWAS+IC |
| 2 | 2516093 | rs79067042 | G | 0.188 | 0.176 | 1.21 | 5.54E-06 | <0.001 | <0.001 | NA | NA | 1.21 | 5.54E-06 | 0.238 | 26.16 | GWAS |
| 2 | 34183606 | rs12611527 | A | 0.678 | 0.639 | 1.22 | 4.34E-07 | 0.368 | 0.341 | 1.07 | 2.62E-02 | 1.04 | 1.08E-01 | 0.000 | 81.92 | GWAS |
| 2 | 595891 | rs35662997 | G | 0.124 | 0.094 | 1.33 | 1.10E-06 | 0.040 | 0.040 | 1.05 | 5.71E-01 | 1.21 | 7.27E-05 | 0.078 | 45.21 | GWAS |
| 4 | 103475444 | rs4648011 | T | 0.655 | 0.604 | 1.14 | 6.27E-07 | 0.489 | 0.501 | 1.01 | 7.89E-01 | 1.08 | 7.14E-05 | 0.002 | 63.71 | GWAS+IC |
| 5 | 71843755 | rs381734 | G | 0.788 | 0.757 | 1.16 | 1.06E-06 | 0.760 | 0.758 | 1.02 | 5.47E-01 | 1.09 | 1.00E-04 | 0.504 | 0 | GWAS+IC |
| 5 | 96370879 | rs248215 | G | 0.490 | 0.477 | 1.14 | 1.63E-06 | 0.536 | 0.535 | 1.01 | 7.09E-01 | 1.07 | 2.99E-04 | 0.057 | 44.15 | GWAS |
| 7 | 120550667 | rs2040762 | T | 0.268 | 0.239 | 1.27 | 5.37E-07 | 0.381 | 0.384 | 0.99 | 6.76E-01 | 1.06 | 2.02E-02 | 0.016 | 57.31 | GWAS |
| 7 | 36707837 | rs73094969 | G | 0.176 | 0.168 | 1.17 | 8.42E-06 | 0.106 | 0.109 | 1.00 | 9.79E-01 | 1.10 | 8.36E-04 | 0.097 | 37.81 | GWAS |
| 8 | 131361477 | rs7018225 | T | 0.570 | 0.550 | 1.17 | 1.50E-06 | 0.336 | 0.337 | 0.97 | 4.00E-01 | 1.10 | 6.46E-05 | 0.410 | 3.37 | GWAS |
| 11 | 64097233 | rs694739 | A | 0.655 | 0.626 | 1.13 | 1.61E-06 | 0.830 | 0.840 | 1.05 | 2.15E-01 | 1.07 | 8.43E-04 | 0.617 | 0 | GWAS+IC |
| 12 | 28850736 | rs10843237 | T | 0.694 | 0.640 | 1.17 | 2.52E-06 | 0.692 | 0.679 | 1.03 | 4.18E-01 | 1.09 | 1.34E-04 | 0.070 | 41.85 | GWAS |
| 16 | 9677083 | rs6498785 | A | 0.584 | 0.541 | 1.15 | 9.94E-06 | 0.501 | 0.505 | 1.04 | 1.76E-01 | 1.04 | 4.23E-02 | 0.226 | 22.82 | GWAS |
| 18 | 11971994 | rs9303756 | G | 0.166 | 0.157 | 1.18 | 2.90E-06 | 0.171 | 0.165 | 1.00 | 9.52E-01 | 1.09 | 1.08E-03 | 0.001 | 67.65 | GWAS |
| 18 | 12737037 | rs2847247 | A | 0.408 | 0.369 | 1.18 | 2.42E-06 | 0.264 | 0.267 | 1.01 | 7.56E-01 | 1.10 | 5.40E-04 | 0.747 | 0 | GWAS |
| 18 | 74983830 | rs75152619 | C | 0.041 | 0.027 | 1.39 | 9.88E-07 | <0.001 | <0.001 | NA | NA | 1.39 | 9.88E-07 | 0.344 | 11.25 | GWAS |
| 19 | 3263999 | rs66654893 | T | 0.481 | 0.432 | 1.25 | 9.14E-06 | 0.262 | 0.271 | 1.05 | 2.53E-01 | 1.08 | 2.94E-02 | 0.151 | 43.45 | GWAS |

##### Table S5. Stepwise conditional analysis to define independent SNPs within each genome-wide significant locus (p<5.0E-08)

| | | | | Unconditional Analysis | | Conditional Analysis | |
|---|---|---|---|---|---|---|---|
| **CHR:BP** | **SNP** | **GENE** | **Risk Allele** | **OR** | **P-value** | **OR** | **P-value** |
| 1:196686918 | rs6677604 | *CFH* | G | 1.21 | 1.50E-17 | | |
| 1:196603302 | rs12029571 | | A | 1.12 | 2.54E-06 | 1.15 | 3.60E-09 |
| 1:196674330 | rs567086291 | | A | 1.26 | 3.19E-10 | 1.22 | 5.70E-08 |
| 1:157542162 | rs849815 | *FCRL3, FCRL4* | A | 1.14 | 3.91E-09 | | |
| 1:157502863 | rs73011558 | | T | 1.21 | 9.37E-05 | 1.21 | 1.02E-04 |
| 1:173146357 | rs4916312 | *TNFSF4* | A | 1.14 | 5.00E-08 | | |
| 1:172903845 | rs12565596 | | A | 1.07 | 3.71E-03 | 1.10 | 9.58E-05 |
| 2:204584759 | rs3769684 | *CD28* | T | 1.19 | 5.14E-11 | | |
| 2:204722752 | rs926169 | | G | 1.09 | 4.74E-06 | 1.09 | 6.28E−5 |
| 2:61092678 | rs842638 | *REL* | T | 1.17 | 9.58E-10 | | |
| 2:61151983 | rs78037521 | | TCA | 1.23 | 3.04E-07 | 1.18 | 1.06E-04 |
| 4:74725320 | rs6828610 | *PF4V1* | G | 1.14 | 3.54E-08 | | |
| 4:75044689 | rs60069701 | | G | 1.11 | 1.96E-03 | 1.12 | 6.65E-04 |
| 6:249571 | rs12201499 | *IRF4* | C | 1.18 | 3.07E-11 | | |
| 6:428486 | rs12526822 | | A | 1.12 | 1.52E-04 | 1.11 | 2.50E-04 |
| 6:7214676 | rs12530084 | *LY86* | C | 1.13 | 1.28E-09 | | |
| 6:7328454 | rs79358471 | | T | 1.86 | 5.38E-04 | 1.81 | 9.45E-04 |
| 8:6808722 | rs2075836 | *DEFA1* | T | 1.21 | 5.85E-11 | | |
| 8:6766505 | rs34087549 | | A | 1.11 | 6.13E-05 | 1.11 | 3.72E-05 |
| 8:56852496 | rs75413466 | *LYN* | A | 1.40 | 1.37E-10 | | |
| 8:56802069 | rs333619 | | T | 1.06 | 3.02E-03 | 1.07 | 1.65E-04 |
| 8:124765474 | rs34354351 | *ANXA13* | T | 1.15 | 3.51E-08 | | |
| 8:124713994 | rs76705386 | | C | 1.22 | 2.35E-03 | 1.21 | 3.50E-03 |
| 9:139266496 | rs4077515 | *CARD9* | T | 1.14 | 2.65E-11 | | |
| 9:139393067 | rs7852557 | | C | 1.09 | 2.51E-04 | 1.07 | 7.09E-03 |
| 9:117643362 | rs13300483 | *TNFSF8, TNFSF15* | T | 1.13 | 1.27E-08 | | |
| 9:117604858 | rs540928691 | | CA | 1.10 | 2.91E-04 | 1.08 | 2.70E-03 |
| 10:81043743 | rs1108618 | *ZMIZ1, PPIF* | A | 1.14 | 1.93E-10 | | |
| 10:81006391 | rs117464630 | | T | 1.20 | 5.34E-04 | 1.20 | 3.50E-04 |
| 10:65363048 | rs57917667 | *REEP3* | G | 1.22 | 1.09E-08 | | |
| 10:65752896 | rs77436363 | | C | 2.03 | 3.12E-04 | 2.08 | 1.97E-04 |
| 11:65555524 | rs10896045 | *RELA* | A | 1.18 | 4.77E-13 | | |
| 11:65562257 | rs2276133 | | G | 1.04 | 5.03E-02 | 1.09 | 5.30E-05 |
| 11:128487069 | rs7121743 | *ETS1* | C | 1.13 | 3.40E-08 | | |
| 11:127683477 | rs7945213 | | C | 1.08 | 1.05E-03 | 1.11 | 2.01E-05 |
| 14:107106655 | rs746398 | *IGH* | A | 1.17 | 1.94E-08 | | |
| 14:107031982 | rs2731189 | | G | 1.04 | 3.91E-01 | 1.06 | 4.21E-03 |
| 16:31357760 | rs11150612 | *ITGAM, ITGAX* | A | 1.16 | 8.37E-14 | | |
| 16:31362711 | rs67898294 | | C | 1.19 | 2.88E-10 | 1.13 | 3.23E-05 |
| 16:86017715 | rs1879210 | *IRF8* | T | 1.14 | 9.92E-09 | | |
| 16:86485321 | rs6540296 | | G | 1.10 | 3.13E-05 | 1.00 | 4.87E-06 |
| 17:7462969 | rs3803800 | *TNFSF13* | A | 1.15 | 1.21E-10 | | |
| 17:7521764 | rs72829406 | | T | 1.08 | 2.63E-03 | 1.10 | 2.83E-04 |
| 17:16851450 | rs57382045 | *TNFRSF13B* | A | 1.16 | 3.45E-09 | | |
| 17:16883047 | rs201993221 | | AT | 1.14 | 3.62E-03 | 1.16 | 8.29E-04 |
| 19:55397217 | rs1865097 | *FCAR* | A | 1.12 | 7.74E-09 | | |
| 19:55177929 | rs760186 | | A | 1.18 | 5.89E-05 | 1.17 | 9.47E-05 |
| 22:30512478 | rs4823074 | *LIF* | G | 1.16 | 7.76E-15 | | |
| 22:30037168 | rs564157878 | | T | 1.15 | 2.89E-08 | 1.13 | 2.04E-06 |

**Table S6. Summary of independent SNPs defined by stepwise conditioning of the HLA locus**. RAF: Risk Allele Frequency, OR: Odds Ratio.

| | | | | | European RAF | | East Asian RAF | | Independent SNPs | |
|---|---|---|---|---|---|---|---|---|---|---|
| **Cohorts** | **SNP** | **BP** | **Closest Gene(s)** | **Risk Allele** | **Cases** | **Controls** | **Cases** | **Controls** | **OR** | **P-value** |
| **Combined** | rs9268557 | 32389305 | *HLA-DRA/BTNL2* | C | 0.614 | 0.509 | 0.6257 | 0.5656 | 1.24 | 4.52E-47 |
| | rs9275355 | 32667829 | *HLA-DQB1/HLA-DQA2* | C | 0.267 | 0.226 | 0.3925 | 0.3249 | 1.26 | 1.71E-34 |
| | rs9272105 | 32599999 | *HLA-DQA1* | A | 0.661 | 0.598 | 0.4778 | 0.4483 | 1.25 | 1.18E-28 |
| | rs9275596 | 32681631 | *HLA-DQB1/HLA-DQA2* | T | 0.748 | 0.663 | 0.8726 | 0.8127 | 1.33 | 3.16E-36 |
| | rs3128927 | 33074288 | *HLA-DPA2/HLA-DPB2* | C | 0.776 | 0.729 | 0.8606 | 0.8271 | 1.22 | 1.50E-25 |
| **European** | rs9268557 | 32389305 | *HLA-DRA/BTNL2* | C | 0.614 | 0.509 | 0.6257 | 0.5656 | 1.27 | 4.25E-38 |
| | rs3128927 | 33074288 | *HLA-DPA2/HLA-DPB2* | C | 0.776 | 0.729 | 0.8606 | 0.8271 | 1.22 | 1.73E-18 |
| | rs9272105 | 32599999 | *HLA-DQA1* | A | 0.661 | 0.598 | 0.4778 | 0.4483 | 1.31 | 2.66E-22 |
| | rs542418 | 31836442 | *SLC44A4* | A | 0.777 | 0.692 | 0.6701 | 0.6716 | 1.30 | 1.27E-12 |
| | rs4496841 | 32389997 | *HLA-DRA/BTNL2* | T | 0.804 | 0.742 | 0.7608 | 0.7324 | 1.29 | 1.76E-11 |
| **East Asian** | rs2760994 | 32574308 | *HLA-DRB1/HLA-DQA1* | T | 0.659 | 0.622 | 0.7943 | 0.7195 | 1.50 | 5.13E-34 |
| | rs1612904 | 32669018 | *HLA-DQB1/HLA-DQA2* | A | 0.730 | 0.681 | 0.8725 | 0.8121 | 1.48 | 1.88E-22 |
| | rs117552038 | 32583937 | *HLA-DRB1/HLA-DQA1* | C | 0.995 | 0.981 | 0.8723 | 0.8508 | 1.20 | 4.65E-05 |
| | rs11459466 | 32853221 | *TAP1* | G | 0.799 | 0.785 | 0.7745 | 0.7476 | 1.20 | 6.99E-07 |

**Table S7. Association analysis for *HLA-DRB1* amino-acid substitutions at positions 11 & 71.** Odds Ratio (OR) and 95% confidence intervals (95%CI) are depicted for each amino-acid substitution in reference to all other substitutions; P-value corresponds to a biallelic test of association; only amino-acid substitutions with frequency in controls greater than 1% are included in the association testing; NA = Not Available (control allele frequency < 1%).

| | | East Asian | | | | European | | | |
|---|---|---|---|---|---|---|---|---|---|
| Position | Substitution | OR | L95 | U95 | P | OR | L95 | U95 | P |
| DRB1_11 | **P** | **0.618** | **0.554** | **0.689** | **3.24E-18** | **0.706** | **0.656** | **0.760** | **9.83E-21** |
| | **V** | **1.498** | **1.362** | **1.647** | **6.71E-17** | **1.146** | **1.075** | **1.221** | **2.93E-05** |
| | G | NA | NA | NA | NA | **0.731** | **0.678** | **0.789** | **6.11E-16** |
| | L | 0.907 | 0.737 | 1.117 | 3.58E-01 | **1.270** | **1.181** | **1.365** | **9.34E-11** |
| | S | 1.120 | 1.040 | 1.207 | 2.81E-03 | 1.126 | 1.074 | 1.180 | 7.41E-07 |
| | D | 0.876 | 0.786 | 0.976 | 1.64E-02 | NA | NA | NA | NA |
| DRB1_71 | **A** | **0.617** | **0.550** | **0.692** | **1.54E-16** | **0.738** | **0.680** | **0.801** | **2.79E-13** |
| | **R** | **1.953** | **1.782** | **2.140** | **1.04E-23** | **1.194** | **1.138** | **1.253** | **5.18E-13** |
| | K | 0.669 | 0.554 | 0.809 | 3.15E-05 | 0.871 | 0.819 | 0.927 | 1.43E-05 |
| | E | 0.806 | 0.688 | 0.945 | 7.98E-03 | 1.041 | 0.973 | 1.114 | 2.45E-01 |

**Table S8. Association testing of the classical HLA alleles in the East Asian case-control cohorts**. OR: Odds Ratio; L95 and U95: lower and upper bounds of the 95% confidence interval.

| HLA Allele | Unconditional | | | | Conditioned on DRB1*1501 and DRB1*0405 | | | |
|---|---|---|---|---|---|---|---|---|
| | OR | L95 | U95 | P | OR | L95 | U95 | P |
| HLA_DRB1_0101 | 0.91 | 0.74 | 1.12 | 3.85E-01 | 0.91 | 0.74 | 1.13 | 3.91E-01 |
| **HLA_DRB1_0405** | **1.67** | **1.46** | **1.90** | **1.73E-14** | | | | |
| HLA_DRB1_0406 | 1.26 | 1.05 | 1.52 | 1.31E-02 | 1.28 | 1.06 | 1.55 | 9.12E-03 |
| HLA_DRB1_0803 | 1.06 | 0.91 | 1.24 | 4.44E-01 | 1.07 | 0.92 | 1.25 | 3.76E-01 |
| HLA_DRB1_0901 | 0.88 | 0.79 | 0.98 | 1.76E-02 | 0.88 | 0.79 | 0.99 | 2.90E-02 |
| HLA_DRB1_1202 | 1.23 | 1.06 | 1.43 | 6.55E-03 | 1.21 | 1.04 | 1.40 | 1.42E-02 |
| HLA_DRB1_1302 | 0.83 | 0.70 | 0.99 | 4.17E-02 | 0.82 | 0.69 | 0.98 | 3.09E-02 |
| **HLA_DRB1_1501** | **0.58** | **0.50** | **0.66** | **2.17E-15** | | | | |
| HLA_DRB1_1502 | 0.76 | 0.62 | 0.92 | 5.27E-03 | 0.77 | 0.63 | 0.94 | 9.57E-03 |
| HLA_DQB1_0301 | 1.20 | 1.09 | 1.31 | 1.36E-04 | 1.21 | 1.10 | 1.33 | 5.50E-05 |
| HLA_DQB1_0302 | 1.20 | 1.05 | 1.37 | 6.57E-03 | 1.22 | 1.07 | 1.40 | 2.94E-03 |
| HLA_DQB1_0303 | 0.89 | 0.80 | 0.99 | 2.51E-02 | 0.89 | 0.80 | 0.99 | 4.00E-02 |
| HLA_DQB1_0401 | 1.68 | 1.46 | 1.94 | 3.05E-13 | 1.19 | 0.91 | 1.57 | 2.08E-01 |
| HLA_DQB1_0501 | 1.01 | 0.86 | 1.18 | 9.21E-01 | 1.01 | 0.86 | 1.18 | 9.36E-01 |
| HLA_DQB1_0502 | 1.01 | 0.83 | 1.22 | 9.09E-01 | 1.04 | 0.86 | 1.26 | 6.82E-01 |
| HLA_DQB1_0503 | 1.36 | 1.13 | 1.63 | 1.10E-03 | 1.39 | 1.16 | 1.67 | 4.39E-04 |
| HLA_DQB1_0601 | 0.95 | 0.84 | 1.08 | 4.48E-01 | 0.99 | 0.88 | 1.13 | 9.35E-01 |
| HLA_DQB1_0602 | 0.55 | 0.48 | 0.64 | 1.20E-14 | 0.79 | 0.57 | 1.08 | 1.41E-01 |
| HLA_DQA1_0101 | 1.21 | 1.08 | 1.36 | 8.18E-04 | 1.22 | 1.09 | 1.37 | 8.21E-04 |
| HLA_DQA1_0102 | 0.63 | 0.57 | 0.71 | 7.80E-17 | 0.78 | 0.67 | 0.90 | 8.15E-04 |
| HLA_DQA1_0103 | 0.90 | 0.80 | 1.02 | 8.99E-02 | 0.92 | 0.81 | 1.04 | 1.74E-01 |
| HLA_DQA1_0301 | 1.23 | 1.14 | 1.33 | 4.15E-07 | 1.02 | 0.94 | 1.12 | 6.05E-01 |
| HLA_DQA1_0501 | 1.08 | 0.97 | 1.19 | 1.64E-01 | 1.09 | 0.98 | 1.21 | 1.16E-01 |
| HLA_DQA1_0601 | 1.20 | 1.03 | 1.38 | 1.57E-02 | 1.18 | 1.02 | 1.36 | 3.03E-02 |
| HLA_DPA1_0103 | 1.10 | 1.02 | 1.18 | 1.88E-02 | 1.10 | 1.02 | 1.19 | 1.29E-02 |
| HLA_DPA1_0201 | 0.73 | 0.65 | 0.82 | 1.07E-07 | 0.73 | 0.65 | 0.82 | 2.87E-07 |
| HLA_DPA1_0202 | 1.05 | 0.97 | 1.13 | 2.50E-01 | 1.04 | 0.96 | 1.12 | 3.56E-01 |
| HLA_DPB1_0201 | 1.10 | 1.01 | 1.20 | 2.31E-02 | 1.17 | 1.08 | 1.28 | 2.92E-04 |
| HLA_DPB1_0202 | 1.16 | 0.96 | 1.39 | 1.22E-01 | 1.16 | 0.96 | 1.39 | 1.26E-01 |
| HLA_DPB1_0402 | 1.06 | 0.91 | 1.24 | 4.25E-01 | 1.01 | 0.87 | 1.18 | 8.76E-01 |
| HLA_DPB1_0501 | 1.08 | 1.00 | 1.17 | 5.06E-02 | 1.05 | 0.97 | 1.14 | 2.07E-01 |
| HLA_DPB1_1301 | 0.77 | 0.64 | 0.93 | 6.99E-03 | 0.77 | 0.63 | 0.93 | 6.07E-03 |
| HLA_A_0206 | 0.97 | 0.85 | 1.10 | 6.13E-01 | 0.97 | 0.85 | 1.11 | 6.42E-01 |
| HLA_A_1101 | 1.16 | 1.04 | 1.28 | 5.47E-03 | 1.16 | 1.04 | 1.28 | 6.52E-03 |
| HLA_A_2402 | 1.06 | 0.96 | 1.16 | 2.33E-01 | 1.05 | 0.96 | 1.15 | 3.09E-01 |
| HLA_A_2601 | 0.99 | 0.84 | 1.17 | 9.16E-01 | 1.02 | 0.86 | 1.20 | 8.31E-01 |
| HLA_A_3101 | 1.05 | 0.88 | 1.25 | 5.82E-01 | 1.08 | 0.91 | 1.29 | 3.83E-01 |
| HLA_A_3303 | 0.83 | 0.73 | 0.95 | 5.29E-03 | 0.82 | 0.72 | 0.94 | 3.59E-03 |
| HLA_B_0702 | 0.75 | 0.59 | 0.95 | 1.85E-02 | 0.82 | 0.64 | 1.04 | 1.06E-01 |
| HLA_B_1301 | 1.04 | 0.84 | 1.27 | 7.32E-01 | 1.07 | 0.87 | 1.31 | 5.43E-01 |
| HLA_B_3501 | 1.14 | 0.97 | 1.35 | 1.20E-01 | 1.16 | 0.98 | 1.37 | 8.69E-02 |
| HLA_B_4001 | 1.13 | 0.97 | 1.31 | 1.12E-01 | 1.14 | 0.98 | 1.33 | 8.33E-02 |
| HLA_B_4403 | 0.86 | 0.72 | 1.02 | 7.99E-02 | 0.85 | 0.71 | 1.01 | 5.93E-02 |
| HLA_B_5101 | 1.02 | 0.89 | 1.16 | 8.04E-01 | 1.03 | 0.90 | 1.17 | 6.77E-01 |
| HLA_B_5201 | 0.67 | 0.55 | 0.82 | 8.80E-05 | 0.68 | 0.56 | 0.84 | 2.11E-04 |
| HLA_B_5401 | 1.24 | 1.06 | 1.45 | 6.03E-03 | 1.04 | 0.89 | 1.23 | 6.08E-01 |
| HLA_C_0102 | 1.10 | 0.99 | 1.21 | 7.38E-02 | 1.01 | 0.91 | 1.12 | 8.76E-01 |
| HLA_C_0303 | 1.01 | 0.90 | 1.15 | 8.17E-01 | 1.06 | 0.93 | 1.21 | 3.60E-01 |
| HLA_C_0304 | 1.06 | 0.94 | 1.19 | 3.43E-01 | 1.08 | 0.96 | 1.22 | 2.11E-01 |
| HLA_C_0401 | 0.96 | 0.83 | 1.11 | 5.55E-01 | 0.96 | 0.83 | 1.11 | 5.61E-01 |
| HLA_C_0602 | 0.89 | 0.77 | 1.02 | 9.44E-02 | 0.88 | 0.76 | 1.01 | 7.09E-02 |
| HLA_C_0702 | 0.99 | 0.88 | 1.11 | 8.07E-01 | 1.01 | 0.90 | 1.14 | 8.70E-01 |
| HLA_C_0801 | 1.14 | 1.02 | 1.28 | 2.66E-02 | 1.16 | 1.03 | 1.31 | 1.19E-02 |
| HLA_C_1202 | 0.73 | 0.61 | 0.88 | 7.77E-04 | 0.75 | 0.62 | 0.90 | 2.37E-03 |
| HLA_C_1402 | 1.08 | 0.92 | 1.27 | 3.60E-01 | 1.09 | 0.93 | 1.29 | 2.84E-01 |
| HLA_C_1502 | 0.99 | 0.82 | 1.20 | 9.06E-01 | 1.03 | 0.85 | 1.25 | 7.58E-01 |

**Table S9. Association testing of the classical HLA alleles in the European case-control cohorts**. OR: Odds Ratio; L95 and U95: lower and upper bounds of the 95% confidence interval.

| HLA Allele | Unconditional | | | | Control for 4 significant DRB1 alleles | | | | Control for DRB1 alleles & DQA1*0102 | | | | Control for DRB1 alleles, DQA1*0102 & DPA1*0103 | | | |
|---|---|---|---|---|---|---|---|---|---|---|---|---|---|---|---|---|
| | OR | L95 | U95 | P | OR | L95 | U95 | P | OR | L95 | U95 | P | OR | L95 | U95 | P |
| **HLA_DRB1_0101** | **1.31** | **1.20** | **1.41** | **6.48E-11** | | | | | | | | | | | | |
| **HLA_DRB1_0301** | **0.77** | **0.71** | **0.83** | **2.84E-10** | | | | | | | | | | | | |
| **HLA_DRB1_0701** | **0.73** | **0.68** | **0.79** | **2.96E-16** | | | | | | | | | | | | |
| HLA_DRB1_1201 | 1.14 | 0.96 | 1.35 | 1.39E-01 | 1.03 | 0.87 | 1.23 | 7.38E-01 | 0.99 | 0.83 | 1.18 | 8.90E-01 | 0.99 | 0.83 | 1.18 | 8.87E-01 |
| HLA_DRB1_1301 | 0.98 | 0.88 | 1.08 | 6.66E-01 | 0.88 | 0.79 | 0.97 | 1.24E-02 | 0.84 | 0.76 | 0.93 | 1.12E-03 | 0.85 | 0.77 | 0.95 | 3.11E-03 |
| HLA_DRB1_1302 | 0.99 | 0.88 | 1.11 | 8.50E-01 | 0.89 | 0.79 | 1.00 | 4.25E-02 | 1.39 | 1.17 | 1.65 | 2.20E-04 | 1.38 | 1.16 | 1.64 | 3.19E-04 |
| **HLA_DRB1_1501** | **0.73** | **0.67** | **0.79** | **2.50E-13** | | | | | | | | | | | | |
| HLA_DQB1_0201 | 0.79 | 0.73 | 0.86 | 1.30E-08 | 1.45 | 1.01 | 2.06 | 4.31E-02 | 1.38 | 0.96 | 1.98 | 8.01E-02 | 1.42 | 0.99 | 2.03 | 5.95E-02 |
| HLA_DQB1_0202 | 0.75 | 0.69 | 0.82 | 1.18E-10 | 1.09 | 0.92 | 1.28 | 3.11E-01 | 1.09 | 0.92 | 1.28 | 3.15E-01 | 1.12 | 0.95 | 1.32 | 1.86E-01 |
| HLA_DQB1_0301 | 1.25 | 1.18 | 1.32 | 5.83E-16 | 1.12 | 1.06 | 1.20 | 1.56E-04 | 1.06 | 1.00 | 1.13 | 6.30E-02 | 1.05 | 0.98 | 1.12 | 1.42E-01 |
| HLA_DQB1_0302 | 1.10 | 1.01 | 1.19 | 2.61E-02 | 0.97 | 0.89 | 1.05 | 4.71E-01 | 0.93 | 0.86 | 1.01 | 9.47E-02 | 0.93 | 0.85 | 1.01 | 8.49E-02 |
| HLA_DQB1_0303 | 0.74 | 0.65 | 0.84 | 5.71E-06 | 0.88 | 0.76 | 1.01 | 7.38E-02 | 0.87 | 0.76 | 1.00 | 5.84E-02 | 0.86 | 0.74 | 0.99 | 3.34E-02 |
| HLA_DQB1_0402 | 1.05 | 0.90 | 1.22 | 5.22E-01 | 0.95 | 0.82 | 1.11 | 5.35E-01 | 0.92 | 0.79 | 1.07 | 2.56E-01 | 0.92 | 0.79 | 1.07 | 2.54E-01 |
| HLA_DQB1_0501 | 1.34 | 1.25 | 1.43 | 1.85E-16 | 1.23 | 1.09 | 1.39 | 8.83E-04 | 1.18 | 1.05 | 1.34 | 7.09E-03 | 1.19 | 1.05 | 1.35 | 5.34E-03 |
| HLA_DQB1_0503 | 1.39 | 1.24 | 1.56 | 1.69E-08 | 1.26 | 1.12 | 1.42 | 8.12E-05 | 1.22 | 1.08 | 1.37 | 1.07E-03 | 1.22 | 1.09 | 1.37 | 7.67E-04 |
| HLA_DQB1_0602 | 0.73 | 0.67 | 0.80 | 6.71E-12 | 1.02 | 0.78 | 1.33 | 8.92E-01 | 1.07 | 0.82 | 1.40 | 6.05E-01 | 1.05 | 0.80 | 1.37 | 7.47E-01 |
| HLA_DQB1_0603 | 0.96 | 0.87 | 1.06 | 3.88E-01 | 0.88 | 0.79 | 0.97 | 1.03E-02 | 0.84 | 0.76 | 0.93 | 1.01E-03 | 0.86 | 0.77 | 0.95 | 3.21E-03 |
| HLA_DQB1_0604 | 1.09 | 0.96 | 1.23 | 2.02E-01 | 0.98 | 0.86 | 1.11 | 7.07E-01 | 1.54 | 1.29 | 1.83 | 1.15E-06 | 1.51 | 1.27 | 1.80 | 3.23E-06 |
| HLA_DQA1_0101 | 1.38 | 1.30 | 1.47 | 1.42E-24 | 1.26 | 1.15 | 1.38 | 2.85E-07 | 1.21 | 1.11 | 1.32 | 3.19E-05 | 1.22 | 1.11 | 1.33 | 1.75E-05 |
| **HLA_DQA1_0102** | **0.76** | **0.71** | **0.81** | **7.04E-17** | **0.75** | **0.68** | **0.82** | **6.35E-10** | | | | | | | | |
| HLA_DQA1_0103 | 0.97 | 0.88 | 1.06 | 4.79E-01 | 0.86 | 0.78 | 0.95 | 3.15E-03 | 0.83 | 0.75 | 0.91 | 1.35E-04 | 0.84 | 0.76 | 0.93 | 6.22E-04 |
| HLA_DQA1_0201 | 0.74 | 0.68 | 0.80 | 2.96E-15 | | | | | | | | | | | | |
| HLA_DQA1_0301 | 1.08 | 1.01 | 1.16 | 1.63E-02 | 0.94 | 0.88 | 1.01 | 9.49E-02 | 0.89 | 0.83 | 0.96 | 2.18E-03 | 0.89 | 0.83 | 0.96 | 1.43E-03 |
| HLA_DQA1_0401 | 0.98 | 0.84 | 1.14 | 7.87E-01 | 0.89 | 0.76 | 1.04 | 1.39E-01 | 0.85 | 0.73 | 1.00 | 4.88E-02 | 0.85 | 0.73 | 1.00 | 4.73E-02 |
| HLA_DQA1_0501 | 1.08 | 1.03 | 1.14 | 2.82E-03 | 1.17 | 1.10 | 1.25 | 1.46E-06 | 1.11 | 1.04 | 1.19 | 1.43E-03 | 1.11 | 1.03 | 1.18 | 3.35E-03 |
| **HLA_DPA1_0103** | **0.77** | **0.72** | **0.82** | **4.96E-16** | **0.82** | **0.76** | **0.87** | **1.12E-09** | **0.82** | **0.77** | **0.88** | **1.06E-08** | | | | |
| HLA_DPA1_0201 | 0.75 | 0.70 | 0.80 | 1.88E-15 | 0.80 | 0.75 | 0.86 | 2.80E-09 | 0.81 | 0.75 | 0.87 | 1.86E-08 | 0.90 | 0.78 | 1.04 | 1.65E-01 |
| HLA_DPA1_0202 | 0.93 | 0.80 | 1.07 | 3.15E-01 | 0.96 | 0.83 | 1.12 | 6.26E-01 | 0.97 | 0.84 | 1.12 | 6.59E-01 | 1.17 | 1.00 | 1.37 | 5.43E-02 |
| HLA_DPB1_0101 | 0.71 | 0.62 | 0.81 | 1.65E-07 | 0.81 | 0.71 | 0.93 | 2.79E-03 | 0.81 | 0.71 | 0.93 | 3.26E-03 | 0.95 | 0.82 | 1.11 | 5.28E-01 |
| HLA_DPB1_0201 | 1.14 | 1.07 | 1.21 | 9.01E-05 | 1.11 | 1.04 | 1.18 | 1.60E-03 | 1.11 | 1.04 | 1.18 | 2.05E-03 | 1.07 | 1.00 | 1.14 | 4.49E-02 |
| HLA_DPB1_0301 | 0.84 | 0.77 | 0.91 | 1.38E-05 | 0.82 | 0.76 | 0.89 | 1.95E-06 | 0.83 | 0.77 | 0.90 | 6.59E-06 | 0.80 | 0.73 | 0.86 | 4.39E-08 |
| HLA_DPB1_0401 | 1.06 | 1.01 | 1.11 | 2.79E-02 | 1.07 | 1.02 | 1.12 | 1.03E-02 | 1.06 | 1.01 | 1.11 | 2.02E-02 | 1.01 | 0.96 | 1.06 | 7.07E-01 |
| HLA_DPB1_0402 | 1.20 | 1.12 | 1.29 | 2.03E-07 | 1.13 | 1.05 | 1.21 | 5.50E-04 | 1.12 | 1.05 | 1.21 | 1.13E-03 | 1.09 | 1.02 | 1.17 | 1.70E-02 |
| HLA_DPB1_0501 | 1.02 | 0.85 | 1.21 | 8.52E-01 | 1.02 | 0.85 | 1.22 | 8.32E-01 | 1.07 | 0.89 | 1.27 | 4.83E-01 | 1.28 | 1.06 | 1.55 | 9.39E-03 |
| HLA_DPB1_1301 | 0.91 | 0.77 | 1.09 | 3.16E-01 | 0.94 | 0.79 | 1.12 | 5.07E-01 | 0.95 | 0.79 | 1.13 | 5.53E-01 | 1.09 | 0.91 | 1.31 | 3.65E-01 |
| HLA_A_0101 | 0.82 | 0.77 | 0.88 | 1.56E-08 | 0.88 | 0.82 | 0.95 | 6.84E-04 | 0.88 | 0.82 | 0.95 | 9.44E-04 | 0.89 | 0.83 | 0.96 | 1.62E-03 |
| HLA_A_0201 | 1.10 | 1.04 | 1.16 | 6.42E-04 | 1.06 | 1.00 | 1.12 | 3.50E-02 | 1.06 | 1.01 | 1.12 | 2.87E-02 | 1.06 | 1.01 | 1.12 | 2.99E-02 |
| HLA_A_0301 | 1.01 | 0.94 | 1.08 | 8.68E-01 | 1.00 | 0.93 | 1.07 | 9.36E-01 | 0.99 | 0.93 | 1.07 | 8.70E-01 | 0.99 | 0.92 | 1.06 | 7.99E-01 |
| HLA_A_1101 | 1.12 | 1.02 | 1.23 | 2.28E-02 | 1.06 | 0.96 | 1.17 | 2.80E-01 | 1.05 | 0.95 | 1.15 | 3.76E-01 | 1.05 | 0.95 | 1.16 | 3.34E-01 |
| HLA_A_2301 | 0.90 | 0.76 | 1.06 | 1.92E-01 | 0.96 | 0.82 | 1.13 | 6.54E-01 | 0.96 | 0.81 | 1.13 | 6.27E-01 | 0.96 | 0.81 | 1.13 | 5.96E-01 |
| HLA_A_2402 | 1.00 | 0.93 | 1.08 | 9.89E-01 | 0.97 | 0.90 | 1.05 | 5.17E-01 | 0.97 | 0.90 | 1.05 | 4.47E-01 | 0.97 | 0.89 | 1.05 | 3.95E-01 |
| HLA_A_2601 | 1.04 | 0.92 | 1.18 | 5.04E-01 | 1.01 | 0.89 | 1.15 | 8.63E-01 | 1.01 | 0.89 | 1.15 | 8.79E-01 | 1.01 | 0.89 | 1.14 | 9.16E-01 |
| HLA_A_3001 | 0.98 | 0.82 | 1.16 | 7.80E-01 | 1.10 | 0.92 | 1.31 | 2.99E-01 | 1.11 | 0.93 | 1.33 | 2.54E-01 | 1.12 | 0.94 | 1.34 | 2.02E-01 |
| HLA_A_3101 | 1.04 | 0.90 | 1.21 | 6.03E-01 | 0.99 | 0.85 | 1.15 | 9.09E-01 | 0.99 | 0.85 | 1.15 | 8.98E-01 | 0.99 | 0.85 | 1.15 | 8.61E-01 |
| HLA_A_3201 | 0.89 | 0.79 | 1.00 | 5.19E-02 | 0.87 | 0.77 | 0.98 | 2.76E-02 | 0.87 | 0.77 | 0.99 | 2.78E-02 | 0.87 | 0.77 | 0.99 | 2.81E-02 |
| HLA_A_6801 | 1.25 | 1.10 | 1.43 | 6.44E-04 | 1.20 | 1.05 | 1.37 | 6.47E-03 | 1.21 | 1.06 | 1.38 | 4.11E-03 | 1.21 | 1.06 | 1.38 | 3.97E-03 |
| HLA_B_0702 | 0.84 | 0.77 | 0.91 | 2.12E-05 | 0.93 | 0.84 | 1.01 | 9.94E-02 | 0.92 | 0.84 | 1.01 | 9.32E-02 | 0.92 | 0.84 | 1.01 | 9.20E-02 |
| HLA_B_0801 | 0.74 | 0.68 | 0.81 | 5.67E-11 | 0.82 | 0.73 | 0.92 | 6.47E-04 | 0.82 | 0.73 | 0.92 | 5.43E-04 | 0.83 | 0.74 | 0.93 | 1.40E-03 |
| HLA_B_1302 | 1.00 | 0.87 | 1.15 | 9.89E-01 | 1.25 | 1.07 | 1.45 | 3.78E-03 | 1.25 | 1.08 | 1.46 | 2.94E-03 | 1.26 | 1.09 | 1.47 | 2.15E-03 |
| HLA_B_1402 | 0.86 | 0.74 | 1.01 | 5.87E-02 | 0.81 | 0.69 | 0.95 | 8.77E-03 | 0.82 | 0.70 | 0.95 | 1.13E-02 | 0.83 | 0.70 | 0.97 | 1.75E-02 |
| HLA_B_1801 | 1.00 | 0.91 | 1.10 | 9.45E-01 | 1.02 | 0.92 | 1.12 | 7.56E-01 | 1.00 | 0.91 | 1.10 | 9.66E-01 | 0.99 | 0.90 | 1.09 | 8.56E-01 |
| HLA_B_2705 | 1.21 | 1.07 | 1.36 | 3.06E-03 | 1.09 | 0.96 | 1.24 | 1.66E-01 | 1.08 | 0.95 | 1.22 | 2.31E-01 | 1.08 | 0.95 | 1.23 | 2.22E-01 |
| HLA_B_4001 | 1.17 | 1.04 | 1.31 | 1.11E-02 | 1.06 | 0.94 | 1.20 | 3.06E-01 | 1.11 | 0.99 | 1.25 | 8.38E-02 | 1.11 | 0.98 | 1.25 | 1.03E-01 |
| HLA_B_4402 | 1.15 | 1.06 | 1.26 | 1.24E-03 | 1.06 | 0.97 | 1.16 | 1.90E-01 | 1.05 | 0.96 | 1.14 | 3.15E-01 | 1.04 | 0.95 | 1.14 | 3.77E-01 |
| HLA_B_4403 | 0.79 | 0.70 | 0.89 | 8.42E-05 | 0.93 | 0.81 | 1.05 | 2.50E-01 | 0.93 | 0.82 | 1.06 | 2.61E-01 | 0.94 | 0.83 | 1.07 | 3.83E-01 |
| HLA_B_4901 | 1.26 | 1.08 | 1.47 | 2.81E-03 | 1.21 | 1.04 | 1.41 | 1.65E-02 | 1.24 | 1.06 | 1.45 | 6.60E-03 | 1.24 | 1.06 | 1.45 | 6.21E-03 |
| HLA_B_5101 | 1.21 | 1.10 | 1.32 | 3.87E-05 | 1.14 | 1.04 | 1.25 | 4.45E-03 | 1.13 | 1.03 | 1.24 | 7.27E-03 | 1.14 | 1.04 | 1.24 | 6.31E-03 |
| HLA_B_5701 | 0.73 | 0.63 | 0.85 | 3.40E-05 | 0.83 | 0.71 | 0.97 | 2.08E-02 | 0.83 | 0.71 | 0.98 | 2.28E-02 | 0.82 | 0.70 | 0.96 | 1.41E-02 |
| HLA_C_0102 | 1.20 | 1.07 | 1.35 | 2.68E-03 | 1.08 | 0.96 | 1.22 | 1.86E-01 | 1.07 | 0.95 | 1.21 | 2.54E-01 | 1.07 | 0.95 | 1.21 | 2.64E-01 |
| HLA_C_0202 | 1.21 | 1.09 | 1.34 | 3.29E-04 | 1.13 | 1.02 | 1.25 | 2.27E-02 | 1.14 | 1.03 | 1.27 | 1.25E-02 | 1.15 | 1.03 | 1.27 | 1.10E-02 |
| HLA_C_0303 | 0.93 | 0.83 | 1.04 | 1.81E-01 | 0.87 | 0.77 | 0.97 | 1.26E-02 | 0.85 | 0.76 | 0.95 | 5.47E-03 | 0.86 | 0.77 | 0.96 | 7.26E-03 |
| HLA_C_0304 | 1.11 | 1.01 | 1.23 | 3.86E-02 | 1.02 | 0.92 | 1.13 | 7.15E-01 | 1.04 | 0.94 | 1.16 | 4.02E-01 | 1.04 | 0.94 | 1.15 | 4.57E-01 |
| HLA_C_0401 | 1.08 | 1.01 | 1.16 | 2.44E-02 | 1.00 | 0.93 | 1.07 | 9.95E-01 | 1.00 | 0.93 | 1.07 | 9.34E-01 | 0.99 | 0.93 | 1.07 | 8.46E-01 |
| HLA_C_0501 | 1.20 | 1.10 | 1.31 | 3.22E-05 | 1.15 | 1.06 | 1.26 | 1.20E-03 | 1.13 | 1.04 | 1.24 | 4.45E-03 | 1.12 | 1.03 | 1.22 | 8.98E-03 |
| HLA_C_0602 | 0.95 | 0.88 | 1.03 | 2.30E-01 | 1.09 | 1.00 | 1.19 | 4.45E-02 | 1.09 | 1.00 | 1.19 | 4.96E-02 | 1.08 | 0.99 | 1.18 | 7.25E-02 |
| HLA_C_0701 | 0.85 | 0.80 | 0.91 | 1.83E-06 | 0.90 | 0.84 | 0.97 | 6.19E-03 | 0.91 | 0.85 | 0.98 | 1.50E-02 | 0.92 | 0.85 | 0.99 | 2.16E-02 |
| HLA_C_0702 | 0.85 | 0.78 | 0.92 | 2.90E-05 | 0.92 | 0.85 | 1.00 | 5.97E-02 | 0.92 | 0.84 | 1.00 | 4.43E-02 | 0.92 | 0.84 | 1.00 | 4.20E-02 |
| HLA_C_0704 | 1.08 | 0.91 | 1.29 | 3.83E-01 | 1.01 | 0.84 | 1.20 | 9.40E-01 | 1.03 | 0.86 | 1.23 | 7.44E-01 | 1.02 | 0.86 | 1.22 | 8.07E-01 |
| HLA_C_0802 | 0.87 | 0.76 | 1.00 | 4.69E-02 | 0.86 | 0.75 | 0.98 | 2.97E-02 | 0.86 | 0.75 | 0.99 | 3.50E-02 | 0.87 | 0.75 | 1.00 | 4.29E-02 |
| HLA_C_1203 | 1.08 | 0.98 | 1.18 | 1.04E-01 | 1.04 | 0.94 | 1.14 | 4.56E-01 | 1.04 | 0.94 | 1.14 | 4.48E-01 | 1.04 | 0.95 | 1.14 | 4.22E-01 |
| HLA_C_1502 | 1.22 | 1.06 | 1.41 | 4.80E-03 | 1.17 | 1.01 | 1.34 | 3.15E-02 | 1.16 | 1.01 | 1.34 | 3.91E-02 | 1.17 | 1.01 | 1.34 | 3.33E-02 |

**Table S10. Pleiotropy analysis of significant non-HLA GWAS loci:** lead IgAN SNPs and their LD proxies (r2>0.5) were annotated against the NHGRI GWAS Catalogue to retrieve previously reported GWAS associations with other traits with $P<5x10^{-8}$.

| Previously published GWAS associations | | | | | | | | | Associations with IgAN | | | | Top SNP in the Locus | LD with the top SNP |
|---|---|---|---|---|---|---|---|---|---|---|---|---|---|---|
| **Locus** | **DISEASE/TRAIT** | **PMID** | **Journal** | **Year** | **Index SNP** | **Risk Allele** | **OR or BETA** | **P value** | **Risk allele** | **OR** | **P value** | **Direction** | **SNP** | **R2** |
| TNFSF4 | Allergy | 29083406 | *Nat Genet* | 2017 | rs4090390 | A | 1.05 | 1.00E-15 | A | 1.08 | 1.57E-02 | Concordant | rs4916312 | 0.750 |
| CFH | Neovascular age-related macular degeneration | 28703135 | *J Hum Genet* | 2017 | rs800292 | G | 1.67 | 3.00E-08 | G | 1.12 | 4.68E-08 | Concordant | rs6677604 | 0.910 |
| CFH | Circulating myeloperoxidase levels (serum) | 23620142 | *Hum Mol Genet* | 2013 | rs800292 | A | 0.15 | 5.00E-41 | G | 1.12 | 4.68E-08 | Opposed | | 0.910 |
| CFH | Sub-foveal choroidal thickness | 29844195 | *Proc Natl Acad Sci USA* | 2018 | rs800292 | A | 10.27 | 2.00E-10 | G | 1.12 | 4.68E-08 | Opposed | | 0.910 |
| CFH | Matrix metalloproteinase-8 levels | 29212897 | *Circ Cardiovasc Genet* | 2017 | rs800292 | A | 0.24 | 2.00E-35 | G | 1.12 | 4.68E-08 | Opposed | | 0.910 |
| REL | Multiple sclerosis | 24076602 | *Nat Genet* | 2013 | rs842639 | A | 1.10 | 2.00E-14 | G | 1.11 | 3.95E-08 | Opposed | rs842638 | 0.672 |
| REL | Rheumatoid arthritis | 23143596 | *Nat Genet* | 2012 | rs13031237 | A | 1.12 | 5.00E-14 | G | 1.08 | 6.73E-04 | Opposed | | 0.661 |
| REL | Psoriasis | 25574825 | *Am J Hum Genet* | 2015 | rs35741374 | T | 1.20 | 4.00E-12 | T | 1.13 | 5.31E-09 | Concordant | | 0.988 |
| CD28 | Primary biliary cirrhosis | 28425483 | *Nat Commun* | 2017 | rs4675369 | G | 1.30 | 1.00E-13 | A | 1.09 | 2.01E-04 | Opposed | rs3769684 | 0.596 |
| IRF4 | Takayasu artheritis | 30498034 | *Proc Natl Acad Sci USA* | 2018 | rs17133698 | C | 1.50 | 3.00E-09 | C | 1.20 | 2.11E-10 | Concordant | rs12201499 | 0.792 |
| LY86 | Neutrophil count | 27863252 | *Cell* | 2016 | rs1334577 | A | 0.02 | 3.00E-09 | G | 1.13 | 2.19E-09 | Opposed | rs12530084 | 0.967 |
| LY86 | Basophil count + eosinophil count | 27863252 | *Cell* | 2016 | rs1334577 | A | 0.02 | 4.00E-09 | G | 1.13 | 2.19E-09 | Opposed | | 0.967 |
| LY86 | Pulse pressure | 30224653 | *Nat Genet* | 2018 | rs1334576 | A | 0.11 | 4.00E-11 | G | 1.09 | 2.07E-05 | Opposed | | 0.778 |
| LY86 | Leukocyte count | 27863252 | *Cell* | 2016 | rs11303054 | C | 0.03 | 2.00E-12 | C | 1.13 | 7.18E-06 | Concordant | | 0.571 |
| LY86 | Urate levels | 23263486 | *Nat Genet* | 2013 | rs675209 | T | 4.39 | 1.00E-23 | C | 1.10 | 1.20E-05 | Opposed | | 0.541 |
| PF4V1 | Sum neutrophil eosinophil counts | 27863252 | *Cell* | 2017 | rs16850073 | T | 0.05 | 2.00E-45 | T | 1.10 | 4.23E-04 | Concordant | rs6828610 | 0.513 |
| PF4V1 | Granulocyte percentage of myeloid white cells | 27863252 | *Cell* | 2017 | rs16850073 | T | 0.04 | 5.00E-23 | T | 1.10 | 4.23E-04 | Concordant | | 0.513 |
| PF4V1 | Leukocyte count | 27863252 | *Cell* | 2017 | rs16850073 | T | 0.05 | 3.00E-34 | T | 1.10 | 4.23E-04 | Concordant | | 0.513 |
| PF4V1 | Monocyte percentage of white cells | 27863252 | *Cell* | 2017 | rs16850073 | T | 0.03 | 5.00E-13 | T | 1.10 | 4.23E-04 | Concordant | | 0.513 |
| TNFSF8/15 | Biliary liver cirrhosis | 23000144 | *Am J Hum Genet* | 2012 | rs4979462 | T | 1.56 | 3.00E-14 | T | 1.11 | 7.53E-04 | Concordant | rs13300483 | 0.563 |
| TNFSF8/15 | Primary biliary cirrhosis | 28062665 | *Hum Mol Genet* | 2017 | rs4979462 | C | 1.60 | 8.00E-19 | T | 1.11 | 7.53E-04 | Opposed | | 0.563 |
| TNFSF8/15 | Serum IgA | 24676358 | *Hum Mol Genet* | 2014 | rs7853287 | A | 0.12 | 3.00E-10 | A | 1.13 | 1.89E-05 | Concordant | | 0.682 |
| CARD9 | Inflammatory bowel diseases | 23128233 | *Nature* | 2012 | rs10781499 | A | 1.19 | 4.00E-56 | A | 1.14 | 7.21E-11 | Concordant | rs4077515 | 0.987 |
| CARD9 | Crohn's disease | 26192919 | *Nat Genet* | 2015 | rs10781499 | A | 1.19 | 8.00E-43 | A | 1.14 | 7.21E-11 | Concordant | | 0.987 |
| CARD9 | Ulcerative colitis | 26192919 | *Nat Genet* | 2015 | rs10781499 | A | 1.14 | 4.00E-26 | A | 1.14 | 7.21E-11 | Concordant | | 0.987 |
| CARD9 | Ankylosing spondylitis | 23749187 | *Nat Genet* | 2013 | rs1128905 | C | 1.12 | 2.00E-09 | C | 1.09 | 1.34E-06 | Concordant | | 0.754 |
| CARD9 | Pediatric chronic inflammatory diseases | 26301688 | *Nat Med* | 2015 | rs11145763 | C | NA | 3.00E-08 | C | 1.12 | 4.00E-08 | Concordant | | 0.983 |
| CARD9 | Granulocyte percentage of myeloid white cells | 27863252 | *Cell* | 2016 | rs3812565 | C | 0.03 | 4.00E-13 | C | 1.12 | 3.66E-08 | Concordant | | 0.815 |
| CARD9 | Vital capacity | 28166213 | *Nat Genet* | 2017 | rs10870202 | C | 0.02 | 9.00E-10 | C | 1.09 | 7.89E-07 | Concordant | | 0.539 |
| REEP3 | Schizophrenia | 28991256 | *Nat Genet* | 2017 | rs111364339 | C | 1.10 | 5.00E-09 | T | 1.10 | 6.91E-03 | Opposed | rs57917667 | 0.641 |
| ZMIZ1/PPIF | Crohn's disease | 26192919 | *Nat Genet* | 2015 | rs1250546 | G | 1.12 | 3.00E-19 | A | 1.12 | 1.44E-08 | Opposed | rs1108618 | 0.877 |
| ZMIZ1/PPIF | Granulocyte count | 27863252 | *Cell* | 2016 | rs2802372 | C | 1.16 | 7.00E-09 | A | 1.13 | 4.90E-10 | Opposed | | 0.832 |
| ZMIZ1/PPIF | Inflammatory bowel diseases | 23128233 | *Nature* | 2012 | rs1250546 | A | 1.10 | 3.00E-18 | A | 1.12 | 1.44E-08 | Concordant | | 0.877 |
| ZMIZ1/PPIF | Lymphocyte percentage of leukocytes | 27863252 | *Cell* | 2016 | rs1250569 | C | 0.02 | 1.00E-11 | T | 1.13 | 2.24E-09 | Opposed | | 0.799 |
| ZMIZ1/PPIF | Multiple sclerosis | 24076602 | *Nat Genet* | 2013 | ZMIZ1/PPIF | A | 1.10 | 3.00E-15 | C | 1.10 | 2.82E-07 | Opposed | | 0.748 |
| ZMIZ1/PPIF | Neutrophil percentage of leukocytes | 27863252 | *Cell* | 2016 | rs1250569 | C | 0.02 | 2.00E-09 | T | 1.13 | 2.24E-09 | Opposed | | 0.799 |
| ZMIZ1/PPIF | Celiac disease | 22057235 | *Nat Genet* | 2011 | rs1250552 | A | 1.12 | 8.00E-17 | A | 1.09 | 1.39E-04 | Concordant | | 0.557 |
| ZMIZ1/PPIF | Pediatric chronic inflammatory diseases | 26301688 | *Nat Med* | 2015 | rs1250563 | G | NA | 1.00E-08 | G | 1.10 | 1.57E-05 | Concordant | | 0.584 |
| OVOL1/RELA | Atopic eczema | 26482879 | *Nat Genet* | 2015 | rs10791824 | G | 1.12 | 2.00E-19 | A | 1.17 | 1.70E-12 | Opposed | rs10896045 | 0.593 |
| OVOL1/RELA | Allergy | 29083406 | *Nat Genet* | 2017 | rs479844 | G | 1.04 | 2.00E-13 | A | 1.09 | 3.85E-04 | Opposed | | 0.552 |
| OVOL1/RELA | Systemic lupus erythematosus | 27399966 | *Nat Genet* | 2016 | rs494003 | A | 1.14 | 6.00E-09 | A | 1.12 | 3.75E-04 | Concordant | | 0.530 |
| ETS1 | Albumin-globulin ratio | 29403010 | *Nat Genet* | 2018 | rs7127911 | T | 0.03 | 2.00E-11 | T | 1.12 | 2.31E-06 | Concordant | rs7121743 | 0.744 |
| ETS1 | Non-albumin protein levels | 29403010 | *Nat Genet* | 2018 | rs7127911 | T | 0.03 | 3.00E-11 | T | 1.12 | 2.31E-06 | Concordant | | 0.744 |
| ITGAM/ITGAX | Autoimmune thyroid disease | 22922229 | *Hum Mol Genet* | 2012 | rs57348955 | G | 1.19 | 5.00E-08 | A | 1.09 | 6.78E-03 | Opposed | rs11150612 | 0.725 |
| IRF8 | Biliary liver cirrhosis | 21399635 | *Nat Genet* | 2011 | rs11117432 | G | 1.31 | 5.00E-11 | G | 1.12 | 8.99E-04 | Concordant | rs1879210 | 0.508 |
| IRF8 | Primary biliary cirrhosis | 22961000 | *Nat Genet* | 2012 | rs11117433 | G | 1.26 | 1.00E-09 | G | 1.09 | 1.43E-03 | Concordant | | 0.510 |
| IRF8 | Multiple sclerosis | 19525953 | *Nat Genet* | 2009 | rs17445836 | G | 1.25 | 4.00E-09 | G | 1.12 | 1.43E-03 | Concordant | | 0.505 |
| IRF8 | Eosinophil count | 27863252 | *Cell* | 2016 | rs17445836 | A | 0.03 | 3.00E-10 | G | 1.12 | 1.43E-03 | Opposed | | 0.505 |
| IRF8 | Systemic lupus erythematosus | 28714469 | *Nat Commun* | 2017 | rs11117433 | G | 1.18 | 2.00E-11 | G | 1.09 | 1.43E-03 | Concordant | | 0.510 |
| TNFRSF13B | Non-albumin protein levels | 29403010 | *Nat Genet* | 2018 | rs4985726 | G | 0.15 | 1.00E-193 | G | 1.14 | 8.58E-09 | Concordant | rs57382045 | 0.979 |
| TNFRSF13B | Albumin-globulin ratio | 29403010 | *Nat Genet* | 2018 | rs57166795 | G | 0.13 | 5.00E-166 | A | 1.16 | 5.98E-09 | Opposed | | 0.989 |
| TNFRSF13B | Total blood protein levels | 29403010 | *Nat Genet* | 2018 | rs4985726 | G | 0.10 | 3.00E-107 | G | 1.14 | 8.58E-09 | Concordant | | 0.979 |
| TNFRSF13B | Albumin level | 29403010 | *Nat Genet* | 2018 | rs34562254 | G | 0.03 | 2.00E-11 | A | 1.16 | 8.00E-09 | Opposed | | 0.884 |
| TNFRSF13B | IgG levels | 29403010 | *Nat Genet* | 2018 | rs74998556 | T | 0.15 | 1.00E-26 | T | 1.11 | 1.70E-03 | Concordant | | 0.628 |
| TNFRSF13B | Na level | 29403010 | *Nat Genet* | 2018 | rs34562254 | G | 0.03 | 8.00E-10 | A | 1.16 | 8.00E-09 | Opposed | | 0.884 |
| TNFRSF13B | Multiple myeloma | 27363682 | *Nat Commun* | 2016 | rs34562254 | A | 1.30 | 4.00E-17 | A | 1.16 | 8.00E-09 | Concordant | | 0.884 |
| LIF | Lymphocyte count | 27863252 | *Cell* | 2016 | rs714027 | G | 0.03 | 3.00E-21 | G | 1.14 | 3.99E-14 | Concordant | rs4823074 | 0.996 |
| LIF | Type 1 diabetes | 25751624 | *Nat Genet* | 2015 | rs4820830 | C | 1.12 | 1.00E-12 | C | 1.07 | 1.21E-03 | Concordant | | 0.585 |
| LIF | Inflammatory bowel diseases | 23128233 | *Nature* | 2012 | rs2412970 | A | 1.08 | 3.00E-14 | G | 1.19 | 1.56E-14 | Opposed | | 1.000 |
| LIF | Crohn's disease | 26192919 | *Nat Genet* | 2015 | rs5763767 | A | 1.09 | 6.00E-13 | G | 1.14 | 3.00E-14 | Opposed | | 1.000 |
| LIF | Lymphocyte percentage of white cells | 27863252 | *Cell* | 2016 | rs5763821 | C | 0.03 | 1.00E-13 | A | 1.14 | 7.61E-07 | Opposed | | 0.966 |
| LIF | Albumin-globulin ratio | 29403010 | *Nat Genet* | 2018 | rs2412974 | T | 0.04 | 2.00E-10 | C | 1.15 | 1.30E-11 | Opposed | | 0.702 |
| LIF | Non-albumin protein levels | 29403010 | *Nat Genet* | 2018 | rs2412975 | T | 0.03 | 3.00E-10 | T | 1.16 | 1.46E-13 | Concordant | | 1.000 |
| LIF | Neutrophil percentage of white cells | 27863252 | *Cell* | 2016 | rs5763821 | C | 0.02 | 7.00E-10 | A | 1.14 | 7.61E-07 | Opposed | | 0.966 |
| LIF | Tonsillectomy | 27941131 | *J Med Genet* | 2016 | rs2412971 | G | 1.22 | 1.00E-09 | G | 1.14 | 3.11E-14 | Concordant | | 1.000 |
| LIF | Mathematical ability | 30038396 | *Nat Genet* | 2018 | rs9614099 | A | 0.01 | 1.00E-09 | T | 1.09 | 8.18E-05 | Opposed | | 0.561 |
| LIF | Creatine kinase levels | 29403010 | *Nat Genet* | 2018 | rs5763790 | G | 0.03 | 1.00E-08 | C | 1.07 | 7.71E-03 | Opposed | | 0.562 |
| LIF | Optic disc area | 25631615 | *Genet Epidemiol* | 2015 | rs2412970 | G | 0.02 | 3.00E-08 | A | 1.19 | 1.56E-14 | Opposed | | 1.000 |

**Table S11. Pleiotropy analysis of suggestive non-HLA GWAS loci ($P<1x10^{-5}$):** lead IgAN SNPs and their LD proxies (r2>0.5) were annotated against the NHGRI GWAS Catalogue to retrieve previously reported GWAS associations with other traits with $P<5x10^{-8}$.

| Previously published GWAS associations | | | | | | | | | Associations with IgAN | | | | Top SNP in the Locus | LD with the top SNP |
|---|---|---|---|---|---|---|---|---|---|---|---|---|---|---|
| **Locus** | **DISEASE/TRAIT** | **PMID** | **Journal** | **Year** | **Index SNP** | **Risk Allele** | **OR or BETA** | **P value** | **Risk allele** | **OR** | **P value** | **Direction** | **SNP** | **R2** |
| FCER1G | Allergy | 27182965 | *Nat Genet* | 2016 | rs2070902 | T | 1.06 | 1.00E-09 | T | 1.09 | 0.001 | Concordant | rs4233368 | 0.507 |
| FCER1G | Crohn's disease | 26192919 | *Nat Genet* | 2015 | rs1801274 | G | 1.08 | 9.00E-11 | A | 1.11 | 4.61E-06 | Opposed | | 0.880 |
| FCER1G | Ulcerative cholitis | 21297633 | *Nat Genet* | 2011 | rs1801274 | A | 1.21 | 2.00E-20 | A | 1.11 | 4.61E-06 | Concordant | | 0.880 |
| FCER1G | Inflammatory bowel diseases | 23128233 | *Nature* | 2012 | rs1801274 | A | 1.12 | 2.00E-38 | A | 1.11 | 4.61E-06 | Concordant | | 0.880 |
| FCER1G | Ankylosing spondylitis | 23749187 | *Nat Genet* | 2013 | rs1801274 | A | 1.12 | 1.00E-09 | A | 1.11 | 4.61E-06 | Concordant | | 0.880 |
| FCER1G | Systemic lupus erithematosus | 27399966 | *Nat Genet* | 2017 | rs1801274 | G | 1.21 | 6.00E-11 | A | 1.11 | 4.61E-06 | Opposed | | 0.880 |
| CXCR2 | Ankylosing spondylitis | 26974007 | *Nat Genet* | 2016 | rs11676348 | T | NA | 2.00E-11 | C | 1.07 | 6.12E-05 | Opposed | rs4674259 | 0.817 |
| CXCR2 | Ulcerative cholitis | 21297633 | *Nat Genet* | 2011 | rs11676348 | T | 1.07 | 1.00E-10 | C | 1.07 | 6.12E-05 | Opposed | | 0.817 |
| INPP5D | Allergy | 29083406 | *Nat Genet* | 2017 | rs1057258 | C | 1.05 | 1.00E-10 | T | 1.14 | 2.01E-06 | Opposed | rs14243 | 0.718 |
| INPP5D | Eosinophil percentage of leukocytes | 27863252 | *Cell* | 2016 | rs56235204 | C | 0.19 | 3.00E-20 | C | 1.12 | 6.84E-07 | Concordant | | 0.785 |
| INPP5D | Eosinophil count | 27863252 | *Cell* | 2016 | rs56235204 | C | 0.03 | 2.00E-14 | C | 1.12 | 6.84E-07 | Concordant | | 0.785 |
| INPP5D | Neutrophil percentage of granulocytes | 27863252 | *Cell* | 2016 | rs56235204 | C | 0.04 | 2.00E-19 | C | 1.12 | 6.84E-07 | Concordant | | 0.785 |
| ZFP36L2 | Multiple sclerosis | 21833088 | *Nature* | 2011 | rs12466022 | C | 1.11 | 6.00E-10 | A | 1.07 | 0.0008 | Opposed | rs737013 | 0.547 |
| ZFP36L2 | granulocyte percentage of myeloid white cells | 27863252 | *Cell* | 2016 | rs28498283 | T | 0.04 | 2.00E-18 | T | 1.07 | 0.0009 | Concordant | | 0.581 |
| ZFP36L2 | monocyte percentage of leukocytes | 27863252 | *Cell* | 2016 | rs28498283 | T | 0.03 | 6.00E-16 | T | 1.07 | 0.0009 | Concordant | | 0.581 |
| RN7SL51P | Crohn's disease | 26192919 | *Nat Genet* | 2015 | rs11679753 | G | 1.13 | 5.00E-11 | A | 1.09 | 2.17E-06 | Opposed | rs10865331 | 0.983 |
| RN7SL51P | Ankylosing spondylitis | 23749187 | *Nat Genet* | 2013 | rs6759298 | C | 1.31 | 4.00E-41 | G | 1.09 | 2.34E-06 | Opposed | | 0.849 |
| TCF7 | Multiple sclerosis | 24076602 | *Nat Genet* | 2013 | rs756699 | A | 1.12 | 9.00E-11 | C | 1.07 | 0.0036 | Opposed | rs151822 | 0.539 |
| IL12B | Psoriasis | 19169254 | *Nat Genet* | 2009 | rs2082412 | G | 1.44 | 2.00E-28 | A | 1.09 | 7.39E-05 | Opposed | rs3213097 | 0.982 |
| IL7R | Atopic eczema | 26482879 | *Nat Genet* | 2015 | rs10214237 | T | 1.06 | 3.00E-14 | T | 1.09 | 0.0002 | Concordant | rs10213865 | 0.934 |
| IL7R | Neutrophil percentage of granulocytes | 27863252 | *Cell* | 2016 | rs4594881 | T | 0.03 | 1.00E-17 | G | 1.09 | 0.0004 | Opposed | | 0.810 |
| IL7R | Eosinophil percentage of leukocytes | 27863252 | *Cell* | 2016 | rs4594881 | T | 0.03 | 3.00E-17 | G | 1.09 | 0.0004 | Opposed | | 0.810 |
| IL7R | Primary biliary cirrhosis | 22961000 | *Nat Genet* | 2012 | rs6871748 | A | 1.30 | 2.00E-13 | T | 1.09 | 0.0003 | Opposed | | 0.934 |
| IL7R | Biliary liver cirrhosis | 23000144 | *Am J Hum Genet* | 2012 | rs6890853 | G | 1.47 | 4.00E-08 | G | 1.09 | 4.04E-05 | Opposed | | 0.962 |
| IL7R | Multiple sclerosis | 24076602 | *Nat Genet* | 2013 | rs6881706 | C | 1.12 | 4.00E-17 | G | 1.09 | 5.86E-05 | Opposed | | 1.000 |
| IL7R | Allergy | 29083406 | *Nat Genet* | 2017 | rs7717955 | C | 1.07 | 9.00E-36 | C | 1.10 | 1.47E-05 | Concordant | | 0.995 |
| PTGER4 | Crohn's disease | 26192919 | *Nat Genet* | 2015 | rs11742570 | A | 1.28 | 4.00E-87 | T | 1.01 | 0.5421 | Opposed | rs4957300 | 0.537 |
| PTGER4 | Inflammatory bowel diseases | 23128233 | *Nature* | 2012 | rs11742570 | C | 1.20 | 2.00E-82 | T | 1.01 | 0.5421 | Opposed | | 0.537 |
| PTGER4 | Ulcerative cholitis | 21297633 | *Nat Genet* | 2011 | rs6451493 | T | 1.08 | 3.00E-09 | G | 1.01 | 0.5605 | Opposed | | 0.537 |
| PTGER4 | Multiple sclerosis | 24076602 | *Nat Genet* | 2013 | rs6880778 | G | 1.12 | 8.00E-20 | A | 1.01 | 0.512 | Opposed | | 0.537 |
| PTGER4 | Allergy | 29083406 | *Nat Genet* | 2017 | rs7714574 | T | 1.03 | 6.00E-10 | T | 1.08 | 0.0008 | Concordant | | 0.522 |
| ANKRD55 | Rheumathoid arthritis | 20453842 | *Nat Genet* | 2010 | rs6859219 | C | 1.28 | 1.00E-11 | C | 1.15 | 1.96E-07 | Concordant | rs10065637 | 0.994 |
| ANKRD55 | Multiple sclerosis | 24076602 | *Nat Genet* | 2013 | rs71624119 | G | 1.13 | 3.00E-13 | G | 1.15 | 4.25E-07 | Concordant | | 0.842 |
| ANKRD55 | Crohn's disease | 26192919 | *Nat Genet* | 2015 | rs71624119 | G | NA | 7.00E-10 | G | 1.15 | 4.25E-07 | Concordant | | 0.842 |
| TNFAIP3 | Systemic lupus erithematosus | 28714469 | *Nat Commun* | 2017 | rs5029939 | C | 1.48 | 5.00E-29 | G | 1.15 | 0.0004 | Opposed | rs58905141 | 0.670 |
| TNFAIP3 | Allergy | 29083406 | *Nat Genet* | 2017 | rs5029937 | G | 1.08 | 2.00E-08 | T | 1.17 | 0.0005 | Opposed | | 0.670 |
| TNFAIP3 | Rheumathoid arthritis | 24390342 | *Nature* | 2013 | rs7752903 | G | 1.38 | 2.00E-29 | G | 1.14 | 0.0036 | Concordant | | 0.813 |
| TAGAP | Crohn's disease | 26192919 | *Nat Genet* | 2015 | rs212388 | G | 1.11 | 2.00E-16 | C | 1.04 | 0.0253 | Opposed | rs2485360 | 0.559 |
| TAGAP | Rheumathoid arthritis | 24390342 | *Nature* | 2013 | rs2451258 | T | 1.10 | 3.00E-11 | C | 1.08 | 0.0011 | Opposed | | 0.735 |
| TAGAP | Psoriasis | 23143594 | *Nat Genet* | 2012 | rs2451258 | C | 1.12 | 3.00E-08 | C | 1.08 | 0.0011 | Concordant | | 0.735 |
| TAGAP | Basophil count + neutrophil count | 27863252 | *Cell* | 2016 | rs2451279 | G | 0.03 | 4.00E-12 | A | 1.07 | 0.0049 | Opposed | | 0.671 |
| TAGAP | Leukocyte count | 27863252 | *Cell* | 2016 | rs2451279 | G | 0.03 | 8.00E-12 | A | 1.07 | 0.0049 | Opposed | | 0.671 |
| IKZF1 | Granulocyte percentage of myeloid white cells | 27863252 | *Cell* | 2016 | rs78697948 | G | 0.08 | 1.00E-30 | G | 1.16 | 8.95E-05 | Concordant | rs79441475 | 0.827 |
| IKZF1 | Monocyte percentage of white cells | 27863252 | *Cell* | 2016 | rs78697948 | G | 0.08 | 3.00E-30 | G | 1.16 | 8.95E-05 | Concordant | | 0.827 |
| IKZF1 | Monocyte count | 27863252 | *Cell* | 2016 | rs78697948 | G | 0.06 | 4.00E-19 | G | 1.16 | 8.95E-05 | Concordant | | 0.827 |
| CCAT1 | Multiple mieloma | 27363682 | *Nat Commun* | 2016 | rs1948915 | C | 1.13 | 4.00E-11 | C | 1.09 | 1.02E-05 | Concordant | rs6989575 | 0.900 |

**Table S12. Genome-wide genetic correlations between IgAN with and without the HLA region and relevant diseases and traits.** All analyses are performed with LDSC software, h2: SNP-based heritability, rg: genetic correlation coefficient, se: standard error, z: z-score, p: p-value.

| Disease/Trait | Reference | h2 | Correlations with IgAN | | | | Correlations with IgAN without MHC region | | | |
|---|---|---|---|---|---|---|---|---|---|---|
| | | | rg | se | z | p | rg | se | z | p |
| Allergy | Ferreira et al. Nat Genet 2017 | 0.03 | 0.0977 | 0.0672 | 1.4545 | 0.1458 | 0.1823 | 0.0652 | 2.7969 | 0.0052 |
| Ankylosing spondylitis | Zhou et al. Nat Genet 2018 | 0.02 | 0.1493 | 0.1559 | 0.9582 | 0.338 | -0.2357 | 0.1818 | -1.2964 | 0.1948 |
| Crohn disease | De Lange et al. Nat Genet 2017 | 0.27 | -0.171 | 0.0666 | -2.5657 | 0.0103 | -0.2163 | 0.0722 | -2.9974 | 0.0027 |
| Celiac disease | Zhou et al. Nat Genet 2018 | 0.05 | -0.2835 | 0.1611 | -1.7594 | 0.0785 | -0.0505 | 0.2704 | -0.1869 | 0.8517 |
| IgA deficiency | Bronson et al. Nat Genet 2016 | 1.00 | 0.2702 | 0.1053 | 2.5649 | 0.0103 | 0.1703 | 0.202 | 0.8432 | 0.3991 |
| Inflammatory bowel diseases | De Lange et al. Nat Genet 2017 | 0.19 | -0.1596 | 0.0618 | -2.5807 | 0.0099 | -0.1832 | 0.0732 | -2.501 | 0.0124 |
| Membranous nephropathy | Xie et al. Nat Comm 2020 | 0.53 | -0.2574 | 0.1757 | -1.4646 | 0.143 | -0.1075 | 0.1612 | -0.6671 | 0.5047 |
| Multiple sclerosis | Int. MS Genetics Consortium. Science 2019 | 0.38 | -0.153 | 0.1369 | -1.1171 | 0.2639 | 0.0635 | 0.0774 | 0.8203 | 0.412 |
| Primary sclerosing cholangitis | Ji et al. Nat Genet 2017 | 0.46 | -0.3696 | 0.1288 | -2.8704 | 0.0041 | -0.2974 | 0.1144 | -2.6004 | 0.0093 |
| Rheumatoid arthritis | Okada et al. Nature 2014 | 0.28 | 0.3417 | 0.1813 | 1.8853 | 0.0594 | 0.1596 | 0.0845 | 1.8889 | 0.0589 |
| Systemic Lupus Erythematosus | Bentham et al. Nat Genet 2015 | 0.39 | -0.2242 | 0.1703 | -1.3165 | 0.188 | -0.0109 | 0.1563 | -0.0696 | 0.9445 |
| Type 1 Diabetes | Zhou et al. Nat Genet 2018 | 0.014 | -0.0384 | 0.0861 | -0.4457 | 0.6558 | -0.0224 | 0.1303 | -0.1721 | 0.8634 |
| Total IgA levels | Liu et al. (accompanying paper) | 0.07 | 0.1862 | 0.1144 | 1.6271 | 0.1037 | 0.3138 | 0.102 | 3.075 | 0.0021 |
| Ulcerative Colitis | De Lange et al. Nat Genet 2017 | 0.16 | -0.0834 | 0.0542 | -1.5376 | 0.1242 | -0.0586 | 0.0787 | -0.7439 | 0.4569 |
| Childhood ear infections | Tian et al. Nat Comm 2017 | 0.04 | 0.0484 | 0.0665 | 0.7266 | 0.4675 | -0.0044 | 0.0768 | -0.0571 | 0.9545 |
| Colds last year | Tian et al. Nat Comm 2017 | 0.016 | 0.0577 | 0.1141 | 0.5055 | 0.6132 | -0.044 | 0.3077 | -0.143 | 0.8863 |
| Mumps | Tian et al. Nat Comm 2017 | 0.012 | -0.0129 | 0.1021 | -0.1262 | 0.8996 | -0.1247 | 0.1338 | -0.9319 | 0.3514 |
| Plantar warts | Tian et al. Nat Comm 2017 | 0.04 | 0.1229 | 0.0805 | 1.5268 | 0.1268 | 0.08 | 0.1612 | 0.4963 | 0.6197 |
| Pneumonia | Tian et al. Nat Comm 2017 | 0.02 | 0.26 | 0.0785 | 3.3133 | 0.0009 | 0.2034 | 0.1008 | 2.0183 | 0.0436 |
| Positive TB test | Tian et al. Nat Comm 2017 | 0.014 | 0.0859 | 0.1326 | 0.648 | 0.517 | 0.1716 | 0.3609 | 0.4754 | 0.6345 |
| Rubella | Tian et al. Nat Comm 2017 | 0.01 | 0.0146 | 0.1549 | 0.0944 | 0.9248 | -0.139 | 0.1517 | -0.9159 | 0.3597 |
| Scarlet fever | Tian et al. Nat Comm 2017 | 0.011 | -0.0893 | 0.2421 | -0.3688 | 0.7123 | 0.244 | 0.154 | 1.5837 | 0.1133 |
| Shingles | Tian et al. Nat Comm 2017 | 0.014 | -0.2477 | 0.1767 | -1.4015 | 0.1611 | -0.0492 | 0.1532 | -0.3213 | 0.748 |
| Strep. Throat | Tian et al. Nat Comm 2017 | 0.03 | 0.0502 | 0.1022 | 0.4906 | 0.6237 | 0.123 | 0.116 | 1.0601 | 0.2891 |
| Tonsillectomy | Tian et al. Nat Comm 2017 | 0.04 | 0.115 | 0.0895 | 1.2844 | 0.199 | 0.1717 | 0.0817 | 2.1016 | 0.0356 |
| UTI frequency | Tian et al. Nat Comm 2017 | 0.04 | 0.253 | 0.0821 | 3.082 | 0.0021 | 0.2007 | 0.0963 | 2.0849 | 0.0371 |
| Yeast infections | Tian et al. Nat Comm 2017 | 0.04 | 0.0713 | 0.0892 | 0.7993 | 0.4241 | 0.0086 | 0.0938 | 0.0921 | 0.9266 |
| Albuminuria | Haas et al. Am J Hum Genet 2018 | 0.03 | 0.0932 | 0.044 | 2.1207 | 0.0339 | 0.0797 | 0.0564 | 1.4145 | 0.1572 |
| Body Mass Index (BMI) | Locke et al. Nature 2015 | 0.14 | 0.0622 | 0.0477 | 1.3038 | 0.1923 | 0.067 | 0.0498 | 1.3444 | 0.1788 |
| Blood Urea Nitrogen (BUN) | Wuttke et al. Nat Genet 2019 | 0.02 | -0.0169 | 0.0377 | -0.4471 | 0.6548 | -0.0263 | 0.0519 | -0.5065 | 0.6125 |
| Coronary Artery Disease | Nikpay et al. Nat Genet 2015 | 0.04 | 0.0965 | 0.0331 | 2.9152 | 0.0036 | 0.0803 | 0.0463 | 1.7332 | 0.0831 |
| Creatinine-based eGFR | Pattaro et al. Nat Comm 2016 | 0.07 | 0.0585 | 0.0969 | 0.6044 | 0.5456 | -0.0073 | 0.0757 | -0.0958 | 0.9236 |
| Cystatin-C-based eGFR | Pattaro et al. Nat Comm 2016 | 0.09 | 0.0707 | 0.1247 | 0.5666 | 0.571 | -0.0047 | 0.1139 | -0.0413 | 0.9671 |
| FEV1/FVC | Shrine et al. Nat Genet 2019 | 0.11 | 0.0875 | 0.0484 | 1.8072 | 0.0707 | 0.0364 | 0.0383 | 0.95 | 0.3421 |
| Height | Wood et al. Nat Genet 2014 | 0.35 | 0.023 | 0.0439 | 0.5241 | 0.6002 | 0.0311 | 0.0464 | 0.67 | 0.5028 |
| Essential Hypertension | Zhou et al. Nat Genet 2018 | 0.04 | 0.0811 | 0.035 | 2.3144 | 0.0206 | 0.0718 | 0.0436 | 1.6472 | 0.0995 |
| LDL cholesterol | Willer et al. Nat Gen 2013 | 0.04 | -0.056 | 0.0919 | -0.6098 | 0.542 | -0.1321 | 0.0694 | -1.9043 | 0.0569 |
| eGFR creatinine | Wuttke et al. Nat Genet 2019 | 0.04 | 0.0157 | 0.0441 | 0.3552 | 0.7224 | -0.025 | 0.0411 | -0.6089 | 0.5426 |
| Serum Cholesterol | Willer et al. Nat Gen 2013 | 0.05 | -0.0196 | 0.124 | -0.1581 | 0.8744 | -0.1222 | 0.0695 | -1.7574 | 0.0788 |
| Serum Triglycerides | Willer et al. Nat Gen 2013 | 0.05 | 0.0631 | 0.0953 | 0.6622 | 0.5078 | -0.0023 | 0.0698 | -0.0333 | 0.9735 |
| Type 2 Diabetes | Xue et al. Nat Comm 1018 | 0.02 | 0.0512 | 0.0637 | 0.8036 | 0.4216 | 0.052 | 0.0657 | 0.7916 | 0.4286 |

**Table S13. Overlapping signals between GWAS for IgAN and GWAS for serum IgA levels.** The effects of top independently significant loci for IgAN in GWAS for IgA levels; four most significant loci with concordant effects overlapping between both GWAS studies are highlighted in yellow; five additional loci with concordant effects that are genome-wide significant in IgAN GWAS and nominally significant (P<0.05) in the GWAS for IgA levels are highlighted in blue. The HLA alleles with opposed effects are highlighted in red.

| | | | | IgAN GWAS | | | IgA level GWAS | | |
|---|---|---|---|---|---|---|---|---|---|
| Locus | CHR | BP | SNP | Risk Allele | OR | P-value | Beta | P-value | Direction |
| *CFH* | 1 | 196686918 | rs6677604 | G | 1.21 | 1.50E-17 | 0.013 | 3.02E-01 | - |
| *CFH* | 1 | 196603302 | rs12029571 | A | 1.12 | 2.50E-06 | 0.015 | 2.11E-01 | - |
| *FCRL3/4* | 1 | 157542162 | rs849815 | A | 1.14 | 3.90E-09 | 0.011 | 3.35E-01 | - |
| *TNFSF4* | 1 | 173146357 | rs4916312 | A | 1.14 | 5.00E-08 | 0.037 | 1.20E-03 | Concordant |
| *CD28* | 2 | 204584759 | rs3769684 | T | 1.19 | 5.10E-11 | 0.027 | 2.05E-01 | - |
| *REL* | 2 | 61092678 | rs842638 | T | 1.17 | 9.60E-10 | 0.039 | 2.19E-04 | Concordant |
| *PF4V1* | 4 | 74725320 | rs6828610 | G | 1.14 | 3.50E-08 | 0.011 | 4.22E-01 | - |
| *IRF4* | 6 | 249571 | rs12201499 | C | 1.18 | 3.10E-11 | 0.034 | 2.21E-02 | Concordant |
| *RREB1* | 6 | 7214676 | rs12530084 | C | 1.13 | 1.30E-09 | 0.013 | 2.81E-01 | - |
| *HLA* | 6 | 32389305 | rs9268557 | C | 1.24 | 4.50E-47 | -0.009 | 3.70E-01 | - |
| *HLA* | 6 | 32667829 | rs9275355 | C | 1.26 | 1.71E-34 | -0.047 | 1.73E-03 | Opposite |
| *HLA* | 6 | 32599999 | rs9272105 | A | 1.25 | 1.18E-28 | -0.049 | 5.15E-13 | Opposite |
| *HLA* | 6 | 32681631 | rs9275596 | T | 1.33 | 3.16E-36 | -0.043 | 1.33E-04 | Opposite |
| *HLA* | 6 | 33074288 | rs3128927 | C | 1.22 | 1.50E-25 | -0.012 | 3.31E-01 | - |
| *DEFA1* | 8 | 6808722 | rs2075836 | T | 1.21 | 5.80E-11 | 0.008 | 4.58E-01 | - |
| *LYN* | 8 | 56852496 | rs75413466 | A | 1.40 | 1.40E-10 | 0.027 | 6.34E-01 | - |
| *ANXA13* | 8 | 124765474 | rs34354351 | T | 1.15 | 3.50E-08 | -0.003 | 8.48E-01 | - |
| *CARD9* | 9 | 139266496 | rs4077515 | T | 1.14 | 2.60E-11 | -0.003 | 7.55E-01 | - |
| *TNFSF8/15* | 9 | 117643362 | rs13300483 | T | 1.13 | 1.30E-08 | 0.079 | 3.17E-10 | Concordant |
| *ZMIZ1/PPIF* | 10 | 81043743 | rs1108618 | A | 1.14 | 1.90E-10 | 0.002 | 8.35E-01 | - |
| *REEP3* | 10 | 65363048 | rs57917667 | G | 1.22 | 1.10E-08 | 0.070 | 2.17E-02 | Concordant |
| *OVOL1/RELA* | 11 | 65555524 | rs10896045 | A | 1.18 | 4.70E-13 | 0.066 | 2.57E-22 | Concordant |
| *ETS1* | 11 | 128487069 | rs7121743 | C | 1.13 | 3.40E-08 | 0.009 | 5.22E-01 | - |
| *IGH* | 14 | 107222014 | rs751081288 | A | 1.17 | 1.90E-08 | 0.030 | 1.67E-01 | - |
| *ITGAM/ITGAX* | 16 | 31357760 | rs11150612 | A | 1.16 | 8.40E-14 | -0.007 | 5.30E-01 | - |
| *IRF8* | 16 | 86017715 | rs1879210 | T | 1.14 | 9.90E-09 | 0.007 | 5.47E-01 | - |
| *TNFSF13* | 17 | 7462969 | rs3803800 | A | 1.15 | 1.20E-10 | 0.065 | 9.41E-08 | Concordant |
| *TNFRSF13B* | 17 | 16851450 | rs57382045 | A | 1.16 | 3.40E-09 | 0.046 | 4.74E-03 | Concordant |
| *FCAR* | 19 | 55397217 | rs1865097 | A | 1.12 | 7.70E-09 | -0.019 | 1.56E-01 | - |
| *LIF/HORMAD2* | 22 | 30512478 | rs4823074 | G | 1.16 | 7.80E-15 | 0.053 | 6.75E-17 | Concordant |

**Table S14. Enrichment tests for the GWAS candidate gene set against human ortholog gene sets producing 27 phenotype categories when knocked out in mice.**

| Knockout mouse phenotype category | No. knockout mouse genes with human ortholog | No. overlap with IgAN genes | P-value |
|---|---|---|---|
| **immune system** | **3701** | **37** | **1.35E-12** |
| **hematopoietic system** | **3667** | **33** | **3.22E-09** |
| cellular | 3782 | 31 | 3.07E-07 |
| integument | 2046 | 19 | 1.58E-05 |
| mortality/aging | 5534 | 35 | 8.99E-05 |
| neoplasm | 996 | 11 | 2.00E-04 |
| endocrine/exocrine glands | 2445 | 18 | 1.80E-03 |
| liver/biliary system | 1359 | 12 | 2.10E-03 |
| skeleton | 2454 | 17 | 6.10E-03 |
| embryo | 1954 | 14 | 1.07E-02 |
| homeostasis/metabolism | 5220 | 28 | 1.83E-02 |
| digestive/alimentary system | 1481 | 11 | 2.10E-02 |
| vision/eye | 1796 | 12 | 3.95E-02 |
| growth/size/body | 4508 | 24 | 3.96E-02 |
| nervous system | 3631 | 20 | 4.98E-02 |
| renal/urinary system | 1377 | 9 | 1.03E-01 |
| respiratory system | 1442 | 9 | 1.36E-01 |
| behavior/neurological | 3944 | 19 | 2.07E-01 |
| pigmentation | 516 | 4 | 2.29E-01 |
| limbs/digits/tail | 1158 | 7 | 2.46E-01 |
| cardiovascular system | 2889 | 14 | 3.07E-01 |
| craniofacial | 1243 | 7 | 3.31E-01 |
| muscle | 1537 | 7 | 6.82E-01 |
| hearing/vestibular/ear | 796 | 2 | 8.17E-01 |
| adipose tissue | 1156 | 4 | 9.23E-01 |
| reproductive system | 2263 | 8 | 9.39E-01 |
| taste/olfaction | 153 | 0 | 9.43E-01 |

**Table S15. Genome-wide cell type-specific heritability enrichment for functional annotations based on FUN-LDA scoring system for all ENCODE and Roadmap Epigenomics cell types and tissues**. Prop. SNPs: proportion of SNPs enriched. Prop. h2: proportion of heritability explained by the SNPs enriched. Prop. h2 SE: standard error of proportion of heritability. Enrichment: heritability enrichment, calculated as (Prop. SNPs)/(Prop. h2). Enrichment SE: standard error of the enrichment. Enrichment p: p-value of the enrichment. Coefficient: regression coefficient of each cell-type specific FUN-LDA score. Coefficient SE: standard error of the regression coefficient. Coefficient z-score: z score of the regression coefficient.

| Epigenome ID | Group | Epigenome Mnemonic | Standardized Epigenome Name | Prop. SNPs | Prop. h2 | Prop. h2 SE | Enrichment | Enrichment SE | Enrichment P-value | Coefficient | Coefficient SE | Coefficient Z-score |
|---|---|---|---|---|---|---|---|---|---|---|---|---|
| E030 | HSC & B-cell | BLD.CD15.PC | Primary neutrophils from peripheral blood | 0.03 | 0.69 | 0.12 | 26.65 | 4.76 | 5.96E-10 | 5.52E-07 | 2.36E-07 | 2.34 |
| E041 | Blood & T-cell | BLD.CD4.CD25M.IL17M.PL.TPC | Primary T helper cells PMA-I stimulated | 0.03 | 0.54 | 0.10 | 20.95 | 3.92 | 2.11E-09 | 5.26E-07 | 2.99E-07 | 1.76 |
| E032 | HSC & B-cell | BLD.CD19.PPC | Primary B cells from peripheral blood | 0.02 | 0.70 | 0.14 | 33.93 | 6.67 | 1.99E-08 | 7.39E-07 | 3.70E-07 | 2.00 |
| E031 | HSC & B-cell | BLD.CD19.CPC | Primary B cells from cord blood | 0.03 | 0.61 | 0.12 | 23.73 | 4.75 | 2.36E-08 | -1.37E-08 | 3.60E-07 | -0.04 |
| E051 | HSC & B-cell | BLD.MOB.CD34.PC.M | Primary hematopoietic stem cells G-CSF-mobilized Male | 0.03 | 0.57 | 0.11 | 21.45 | 4.33 | 1.14E-07 | 5.45E-07 | 3.81E-07 | 1.43 |
| E115 | ENCODE2012 | BLD.DND41.CNCR | Dnd41 TCell Leukemia Cell Line | 0.02 | 0.53 | 0.11 | 29.81 | 6.21 | 2.55E-07 | 6.87E-07 | 2.54E-07 | 2.70 |
| E124 | ENCODE2012 | BLD.CD14.MONO | Monocytes-CD14+ RO01746 Primary Cells | 0.03 | 0.53 | 0.11 | 20.66 | 4.47 | 3.10E-07 | -1.07E-07 | 2.63E-07 | -0.41 |
| E116 | ENCODE2012 | BLD.GM12878 | GM12878 Lymphoblastoid Cells | 0.02 | 0.56 | 0.14 | 29.43 | 7.23 | 3.51E-06 | 2.27E-07 | 3.34E-07 | 0.68 |
| E113 | Other | SPLN | Spleen | 0.02 | 0.41 | 0.10 | 21.84 | 5.35 | 4.50E-06 | 4.49E-07 | 3.16E-07 | 1.42 |
| E101 | Digestive | GI.RECT.MUC.29 | Rectal Mucosa Donor 29 | 0.02 | 0.50 | 0.11 | 25.04 | 5.45 | 6.42E-06 | 1.28E-06 | 4.90E-07 | 2.62 |
| E066 | Other | LIV.ADLT | Liver | 0.03 | 0.41 | 0.10 | 15.91 | 3.70 | 8.61E-06 | 1.97E-07 | 1.58E-07 | 1.25 |
| E042 | Blood & T-cell | BLD.CD4.CD25M.IL17P.PL.TPC | Primary T helper 17 cells PMA-I stimulated | 0.02 | 0.44 | 0.11 | 22.25 | 5.38 | 1.83E-05 | 3.49E-07 | 3.86E-07 | 0.91 |
| E047 | Blood & T-cell | BLD.CD8.NPC | Primary T CD8+ naive cells from peripheral blood | 0.02 | 0.42 | 0.11 | 22.74 | 5.89 | 2.32E-05 | 5.50E-07 | 4.82E-07 | 1.14 |
| E046 | HSC & B-cell | BLD.CD56.PC | Primary Natural Killer cells from peripheral blood | 0.02 | 0.49 | 0.12 | 27.38 | 6.61 | 2.63E-05 | 2.27E-07 | 3.90E-07 | 0.58 |
| E093 | Thymus | THYM.FET | Fetal Thymus | 0.02 | 0.44 | 0.12 | 22.04 | 5.80 | 6.03E-05 | -2.85E-08 | 3.28E-07 | -0.09 |
| E037 | Blood & T-cell | BLD.CD4.MPC | Primary T helper memory cells from peripheral blood 2 | 0.02 | 0.38 | 0.10 | 18.42 | 4.73 | 9.27E-05 | 1.55E-07 | 4.35E-07 | 0.35 |
| E029 | HSC & B-cell | BLD.CD14.PC | Primary monocytes from peripheral blood | 0.02 | 0.49 | 0.13 | 22.31 | 6.14 | 9.89E-05 | -1.53E-07 | 3.60E-07 | -0.43 |
| E033 | Blood & T-cell | BLD.CD3.CPC | Primary T cells from cord blood | 0.02 | 0.39 | 0.11 | 24.76 | 6.76 | 1.10E-04 | 2.41E-07 | 4.92E-07 | 0.49 |
| E045 | Blood & T-cell | BLD.CD4.CD25I.CD127.TMEMPC | Primary T cells effector/memory enriched from periph. blood | 0.02 | 0.36 | 0.11 | 19.45 | 5.73 | 1.60E-04 | 7.94E-08 | 4.76E-07 | 0.17 |
| E043 | Blood & T-cell | BLD.CD4.CD25M.TPC | Primary T helper cells from peripheral blood | 0.02 | 0.40 | 0.11 | 17.41 | 4.95 | 2.02E-04 | -4.61E-08 | 4.18E-07 | -0.11 |
| E063 | Adipose | FAT.ADIP.NUC | Adipose Nuclei | 0.03 | 0.45 | 0.12 | 16.80 | 4.42 | 2.11E-04 | 3.01E-07 | 2.67E-07 | 1.13 |
| E110 | Digestive | GI.STMC.MUC | Stomach Mucosa | 0.02 | 0.39 | 0.11 | 18.54 | 5.05 | 2.16E-04 | 4.31E-07 | 2.92E-07 | 1.48 |
| E129 | ENCODE2012 | BONE.OSTEO | Osteoblast Primary Cells | 0.03 | 0.40 | 0.10 | 15.19 | 3.95 | 2.21E-04 | 6.23E-07 | 2.87E-07 | 2.17 |
| E040 | Blood & T-cell | BLD.CD4.CD25M.CD45RO.MPC | Primary T helper memory cells from peripheral blood 1 | 0.02 | 0.36 | 0.10 | 16.85 | 4.69 | 2.40E-04 | -1.42E-07 | 5.27E-07 | -0.27 |
| E119 | ENCODE2012 | BRST.HMEC | HMEC Mammary Epithelial Primary Cells | 0.02 | 0.37 | 0.10 | 19.38 | 5.07 | 2.40E-04 | 4.33E-07 | 3.98E-07 | 1.09 |
| E109 | Digestive | GI.S.INT | Small Intestine | 0.02 | 0.31 | 0.09 | 17.02 | 4.96 | 3.20E-04 | 6.62E-08 | 3.46E-07 | 0.19 |
| E034 | Blood & T-cell | BLD.CD3.PPC | Primary T cells from peripheral blood | 0.02 | 0.38 | 0.10 | 19.75 | 5.38 | 3.46E-04 | -3.19E-07 | 4.19E-07 | -0.76 |
| E044 | Blood & T-cell | BLD.CD4.CD25.CD127M.TREGPC | Primary T regulatory cells from peripheral blood | 0.02 | 0.33 | 0.10 | 17.13 | 4.94 | 4.35E-04 | -3.28E-07 | 3.66E-07 | -0.90 |
| E028 | Epithelial | BRST.HMEC.35 | Breast variant Human Mammary Epithelial Cells (vHMEC) | 0.02 | 0.35 | 0.10 | 18.54 | 5.09 | 4.47E-04 | 2.90E-07 | 4.05E-07 | 0.72 |
| E025 | Mesench | FAT.ADIP.DR.MSC | Adipose Derived Mesenchymal Stem Cell Cultured Cells | 0.03 | 0.36 | 0.10 | 11.14 | 3.04 | 6.46E-04 | 8.05E-08 | 1.97E-07 | 0.41 |
| E056 | Epithelial | SKIN.PEN.FRSK.FIB.02 | Foreskin Fibroblast Primary Cells skin02 | 0.02 | 0.38 | 0.11 | 15.18 | 4.27 | 7.13E-04 | 3.21E-07 | 4.84E-07 | 0.66 |
| E091 | Other | PLCNT.FET | Placenta | 0.02 | 0.31 | 0.09 | 15.82 | 4.74 | 7.44E-04 | 1.37E-07 | 1.83E-07 | 0.75 |
| E059 | Epithelial | SKIN.PEN.FRSK.MEL.01 | Foreskin Melanocyte Primary Cells skin01 | 0.01 | 0.28 | 0.08 | 19.06 | 5.82 | 8.17E-04 | 3.11E-07 | 3.59E-07 | 0.87 |
| E035 | HSC & B-cell | BLD.CD34.PC | Primary hematopoietic stem cells | 0.02 | 0.43 | 0.13 | 22.27 | 6.85 | 8.59E-04 | 1.48E-07 | 4.91E-07 | 0.30 |
| E106 | Digestive | GI.CLN.SIG | Sigmoid Colon | 0.02 | 0.30 | 0.10 | 16.40 | 5.32 | 8.73E-04 | -1.48E-07 | 4.13E-07 | -0.36 |
| E036 | HSC & B-cell | BLD.CD34.CC | Primary hematopoietic stem cells short term culture | 0.02 | 0.41 | 0.13 | 17.67 | 5.80 | 1.03E-03 | -3.62E-07 | 3.61E-07 | -1.00 |
| E084 | Digestive | GI.L.INT.FET | Fetal Intestine Large | 0.02 | 0.29 | 0.09 | 13.13 | 4.01 | 1.14E-03 | 2.10E-07 | 3.26E-07 | 0.65 |
| E112 | Thymus | THYM | Thymus | 0.02 | 0.32 | 0.11 | 21.22 | 7.23 | 1.30E-03 | -3.14E-07 | 4.83E-07 | -0.65 |
| E061 | Epithelial | SKIN.PEN.FRSK.MEL.03 | Foreskin Melanocyte Primary Cells skin03 | 0.02 | 0.28 | 0.09 | 14.32 | 4.44 | 1.48E-03 | -3.95E-08 | 2.41E-07 | -0.16 |
| E127 | ENCODE2012 | SKIN.NHEK | NHEK-Epidermal Keratinocyte Primary Cells | 0.02 | 0.31 | 0.09 | 14.10 | 4.20 | 1.56E-03 | -1.26E-07 | 3.15E-07 | -0.40 |
| E057 | Epithelial | SKIN.PEN.FRSK.KER.02 | Foreskin Keratinocyte Primary Cells skin02 | 0.02 | 0.33 | 0.10 | 16.92 | 5.02 | 1.56E-03 | 1.70E-07 | 3.82E-07 | 0.44 |
| E075 | Digestive | GI.CLN.MUC | Colonic Mucosa | 0.02 | 0.31 | 0.10 | 16.15 | 5.17 | 1.74E-03 | -5.20E-08 | 3.84E-07 | -0.14 |
| E107 | Muscle | MUS.SKLT.M | Skeletal Muscle Male | 0.02 | 0.31 | 0.09 | 12.70 | 3.86 | 1.76E-03 | 1.00E-07 | 3.20E-07 | 0.31 |
| E123 | ENCODE2012 | BLD.K562.CNCR | K562 Leukemia Cells | 0.02 | 0.28 | 0.09 | 16.87 | 5.35 | 1.85E-03 | 2.34E-09 | 2.13E-07 | 0.01 |
| E090 | Muscle | MUS.LEG.FET | Fetal Muscle Leg | 0.02 | 0.33 | 0.10 | 14.24 | 4.50 | 1.86E-03 | 5.08E-07 | 3.56E-07 | 1.43 |
| E050 | HSC & B-cell | BLD.MOB.CD34.PC.F | Primary hematopoietic stem cells G-CSF-mobilized Female | 0.02 | 0.36 | 0.11 | 14.74 | 4.69 | 1.97E-03 | -5.89E-07 | 3.39E-07 | -1.74 |
| E120 | ENCODE2012 | MUS.HSMM | HSMM Skeletal Muscle Myoblasts Cells | 0.02 | 0.31 | 0.09 | 16.45 | 5.01 | 1.99E-03 | 5.71E-07 | 3.36E-07 | 1.70 |
| E126 | ENCODE2012 | SKIN.NHDFAD | NHDF-Ad Adult Dermal Fibroblast Primary Cells | 0.02 | 0.32 | 0.10 | 17.81 | 5.69 | 2.02E-03 | 4.58E-07 | 3.02E-07 | 1.51 |
| E108 | Muscle | MUS.SKLT.F | Skeletal Muscle Female | 0.02 | 0.31 | 0.10 | 12.69 | 3.95 | 2.03E-03 | -1.18E-09 | 2.76E-07 | 0.00 |
| E079 | Digestive | GI.ESO | Esophagus | 0.01 | 0.27 | 0.08 | 19.41 | 6.01 | 2.12E-03 | 3.99E-07 | 3.91E-07 | 1.02 |
| E074 | Brain | BRN.SUB.NIG | Brain Substantia Nigra | 0.03 | 0.29 | 0.09 | 10.79 | 3.38 | 2.22E-03 | 3.41E-07 | 3.03E-07 | 1.13 |
| E122 | ENCODE2012 | VAS.HUVEC | HUVEC Umbilical Vein Endothelial Primary Cells | 0.02 | 0.27 | 0.09 | 14.57 | 4.58 | 2.32E-03 | 5.25E-08 | 1.74E-07 | 0.30 |
| E062 | Blood & T-cell | BLD.PER.MONUC.PC | Primary mononuclear cells from peripheral blood | 0.01 | 0.33 | 0.12 | 23.97 | 8.50 | 2.51E-03 | -1.15E-07 | 5.19E-07 | -0.22 |
| E017 | IMR90 | LNG.IMR90 | IMR90 fetal lung fibroblasts Cell Line | 0.02 | 0.32 | 0.10 | 16.62 | 5.07 | 2.77E-03 | 2.59E-07 | 3.13E-07 | 0.83 |
| E049 | Mesench | STRM.CHON.MRW.DR.MSC | Mesenchymal Stem Cell Derived Chondrocyte Cultured Cells | 0.03 | 0.31 | 0.10 | 11.20 | 3.49 | 2.94E-03 | 3.23E-08 | 2.49E-07 | 0.13 |
| E098 | Other | PANC | Pancreas | 0.01 | 0.23 | 0.08 | 18.67 | 6.56 | 3.08E-03 | 5.13E-07 | 3.61E-07 | 1.42 |
| E068 | Brain | BRN.ANT.CAUD | Brain Anterior Caudate | 0.02 | 0.29 | 0.09 | 11.81 | 3.78 | 3.10E-03 | 4.30E-07 | 2.98E-07 | 1.44 |
| E038 | Blood & T-cell | BLD.CD4.NPC | Primary T helper naive cells from peripheral blood | 0.02 | 0.30 | 0.10 | 17.35 | 5.80 | 3.17E-03 | -2.29E-07 | 5.52E-07 | -0.41 |
| E039 | Blood & T-cell | BLD.CD4.CD25M.CD45RA.NPC | Primary T helper naive cells from peripheral blood | 0.02 | 0.28 | 0.09 | 13.96 | 4.59 | 3.28E-03 | -5.34E-07 | 4.10E-07 | -1.30 |
| E078 | Sm. Muscle | GI.DUO.SM.MUS | Duodenum Smooth Muscle | 0.02 | 0.32 | 0.11 | 14.61 | 5.08 | 3.68E-03 | -1.88E-08 | 3.15E-07 | -0.06 |
| E054 | Neurosph | BRN.GANGEM.DR.NRSPHR | Ganglion Eminence derived primary cultured neurospheres | 0.01 | 0.27 | 0.10 | 19.57 | 6.87 | 4.06E-03 | 9.11E-07 | 4.73E-07 | 1.93 |
| E114 | ENCODE2012 | LNG.A549.ETOH002.CNCR | A549 EtOH 0.02pct Lung Carcinoma Cell Line | 0.02 | 0.25 | 0.08 | 11.42 | 3.64 | 4.11E-03 | -5.50E-08 | 1.56E-07 | -0.35 |
| E058 | Epithelial | SKIN.PEN.FRSK.KER.03 | Foreskin Keratinocyte Primary Cells skin03 | 0.02 | 0.26 | 0.09 | 12.48 | 4.08 | 4.84E-03 | -3.30E-07 | 3.57E-07 | -0.92 |
| E085 | Digestive | GI.S.INT.FET | Fetal Intestine Small | 0.02 | 0.27 | 0.10 | 12.27 | 4.36 | 5.08E-03 | -1.22E-08 | 3.70E-07 | -0.03 |
| E023 | Mesench | FAT.MSC.DR.ADIP | Mesenchymal Stem Cell Derived Adipocyte Cultured Cells | 0.03 | 0.30 | 0.10 | 9.90 | 3.22 | 5.22E-03 | -1.41E-07 | 2.27E-07 | -0.62 |
| E080 | Other | ADRL.GLND.FET | Fetal Adrenal Gland | 0.02 | 0.29 | 0.10 | 12.86 | 4.57 | 5.26E-03 | 4.44E-08 | 1.85E-07 | 0.24 |
| E076 | Sm. Muscle | GI.CLN.SM.MUS | Colon Smooth Muscle | 0.02 | 0.27 | 0.09 | 11.54 | 4.03 | 5.90E-03 | 2.76E-08 | 2.62E-07 | 0.11 |
| E055 | Epithelial | SKIN.PEN.FRSK.FIB.01 | Foreskin Fibroblast Primary Cells skin01 | 0.03 | 0.26 | 0.09 | 9.89 | 3.43 | 6.37E-03 | -4.50E-07 | 3.74E-07 | -1.21 |
| E027 | Epithelial | BRST.MYO | Breast Myoepithelial Primary Cells | 0.02 | 0.25 | 0.09 | 11.56 | 4.01 | 6.42E-03 | -2.40E-07 | 2.01E-07 | -1.19 |
| E006 | ES-deriv | ESDR.H1.MSC | H1 Derived Mesenchymal Stem Cells | 0.02 | 0.27 | 0.10 | 16.36 | 5.84 | 6.64E-03 | 1.30E-07 | 3.41E-07 | 0.38 |
| E117 | ENCODE2012 | CRVX.HELAS3.CNCR | HeLa-S3 Cervical Carcinoma Cell Line | 0.02 | 0.25 | 0.09 | 12.23 | 4.28 | 8.35E-03 | -7.80E-08 | 1.89E-07 | -0.41 |
| E088 | Other | LNG.FET | Fetal Lung | 0.02 | 0.25 | 0.09 | 13.65 | 5.04 | 8.65E-03 | 3.40E-08 | 2.51E-07 | 0.14 |
| E128 | ENCODE2012 | LNG.NHLF | NHLF Lung Fibroblast Primary Cells | 0.02 | 0.27 | 0.10 | 14.90 | 5.44 | 9.96E-03 | -1.04E-07 | 4.17E-07 | -0.25 |
| E086 | Other | KID.FET | Fetal Kidney | 0.01 | 0.24 | 0.09 | 17.02 | 6.68 | 1.08E-02 | 2.28E-07 | 3.55E-07 | 0.64 |
| E077 | Digestive | GI.DUO.MUC | Duodenum Mucosa | 0.02 | 0.26 | 0.10 | 11.60 | 4.57 | 1.33E-02 | -6.39E-07 | 3.20E-07 | -1.99 |
| E013 | ES-deriv | ESDR.CD56.MESO | hESC Derived CD56+ Mesoderm Cultured Cells | 0.02 | 0.25 | 0.09 | 12.84 | 4.90 | 1.36E-02 | 1.96E-07 | 2.55E-07 | 0.77 |
| E001 | ESC | ESC.I3 | ES-I3 Cells | 0.02 | 0.22 | 0.09 | 10.59 | 4.02 | 1.51E-02 | 6.02E-07 | 2.95E-07 | 2.04 |
| E103 | Sm. Muscle | GI.RECT.SM.MUS | Rectal Smooth Muscle | 0.02 | 0.23 | 0.09 | 12.21 | 4.82 | 1.59E-02 | -2.81E-08 | 3.65E-07 | -0.08 |
| E099 | Other | PLCNT.AMN | Placenta Amnion | 0.01 | 0.22 | 0.09 | 15.77 | 6.49 | 1.65E-02 | -3.39E-08 | 3.18E-07 | -0.11 |
| E081 | Brain | BRN.FET.M | Fetal Brain Male | 0.02 | 0.24 | 0.09 | 12.25 | 4.67 | 1.70E-02 | 1.68E-07 | 3.05E-07 | 0.55 |
| E095 | Heart | HRT.VENT.L | Left Ventricle | 0.02 | 0.21 | 0.08 | 12.48 | 4.99 | 1.84E-02 | 1.20E-07 | 3.42E-07 | 0.35 |
| E105 | Heart | HRT.VNT.R | Right Ventricle | 0.02 | 0.23 | 0.09 | 12.51 | 5.07 | 1.86E-02 | 2.80E-08 | 3.74E-07 | 0.07 |
| E083 | Heart | HRT.FET | Fetal Heart | 0.02 | 0.20 | 0.08 | 8.54 | 3.53 | 2.44E-02 | -8.68E-08 | 1.35E-07 | -0.65 |
| E092 | Digestive | GI.STMC.FET | Fetal Stomach | 0.02 | 0.25 | 0.10 | 11.47 | 4.87 | 2.57E-02 | -5.78E-09 | 2.64E-07 | -0.02 |
| E048 | Blood & T-cell | BLD.CD8.MPC | Primary T CD8+ memory cells from peripheral blood | 0.02 | 0.22 | 0.10 | 12.39 | 5.41 | 2.84E-02 | -8.67E-07 | 4.43E-07 | -1.96 |
| E071 | Brain | BRN.HIPP.MID | Brain Hippocampus Middle | 0.03 | 0.25 | 0.10 | 8.38 | 3.46 | 2.85E-02 | -3.30E-08 | 3.27E-07 | -0.10 |
| E104 | Heart | HRT.ATR.R | Right Atrium | 0.01 | 0.19 | 0.08 | 12.84 | 5.55 | 2.88E-02 | 1.22E-07 | 3.80E-07 | 0.32 |
| E118 | ENCODE2012 | LIV.HEPG2.CNCR | HepG2 Hepatocellular Carcinoma Cell Line | 0.02 | 0.20 | 0.09 | 10.82 | 4.70 | 3.59E-02 | -2.47E-07 | 1.95E-07 | -1.27 |
| E111 | Sm. Muscle | GI.STMC.MUS | Stomach Smooth Muscle | 0.02 | 0.20 | 0.09 | 10.12 | 4.60 | 3.77E-02 | -2.05E-07 | 3.10E-07 | -0.66 |
| E073 | Brain | BRN.DL.PRFRNTL.CRTX | Brain_Dorsolateral_Prefrontal_Cortex | 0.02 | 0.21 | 0.09 | 9.21 | 4.15 | 4.60E-02 | 2.88E-08 | 3.22E-07 | 0.09 |
| E005 | ES-deriv | ESDR.H1.BMP4.TROP | H1 BMP4 Derived Trophoblast Cultured Cells | 0.01 | 0.16 | 0.08 | 11.20 | 5.38 | 4.96E-02 | -1.52E-07 | 2.88E-07 | -0.53 |
| E026 | Mesench | STRM.MRW.MSC | Bone Marrow Derived Cultured Mesenchymal Stem Cells | 0.02 | 0.23 | 0.11 | 9.48 | 4.36 | 0.05 | -2.30E-07 | 3.19E-07 | -0.72 |
| E082 | Brain | BRN.FET.F | Fetal Brain Female | 0.01 | 0.17 | 0.08 | 12.90 | 6.28 | 0.05 | 1.62E-08 | 3.91E-07 | 0.04 |
| E012 | ES-deriv | ESDR.CD56.ECTO | hESC Derived CD56+ Ectoderm Cultured Cells | 0.02 | 0.15 | 0.07 | 9.36 | 4.73 | 0.07 | 4.10E-07 | 3.15E-07 | 1.30 |
| E022 | iPSC | IPSC.DF.19.11 | iPS DF 19.11 Cells | 0.01 | 0.14 | 0.07 | 11.07 | 5.65 | 0.07 | 1.02E-07 | 4.04E-07 | 0.25 |
| E070 | Brain | BRN.GRM.MTRX | Brain Germinal Matrix | 0.01 | 0.16 | 0.08 | 12.11 | 6.26 | 0.07 | -1.24E-07 | 4.13E-07 | -0.30 |
| E100 | Muscle | MUS.PSOAS | Psoas Muscle | 0.01 | 0.16 | 0.08 | 12.36 | 6.54 | 0.08 | -1.08E-07 | 3.48E-07 | -0.31 |
| E094 | Digestive | GI.STMC.GAST | Gastric | 0.01 | 0.14 | 0.08 | 13.84 | 7.38 | 0.08 | -4.59E-07 | 4.86E-07 | -0.95 |
| E096 | Other | LNG | Lung | 0.02 | 0.16 | 0.09 | 10.50 | 5.75 | 0.09 | -7.25E-07 | 3.86E-07 | -1.88 |
| E052 | Myosat | MUS.SAT | Muscle Satellite Cultured Cells | 0.03 | 0.21 | 0.11 | 8.37 | 4.45 | 0.09 | -3.42E-07 | 2.85E-07 | -1.20 |
| E067 | Brain | BRN.ANG.GYR | Brain Angular Gyrus | 0.02 | 0.18 | 0.09 | 7.51 | 3.97 | 0.10 | -1.14E-07 | 3.70E-07 | -0.31 |
| E121 | ENCODE2012 | MUS.HSMMT | HSMM cell derived Skeletal Muscle Myotubes Cells | 0.02 | 0.18 | 0.10 | 8.75 | 4.75 | 0.10 | -3.97E-07 | 2.62E-07 | -1.52 |
| E087 | Other | PANC.ISLT | Pancreatic Islets | 0.01 | 0.13 | 0.08 | 10.83 | 6.30 | 0.11 | -4.88E-07 | 3.97E-07 | -1.23 |
| E007 | ES-deriv | ESDR.H1.NEUR.PROG | H1 Derived Neuronal Progenitor Cultured Cells | 0.01 | 0.12 | 0.07 | 12.80 | 7.69 | 0.11 | 2.83E-07 | 4.85E-07 | 0.58 |
| E102 | Digestive | GI.RECT.MUC.31 | Rectal Mucosa Donor 31 | 0.02 | 0.17 | 0.10 | 8.78 | 5.07 | 0.12 | -1.05E-06 | 4.66E-07 | -2.25 |
| E097 | Other | OVRY | Ovary | 0.01 | 0.13 | 0.08 | 11.28 | 6.87 | 0.12 | -1.38E-07 | 3.14E-07 | -0.44 |
| E053 | Neurosph | BRN.CRTX.DR.NRSPHR | Cortex derived primary cultured neurospheres | 0.02 | 0.14 | 0.09 | 9.20 | 5.50 | 0.13 | -4.83E-07 | 3.67E-07 | -1.32 |
| E003 | ESC | ESC.H1 | H1 Cells | 0.02 | 0.12 | 0.07 | 7.37 | 4.71 | 0.17 | 3.69E-07 | 4.28E-07 | 0.86 |
| E072 | Brain | BRN.INF.TMP | Brain Inferior Temporal Lobe | 0.03 | 0.14 | 0.09 | 5.51 | 3.44 | 0.18 | -3.49E-07 | 3.84E-07 | -0.91 |
| E065 | Heart | VAS.AOR | Aorta | 0.01 | 0.09 | 0.07 | 8.38 | 6.18 | 0.22 | -3.86E-07 | 3.34E-07 | -1.16 |
| E069 | Brain | BRN.CING.GYR | Brain Cingulate Gyrus | 0.03 | 0.13 | 0.10 | 5.06 | 3.75 | 0.27 | -4.72E-07 | 3.90E-07 | -1.21 |
| E008 | ESC | ESC.H9 | H9 Cells | 0.01 | 0.09 | 0.08 | 6.92 | 5.61 | 0.27 | 1.03E-07 | 4.37E-07 | 0.24 |
| E024 | ESC | ESC.4STAR | ES-UCSF4 Cells | 0.01 | 0.09 | 0.07 | 6.29 | 5.04 | 0.29 | -1.10E-07 | 3.65E-07 | -0.30 |
| E021 | iPSC | IPSC.DF.6.9 | iPS DF 6.9 Cells | 0.01 | 0.07 | 0.06 | 7.37 | 6.42 | 0.31 | -1.30E-07 | 5.39E-07 | -0.24 |
| E125 | ENCODE2012 | BRN.NHA | NH-A Astrocytes Primary Cells | 0.02 | 0.12 | 0.10 | 6.00 | 5.04 | 0.33 | -8.24E-07 | 3.09E-07 | -2.67 |
| E019 | iPSC | IPSC.18 | iPS-18 Cells | 0.02 | 0.10 | 0.08 | 5.13 | 4.48 | 0.35 | 2.19E-07 | 3.74E-07 | 0.59 |
| E089 | Muscle | MUS.TRNK.FET | Fetal Muscle Trunk | 0.02 | 0.11 | 0.10 | 5.11 | 4.49 | 0.36 | -7.80E-07 | 3.82E-07 | -2.04 |
| E014 | ESC | ESC.HUES48 | HUES48 Cells | 0.02 | 0.09 | 0.08 | 4.77 | 4.35 | 0.38 | 1.62E-07 | 4.19E-07 | 0.39 |
| E018 | iPSC | IPSC.15b | iPS-15b Cells | 0.02 | 0.07 | 0.08 | 4.19 | 4.42 | 0.47 | -1.27E-07 | 3.73E-07 | -0.34 |
| E002 | ESC | ESC.WA7 | ES-WA7 Cells | 0.01 | 0.06 | 0.07 | 5.29 | 6.46 | 0.50 | -2.51E-07 | 4.62E-07 | -0.54 |
| E009 | ES-deriv | ESDR.H9.NEUR.PROG | H9 Derived Neuronal Progenitor Cultured Cells | 0.01 | 0.07 | 0.08 | 4.69 | 5.46 | 0.50 | -6.17E-08 | 3.89E-07 | -0.16 |
| E011 | ES-deriv | ESDR.CD184.ENDO | hESC Derived CD184+ Endoderm Cultured Cells | 0.02 | 0.05 | 0.08 | 3.12 | 4.97 | 0.67 | -2.23E-07 | 2.89E-07 | -0.77 |
| E016 | ESC | ESC.HUES64 | HUES64 Cells | 0.02 | 0.04 | 0.07 | 2.56 | 4.27 | 0.71 | -2.01E-07 | 3.97E-07 | -0.51 |
| E010 | ES-deriv | ESDR.H9.NEUR | H9 Derived Neuron Cultured Cells | 0.02 | -0.01 | 0.08 | -0.65 | 4.95 | 0.74 | -6.16E-07 | 3.46E-07 | -1.78 |
| E015 | ESC | ESC.HUES6 | HUES6 Cells | 0.02 | 0.04 | 0.08 | 1.85 | 4.06 | 0.83 | -4.19E-07 | 3.46E-07 | -1.21 |
| E004 | ES-deriv | ESDR.H1.BMP4.MESO | H1 BMP4 Derived Mesendoderm Cultured Cells | 0.01 | 0.02 | 0.07 | 1.56 | 5.11 | 0.91 | -4.33E-07 | 3.22E-07 | -1.35 |
| E020 | iPSC | IPSC.20B | iPS-20b Cells | 0.02 | 0.02 | 0.08 | 1.23 | 4.61 | 0.96 | -3.94E-07 | 3.98E-07 | -0.99 |

**Table S16. Top ranked tissues and cell types based on DEPICT enrichment.** The analysis was performed for 209 Medical Subject Heading (MeSH) tissue- and cell-type annotations, only results at nominal P<0.05 are displayed and sorted by the level of statistical significance, the results meeting FDR cut-off <5% are flagged (top part, graphically depicted in Figure 3c).

| MeSH term | MeSH first level term | MeSH second level term | Nominal P value | False discovery rate < 5% |
|---|---|---|---|---|
| A15.378.316 | Bone Marrow Cells | Hemic and Immune Systems | 4.90E-06 | Yes |
| A15.378 | Hematopoietic System | Hemic and Immune Systems | 4.90E-06 | Yes |
| A11.118.637.415 | Granulocytes | Cells | 5.88E-06 | Yes |
| A15.382.490.315.583 | Neutrophils | Hemic and Immune Systems | 6.31E-06 | Yes |
| A15.145 | Blood | Hemic and Immune Systems | 6.66E-06 | Yes |
| A11.627 | Myeloid Cells | Cells | 1.11E-05 | Yes |
| A15.145.229 | Blood Cells | Hemic and Immune Systems | 1.30E-05 | Yes |
| A15.382.680 | Phagocytes | Hemic and Immune Systems | 1.64E-05 | Yes |
| A11.118.637 | Leukocytes | Cells | 2.07E-05 | Yes |
| A15.145.300 | Fetal Blood | Hemic and Immune Systems | 9.39E-05 | Yes |
| A15.382.520.604.700 | Spleen | Hemic and Immune Systems | 1.51E-04 | Yes |
| A02.835.583.443.800.800 | Synovial Fluid | Musculoskeletal System | 1.56E-04 | Yes |
| A15.378.316.580 | Monocytes | Hemic and Immune Systems | 2.24E-04 | Yes |
| A15.382.812 | Mononuclear Phagocyte System | Hemic and Immune Systems | 2.77E-04 | Yes |
| A15.382 | Immune System | Hemic and Immune Systems | 3.02E-04 | Yes |
| A11.066 | Antigen Presenting Cells | Cells | 4.26E-04 | Yes |
| A15.382.812.260 | Dendritic Cells | Hemic and Immune Systems | 4.26E-04 | Yes |
| A15.382.520 | Lymphatic System | Hemic and Immune Systems | 4.92E-04 | Yes |
| A10.549 | Lymphoid Tissue | Tissues | 4.92E-04 | Yes |
| A10.549.400 | Lymph Nodes | Tissues | 6.11E-04 | Yes |
| A15.382.812.522 | Macrophages | Hemic and Immune Systems | 6.40E-04 | Yes |
| A02.835.232.043 | Bones of Lower Extremity | Musculoskeletal System | 1.21E-03 | Yes |
| A02.835.232.043.300.710 | Tarsal Bones | Musculoskeletal System | 1.35E-03 | Yes |
| A02.835.232.043.300 | Foot Bones | Musculoskeletal System | 1.35E-03 | Yes |
| A11.872.378.590.635 | Granulocyte Macrophage Progenitor Cells | Cells | 2.03E-03 | Yes |
| A15.145.229.637.555 | Leukocytes Mononuclear | Hemic and Immune Systems | 2.66E-03 | Yes |
| A09.371.060 | Anterior Eye Segment | Sense Organs | 2.66E-03 | Yes |
| A09.371.337.168 | Conjunctiva | Sense Organs | 2.67E-03 | Yes |
| A09.371.337 | Eyelids | Sense Organs | 2.67E-03 | Yes |
| A11.627.340.360 | Granulocyte Precursor Cells | Cells | 4.55E-03 | Yes |
| A11.118.637.555.567.569 | T Lymphocytes | Cells | 9.17E-03 | No |
| A11.627.624.249 | Monocyte Macrophage Precursor Cells | Cells | 1.09E-02 | No |
| A11.872.378 | Hematopoietic Stem Cells | Cells | 1.10E-02 | No |
| A11.627.635 | Myeloid Progenitor Cells | Cells | 1.48E-02 | No |
| A15.382.520.604.800 | Palatine Tonsil | Hemic and Immune Systems | 1.52E-02 | No |
| A04.623.603 | Oropharynx | Respiratory System | 1.52E-02 | No |
| A15.382.216 | Bone Marrow | Hemic and Immune Systems | 1.96E-02 | No |
| A14.724 | Pharynx | Stomatognathic System | 2.01E-02 | No |
| A10.165 | Connective Tissue | Tissues | 2.33E-02 | No |
| A02.835 | Skeleton | Musculoskeletal System | 2.34E-02 | No |
| A02.835.232 | Bone and Bones | Musculoskeletal System | 2.36E-02 | No |
| A15.382.490.555.567.537 | Natural Killer Cells | Hemic and Immune Systems | 2.76E-02 | No |
| A11.329.372.600 | Macrophages Alveolar | Cells | 2.82E-02 | No |
| A03.556.249.124 | Ileum | Digestive System | 3.06E-02 | No |
| A03.556.124.684 | Intestine Small | Digestive System | 3.17E-02 | No |
| A03.556.124.526.767 | Rectum | Digestive System | 3.25E-02 | No |
| A03.556.249.249.209 | Cecum | Digestive System | 3.62E-02 | No |
| A03.556.249 | Lower Gastrointestinal Tract | Digestive System | 3.84E-02 | No |
| A11.872.378.294 | Lymphoid Progenitor Cells | Cells | 4.15E-02 | No |
| A11.118.637.555.567.562.440 | Precursor Cells B Lymphoid | Cells | 4.15E-02 | No |
| A03.556.249.249 | Intestine Large | Digestive System | 4.40E-02 | No |
| A03.556.249.249.356 | Colon | Digestive System | 4.69E-02 | No |

**Table S17. Top ranked tissues and cell types from GARFIELD enrichment analysis.** Enrichment in tissue and cell-type specific ENCODE and Roadmap functional annotations at GWAS P-value threshold $T<1.0x10^{-5}$. Results sorted by significance level and only the top 100 enrichments are depicted; the complete analysis involves 2013 functional annotations across all Roadmap and ENCODE tissues and cell types. Hotspots refer to DNase I–hypersensitive sites. The top scoring cell types across multiple annotations include immune, blood, intestinal mucosa, thymic, and splenic cells.

| Cell type | Tissue | Type | Category | OR | Beta | SE | P-value |
|---|---|---|---|---|---|---|---|
| GM12878 | blood | H3K27ac | Histone_Modifications | 5.33 | 1.67 | 0.14 | 4.33E-32 |
| CD19_Primary_Cells | blood | hotspots | Hotspots | 4.99 | 1.61 | 0.15 | 4.41E-28 |
| GM06990 | blood | hotspots | Hotspots | 4.71 | 1.55 | 0.15 | 1.53E-25 |
| CD56_Primary_Cells | blood | hotspots | Hotspots | 4.66 | 1.54 | 0.15 | 2.38E-24 |
| CD3_Primary_Cells | blood | hotspots | Hotspots | 4.39 | 1.48 | 0.15 | 3.96E-23 |
| CD19_Primary_Cells | blood | hotspots | Hotspots | 4.30 | 1.46 | 0.15 | 4.83E-23 |
| GM12878 | blood | H3K4me2 | Histone_Modifications | 4.09 | 1.41 | 0.14 | 4.86E-23 |
| CD20+ | blood | hotspots | Hotspots | 4.19 | 1.43 | 0.15 | 5.86E-23 |
| GM12878 | blood | H3K9ac | Histone_Modifications | 4.29 | 1.46 | 0.15 | 1.74E-22 |
| GM12878 | blood | H3K4me1 | Histone_Modifications | 3.82 | 1.34 | 0.14 | 3.67E-22 |
| CD14_Primary_Cells | blood | peaks | Peaks | 5.22 | 1.65 | 0.17 | 1.51E-21 |
| CD4_Primary_Cells | blood | hotspots | Hotspots | 4.06 | 1.40 | 0.15 | 5.74E-21 |
| GM12878 | blood | H3K4me3 | Histone_Modifications | 3.96 | 1.38 | 0.15 | 5.89E-21 |
| CD8_Primary_Cells | blood | hotspots | Hotspots | 4.04 | 1.40 | 0.15 | 2.14E-20 |
| CD19_Primary_Cells | blood | hotspots | Hotspots | 3.92 | 1.37 | 0.15 | 2.84E-20 |
| CD3_Primary_Cells | blood | hotspots | Hotspots | 4.01 | 1.39 | 0.15 | 8.27E-20 |
| CD56_Primary_Cells | blood | hotspots | Hotspots | 3.91 | 1.36 | 0.15 | 9.58E-20 |
| GM12878 | blood | ENHANCER | Chromatin_States | 4.41 | 1.48 | 0.17 | 2.88E-19 |
| CD8_Primary_Cells | blood | hotspots | Hotspots | 3.62 | 1.29 | 0.15 | 2.25E-18 |
| CD14+ | blood | hotspots | Hotspots | 3.55 | 1.27 | 0.15 | 5.77E-18 |
| GM12878 | blood | TSS | Chromatin_States | 4.15 | 1.42 | 0.17 | 6.52E-18 |
| GM12878 | blood | H3K27ac | Histone_Modifications | 7.71 | 2.04 | 0.24 | 1.83E-17 |
| CD4_Primary_Cells | blood | hotspots | Hotspots | 3.56 | 1.27 | 0.15 | 1.97E-17 |
| GM12878 | blood | H3K79me2 | Histone_Modifications | 3.40 | 1.22 | 0.15 | 4.96E-17 |
| CD14_Primary_Cells | blood | peaks | Peaks | 4.33 | 1.46 | 0.18 | 6.58E-17 |
| CD4_Primary_Cells | blood | hotspots | Hotspots | 3.40 | 1.22 | 0.15 | 7.89E-17 |
| CD3_Primary_Cells | blood | hotspots | Hotspots | 3.50 | 1.25 | 0.15 | 1.86E-16 |
| GM12865 | blood | hotspots | Hotspots | 3.42 | 1.23 | 0.15 | 2.51E-16 |
| GM12864 | blood | hotspots | Hotspots | 3.42 | 1.23 | 0.15 | 4.07E-16 |
| GM12878 | blood | H2AFZ | Histone_Modifications | 3.37 | 1.22 | 0.15 | 4.20E-16 |
| Mobilized_CD4_Primary_Cells | blood | hotspots | Hotspots | 3.39 | 1.22 | 0.15 | 4.36E-16 |
| Fetal_Intestine_Large | fetal_intestine,_large | hotspots | Hotspots | 3.46 | 1.24 | 0.15 | 4.69E-16 |
| CD14_Primary_Cells | blood | hotspots | Hotspots | 3.46 | 1.24 | 0.15 | 6.37E-16 |
| CD4_Primary_Cells | blood | hotspots | Hotspots | 3.37 | 1.21 | 0.15 | 9.36E-16 |
| Fetal_Thymus | fetal_thymus | hotspots | Hotspots | 3.30 | 1.20 | 0.15 | 1.13E-15 |
| CD14_Primary_Cells | blood | hotspots | Hotspots | 3.35 | 1.21 | 0.15 | 1.40E-15 |
| CD8_Primary_Cells | blood | hotspots | Hotspots | 3.32 | 1.20 | 0.15 | 1.46E-15 |
| CD20+ | blood | peaks | Peaks | 4.13 | 1.42 | 0.18 | 2.00E-15 |
| CD3_Primary_Cells | blood | peaks | Peaks | 4.18 | 1.43 | 0.18 | 5.79E-15 |
| Mobilized_CD4_Primary_Cells | blood | peaks | Peaks | 4.10 | 1.41 | 0.18 | 6.52E-15 |
| Mobilized_CD4_Primary_Cells | blood | hotspots | Hotspots | 3.14 | 1.14 | 0.15 | 6.85E-15 |
| CD8_Primary_Cells | blood | hotspots | Hotspots | 3.30 | 1.19 | 0.15 | 7.78E-15 |
| GM12865 | blood | footprints | Footprints | 4.26 | 1.45 | 0.19 | 1.04E-14 |
| CD19_Primary_Cells | blood | hotspots | Hotspots | 6.85 | 1.92 | 0.25 | 1.08E-14 |
| CD19_Primary_Cells | blood | peaks | Peaks | 4.07 | 1.40 | 0.18 | 1.96E-14 |
| Mobilized_CD34_Primary_Cells | blood | hotspots | Hotspots | 3.24 | 1.18 | 0.15 | 1.98E-14 |
| GM12878 | blood | tfbs | TFBS | 3.39 | 1.22 | 0.16 | 2.66E-14 |
| Mobilized_CD34_Primary_Cells | blood | hotspots | Hotspots | 3.17 | 1.15 | 0.15 | 3.80E-14 |
| Mobilized_CD56_Primary_Cells | blood | hotspots | Hotspots | 3.09 | 1.13 | 0.15 | 4.78E-14 |
| GM12878 | blood | hotspots | Hotspots | 3.00 | 1.10 | 0.15 | 6.32E-14 |
| Fetal_Spleen | fetal_spleen | hotspots | Hotspots | 3.02 | 1.11 | 0.15 | 1.39E-13 |
| Mobilized_CD34_Primary_Cells | blood | hotspots | Hotspots | 3.08 | 1.13 | 0.15 | 1.45E-13 |
| GM12878 | blood | H3K4me2 | Histone_Modifications | 6.00 | 1.79 | 0.24 | 1.58E-13 |
| HeLa-S3 | cervix | H3K27me3 | Histone_Modifications | 5.13 | 1.63 | 0.22 | 1.63E-13 |
| Fetal_Thymus | fetal_thymus | hotspots | Hotspots | 2.98 | 1.09 | 0.15 | 1.92E-13 |
| Fetal_Intestine_Small | fetal_intestine,_small | hotspots | Hotspots | 2.95 | 1.08 | 0.15 | 4.09E-13 |
| Mobilized_CD4_Primary_Cells | blood | peaks | Peaks | 3.67 | 1.30 | 0.18 | 6.56E-13 |
| Mobilized_CD34_Primary_Cells | blood | hotspots | Hotspots | 2.98 | 1.09 | 0.15 | 7.04E-13 |
| GM12864 | blood | peaks | Peaks | 3.55 | 1.27 | 0.18 | 7.17E-13 |
| Mobilized_CD8_Primary_Cells | blood | hotspots | Hotspots | 2.93 | 1.07 | 0.15 | 8.93E-13 |
| HeLa-S3 | cervix | H3K27me3 | Histone_Modifications | 2.61 | 0.96 | 0.13 | 1.14E-12 |
| CD34+ | blood | hotspots | Hotspots | 2.84 | 1.04 | 0.15 | 1.32E-12 |
| Th2 | blood | hotspots | Hotspots | 2.95 | 1.08 | 0.15 | 1.44E-12 |
| Mobilized_CD34_Primary_Cells | blood | hotspots | Hotspots | 2.94 | 1.08 | 0.15 | 1.61E-12 |
| Th1 | blood | hotspots | Hotspots | 2.81 | 1.03 | 0.15 | 1.68E-12 |
| Mobilized_CD34_Primary_Cells | blood | hotspots | Hotspots | 2.95 | 1.08 | 0.15 | 2.29E-12 |
| CD8_Primary_Cells | blood | peaks | Peaks | 3.71 | 1.31 | 0.19 | 2.75E-12 |
| Mobilized_CD34_Primary_Cells | blood | hotspots | Hotspots | 2.89 | 1.06 | 0.15 | 3.11E-12 |
| GM06990 | blood | hotspots | Hotspots | 5.80 | 1.76 | 0.25 | 3.97E-12 |
| CD19_Primary_Cells | blood | peaks | Peaks | 3.57 | 1.27 | 0.18 | 5.17E-12 |
| CD34+ | blood | footprints | Footprints | 3.64 | 1.29 | 0.19 | 6.76E-12 |
| GM12878 | blood | H3K4me1 | Histone_Modifications | 4.95 | 1.60 | 0.23 | 9.08E-12 |
| CD56_Primary_Cells | blood | peaks | Peaks | 3.61 | 1.28 | 0.19 | 1.47E-11 |
| CD8_Primary_Cells | blood | peaks | Peaks | 3.49 | 1.25 | 0.19 | 1.64E-11 |
| GM06990 | blood | peaks | Peaks | 3.60 | 1.28 | 0.19 | 1.70E-11 |
| Mobilized_CD34_Primary_Cells | blood | hotspots | Hotspots | 2.79 | 1.03 | 0.15 | 2.01E-11 |
| Mobilized_CD34_Primary_Cells | blood | hotspots | Hotspots | 2.76 | 1.01 | 0.15 | 2.64E-11 |
| Mobilized_CD34_Primary_Cells | blood | hotspots | Hotspots | 2.77 | 1.02 | 0.15 | 3.20E-11 |
| CD3_Primary_Cells | blood | hotspots | Hotspots | 2.67 | 0.98 | 0.15 | 4.05E-11 |
| Mobilized_CD34_Primary_Cells | blood | hotspots | Hotspots | 2.71 | 1.00 | 0.15 | 4.18E-11 |
| Fetal_Thymus | fetal_thymus | hotspots | Hotspots | 2.73 | 1.00 | 0.15 | 4.75E-11 |
| CD19_Primary_Cells | blood | peaks | Peaks | 3.45 | 1.24 | 0.19 | 5.02E-11 |
| Fetal_Thymus | fetal_thymus | hotspots | Hotspots | 2.67 | 0.98 | 0.15 | 5.75E-11 |
| CD3_Primary_Cells | blood | hotspots | Hotspots | 5.31 | 1.67 | 0.26 | 6.56E-11 |
| Mobilized_CD34_Primary_Cells | blood | hotspots | Hotspots | 2.66 | 0.98 | 0.15 | 6.84E-11 |
| Fetal_Thymus | fetal_thymus | peaks | Peaks | 3.26 | 1.18 | 0.18 | 7.09E-11 |
| Mobilized_CD34_Primary_Cells | blood | peaks | Peaks | 3.18 | 1.16 | 0.18 | 7.30E-11 |
| Th2 | blood | peaks | Peaks | 3.48 | 1.25 | 0.19 | 8.26E-11 |
| GM18507 | blood | peaks | Peaks | 3.28 | 1.19 | 0.18 | 8.84E-11 |
| GM12878 | blood | H3K9ac | Histone_Modifications | 5.20 | 1.65 | 0.25 | 9.62E-11 |
| CD20+ | blood | footprints | Footprints | 3.82 | 1.34 | 0.21 | 1.17E-10 |
| CD3_Primary_Cells | blood | peaks | Peaks | 3.37 | 1.22 | 0.19 | 1.19E-10 |
| Mobilized_CD56_Primary_Cells | blood | peaks | Peaks | 3.25 | 1.18 | 0.18 | 1.45E-10 |
| NB4 | blood | footprints | Footprints | 3.31 | 1.20 | 0.19 | 1.56E-10 |
| CD34+ | blood | peaks | Peaks | 3.05 | 1.11 | 0.17 | 1.80E-10 |
| GM12878 | blood | H3K4me3 | Histone_Modifications | 4.90 | 1.59 | 0.25 | 2.20E-10 |
| Fetal_Thymus | fetal_thymus | hotspots | Hotspots | 2.63 | 0.97 | 0.15 | 2.29E-10 |
| HUVEC | blood_vessel | H3K27me3 | Histone_Modifications | 2.45 | 0.90 | 0.14 | 2.83E-10 |
| CD56_Primary_Cells | blood | hotspots | Hotspots | 5.15 | 1.64 | 0.26 | 3.09E-10 |

**Table S18. Regulatory Element Locus Intersection (RELI) analysis of IgAN GWAS loci against transcription factor binding sites: (a)** limited to genome-wide significant GWAS loci (28 loci), and **(b)** extended to genome-wide significant and suggestive GWAS loci (76 loci). Enrichment statistics and corrected P-values as described in Harley JB et al. (*Nat Gen* 2018)[19].

**(a) Significant GWAS loci (total of 28 loci)**

| Track | Cell | TF | Overlap | Total | Ratio | Mean | STD | Z-score | Enrichment | P-val | Corrected P | Species |
|---|---|---|---|---|---|---|---|---|---|---|---|---|
| ReMap_ChIP | lclgm12878 | NFKB1 | 14 | 28 | 0.50 | 2.13 | 1.39 | 8.52 | 6.57 | 8.15E-18 | 1.26E-14 | human |
| TxnFactorChIPV3 | H1-hESC | RFX5 | 3 | 28 | 0.11 | 0.17 | 0.42 | 6.76 | 17.35 | 7.07E-12 | 1.09E-08 | human |
| TxnFactorChIPV3 | GM12878 | ATF2 | 10 | 28 | 0.36 | 1.86 | 1.31 | 6.20 | 5.39 | 2.86E-10 | 4.41E-07 | human |
| TxnFactorChIPV3 | GM18505 | POLR2A | 11 | 28 | 0.39 | 2.31 | 1.41 | 6.17 | 4.76 | 3.33E-10 | 5.14E-07 | human |
| TxnFactorChIPV3 | GM12878 | RUNX3 | 14 | 28 | 0.50 | 3.50 | 1.70 | 6.16 | 4.00 | 3.63E-10 | 5.61E-07 | human |
| ReMap_ChIP | lclgm18951 | NFKB1 | 6 | 28 | 0.21 | 0.78 | 0.87 | 5.97 | 7.66 | 1.19E-09 | 1.84E-06 | human |
| TxnFactorChIPV3 | GM12878 | POLR2A | 12 | 28 | 0.43 | 2.75 | 1.56 | 5.94 | 4.36 | 1.45E-09 | 2.24E-06 | human |
| ReMap_ChIP | cd4_il12 | STAT4 | 2 | 28 | 0.07 | 0.10 | 0.32 | 5.89 | 19.15 | 1.94E-09 | 2.99E-06 | human |
| TxnFactorChIPV3 | GM12878 | CEBPB | 5 | 28 | 0.18 | 0.58 | 0.77 | 5.76 | 8.66 | 4.15E-09 | 6.41E-06 | human |
| TxnFactorChIPV3 | GM18505+TNFa | RELA | 6 | 28 | 0.21 | 0.82 | 0.90 | 5.74 | 7.32 | 4.87E-09 | 7.53E-06 | human |
| TxnFactorChIPV3 | GM12892 | POLR2A | 11 | 28 | 0.39 | 2.54 | 1.48 | 5.72 | 4.32 | 5.28E-09 | 8.15E-06 | human |
| TxnFactorChIPV3 | GM12878+TNFa | RELA | 8 | 28 | 0.29 | 1.42 | 1.15 | 5.71 | 5.64 | 5.51E-09 | 8.51E-06 | human |
| TxnFactorChIPV3 | GM12878 | TBL1XR1 | 7 | 28 | 0.25 | 1.10 | 1.05 | 5.63 | 6.34 | 9.26E-09 | 1.43E-05 | human |
| ReMap_ChIP | B-cell | SMARCA4 | 2 | 28 | 0.07 | 0.12 | 0.34 | 5.52 | 17.10 | 1.68E-08 | 2.60E-05 | human |

**(b) Significant and suggestive loci (total of 76 loci)**

| Track | Cell | TF | Overlap | Total | Ratio | Mean | STD | Z-score | Enrichment | P-val | Corrected P | Species |
|---|---|---|---|---|---|---|---|---|---|---|---|---|
| TxnFactorChIPV3 | GM12878+TNFa | RELA | 19 | 76 | 0.25 | 3.98 | 1.86 | 8.07 | 4.78 | 3.58E-16 | 5.53E-13 | human |
| ReMap_ChIP | lclgm12878 | NFKB1 | 24 | 76 | 0.32 | 5.92 | 2.28 | 7.91 | 4.06 | 1.24E-15 | 1.92E-12 | human |
| TxnFactorChIPV3 | GM12878 | ATF2 | 21 | 76 | 0.28 | 5.15 | 2.08 | 7.62 | 4.08 | 1.27E-14 | 1.96E-11 | human |
| TxnFactorChIPV3 | GM12878 | FOXM1 | 20 | 76 | 0.26 | 5.00 | 2.02 | 7.43 | 4.00 | 5.43E-14 | 8.38E-11 | human |
| TxnFactorChIPV3 | GM12878 | CEBPB | 11 | 76 | 0.14 | 1.67 | 1.28 | 7.28 | 6.57 | 1.64E-13 | 2.53E-10 | human |
| TxnFactorChIPV3 | GM12878 | TBL1XR1 | 15 | 76 | 0.20 | 3.06 | 1.68 | 7.13 | 4.91 | 5.06E-13 | 7.82E-10 | human |
| TxnFactorChIPV3 | GM12878 | MTA3 | 15 | 76 | 0.20 | 3.20 | 1.68 | 7.02 | 4.69 | 1.13E-12 | 1.74E-09 | human |
| ReMap_ChIP | lclgm18951 | NFKB1 | 12 | 76 | 0.16 | 2.15 | 1.41 | 6.99 | 5.57 | 1.41E-12 | 2.18E-09 | human |
| Cistrome_ChIP | CD14+ | IRF1 | 23 | 76 | 0.30 | 6.47 | 2.39 | 6.90 | 3.55 | 2.52E-12 | 3.89E-09 | human |
| TxnFactorChIPV3 | GM12878 | SPI1 | 22 | 76 | 0.29 | 6.49 | 2.31 | 6.73 | 3.39 | 8.68E-12 | 1.34E-08 | human |
| Misc_ChIP | GM12878 | p52 | 19 | 76 | 0.25 | 4.93 | 2.11 | 6.65 | 3.85 | 1.43E-11 | 2.20E-08 | human |
| TxnFactorChIPV3 | GM12891 | POLR2A | 22 | 76 | 0.29 | 6.69 | 2.31 | 6.62 | 3.29 | 1.81E-11 | 2.79E-08 | human |
| Misc_ChIP | MutuIII | EBNA2 | 23 | 76 | 0.30 | 6.81 | 2.48 | 6.54 | 3.38 | 3.08E-11 | 4.75E-08 | EBV |
| TxnFactorChIPV3 | GM18505+TNFa | RELA | 12 | 76 | 0.16 | 2.36 | 1.50 | 6.41 | 5.09 | 7.30E-11 | 1.13E-07 | human |
| TxnFactorChIPV3 | GM18505 | POLR2A | 21 | 76 | 0.28 | 6.29 | 2.30 | 6.40 | 3.34 | 7.89E-11 | 1.22E-07 | human |
| TxnFactorChIPV3 | GM12878 | POLR2A | 24 | 76 | 0.32 | 7.71 | 2.56 | 6.36 | 3.11 | 1.03E-10 | 1.59E-07 | human |
| Cistrome_ChIP | GM12878 | NFATC1 | 21 | 76 | 0.28 | 6.57 | 2.29 | 6.30 | 3.20 | 1.51E-10 | 2.33E-07 | human |
| Cistrome_ChIP | GM12878 | STAT5A | 23 | 76 | 0.30 | 7.41 | 2.48 | 6.29 | 3.10 | 1.54E-10 | 2.38E-07 | human |
| TxnFactorChIPV3 | GM12878 | PAX5 | 18 | 76 | 0.24 | 5.15 | 2.05 | 6.26 | 3.49 | 1.90E-10 | 2.93E-07 | human |
| Pazar_Chip | CD4+ | HMGN1 | 38 | 76 | 0.50 | 17.37 | 3.32 | 6.21 | 2.19 | 2.66E-10 | 4.11E-07 | human |
| ReMap_ChIP | lclgm12891 | NFKB1 | 22 | 76 | 0.29 | 7.05 | 2.41 | 6.21 | 3.12 | 2.67E-10 | 4.13E-07 | human |
| Misc_ChIP | GM12878 | RelB | 19 | 76 | 0.25 | 5.48 | 2.18 | 6.20 | 3.47 | 2.85E-10 | 4.41E-07 | human |
| ReMap_ChIP | lclgm10861_calcitriol | VDR | 15 | 76 | 0.20 | 3.74 | 1.83 | 6.16 | 4.01 | 3.65E-10 | 5.64E-07 | human |
| TxnFactorChIPV3 | GM12891 | SPI1 | 22 | 76 | 0.29 | 7.09 | 2.42 | 6.15 | 3.10 | 3.77E-10 | 5.81E-07 | human |
| Cistrome_ChIP | Monocyte_CD14+ | IRF1 | 13 | 76 | 0.17 | 2.99 | 1.66 | 6.04 | 4.34 | 7.69E-10 | 1.19E-06 | human |
| Misc_ChIP | BCells+Sendai_Virus | IRF3 | 13 | 76 | 0.17 | 2.92 | 1.68 | 6.00 | 4.45 | 9.83E-10 | 1.52E-06 | human |
| TxnFactorChIPV3 | GM12878 | RUNX3 | 25 | 76 | 0.33 | 9.27 | 2.64 | 5.95 | 2.70 | 1.32E-09 | 2.04E-06 | human |
| Cistrome_ChIP | GM12878 | PML | 22 | 76 | 0.29 | 7.20 | 2.50 | 5.93 | 3.06 | 1.52E-09 | 2.34E-06 | human |
| TxnFactorChIPV3 | GM12878 | NFATC1 | 13 | 76 | 0.17 | 3.00 | 1.69 | 5.92 | 4.34 | 1.62E-09 | 2.51E-06 | human |
| Pazar_Chip | U937+BMP4 | SMAD1 | 17 | 76 | 0.22 | 4.74 | 2.08 | 5.89 | 3.59 | 1.98E-09 | 3.06E-06 | human |
| Artem-ChIP | CD4+_T_cells+invitrogen | POLR2A | 25 | 76 | 0.33 | 9.21 | 2.69 | 5.87 | 2.71 | 2.15E-09 | 3.31E-06 | human |
| Cistrome_ChIP | OCI-Ly10 | IRF4 | 8 | 76 | 0.11 | 1.37 | 1.13 | 5.87 | 5.84 | 2.18E-09 | 3.37E-06 | human |
| TxnFactorChIPV3 | GM12878 | NFIC | 19 | 76 | 0.25 | 5.93 | 2.24 | 5.84 | 3.20 | 2.67E-09 | 4.12E-06 | human |
| Artem-ChIP | Naive_CD4_Tcell_EI_5h_6 | NFATC2 | 21 | 76 | 0.28 | 6.91 | 2.43 | 5.80 | 3.04 | 3.24E-09 | 5.01E-06 | human |
| Misc_ChIP | Kasumi-1 | RUNX1 | 14 | 76 | 0.18 | 3.53 | 1.81 | 5.77 | 3.97 | 3.87E-09 | 5.97E-06 | human |
| TxnFactorChIPV3 | GM12878 | BCL3 | 13 | 76 | 0.17 | 3.23 | 1.69 | 5.76 | 4.02 | 4.12E-09 | 6.37E-06 | human |
| Cistrome_ChIP | GM12891 | SPI1 | 20 | 76 | 0.26 | 6.60 | 2.35 | 5.70 | 3.03 | 5.95E-09 | 9.18E-06 | human |
| Cistrome_ChIP | CD14+ | STAT1 | 16 | 76 | 0.21 | 4.70 | 2.00 | 5.66 | 3.40 | 7.77E-09 | 1.20E-05 | human |
| TxnFactorChIPV3 | GM12878 | EBF1 | 21 | 76 | 0.28 | 7.27 | 2.43 | 5.65 | 2.89 | 7.80E-09 | 1.20E-05 | human |
| Pazar_Chip | CD34+prog | SMAD1 | 12 | 76 | 0.16 | 2.91 | 1.61 | 5.63 | 4.13 | 8.81E-09 | 1.36E-05 | human |
| Cistrome_ChIP | GM12878 | SP1 | 20 | 76 | 0.26 | 6.65 | 2.38 | 5.61 | 3.01 | 9.90E-09 | 1.53E-05 | human |
| TxnFactorChIPV3 | GM12878 | EP300 | 15 | 76 | 0.20 | 4.20 | 1.94 | 5.58 | 3.57 | 1.23E-08 | 1.91E-05 | human |
| Cistrome_ChIP | GM12878 | PAX5 | 21 | 76 | 0.28 | 7.25 | 2.47 | 5.57 | 2.90 | 1.28E-08 | 1.98E-05 | human |
| ReMap_ChIP | blood_monocyte | SPI1 | 23 | 76 | 0.30 | 8.63 | 2.58 | 5.56 | 2.66 | 1.35E-08 | 2.08E-05 | human |
| TxnFactorChIPV3 | GM18951+TNFa | RELA | 13 | 76 | 0.17 | 3.32 | 1.74 | 5.55 | 3.91 | 1.41E-08 | 2.17E-05 | human |
| Cistrome_ChIP | Monocyte_CD14+ | STAT1 | 16 | 76 | 0.21 | 4.71 | 2.04 | 5.52 | 3.40 | 1.70E-08 | 2.62E-05 | human |
| Misc_ChIP | CD34+ | RUNX1 | 19 | 76 | 0.25 | 6.27 | 2.31 | 5.51 | 3.03 | 1.77E-08 | 2.73E-05 | human |
| TxnFactorChIPV3 | GM12878 | IRF4 | 13 | 76 | 0.17 | 3.26 | 1.77 | 5.50 | 3.98 | 1.88E-08 | 2.90E-05 | human |
| Cistrome_ChIP | GM19193 | RELA | 8 | 76 | 0.11 | 1.49 | 1.20 | 5.41 | 5.37 | 3.13E-08 | 4.83E-05 | human |
| TxnFactorChIPV3 | GM12878 | STAT5A | 10 | 76 | 0.13 | 2.17 | 1.45 | 5.38 | 4.60 | 3.72E-08 | 5.75E-05 | human |
| Misc_ChIP | BCells+Sendai_Virus | MED1 | 9 | 76 | 0.12 | 1.86 | 1.33 | 5.36 | 4.83 | 4.13E-08 | 6.38E-05 | human |

**Table S19. PPI network confidence scores and module enrichments: (a)** pathway enrichment analysis of individual modules, **(b)** confidence scores and network types for each edge in the PPI network.

**(a) PPI network module pathway enrichment analysis (only two most significant pathways depicted)**

| Module | PPI module enrichment p-value | Number of nodes | Number of edges | Functional Enrichment | | |
|---|---|---|---|---|---|---|
| | | | | Network | Count in gene set | Adjusted P-value |
| 1 | 2.82E-05 | 11 | 19 | GO:0006950: response to stress | 11 of 3267 | 4.34E-06 |
| | | | | GO:0006952: defense response | 8 of 1234 | 2.14E-05 |
| 2 | 1.48E-03 | 3 | 2 | GO:0050727: regulation of inflammatory response | 3 of 338 | 1.20E-03 |
| | | | | REACTOME: HSA-168249: Innate Immune System | 3 of 1012 | 6.50E-04 |
| 3 | 1.07E-01 | 6 | 3 | GO:0000398: mRNA splicing, via spliceosome | 3 of 284 | 1.90E-02 |
| | | | | REACTOME: HSA-8953854 Metabolism of RNA | 4 of 652 | 1.60E-03 |
| 4 | < 1.0E-16 | 10 | 45 | GO:0070098: chemokine-mediated signaling pathway | 10 of 75 | 5.60E-22 |
| | | | | REACTOME: HSA-380108: Chemokine receptors bind chemokines | 10 of 48 | 8.04E-25 |
| 5 | 4.44E-16 | 9 | 17 | GO:0006955: immune response | 9 of 1560 | 9.63E-08 |
| | | | | REACTOME: HSA-168256: Immune System | 9 of 1925 | 6.17E-08 |
| 6 | < 1.0E-16 | 6 | 14 | GO:0019221: cytokine-mediated signaling pathway | 6 of 655 | 2.12E-07 |
| | | | | REACTOME: HSA-6788467: IL-6-type cytokine receptor ligand interactions | 6 of 17 | 2.07E-17 |
| 7 | 1.83E-06 | 5 | 5 | GO:0043122: regulation of I-kappaB kinase/NF-kappaB signaling | 4 of 167 | 1.89E-05 |
| | | | | GO:0097190: apoptotic signaling pathway | 4 of 295 | 8.76E-05 |
| 8 | 8.54E-04 | 2 | 1 | GO:0002227: innate immune response in mucosa | 2 of 22 | 1.60E-04 |
| | | | | GO:0019731: antibacterial humoral response | 2 of 47 | 1.60E-04 |

**(b) Confidence scores and network types for each edge in the PPI network**

| Protein 1 | Protein 2 | Confidence Score | Network group | Network |
|---|---|---|---|---|
| CASP8 | PYCARD | 1.00 | Physical Interactions/Predicted | IREF-INTACT/Wu-Stein-2010 |
| CFH | CFHR1 | 1.00 | Co-expression | Mallon-McKay-2013 |
| CFH | ITGAM | 0.11 | Physical Interactions | IREF-HPRD |
| CXCL8 | RELA | 1.00 | Pathway | Wu-Stein-2010 |
| CXCL8 | PF4 | 1.00 | Co-expression | Ramaswamy-Golub-2001 |
| CXCR1 | CXCL6 | 1.00 | Pathway | Wu-Stein-2010 |
| CXCR1 | CXCL3 | 1.00 | Physical Interactions | IREF-HPRD |
| CXCR1 | CXCL2 | 1.00 | Physical Interactions | IREF-HPRD |
| CXCR1 | PPBP | 1.00 | Physical Interactions | IREF-DIP |
| CXCR1 | CXCL5 | 1.00 | Physical Interactions | IREF-DIP |
| CXCR2 | CXCL8 | 1.00 | Co-expression/Pathway/Physical Interactions | Noble-Diehl-2008/NCI_NATURE/IREF-HPRD |
| CXCR2 | CXCL1 | 1.00 | Co-expression/Pathway/Physical Interactions | Noble-Diehl-2008/Wu-Stein-2010/IREF-HPRD |
| CXCR2 | CXCL3 | 1.00 | Pathway/Physical Interactions/Predicted | Wu-Stein-2010/IREF-HPRD/I2D-INNATEDB-Mouse2Human |
| CXCR2 | CXCR1 | 1.00 | Physical Interactions/Predicted/Shared protein domains | IREF-HPRD/Wu-Stein-2010/INTERPRO |
| CXCR2 | CXCL2 | 1.00 | Co-expression/Pathway/Physical Interactions/Predicted | Noble-Diehl-2008/Wu-Stein-2010/IREF-HPRD/I2D-INNATEDB-Mouse2Human |
| CXCR2 | CXCL5 | 1.00 | Physical Interactions/Co-expression | IREF-DIP/Bild-Nevins-2006 |
| CXCR2 | CXCL6 | 1.00 | Co-expression/Pathway/Physical Interactions | Noble-Diehl-2008/Wu-Stein-2010/IREF-HPRD |
| CXCR2 | PPBP | 1.00 | Pathway/Physical Interactions | Wu-Stein-2010/IREF-HPRD |
| DEFA3 | DEFA1 | 1.00 | Physical Interactions/Shared protein domains | IREF-HPRD/INTERPRO |
| FASLG | CASP10 | 1.00 | Pathway | NCI_NATURE |
| FASLG | CASP8 | 1.00 | Co-localization | Schadt-Shoemaker-2004 |
| FUS | CFL1 | 0.13 | Physical Interactions/Co-expression | Wang-Xu-2015/Ramaswamy-Golub-2001 |
| FUS | EIF4A1 | 0.22 | Co-expression | Innocenti-Brown-2011 |
| FUS | SF3A1 | 1.00 | Co-expression | Mallon-McKay-2013 |
| FUS | POLR2A | 1.00 | Physical Interactions | IREF-HPRD |
| FUS | RELA | 0.35 | Physical Interactions | IREF-HPRD |
| FUS | KAT5 | 0.18 | Physical Interactions | IREF-DIP |
| IL31A | LIFR | 1.00 | Physical Interactions | IREF-HPRD |
| IL6ST | IL31A | 1.00 | Physical Interactions | IREF-HPRD |
| IL6ST | LIFR | 1.00 | Shared protein domains/Physical Interactions | INTERPRO/IREF-HPRD |
| IL6ST | LIF | 1.00 | Physical Interactions | IREF-INTACT |
| IL6ST | OSM | 1.00 | Physical Interactions | IREF-INTACT |
| IL6ST | OSMR | 1.00 | Shared protein domains/Physical Interactions | INTERPRO/IREF-HPRD |
| IRF8 | LY86 | 1.00 | Co-expression | Wang-Maris-2006 |
| IRF8 | IRF4 | 0.39 | Co-expression/Shared protein domains | Boldrick-Relman-2002/INTERPRO |
| LIF | LIFR | 1.00 | Physical Interactions/Predicted | IREF-HPRD/I2D-BIND-Rat2Human |
| LYN | FCGR2B | 0.39 | Co-expression/Physical Interactions/Predicted | Perou-Botstein-2000/IREF-HPRD/Wu-Stein-2010 |
| LYN | FCGR2A | 0.37 | Physical Interactions/Predicted | IREF-HPRD/Wu-Stein-2010 |
| LYN | CTLA4 | 1.00 | Physical Interactions | IREF-HPRD |
| LYN | FCAR | 0.38 | Physical Interactions | IREF-HPRD |
| LYN | TNFSF12 | 0.15 | Physical Interactions | BIOGRID-SMALL-SCALE-STUDIES |
| LYN | FASLG | 0.25 | Physical Interactions | IREF-INTACT |
| LYN | IL6ST | 1.00 | Pathway | Hallek-1997 |
| LYN | IL7R | 0.11 | Co-expression/Physical Interactions | Wu-Garvey-2007/IREF-HPRD |
| NF2 | MAP3K11 | 0.37 | Physical Interactions | IREF-INTACT |
| NOTCH1 | REL | 0.36 | Physical Interactions | IREF-HPRD |
| OSM | OSMR | 1.00 | Pathway/Physical Interactions | Wu-Stein-2010/IREF-HPRD |
| POLR2A | EHBP1L1 | 0.11 | Physical Interactions | Kristensen-Foster-2012 |
| PYCARD | PYDC1 | 0.41 | Co-expression/Physical Interactions/Shared protein domains | Mallon-McKay-2013/IREF-INTACT/INTERPRO |
| PYCARD | TP53 | 0.12 | Pathway | NCI_NATURE |
| REL | IL12B | 0.25 | Pathway/Predicted | Wu-Stein-2010/I2D-INNATEDB-Mouse2Human |
| RELA | ASCC2 | 0.17 | Physical Interactions | IREF-SMALL-SCALE-STUDIES |
| RELA | IL12B | 0.33 | Pathway/Predicted | Wu-Stein-2010/I2D-INNATEDB-Mouse2Human |
| RELA | IL7R | 0.12 | Co-expression | Rosenwald-Staudt-2001 |
| RELA | IRF8 | 0.25 | Physical Interactions | IREF-SMALL-SCALE-STUDIES |
| RELA | IRF4 | 0.15 | Co-expression/Pathway | Boldrick-Relman-2002/Wu-Stein-2010 |
| RELA | KAT5 | 0.35 | Co-expression | Smirnov-Cheung-2009 |
| RELA | NOTCH1 | 0.35 | Physical Interactions/Predicted | IREF-HPRD/Wu-Stein-2010 |
| RELA | CXCL1 | 0.21 | Pathway/Co-expression | Wu-Stein-2010/Boldrick-Relman-2002 |
| RELA | REL | 1.00 | Pathway/Physical Interactions/Shared protein domains | Wu-Stein-2010/IREF-HPRD/INTERPRO |
| RELA | TP53 | 0.18 | Co-expression/Pathway/Shared protein domains | Roth-Zlotnik-2006/Wu-Stein-2010/INTERPRO |
| TNFSF12 | TNFRSF13B | 0.26 | Pathway | Reactome |
| TNFSF13 | TNFRSF13B | 1.00 | Physical Interactions/Pathway | IREF-DIP/Wu-Stein-2010 |
| TP53 | KAT5 | 1.00 | Physical Interactions/Predicted | IREF-HPRD/Wu-Stein-2010 |

**Table S20. Ligand-receptor pairs encoded by GWAS loci with P<1.0E-05 for at least one member of the pair.** The overall enrichment p-value for ligand-receptor pairs in DLRP database: P=0.01. *Blood cis-eQTL effect of the risk allele (eQTL Gen).

| LIGAND | | | | | | | RECEPTOR | | | | | | |
|---|---|---|---|---|---|---|---|---|---|---|---|---|---|
| **GENE** | **CHR:BP** | **SNP** | **RISK ALLELE** | **OR** | **GWAS P-value** | **eQTL effect of risk allele*** | **GENE** | **CHR:BP** | **SNP** | **RISK ALLELE** | **OR** | **GWAS P-value** | **eQTL effect of risk allele*** |
| ***CXCL1*** | 4:74725320 | rs6828610 | G | 1.14 | 3.54E-08 | ↓ | ***CXCR1*** | 2:218991005 | rs4674259 | G | 1.08 | 3.73E-06 | ↑ |
| ***CXCL2*** | 4:74725320 | rs6828610 | G | 1.14 | 3.54E-08 | ↓ | ***CXCR2*** | 2:218991005 | rs4674259 | G | 1.08 | 3.73E-06 | ↑ |
| ***CXCL3*** | 4:74725320 | rs6828610 | G | 1.14 | 3.54E-08 | - | | | | | | | |
| ***CXCL5*** | 4:74725320 | rs6828610 | G | 1.14 | 3.54E-08 | ↑ | | | | | | | |
| ***CXCL6*** | 4:74725320 | rs6828610 | G | 1.14 | 3.54E-08 | - | | | | | | | |
| ***CXCL7 (PPBP)*** | 4:74725320 | rs6828610 | G | 1.14 | 3.54E-08 | - | | | | | | | |
| ***CXCL8 (IL8)*** | 4:74725320 | rs6828610 | G | 1.14 | 3.54E-08 | ↑ | | | | | | | |
| ***OSM*** | 22:30512478 | rs4823074 | G | 1.16 | 7.76E-15 | ↓ | ***OSMR*** | 5:38867732 | rs395157 | T | 1.11 | 1.55E-06 | - |
| | | | | | | | ***LIFR*** | 5:38867732 | rs395157 | T | 1.11 | 1.55E-06 | - |
| | | | | | | | ***IL6ST*** | 5:55438851 | rs10065637 | C | 1.15 | 1.77E-07 | ↑ |
| ***LIF*** | 22:30577771 | rs714027 | G | 1.14 | 3.99E-14 | ↑ | ***LIFR*** | 5:38867732 | rs395157 | T | 1.11 | 1.55E-06 | - |
| | | | | | | | ***IL6ST*** | 5:55438851 | rs10065637 | C | 1.15 | 1.77E-07 | ↑ |
| ***IL6*** | 7:22752731 | rs6969599 | A | 1.09 | 5.61E-04 | - | ***IL6ST*** | 5:55438851 | rs10065637 | C | 1.15 | 1.77E-07 | ↑ |
| ***TNFSF13 (APRIL)*** | 17:7462969 | rs3803800 | A | 1.15 | 1.21E-10 | ↑ | ***TNFRSF13B (TACI)*** | 17:16851450 | rs57382045 | A | 1.16 | 3.45E-09 | ↑ |
| ***IL7*** | 8:79561083 | rs11776243 | C | 1.09 | 2.03E-04 | ↓ | ***IL7R*** | 5:35857850 | rs10213865 | A | 1.11 | 6.57E-06 | ↑ |
| ***IL31*** | 12:122652365 | rs796259997 | GT | 1.18 | 6.26E-04 | - | ***IL31RA*** | 5:55438851 | rs10065637 | C | 1.15 | 1.77E-07 | - |
| ***TNFSF8*** | 9:117643362 | rs13300483 | T | 1.13 | 1.27E-08 | ↓ | ***TNFRSF8*** | 1:12096379 | rs1208994 | A | 1.10 | 3.73E-04 | - |
| ***TNFSF4*** | 1:173146357 | rs4916312 | A | 1.14 | 5.00E-08 | ↓ | ***TNFRSF4*** | 1:1193586 | rs59549363 | A | 1.23 | 7.60E-04 | ↓ |
| ***TNFSF3 (LTB)*** | 6:31603591 | rs2261033 | G | 1.17 | 3.38E-22 | - | ***TNFRSF3 (LTBR)*** | 12:6492649 | rs11064157 | C | 1.11 | 1.51E-04 | ↓ |

**Table S21. Expression QTLs (eQTL) of top IgAN risk loci and their LD proxies in primary immune cells from the DICE project:** the top SNP is reported for each eGene in each cell type, R2 is the measure of linkage disequilibrium between top SNP from GWAS and top SNP for a given eGene, effect sizes are provided for each IgAN risk allele, co-localization probabilities between IgAN GWAS and eQTL summary statistics are also provided: PP3 is the posterior probability that there are two different causal variants. PP4 is the posterior probability that the same causal variant is shared.

| | | | | | | | | | | | Co-localization | |
|---|---|---|---|---|---|---|---|---|---|---|---|---|
| **LOCUS** | **CHR** | **BP** | **SNP-risk allele** | **GENE** | **Tissue** | **Effect** | **P-value** | **top SNP** | **R2** | **Effect of risk allele** | **PP3** | **PP4** |
| REL | 2 | 61091950 | rs842636-A | NONOP2 | Naïve B cells | 0.6695 | 2.29E-06 | rs842638 | 0.973 | ↓ | 0.04 | **0.83** |
| CD28 | 2 | 204625370 | rs59789950-C | RAPH1 | Naïve B cells | 0.9634 | 5.20E-05 | rs3769684 | 0.809 | ↓ | 0.04 | 0.30 |
| CCR6 | 6 | 167368885 | rs112074021-A | FGFR1OP | Monocytes | 1.5011 | 4.56E-07 | rs2282859 | 0.616 | ↑ | 0.03 | 0.47 |
| IRF4/DUSP22 | 6 | 251685 | rs12195338-T | IRF4 | Non-classical Monocytes | 0.8396 | 1.02E-05 | rs12201499 | 0.888 | ↓ | 0.01 | **0.65** |
| IRF4/DUSP22 | 6 | 253428 | rs6596865-T | DUSP22 | Non-classical Monocytes | 0.7970 | 3.51E-05 | | 0.888 | ↓ | 0.01 | **0.63** |
| TNFSF8/15 | 9 | 117597885 | rs34187507-G | TNC | TH1/17 cells | 0.4673 | 5.88E-06 | rs13300483 | 0.886 | ↓ | 0.94 | 0.01 |
| TNFSF8/15 | 9 | 117624799 | rs35097049-C | TNFSF8 | NK cells | 0.4491 | 1.82E-05 | | 0.901 | ↓ | 0.71 | 0.04 |
| ZMIZ1/PPIF | 10 | 81048158 | rs942793-G | ZMIZ1 | NK cells | 0.6207 | 1.75E-05 | rs1108618 | 0.707 | ↓ | 0.02 | **0.58** |
| REEP3 | 10 | 65013950 | rs76460998-C | NRBF2 | Non-classical Monocytes | 1.2179 | 2.23E-05 | rs10896045 | 0.916 | ↓ | 0.18 | 0.37 |
| IGH | 14 | 107166471 | rs11624734-T | IGHV3-65 | Naïve B cells | 0.8644 | 1.84E-11 | rs751081288 | 0.817 | ↑ | 0.32 | **0.68** |
| IGH | 14 | 107141916 | rs10139058-C | IGHV1-69 | Naïve B cells | 0.4402 | 1.75E-09 | | 0.872 | ↓ | 0.39 | **0.59** |
| IGH | 14 | 107166471 | rs11624734-G | IGHV4-61 | Naïve B cells | 0.4134 | 1.44E-08 | | 0.817 | ↓ | 0.06 | **0.94** |
| IGH | 14 | 107157003 | rs6576205-C | IGHV3-33-2 | TH1/17 cells | 0.3894 | 1.91E-06 | | 0.918 | ↓ | 0.22 | **0.67** |
| IGH | 14 | 107166471 | rs11624734-G | IGHV3-47 | Naïve CD4+ T cells | 0.3853 | 5.23E-06 | | 0.817 | ↓ | 0.27 | 0.43 |
| IGH | 14 | 107169013 | rs7140392-T | IGHV3-66 | Naïve B cells | 0.6112 | 5.66E-06 | | 0.905 | ↑ | 0.41 | **0.57** |
| IGH | 14 | 107169013 | rs7140392-C | IGHVII-78-1 | Naïve CD8+ T cells | 0.3771 | 1.44E-05 | | 0.905 | ↓ | 0.20 | **0.62** |
| IGH | 14 | 107190574 | rs10142951-G | IGHV3-38 | Naïve B cells | 0.5932 | 1.84E-05 | | 0.945 | ↑ | 0.30 | 0.33 |
| IGH | 14 | 107153577 | rs8018138-T | IGHV3-64 | Naïve B cells | 0.5922 | 4.02E-05 | | 0.881 | ↑ | 0.13 | **0.76** |
| ITGAM/ITGAX | 16 | 31353796 | rs12599388-T | ITGAX | Monocytes | 0.6361 | 7.47E-06 | rs11150612 | 0.982 | ↑ | 0.35 | **0.60** |
| ITGAM/ITGAX | 16 | 31353796 | rs12599388-T | ITGAX | Non-classical Monocytes | 0.6148 | 2.44E-05 | | 0.982 | ↑ | 0.04 | **0.79** |
| TNFSF13 | 17 | 7448003 | rs11078694-T | ATP1B2 | Naïve CD8+ T cells | 0.7121 | 3.01E-05 | rs3803800 | 0.500 | ↑ | 0.03 | 0.13 |
| FCAR | 19 | 55396613 | rs10402324-G | FCAR | Non-classical Monocytes | 0.4094 | 1.89E-07 | rs1865097 | 1.000 | ↑ | 0.98 | 0.02 |
| FCAR | 19 | 55398101 | rs12972637-A | NLRP7 | Memory T-reg cells | 0.3963 | 2.09E-06 | | 0.996 | ↑ | 0.01 | **0.92** |
| FCAR | 19 | 55396613 | rs10402324-G | CTB-61M7.2 | Non-classical Monocytes | 0.3799 | 6.45E-06 | | 1.000 | ↑ | 0.98 | 0.02 |
| FCAR | 19 | 55396613 | rs10402324-G | NCR1 | Naïve CD4+ T cells | 0.3756 | 1.48E-05 | | 1.000 | ↑ | 0.01 | **0.79** |
| LIF | 22 | 30486826 | rs2412970-G | LIF | NK cells | 0.5749 | 3.45E-05 | rs48230748 | 1.000 | ↓ | 0.06 | **0.75** |
| LIF | 22 | 30549071 | rs5763821-A | NIPSNAP1 | TH1/17 cells | 0.3455 | 5.54E-05 | | 0.966 | ↑ | 0.10 | 0.42 |

**Table S22. Blood eQTLs for the top IgAN risk loci:** only significant blood cis-eQTL effects of non-HLA IgAN genome-wide significant SNPs are listed along with co-localization results. PP3: posterior probability that there are two different causal variants. PP4: posterior probability that the same causal variant is shared. Based on the blood eQTL meta-analysis of the eQTLGen Consortium.

| | | | | | | | | | Colocalization | |
|---|---|---|---|---|---|---|---|---|---|---|
| **LOCUS** | **CHR** | **BP** | **SNP-risk allele** | **eGENE** | **Z-score** | **p-value** | **FDR** | **Effect of risk allele** | **PP3** | **PP4** |
| *FCRL3/4* | 1 | 157542162 | rs849815-G | *FCRL5* | 46.5073 | 3.27E-296 | 0 | ↓ | 0.09 | **0.91** |
| *FCRL3/4* | 1 | 157542162 | rs849815-G | *FCRL3* | 49.1059 | 3.27E-296 | 0 | ↓ | 0.25 | **0.75** |
| *FCRL3/4* | 1 | 157542162 | rs849815-A | *FCRL1* | 7.2454 | 4.31E-13 | 0 | ↑ | 1.00 | 0.00 |
| *FCRL3/4* | 1 | 157542162 | rs849815-G | *RP11-367J7.2* | 5.9941 | 2.05E-09 | 1.96E-05 | ↑ | 0.99 | 0.00 |
| *TNFSF4* | 1 | 173146357 | rs4916312-G | *TNFSF4* | 16.3284 | 6.20E-60 | 0 | ↓ | 0.11 | **0.89** |
| *CFH* | 1 | 196686918 | rs6677604-A | *CFH* | 9.9646 | 2.18E-23 | 0 | ↓ | 1.00 | 0.00 |
| *REL* | 2 | 61092678 | rs842638-C | *PUS10* | 19.5107 | 8.92E-85 | 0 | ↓ | 1.00 | 0.00 |
| *REL* | 2 | 61092678 | rs842638-C | *RP11-373L24.1* | 9.5193 | 1.75E-21 | 0 | ↓ | 1.00 | 0.00 |
| *REL* | 2 | 61092678 | rs842638-C | *REL* | 9.0551 | 1.37E-19 | 0 | ↓ | 1.00 | 0.00 |
| *REL* | 2 | 61092678 | rs842638-C | *USP34* | 8.1205 | 4.65E-16 | 0 | ↓ | 1.00 | 0.00 |
| *REL* | 2 | 61092678 | rs842638-C | *AC010733.5* | 7.6041 | 2.87E-14 | 0 | ↓ | 0.05 | **0.96** |
| *PF4V1* | 4 | 74725320 | rs6828610-G | *PF4V1* | 49.2489 | 3.27E-296 | 0 | ↑ | 0.04 | **0.96** |
| *PF4V1* | 4 | 74725320 | rs6828610-G | *CXCL5* | 8.3989 | 4.51E-17 | 0 | ↑ | 1.00 | 0.00 |
| *PF4V1* | 4 | 74725320 | rs6828610-A | *CXCL1* | 6.7486 | 1.49E-11 | 0 | ↓ | 0.02 | **0.98** |
| *PF4V1* | 4 | 74725320 | rs6828610-G | *IL8* | 5.3337 | 9.63E-08 | 3.39E-04 | ↑ | 0.98 | 0.01 |
| *LY86* | 6 | 7214676 | rs12530084-T | *DSP* | 5.6881 | 1.29E-08 | 7.06E-05 | ↓ | 1.00 | 0.00 |
| *LY86* | 6 | 7214676 | rs12530084-C | *SSR1* | 4.7907 | 1.66E-06 | 0.005 | ↑ | 1.00 | 0.00 |
| *CCR6* | 6 | 167445139 | rs2282859-C | *FGFR1OP* | 25.0468 | 1.90E-138 | 0 | ↑ | 0.70 | 0.26 |
| *CCR6* | 6 | 167445139 | rs2282859-C | *RP1-167A14.2* | 9.5649 | 1.12E-21 | 0 | ↑ | 0.93 | 0.01 |
| *CCR6* | 6 | 167445139 | rs2282859-C | *RNASET2* | 8.601 | 7.90E-18 | 0 | ↑ | 0.57 | 0.40 |
| *CCR6* | 6 | 167445139 | rs2282859-C | *AL133458.1* | 4.7261 | 2.29E-06 | 0.007 | ↑ | 0.66 | 0.30 |
| *CCR6* | 6 | 167445139 | rs2282859-T | *CCR6* | 4.3735 | 1.22E-05 | 0.034 | ↓ | 0.67 | 0.29 |
| *DEFA1* | 8 | 6808722 | rs2075836-T | *DEFA8P* | 14.1255 | 2.64E-45 | 0 | ↑ | 0.89 | 0.11 |
| *DEFA1* | 8 | 6808722 | rs2075836-G | *DEFA3* | 7.2642 | 3.76E-13 | 0 | ↓ | 1.00 | 0.00 |
| *ANXA13* | 8 | 124765474 | rs34354351-G | *FAM91A1* | 10.1885 | 2.23E-24 | 0 | ↓ | 0.80 | 0.19 |
| *TNFSF8/15* | 9 | 117643362 | rs13300483-C | *TNFSF8* | 8.3302 | 8.07E-17 | 0 | ↓ | 0.89 | 0.10 |
| *CARD9* | 9 | 139266496 | rs4077515-T | *CARD9* | 72.772 | 3.27E-296 | 0 | ↑ | 0.11 | **0.90** |
| *CARD9* | 9 | 139266496 | rs4077515-C | *INPP5E* | 62.5426 | 3.27E-296 | 0 | ↓ | 0.09 | **0.91** |
| *CARD9* | 9 | 139266496 | rs4077515-C | *SDCCAG3* | 44.0373 | 3.27E-296 | 0 | ↓ | 0.05 | **0.95** |
| *CARD9* | 9 | 139266496 | rs4077515-C | *SEC16A* | 23.46 | 1.04E-121 | 0 | ↓ | 1.00 | 0.00 |
| *CARD9* | 9 | 139266496 | rs4077515-T | *SNAPC4* | 12.5376 | 4.66E-36 | 0 | ↑ | 1.00 | 0.00 |
| *CARD9* | 9 | 139266496 | rs4077515-C | *DNLZ* | 5.4878 | 4.07E-08 | 1.59E-04 | ↓ | 1.00 | 0.00 |
| *CARD9* | 9 | 139266496 | rs4077515-T | *PMPCA* | 5.2489 | 1.53E-07 | 5.19E-04 | ↑ | 0.36 | **0.64** |
| *REEP3* | 10 | 65363048 | rs57917667-A | *NRBF2* | 14.5229 | 8.69E-48 | 0 | ↓ | 1.00 | 0.00 |
| *REEP3* | 10 | 65363048 | rs57917667-G | *REEP3* | 4.9702 | 6.69E-07 | 0.002 | ↑ | 1.00 | 0.00 |
| *ZMIZ1/PPIF* | 10 | 81043743 | rs1108618-G | *ZMIZ1* | 13.9775 | 2.14E-44 | 0 | ↓ | 0.08 | **0.93** |
| *ZMIZ1/PPIF* | 10 | 81043743 | rs1108618-G | *PPIF* | 10.4062 | 2.32E-25 | 0 | ↓ | 0.45 | **0.55** |
| *RELA* | 11 | 65555524 | rs10896045-A | *EFEMP2* | 21.8385 | 1.00E-105 | 0 | ↑ | 0.01 | **1.00** |
| *RELA* | 11 | 65555524 | rs10896045-G | *CTSW* | 18.8187 | 5.30E-79 | 0 | ↓ | 1.00 | 0.00 |
| *RELA* | 11 | 65555524 | rs10896045-A | *SNX32* | 13.2675 | 3.58E-40 | 0 | ↑ | 1.00 | 0.00 |
| *RELA* | 11 | 65555524 | rs10896045-A | *FIBP* | 12.0861 | 1.25E-33 | 0 | ↑ | 1.00 | 0.00 |
| *RELA* | 11 | 65555524 | rs10896045-G | *MAP3K11* | 11.6803 | 1.61E-31 | 0 | ↓ | 1.00 | 0.00 |
| *RELA* | 11 | 65555524 | rs10896045-G | *FAM89B* | 10.5176 | 7.15E-26 | 0 | ↓ | 0.04 | 0.96 |
| *RELA* | 11 | 65555524 | rs10896045-G | *CFL1* | 9.8511 | 6.77E-23 | 0 | ↓ | 0.86 | 0.14 |
| *RELA* | 11 | 65555524 | rs10896045-A | *RP11-770G2.2* | 8.6002 | 7.96E-18 | 0 | ↑ | 1.00 | 0.00 |
| *RELA* | 11 | 65555524 | rs10896045-G | *MUS81* | 8.5381 | 1.36E-17 | 0 | ↓ | 0.81 | 0.19 |
| *RELA* | 11 | 65555524 | rs10896045-A | *KRT8P26* | 7.3516 | 1.96E-13 | 0 | ↑ | 1.00 | 0.00 |
| *RELA* | 11 | 65555524 | rs10896045-G | *RNASEH2C* | 7.2179 | 5.28E-13 | 0 | ↓ | 1.00 | 0.00 |
| *RELA* | 11 | 65555524 | rs10896045-G | *BANF1* | 6.4199 | 1.36E-10 | 0 | ↓ | 1.00 | 0.00 |
| *RELA* | 11 | 65555524 | rs10896045-A | *LTBP3* | 4.635 | 3.57E-06 | 0.01 | ↑ | 1.00 | 0.00 |
| *ITGAM/ITGAX* | 16 | 31357760 | rs11150612-A | *ITGAX* | 46.6839 | 3.27E-296 | 0 | ↑ | 0.09 | **0.91** |
| *ITGAM/ITGAX* | 16 | 31357760 | rs11150612-A | *RP11-120K18.3* | 20.4771 | 3.45E-93 | 0 | ↑ | 1.00 | 0.00 |
| *ITGAM/ITGAX* | 16 | 31357760 | rs11150612-A | *BCKDK* | 19.6062 | 1.37E-85 | 0 | ↑ | 1.00 | 0.00 |
| *ITGAM/ITGAX* | 16 | 31357760 | rs11150612-A | *ZNF668* | 19.0704 | 4.47E-81 | 0 | ↑ | 1.00 | 0.00 |
| *ITGAM/ITGAX* | 16 | 31357760 | rs11150612-A | *ITGAD* | 18.4896 | 2.50E-76 | 0 | ↑ | 1.00 | 0.00 |
| *ITGAM/ITGAX* | 16 | 31357760 | rs11150612-A | *HSD3B7* | 12.7532 | 3.00E-37 | 0 | ↑ | 1.00 | 0.00 |
| *ITGAM/ITGAX* | 16 | 31357760 | rs11150612-G | *STX4* | 12.2608 | 1.47E-34 | 0 | ↓ | 1.00 | 0.00 |
| *ITGAM/ITGAX* | 16 | 31357760 | rs11150612-A | *ITGAM* | 11.279 | 1.67E-29 | 0 | ↑ | 1.00 | 0.00 |
| *ITGAM/ITGAX* | 16 | 31357760 | rs11150612-G | *RP11-196G11.2* | 10.8147 | 2.93E-27 | 0 | ↓ | 1.00 | 0.00 |
| *ITGAM/ITGAX* | 16 | 31357760 | rs11150612-G | *KAT8* | 7.0899 | 1.34E-12 | 0 | ↓ | 1.00 | 0.00 |
| *ITGAM/ITGAX* | 16 | 31357760 | rs11150612-A | *VKORC1* | 7.075 | 1.50E-12 | 0 | ↑ | 0.89 | 0.11 |
| *ITGAM/ITGAX* | 16 | 31357760 | rs11150612-G | *ARMC5* | 6.2347 | 4.52E-10 | 0 | ↓ | 1.00 | 0.00 |
| *ITGAM/ITGAX* | 16 | 31357760 | rs11150612-A | *C16orf93* | 5.7421 | 9.36E-09 | 5.79E-05 | ↑ | 0.36 | **0.64** |
| *ITGAM/ITGAX* | 16 | 31357760 | rs11150612-G | *SETD1A* | 5.0383 | 4.69E-07 | 0.002 | ↓ | 0.06 | **0.94** |
| *ITGAM/ITGAX* | 16 | 31357760 | rs11150612-A | *PRSS53* | 4.9511 | 7.38E-07 | 0.002 | ↑ | 1.00 | 0.00 |
| *IRF8* | 16 | 86017715 | rs1879210-C | *RP11-542M13.2* | 8.7386 | 2.36E-18 | 0 | ↓ | 0.99 | 0.01 |
| *IRF8* | 16 | 86017715 | rs1879210-C | *RP11-542M13.3* | 4.7013 | 2.59E-06 | 0.007 | ↓ | 0.70 | 0.30 |
| *TNFSF13* | 17 | 7462969 | rs3803800-G | *TNFSF12* | 15.0379 | 4.14E-51 | 0 | ↓ | 1.00 | 0.00 |
| *TNFSF13* | 17 | 7462969 | rs3803800-A | *TNFSF13* | 14.608 | 2.50E-48 | 0 | ↑ | 0.98 | 0.03 |
| *TNFSF13* | 17 | 7462969 | rs3803800-A | *ATP1B2* | 12.9661 | 1.91E-38 | 0 | ↑ | 1.00 | 0.00 |
| *TNFSF13* | 17 | 7462969 | rs3803800-G | *SAT2* | 8.5808 | 9.42E-18 | 0 | ↓ | 1.00 | 0.00 |
| *TNFSF13* | 17 | 7462969 | rs3803800-A | *CD68* | 7.0055 | 2.46E-12 | 0 | ↑ | 1.00 | 0.00 |
| *TNFSF13* | 17 | 7462969 | rs3803800-A | *NLGN2* | 5.4252 | 5.79E-08 | 2.40E-04 | ↑ | 0.03 | **0.98** |
| *TNFSF13* | 17 | 7462969 | rs3803800-G | *TP53* | 4.3404 | 1.42E-05 | 0.04 | ↓ | 1.00 | 0.00 |
| *TNFRSF13B* | 17 | 16851450 | rs57382045-A | *NT5M* | 9.7146 | 2.61E-22 | 0 | ↑ | 1.00 | 0.00 |
| *TNFRSF13B* | 17 | 16851450 | rs57382045-G | *COPS3* | 8.1101 | 5.05E-16 | 0 | ↓ | 1.00 | 0.00 |
| *FCAR* | 19 | 55397217 | rs1865097-A | *FCAR* | 25.9667 | 1.18E-148 | 0 | ↑ | 0.99 | 0.01 |
| *FCAR* | 19 | 55397217 | rs1865097-A | *CTB-61M7.2* | 23.6104 | 3.01E-123 | 0 | ↑ | 0.98 | 0.02 |
| *FCAR* | 19 | 55397217 | rs1865097-A | *KIR2DL1* | 7.3729 | 1.67E-13 | 0 | ↑ | 1.00 | 0.00 |
| *LIF* | 22 | 30512478 | rs4823074-G | *MTMR3* | 16.8319 | 1.42E-63 | 0 | ↑ | 1.00 | 0.00 |
| *LIF* | 22 | 30512478 | rs4823074-G | *NIPSNAP1* | 15.9373 | 3.49E-57 | 0 | ↑ | 1.00 | 0.00 |
| *LIF* | 22 | 30512478 | rs4823074-A | *UQCR10* | 14.2496 | 4.51E-46 | 0 | ↓ | 1.00 | 0.00 |
| *LIF* | 22 | 30512478 | rs4823074-G | *SF3A1* | 9.9708 | 2.04E-23 | 0 | ↑ | 1.00 | 0.00 |
| *LIF* | 22 | 30512478 | rs4823074-G | *ASCC2* | 9.4183 | 4.58E-21 | 0 | ↑ | 1.00 | 0.00 |
| *LIF* | 22 | 30512478 | rs4823074-G | *SEC14L3* | 7.6887 | 1.49E-14 | 0 | ↑ | 1.00 | 0.00 |
| *LIF* | 22 | 30512478 | rs4823074-A | *DUSP18* | 6.1692 | 6.87E-10 | 0 | ↓ | 1.00 | 0.00 |
| *LIF* | 22 | 30512478 | rs4823074-A | *ZMAT5* | 4.8488 | 1.24E-06 | 0.004 | ↓ | 1.00 | 0.00 |

#### Table S23. Significant blood pQTL effects of genome-wide significant IgAN risk loci.

| LOCUS | CHR | BP | SNP-risk allele | Target | IgAN top SNP in the Locus | r2 | beta | p | IgAN risk allele effect | Cis/Trans | Study |
|---|---|---|---|---|---|---|---|---|---|---|---|
| FCRL3/4 | 1 | 157670816 | rs7528684-G | FCRL3 | rs849815 | 0.24 | 0.53 | 1.40E-112 | ↓ | cis | Sun BB et al |
| FCRL3/4 | 1 | 157779182 | rs4971155-T | FCRL1 | | 0.21 | 0.21 | 6.30E-26 | ↓ | cis | Sun BB et al |
| FCRL3/4 | 1 | 157635373 | rs6677006-T | FCRL4 | | 0.17 | 0.55 | 5.50E-38 | ↓ | cis | Sun BB et al |
| CFH | 1 | 196825287 | rs7519758-C | LRC19 | rs6677604 | 0.97 | 0.48 | 1.40E-307 | ↑ | trans | Sun BB et al |
| CFH | 1 | 196819479 | rs149369377-A | XRCC4 | | 0.97 | 0.21 | 3.50E-17 | ↑ | trans | Sun BB et al |
| CFH | 1 | 196821380 | rs67908756-T | Calnexin | | 0.97 | 0.19 | 3.90E-14 | ↑ | trans | Sun BB et al |
| CFH | 1 | 196814850 | rs148235292-A | LIRA5 | | 0.96 | 0.26 | 5.90E-18 | ↓ | trans | Sun BB et al |
| CFH | 1 | 196632004 | rs11390840-CA | FYN | | 0.92 | 0.48 | 6.50E-66 | ↑ | trans | Sun BB et al |
| CFH | 1 | 196584321 | rs4657825-A | CFB | | 0.91 | 0.22 | 6.30E-22 | ↓ | trans | Sun BB et al |
| CFH | 1 | 196886770 | rs4915559-T | HPX | | 0.68 | 0.68 | 1.78E-140 | ↑ | trans | Emilsson V et al |
| CFH | 1 | 196886770 | rs4915559-T | CFHR4 | | 0.68 | 0.66 | 1.67E-131 | ↑ | cis | Emilsson V et al |
| CFH | 1 | 196886770 | rs4915559-C | NDUFS4 | | 0.68 | 0.27 | 5.50E-42 | ↓ | trans | Emilsson V et al |
| CFH | 1 | 196886770 | rs4915559-C | TST | | 0.68 | 0.26 | 1.53E-40 | ↓ | trans | Emilsson V et al |
| CFH | 1 | 196886770 | rs4915559-T | F13B | | 0.68 | 0.34 | 3.55E-36 | ↑ | trans | Emilsson V et al |
| CFH | 1 | 196886770 | rs4915559-T | C1S | | 0.68 | 0.27 | 5.86E-22 | ↑ | trans | Emilsson V et al |
| CFH | 1 | 196886770 | rs4915559-C | CFP | | 0.68 | 0.21 | 7.81E-20 | ↓ | trans | Emilsson V et al |
| CFH | 1 | 196886770 | rs4915559-C | CEACAM7 | | 0.68 | 0.20 | 1.46E-19 | ↓ | trans | Emilsson V et al |
| CFH | 1 | 196886770 | rs4915559-C | ANAPC7 | | 0.68 | 0.17 | 3.27E-13 | ↓ | trans | Emilsson V et al |
| CFH | 1 | 196886770 | rs4915559-T | CFHR5 | | 0.68 | 0.20 | 4.44E-13 | ↑ | cis | Emilsson V et al |
| CFH | 1 | 196886770 | rs4915559-T | PPP2R3A | | 0.68 | 0.19 | 1.15E-11 | ↑ | trans | Emilsson V et al |
| CFH | 1 | 196886770 | rs4915559-T | PDGFRA | | 0.68 | 0.17 | 3.07E-09 | ↑ | trans | Emilsson V et al |
| CFH | 1 | 196841377 | rs57809726-A | CFHR1 | | 0.59 | 1.30 | 1.00E-300 | ↑ | cis | Emilsson V et al |
| CFH | 1 | 196841377 | rs57809726-A | PRLR | | 0.59 | 0.40 | 2.26E-28 | ↑ | trans | Emilsson V et al |
| CFH | 1 | 196841377 | rs57809726-A | GALK1 | | 0.59 | 0.31 | 2.87E-19 | ↑ | trans | Emilsson V et al |
| CFH | 1 | 196841377 | rs57809726-G | CFH | | 0.59 | 0.22 | 4.71E-16 | ↓ | cis | Emilsson V et al |
| CFH | 1 | 196815708 | rs71524421-C | GSKIP | | 0.21 | 0.14 | 1.56E-10 | ↓ | trans | Emilsson V et al |
| CFH | 1 | 196815708 | rs71524421-G | EFCAB14 | | 0.21 | 0.16 | 6.80E-10 | ↑ | trans | Emilsson V et al |
| CFH | 1 | 196673993 | rs1831282-A | BATF3 | | 0.21 | 0.19 | 1.80E-21 | ↑ | trans | Sun BB et al |
| CFH | 1 | 196719716 | rs529541-G | ARHGA | | 0.21 | 0.31 | 7.20E-23 | ↑ | trans | Sun BB et al |
| CFH | 1 | 196667252 | rs7539005-A | GL8D1 | | 0.19 | 0.40 | 8.30E-206 | ↑ | trans | Sun BB et al |
| CFH | 1 | 196659237 | rs1061170-T | PGRC2 | | 0.19 | 0.38 | 1.70E-52 | ↓ | trans | Sun BB et al |
| CFH | 1 | 196667252 | rs7539005-A | TSNA1 | | 0.19 | 0.26 | 1.90E-47 | ↑ | trans | Sun BB et al |
| CFH | 1 | 196668360 | rs34813609-GT | APOD | | 0.19 | 0.19 | 4.20E-21 | ↑ | trans | Sun BB et al |
| CFH | 1 | 196668360 | rs34813609-GT | ZNF276 | | 0.19 | 0.19 | 7.90E-21 | ↑ | trans | Sun BB et al |
| CFH | 1 | 196668360 | rs34813609-GT | GPD1L | | 0.19 | 0.19 | 9.30E-20 | ↑ | trans | Sun BB et al |
| CFH | 1 | 196668360 | rs34813609-GT | PCDB4 | | 0.19 | 0.18 | 5.20E-18 | ↑ | trans | Sun BB et al |
| CFH | 1 | 196668360 | rs34813609-GT | SLAF7 | | 0.19 | 0.17 | 2.20E-17 | ↑ | trans | Sun BB et al |
| CFH | 1 | 196671217 | rs28664709-G | SPTA2 | | 0.19 | 0.17 | 4.40E-17 | ↑ | trans | Sun BB et al |
| CFH | 1 | 196668360 | rs34813609-GT | KLK14 | | 0.19 | 0.17 | 2.30E-16 | ↑ | trans | Sun BB et al |
| CFH | 1 | 196672454 | rs12038333-G | PRP16 | | 0.19 | 0.17 | 6.00E-16 | ↑ | trans | Sun BB et al |
| CFH | 1 | 196668360 | rs34813609-GT | sFRP-3 | | 0.19 | 0.17 | 1.20E-15 | ↑ | trans | Sun BB et al |
| CFH | 1 | 196668360 | rs34813609-GT | IGFL3 | | 0.19 | 0.17 | 2.80E-15 | ↑ | trans | Sun BB et al |
| CFH | 1 | 196668360 | rs34813609-GT | AUGN | | 0.19 | 0.17 | 5.80E-15 | ↑ | trans | Sun BB et al |
| CFH | 1 | 196668360 | rs34813609-GT | MCEM1 | | 0.19 | 0.16 | 2.30E-14 | ↑ | trans | Sun BB et al |
| CFH | 1 | 196668360 | rs34813609-GT | C1T9A | | 0.19 | 0.16 | 5.50E-14 | ↑ | trans | Sun BB et al |
| CFH | 1 | 196672454 | rs12038333-G | HXK4 | | 0.19 | 0.16 | 5.80E-14 | ↑ | trans | Sun BB et al |
| CFH | 1 | 196668360 | rs34813609-GT | PTP-1B | | 0.19 | 0.16 | 7.20E-14 | ↑ | trans | Sun BB et al |
| CFH | 1 | 196668360 | rs34813609-GT | RGS8 | | 0.19 | 0.16 | 8.70E-14 | ↑ | trans | Sun BB et al |
| CFH | 1 | 196668360 | rs34813609-GT | VTN | | 0.19 | 0.16 | 1.00E-13 | ↑ | trans | Sun BB et al |
| CFH | 1 | 196668360 | rs34813609-GT | P3C2A | | 0.19 | 0.16 | 2.40E-13 | ↑ | trans | Sun BB et al |
| CFH | 1 | 196668360 | rs34813609-GT | BIRC5 | | 0.19 | 0.15 | 3.70E-13 | ↑ | trans | Sun BB et al |
| CFH | 1 | 196667252 | rs7539005-A | KLK6 | | 0.19 | 0.15 | 4.60E-13 | ↑ | trans | Sun BB et al |
| CFH | 1 | 196668360 | rs34813609-GT | SARP-1 | | 0.19 | 0.15 | 9.50E-13 | ↑ | trans | Sun BB et al |
| CFH | 1 | 196672454 | rs12038333-G | JAM-B | | 0.19 | 0.15 | 5.80E-12 | ↑ | trans | Sun BB et al |
| CFH | 1 | 196668360 | rs34813609-GT | OSM | | 0.19 | 0.15 | 7.40E-12 | ↑ | trans | Sun BB et al |
| CFH | 1 | 196668360 | rs34813609-GT | RMTL1 | | 0.19 | 0.15 | 8.10E-12 | ↑ | trans | Sun BB et al |
| CFH | 1 | 196670839 | rs10754199-A | CD63 | | 0.19 | 0.15 | 1.10E-11 | ↑ | trans | Sun BB et al |
| CFH | 1 | 196657064 | rs570618-T | ERVV1 | | 0.18 | 0.17 | 5.90E-15 | ↑ | trans | Sun BB et al |
| CFH | 1 | 196657064 | rs570618-T | OPG | | 0.18 | 0.15 | 5.20E-13 | ↑ | trans | Sun BB et al |
| CFH | 1 | 196657064 | rs570618-T | LTBR | | 0.18 | 0.15 | 1.10E-12 | ↑ | trans | Sun BB et al |
| CFH | 1 | 196671981 | rs368465-T | CD22 | | 0.18 | 0.15 | 1.40E-12 | ↑ | trans | Sun BB et al |
| CFH | 1 | 196657064 | rs570618-T | CLK2 | | 0.18 | 0.15 | 1.50E-12 | ↑ | trans | Sun BB et al |
| CFH | 1 | 196657064 | rs570618-T | FRK | | 0.18 | 0.15 | 1.10E-11 | ↑ | trans | Sun BB et al |
| CFH | 1 | 196660995 | rs528298-T | MLL2 | | 0.18 | 0.76 | 3.70E-268 | ↓ | trans | Sun BB et al |
| CFH | 1 | 196660995 | rs528298-T | MA1B1 | | 0.18 | 0.37 | 6.30E-50 | ↓ | trans | Sun BB et al |
| CFH | 1 | 196660995 | rs528298-A | DCNL5 | | 0.18 | 0.15 | 8.90E-13 | ↑ | trans | Sun BB et al |
| CFH | 1 | 196660995 | rs528298-T | APC7 | | 0.18 | 0.88 | 3.5E-401 | ↓ | trans | Sun BB et al |
| CFH | 1 | 196660995 | rs528298-T | APC8 | | 0.18 | 0.32 | 3.5E-402 | ↓ | trans | Sun BB et al |
| CFH | 1 | 196867233 | rs6685931-C | DLL3 | | 0.17 | 0.14 | 1.75E-10 | ↑ | trans | Emilsson V et al |
| CFH | 1 | 196867233 | rs6685931-T | TNFRSF18 | | 0.17 | 0.15 | 2.43E-09 | ↓ | trans | Emilsson V et al |
| CFH | 1 | 196867233 | rs6685931-C | IL10RA | | 0.17 | 0.12 | 5.49E-09 | ↑ | trans | Emilsson V et al |
| PF4V1 | 4 | 74718941 | rs941758-C | SNAB | rs6828610 | 0.40 | 0.15 | 5.00E-12 | ↑ | trans | Sun BB et al |
| PF4V1 | 4 | 74718101 | rs872914-G | SLC3A2 | | 0.39 | 0.45 | 2.23E-250 | ↑ | trans | Emilsson V et al |
| PF4V1 | 4 | 74718101 | rs872914-G | PF4V1 | | 0.39 | 0.43 | 1.93E-214 | ↑ | cis | Emilsson V et al |
| PF4V1 | 4 | 74718101 | rs872914-G | EMC1 | | 0.39 | 0.43 | 8.32E-192 | ↑ | trans | Emilsson V et al |
| PF4V1 | 4 | 74718101 | rs872914-G | TNFAIP8 | | 0.39 | 0.40 | 2.55E-160 | ↑ | trans | Emilsson V et al |
| PF4V1 | 4 | 74718101 | rs872914-G | ID2 | | 0.39 | 0.38 | 3.39E-124 | ↑ | trans | Emilsson V et al |
| PF4V1 | 4 | 74718101 | rs872914-G | RAB39B | | 0.39 | 0.37 | 1.36E-112 | ↑ | trans | Emilsson V et al |
| PF4V1 | 4 | 74718101 | rs872914-G | EIF4EBP2 | | 0.39 | 0.32 | 3.02E-73 | ↑ | trans | Emilsson V et al |
| PF4V1 | 4 | 74718101 | rs872914-G | EVA1B | | 0.39 | 0.29 | 2.05E-57 | ↑ | trans | Emilsson V et al |
| PF4V1 | 4 | 74718101 | rs872914-G | ETNK1 | | 0.39 | 0.28 | 8.15E-52 | ↑ | trans | Emilsson V et al |
| PF4V1 | 4 | 74718101 | rs872914-G | KITLG | | 0.39 | 0.28 | 2.12E-49 | ↑ | trans | Emilsson V et al |
| PF4V1 | 4 | 74718101 | rs872914-G | CREBBP | | 0.39 | 0.27 | 1.01E-43 | ↑ | trans | Emilsson V et al |
| PF4V1 | 4 | 74718101 | rs872914-G | SCG2 | | 0.39 | 0.24 | 3.33E-34 | ↑ | trans | Emilsson V et al |
| PF4V1 | 4 | 74718101 | rs872914-G | STX1B | | 0.39 | 0.24 | 1.34E-31 | ↑ | trans | Emilsson V et al |
| PF4V1 | 4 | 74718101 | rs872914-G | KPNA4 | | 0.39 | 0.23 | 2.59E-31 | ↑ | trans | Emilsson V et al |
| PF4V1 | 4 | 74718101 | rs872914-G | AXIN2 | | 0.39 | 0.24 | 3.08E-30 | ↑ | trans | Emilsson V et al |
| PF4V1 | 4 | 74718101 | rs872914-G | NLGN4X | | 0.39 | 0.23 | 2.33E-29 | ↑ | trans | Emilsson V et al |
| PF4V1 | 4 | 74718101 | rs872914-G | ARL1 | | 0.39 | 0.22 | 1.15E-27 | ↑ | trans | Emilsson V et al |
| PF4V1 | 4 | 74718101 | rs872914-G | GSTT2B | | 0.39 | 0.21 | 2.84E-23 | ↑ | trans | Emilsson V et al |
| PF4V1 | 4 | 74718101 | rs872914-G | SCGB2A1 | | 0.39 | 0.21 | 1.67E-22 | ↑ | trans | Emilsson V et al |
| PF4V1 | 4 | 74718101 | rs872914-G | ARMC5 | | 0.39 | 0.20 | 5.84E-22 | ↑ | trans | Emilsson V et al |
| PF4V1 | 4 | 74718101 | rs872914-G | CXCL6 | | 0.39 | 0.20 | 9.50E-22 | ↑ | cis | Emilsson V et al |
| PF4V1 | 4 | 74718101 | rs872914-G | WFDC13 | | 0.39 | 0.20 | 1.03E-21 | ↑ | trans | Emilsson V et al |
| PF4V1 | 4 | 74718101 | rs872914-G | RIC3 | | 0.39 | 0.19 | 5.13E-18 | ↑ | trans | Emilsson V et al |
| PF4V1 | 4 | 74718101 | rs872914-G | RAP1GDS1 | | 0.39 | 0.19 | 1.04E-17 | ↑ | trans | Emilsson V et al |
| PF4V1 | 4 | 74718101 | rs872914-G | PGK1 | | 0.39 | 0.18 | 5.64E-17 | ↑ | trans | Emilsson V et al |
| PF4V1 | 4 | 74718101 | rs872914-G | CXCL5 | | 0.39 | 0.18 | 4.04E-16 | ↑ | cis | Emilsson V et al |
| PF4V1 | 4 | 74718101 | rs872914-G | VPS29 | | 0.39 | 0.17 | 5.39E-15 | ↑ | trans | Emilsson V et al |
| PF4V1 | 4 | 74718101 | rs872914-G | RELB | | 0.39 | 0.17 | 4.09E-14 | ↑ | trans | Emilsson V et al |
| PF4V1 | 4 | 74718101 | rs872914-G | PSG5 | | 0.39 | 0.17 | 4.18E-14 | ↑ | trans | Emilsson V et al |
| PF4V1 | 4 | 74718101 | rs872914-G | HERC1 | | 0.39 | 0.17 | 4.68E-14 | ↑ | trans | Emilsson V et al |
| PF4V1 | 4 | 74718101 | rs872914-G | DNAJC11 | | 0.39 | 0.16 | 8.37E-13 | ↑ | trans | Emilsson V et al |
| PF4V1 | 4 | 74718101 | rs872914-G | WISP3 | | 0.39 | 0.16 | 1.44E-12 | ↑ | trans | Emilsson V et al |
| PF4V1 | 4 | 74718101 | rs872914-G | IFNLR1 | | 0.39 | 0.15 | 3.48E-11 | ↑ | trans | Emilsson V et al |
| PF4V1 | 4 | 74718101 | rs872914-G | TMCC3 | | 0.39 | 0.14 | 3.39E-10 | ↑ | trans | Emilsson V et al |
| PF4V1 | 4 | 74718101 | rs872914-G | DDR1 | | 0.39 | 0.14 | 1.13E-09 | ↑ | trans | Emilsson V et al |
| PF4V1 | 4 | 74718101 | rs872914-G | FGFR3 | | 0.39 | 0.13 | 1.20E-09 | ↑ | trans | Emilsson V et al |
| PF4V1 | 4 | 74718101 | rs872914-G | PCDH15 | | 0.39 | 0.14 | 1.40E-09 | ↑ | trans | Emilsson V et al |
| PF4V1 | 4 | 74718101 | rs872914-G | PSMD9 | | 0.39 | 0.14 | 1.41E-09 | ↑ | trans | Emilsson V et al |
| PF4V1 | 4 | 74718101 | rs872914-G | SULT1E1 | | 0.39 | 0.14 | 3.62E-09 | ↑ | trans | Emilsson V et al |
| PF4V1 | 4 | 74718101 | rs872914-G | NPPA | | 0.39 | 0.14 | 3.95E-09 | ↑ | trans | Emilsson V et al |
| PF4V1 | 4 | 74718101 | rs872914-G | ZPBP | | 0.39 | 0.13 | 5.93E-09 | ↑ | trans | Emilsson V et al |
| PF4V1 | 4 | 74718101 | rs872914-G | TRIL | | 0.39 | 0.13 | 6.96E-09 | ↑ | trans | Emilsson V et al |
| PF4V1 | 4 | 74733558 | rs3117603-T | CXCL1 | | 0.23 | 0.70 | 1.00E-62 | ↑ | cis | Sun BB et al |
| DEFA1 | 8 | 6878257 | rs4284061-T | DEFA1 | rs2075836 | 0.19 | 0.17 | 3.30E-12 | ↓ | cis | Emilsson V et al |
| IGH | 14 | 107211211 | rs2073668-G | ATP4B | rs751081288 | 0.93 | 0.15 | 3.20E-12 | ↓ | trans | Sun BB et al |
| IGH | 14 | 107141916 | rs10139058-C | ACP2 | | 0.87 | 0.21 | 1.20E-25 | ↓ | trans | Sun BB et al |
| IGH | 14 | 107167782 | rs3814922-G | STIM1 | | 0.86 | 0.16 | 2.60E-13 | ↓ | trans | Sun BB et al |
| IGH | 14 | 107170077 | rs11845244-T | DB119 | | 0.71 | 0.24 | 2.30E-20 | ↑ | trans | Sun BB et al |
| IGH | 14 | 107100923 | rs12882946-A | EPHB3 | | 0.70 | 0.21 | 1.80E-24 | ↓ | trans | Sun BB et al |
| FCAR | 19 | 55389032 | rs73065472-T | KI2LA | rs1865097 | 0.43 | 0.73 | 6.50E-163 | ↑ | trans | Sun BB et al |

**Table S24. Significant blood trans-eQTL effects of IgAN risk alleles.** Based on the blood eQTL meta-analysis of the eQTLGen Consortium.

| LOCUS | CHR | BP | SNP-risk allele | eGENE | CHR | BP | Z-score | P-value | FDR | Effect of IgAN risk allele |
|---|---|---|---|---|---|---|---|---|---|---|
| *TNFSF8/15* | 9 | 117643362 | rs13300483-T | *ALPK2* | 18 | 56222334 | 8.0631 | 7.44E-16 | 0 | ↑ |
| *TNFSF8/15* | 9 | 117643362 | rs13300483-T | *MSC* | 8 | 72755243 | 6.4489 | 1.13E-10 | 0 | ↑ |
| *TNFSF8/15* | 9 | 117643362 | rs13300483-T | *HDGFRP3* | 15 | 83830545 | 5.1624 | 2.44E-07 | 0.002354861 | ↑ |
| *TNFSF8/15* | 9 | 117643362 | rs13300483-T | *GZMK* | 5 | 54325239 | 5.1005 | 3.39E-07 | 0.00313024 | ↑ |
| *TNFSF8/15* | 9 | 117643362 | rs13300483-C | *SOCS2* | 12 | 93970426 | 4.6913 | 2.71E-06 | 0.019953826 | ↓ |
| *TNFSF8/15* | 9 | 117643362 | rs13300483-T | *EOMES* | 3 | 27760823 | 4.6907 | 2.72E-06 | 0.020014832 | ↑ |
| *TNFSF8/15* | 9 | 117643362 | rs13300483-T | *COCH* | 14 | 31353995 | 4.5529 | 5.29E-06 | 0.034565822 | ↑ |
| *TNFSF8/15* | 9 | 117643362 | rs13300483-T | *VCAM1* | 1 | 101194953 | 4.532 | 5.84E-06 | 0.037549294 | ↑ |
| *CARD9* | 9 | 139266496 | rs4077515-C | *MX1* | 21 | 42811686 | 10.1331 | 3.94E-24 | 0 | ↓ |
| *CARD9* | 9 | 139266496 | rs4077515-C | *IFIT1* | 10 | 91158024 | 9.0114 | 2.03E-19 | 0 | ↓ |
| *CARD9* | 9 | 139266496 | rs4077515-C | *HERC5* | 4 | 89402791 | 7.8329 | 4.76E-15 | 0 | ↓ |
| *CARD9* | 9 | 139266496 | rs4077515-C | *IFI44L* | 1 | 79097045 | 7.2547 | 4.02E-13 | 0 | ↓ |
| *CARD9* | 9 | 139266496 | rs4077515-C | *IFI44* | 1 | 79122622 | 7.1705 | 7.47E-13 | 0 | ↓ |
| *CARD9* | 9 | 139266496 | rs4077515-C | *RSAD2* | 2 | 7022153 | 6.1956 | 5.81E-10 | 2.07E-05 | ↓ |
| *CARD9* | 9 | 139266496 | rs4077515-C | *OAS3* | 12 | 113393605 | 5.4093 | 6.32E-08 | 0.00081666 | ↓ |
| *CARD9* | 9 | 139266496 | rs4077515-C | *IFIT2* | 10 | 91065372 | 5.167 | 2.38E-07 | 0.00226768 | ↓ |
| *CARD9* | 9 | 139266496 | rs4077515-C | *ISG15* | 1 | 949361 | 5.0655 | 4.07E-07 | 0.003626038 | ↓ |
| *CARD9* | 9 | 139266496 | rs4077515-C | *OAS2* | 12 | 113432864 | 4.9992 | 5.75E-07 | 0.004983771 | ↓ |
| *CARD9* | 9 | 139266496 | rs4077515-C | *XAF1* | 17 | 6668866 | 4.827 | 1.39E-06 | 0.010901042 | ↓ |
| *CARD9* | 9 | 139266496 | rs4077515-C | *MX2* | 21 | 42757593 | 4.6029 | 4.16E-06 | 0.028405265 | ↓ |
| *ITGAM/ITGAX* | 16 | 31357760 | rs11150612-G | *IGHG4* | 14 | 106091545 | 4.7301 | 2.24E-06 | 0.01693569 | ↓ |

**Table S25. Significant cis-eQTL effects (q<0.05) in tissues and cell types from GTEx.**

| LOCUS | SNP allele | CHR | BP | eGENE | Tissue | Slope | Slope s.e. | q-val | Top GWAS signal | R2 | Effect of IgAN risk allele |
|---|---|---|---|---|---|---|---|---|---|---|---|
| CFH | rs7542235-A | 1 | 196823613 | CFHR1 | Liver | 0.51 | 0.06 | 1.28E-30 | rs6677604 | 0.97 | ↑ |
| CFH | rs35253683-G | 1 | 196704039 | CFHR3 | Lung | 0.44 | 0.06 | 2.52E-23 | | 0.99 | ↑ |
| CFH | rs6677460-A | 1 | 196684575 | CFHR3 | Liver | 0.51 | 0.08 | 9.60E-21 | | 1.00 | ↑ |
| CFH | rs2064456-A | 1 | 196704443 | CFHR3 | Artery_Aorta | 0.39 | 0.08 | 3.16E-10 | | 0.99 | ↑ |
| CFH | rs60642321-A | 1 | 196822368 | CFHR3 | Adrenal_gland | 0.44 | 0.09 | 4.21E-10 | | 0.97 | ↑ |
| CFH | rs731557-C | 1 | 196705015 | CFHR3 | Esophagus_Mucosa | 0.37 | 0.08 | 1.37E-09 | | 0.99 | ↑ |
| CFH | rs2064456-A | 1 | 196704443 | CFHR3 | Heart_Atrial_Appendage | 0.37 | 0.08 | 3.94E-09 | | 0.99 | ↑ |
| CFH | rs71631868-T | 1 | 196815711 | CFHR3 | Testis | 0.41 | 0.09 | 8.15E-09 | | 0.93 | ↑ |
| CFH | rs2300429-G | 1 | 196675356 | CFHR3 | Artery_Coronary | 0.46 | 0.11 | 7.46E-08 | | 0.99 | ↑ |
| CFH | rs35253683-G | 1 | 196704039 | CFHR3 | Stomach | 0.41 | 0.09 | 1.13E-07 | | 0.99 | ↑ |
| CFH | rs67908756-T | 1 | 196821380 | CFHR1 | Testis | 0.38 | 0.09 | 2.40E-06 | | 0.97 | ↑ |
| CFH | rs6664877-C | 1 | 196684574 | CFHR3 | Vagina | 0.41 | 0.13 | 0.00158222 | | 0.91 | ↑ |
| CFH | rs35617250-C | 1 | 196679682 | CFHR3 | Ovary | 0.38 | 0.11 | 0.00358355 | | 1.00 | ↑ |
| CFH | rs71631868-T | 1 | 196815711 | CFHR1 | Brain_Hypothalamus | 0.41 | 0.13 | 0.00715966 | | 0.93 | ↑ |
| CFH | rs74696321-A | 1 | 196821120 | CFHR4 | Testis | 0.46 | 0.11 | 0.0266843 | | 0.89 | ↓ |
| CFH | rs6664877-C | 1 | 196684574 | CFHR1 | Colon_Sigmoid | 0.34 | 0.12 | 0.0271913 | | 0.91 | ↑ |
| TNFSF4 | rs4090391-T | 1 | 173147090 | TNFSF18 | Brain_Cerebellar_Hemisphere | 0.66 | 0.12 | 0.000343688 | rs4916312 | 1.00 | ↑ |
| TNFSF4 | rs7553711-T | 1 | 173131908 | RP3-395P12.2 | Brain_Cerebellum | 0.37 | 0.11 | 0.00129703 | | 1.00 | ↓ |
| TNFSF4 | rs7514229-T | 1 | 173154304 | TNFSF18 | Brain_Cerebellum | 0.56 | 0.11 | 0.00258856 | | 0.88 | ↑ |
| REL | rs397724941-T | 2 | 61082633 | REL | Testis | 0.15 | 0.04 | 0.00310488 | rs842638 | 0.99 | ↑ |
| REL | rs1432297-A | 2 | 61070652 | REL | Esophagus_Mucosa | 0.11 | 0.03 | 0.0283461 | | 0.97 | ↑ |
| TNFSF8/15 | rs4979462-T | 9 | 117567013 | TNFSF15 | Prostate | 0.41 | 0.14 | 0.0112187 | rs13300483 | 0.56 | ↑ |
| CARD9 | rs4078099-A | 9 | 139267533 | CARD9 | Whole_Blood | 0.30 | 0.03 | 6.33E-22 | rs4077515 | 1.00 | ↑ |
| CARD9 | rs10781499-A | 9 | 139266405 | CARD9 | Lung | 0.16 | 0.03 | 5.21E-05 | | 0.99 | ↑ |
| CARD9 | rs35670379-T | 9 | 139270606 | SEC16A | Liver | 0.18 | 0.04 | 0.00596325 | | 0.86 | ↓ |
| CARD9 | rs4498662-C | 9 | 139290251 | INPP5E | Lung | 0.12 | 0.03 | 0.020074 | | 0.84 | ↓ |
| CARD9 | rs11145763-C | 9 | 139263596 | SDCCAG3 | Muscle_Skeletal | 0.10 | 0.02 | 0.0275688 | | 0.98 | ↑ |
| RELA | rs10791824-A | 11 | 65559266 | OVOL1 | EBV_transformed_Lymphocytes | 0.43 | 0.10 | 1.25E-07 | rs10896045 | 0.59 | ↑ |
| RELA | rs12273982-C | 11 | 65535464 | AP5B1 | Nerve_Tibial | 0.20 | 0.05 | 0.0278015 | | 0.53 | ↓ |
| IGH | rs2105987-C | 14 | 107163289 | IGHV1-69 | Whole_Blood | 0.28 | 0.03 | 2.03E-26 | rs751081288 | 0.91 | ↓ |
| IGH | rs8004835-C | 14 | 107142378 | IGHV3-66 | Lung | 0.55 | 0.04 | 1.64E-22 | | 0.87 | ↑ |
| IGH | rs3944157-T | 14 | 107138303 | IGHV3-66 | Whole_Blood | 0.51 | 0.04 | 1.55E-21 | | 0.87 | ↑ |
| IGH | rs756583-T | 14 | 107136019 | IGHV3-66 | Colon_Transverse | 0.38 | 0.03 | 3.40E-20 | | 0.87 | ↑ |
| IGH | rs2097580-C | 14 | 107143461 | IGHV3-66 | Esophagus_Mucosa | 0.41 | 0.04 | 1.75E-19 | | 0.87 | ↑ |
| IGH | rs756583-T | 14 | 107136019 | IGHV3-66 | Spleen | 0.66 | 0.06 | 3.08E-15 | | 0.87 | ↑ |
| IGH | rs756583-T | 14 | 107136019 | IGHV3-66 | Stomach | 0.38 | 0.04 | 4.01E-13 | | 0.87 | ↑ |
| IGH | rs756583-T | 14 | 107136019 | IGHV3-66 | Adipose_visceral_Omentum | 0.44 | 0.06 | 8.15E-09 | | 0.87 | ↑ |
| IGH | rs756583-T | 14 | 107136019 | IGHV3-66 | Minor_Salivary_Gland | 0.71 | 0.07 | 9.32E-09 | | 0.87 | ↑ |
| IGH | rs11624912-G | 14 | 107129907 | IGHV3-66 | Liver | 0.62 | 0.07 | 2.52E-08 | | 0.91 | ↑ |
| IGH | rs8018138-G | 14 | 107153577 | IGHV1-69 | Esophagus_Mucosa | 0.17 | 0.03 | 1.11E-07 | | 0.88 | ↓ |
| IGH | rs55848268-A | 14 | 107207561 | IGHV3-66 | Thyroid | 0.26 | 0.04 | 1.97E-07 | | 0.96 | ↑ |
| IGH | rs8004895-T | 14 | 107193377 | IGHV1-69 | Colon_Transverse | 0.17 | 0.03 | 1.56E-05 | | 0.94 | ↓ |
| IGH | rs11627315-T | 14 | 107210420 | IGHV3-73 | Whole_Blood | 0.17 | 0.04 | 0.000634228 | | 0.96 | ↓ |
| IGH | rs10220646-T | 14 | 107128564 | IGHV1-69 | Minor_Salivary_Gland | 0.29 | 0.06 | 0.000738176 | | 0.91 | ↓ |
| IGH | rs3814917-T | 14 | 107167927 | IGHV4-59 | Adipose_visceral_Omentum | 0.31 | 0.06 | 0.000871219 | | 0.85 | ↑ |
| IGH | rs55848268-G | 14 | 107207561 | IGHV1-69 | Artery_Aorta | 0.25 | 0.06 | 0.00090114 | | 0.96 | ↓ |
| IGH | rs6576200-G | 14 | 107124719 | IGHV1-69 | Spleen | 0.26 | 0.07 | 0.00506229 | | 0.90 | ↓ |
| IGH | rs2006284-G | 14 | 107132201 | IGHV1-69 | Esophagus_Muscolaris | 0.24 | 0.07 | 0.0342393 | | 0.90 | ↓ |
| LIF | rs9614090-T | 22 | 30269907 | LIF | Muscle_Skeletal | 0.18 | 0.04 | 0.000851027 | rs48230748 | 0.90 | ↓ |
| LIF | rs5752989-G | 22 | 30365780 | RP4-539M6.20 | Testis | 0.27 | 0.08 | 0.0107755 | | 0.86 | ↓ |

**Table S26. Significant kidney compartment-specific eQTL effects of genome-wide significant IgAN risk loci.**

| LOCUS | CHR | BP | SNP-risk allele | GENE | Effect | s.e. | p-VALUE | Compartment | Effect of IgAN risk allele |
|---|---|---|---|---|---|---|---|---|---|
| CFH | 1 | 196686918 | rs6677604-G | *CFHR1* | 0.4082 | 0.1481 | 6.5067E-06 | Glomerulus | ↑ |
| CFH | 1 | 196686918 | rs6677604-G | *CFHR1* | 0.5004 | 0.1431 | 3.8659E-11 | Tubule | ↑ |
| RELA | 11 | 65555524 | rs10896045-A | *SNX32* | 0.4339 | 0.133 | 4.16E-08 | Glomerulus | ↑ |
| LIF | 22 | 30512478 | rs4823074-G | *NIPSNAP1* | 0.3662 | 0.1142 | 0.00000133 | Glomerulus | ↑ |

#### Table S27. Significant blood mQTL effects of genome-wide significant IgAN risk loci (Metabolomics GWAS Server)

| Locus | CHR | BP | SNP-risk allele | Metabolite | Effect | p-VALUE | top GWAS SNP | R2 | Effect of risk allele | Study |
|---|---|---|---|---|---|---|---|---|---|---|
| CFH | 1 | 196823613 | rs7542235-A | 1-stearoylglycerophosphoinositol:HWESASXX ratio | 0.06733 | 5.22E-09 | rs6677604 | 0.9533 | ↑ | Suhre et al |
| CFH | 1 | 196823613 | rs7542235-A | 1-linoleoylglycerophosphocholine:HWESASXX ratio | 0.05715 | 6.74E-08 | | 0.9533 | ↑ | Suhre et al |
| CFH | 1 | 196823613 | rs7542235-A | arachidonate (20:4n6):HWESASXX ratio | 0.05476 | 6.90E-08 | | 0.9533 | ↑ | Suhre et al |
| CFH | 1 | 196674714 | rs2019727-A | 1-stearoylglycerophosphoinositol:HWESASXX ratio | 0.06348 | 7.99E-08 | | 0.9108 | ↓ | Suhre et al |
| CFH | 1 | 196513694 | rs10922082-A | HWESASXX:pipecolate ratio | 0.049 | 2.23E-07 | | 0.6755 | ↓ | Suhre et al |
| CFH | 1 | 196674714 | rs2019727-A | arachidonate (20:4n6):HWESASXX ratio | 0.05364 | 2.39E-07 | | 0.9108 | ↓ | Suhre et al |
| CFH | 1 | 196674714 | rs2019727-T | X-11852:X-11858 ratio | 0.154 | 3.50E-07 | | 0.9108 | ↑ | Shin et al |
| CFH | 1 | 196674714 | rs2019727-A | 1-eicosadienoylglycerophosphocholine:HWESASXX ratio | 0.06188 | 3.69E-07 | | 0.9108 | ↓ | Suhre et al |
| CFH | 1 | 196674714 | rs2019727-A | 1-linoleoylglycerophosphocholine:HWESASXX ratio | 0.05506 | 3.71E-07 | | 0.9108 | ↓ | Suhre et al |
| CFH | 1 | 196603302 | rs12029571-A | X-12450:stachydrine ratio | 0.175 | 4.32E-07 | | 1 | ↑ | Shin et al |
| CFH | 1 | 196513694 | rs10922082-G | asparagine:HWESASXX ratio | 0.04621 | 4.34E-07 | | 0.6755 | ↑ | Suhre et al |
| CFH | 1 | 196569681 | rs16840224-T | HWESASXX:pipecolate ratio | 0.047 | 4.99E-07 | | 0.7059 | ↓ | Suhre et al |
| CFH | 1 | 196686918 | rs6677604-G | gamma-tocopherol:oleoylcarnitine ratio | 0.124 | 5.25E-07 | | 1 | ↑ | Suhre et al |
| CFH | 1 | 196825287 | rs7519758-T | HWESASXX | 0.043 | 5.29E-07 | | 0.9013 | ↓ | Shin et al |
| CFH | 1 | 196584468 | rs12061508-G | X-12450:stachydrine ratio | 0.177 | 5.80E-07 | | 0.9528 | ↑ | Shin et al |
| CFH | 1 | 196569681 | rs16840224-C | asparagine:HWESASXX ratio | 0.0448 | 6.98E-07 | | 0.7059 | ↑ | Suhre et al |
| CFH | 1 | 196584468 | rs12061508-A | X-12855:X-14662 ratio | 0.0593 | 7.65E-07 | | 0.9528 | ↓ | Shin et al |
| CFH | 1 | 196823613 | rs7542235-G | HWESASXX | 0.0421 | 7.76E-07 | | 0.9533 | ↓ | Suhre et al |
| CFH | 1 | 196686918 | rs6677604-A | X-11852:X-11858 ratio | 0.1103 | 8.50E-07 | | 1 | ↓ | Shin et al |
| CFH | 1 | 196632470 | rs6680396-G | X-12855:X-14662 ratio | 0.0584 | 8.66E-07 | | 0.9503 | ↓ | Shin et al |
| CFH | 1 | 196584321 | rs4657825-A | X-12855:X-14662 ratio | 0.0575 | 8.72E-07 | | 0.864 | ↓ | Shin et al |
| CFH | 1 | 196603302 | rs12029571-A | 5-alpha-pregnan-3beta,20alpha-disulfate:stachydrine ratio | 0.188 | 9.50E-07 | | 1 | ↑ | Shin et al |
| CFH | 1 | 196710916 | rs16840522-T | HWESASXX | 0.0433 | 1.82E-06 | | 0.981 | ↑ | Shin et al |
| CFH | 1 | 196674714 | rs2019727-T | HWESASXX | 0.0398 | 5.04E-06 | | 0.9108 | ↑ | Suhre et al |
| CFH | 1 | 196573505 | rs10754198-A | HWESASXX | 0.038 | 6.12E-06 | | 0.7116 | ↑ | Shin et al |
| CFH | 1 | 196513694 | rs10922082-A | HWESASXX | 0.0332 | 2.09E-05 | | 0.6755 | ↓ | Suhre et al |
| CFH | 1 | 196397775 | rs1538687-G | HWESASXX | 0.0318 | 5.23E-05 | | 0.5518 | ↓ | Suhre et al |
| CFH | 1 | 196387741 | rs12069983-A | HWESASXX | 0.0307 | 7.02E-05 | | 0.5513 | ↓ | Suhre et al |
| CFH | 1 | 196584321 | rs4657825-A | X-14662 | 0.0304 | 9.35E-05 | | 0.864 | ↓ | Shin et al |
| CFH | 1 | 196584468 | rs12061508-A | X-14662 | 0.0304 | 9.50E-05 | | 0.9528 | ↓ | Shin et al |
| CFH | 1 | 196405652 | rs7531611-T | HWESASXX | 0.0293 | 0.0001736 | | 0.5489 | ↓ | Suhre et al |
| CFH | 1 | 196686918 | rs6677604-G | gamma-tocopherol | 0.058 | 4.20E-04 | | 1 | ↑ | Suhre et al |
| CFH | 1 | 196702525 | rs2284664-A | stachydrine | 0.0876 | 0.0005332 | | 0.6466 | ↓ | Suhre et al |
| CFH | 1 | 196642233 | rs800292-T | HWESASXX | 0.0288 | 0.000645 | | 0.8085 | ↓ | Suhre et al |
| CFH | 1 | 196464754 | rs12065463-C | stachydrine | 0.0975 | 0.000731 | | 0.6597 | ↓ | Suhre et al |
| CFH | 1 | 196838060 | rs6657442-C | stachydrine | 0.0842 | 0.0007412 | | 0.5731 | ↓ | Suhre et al |
| CFH | 1 | 196387809 | rs12069990-T | HWESASXX | 0.0273 | 0.000746 | | 0.5513 | ↓ | Suhre et al |
| CD28 | 2 | 204584456 | rs4675360-A | myristoleate (14:1n5) | 0.0369 | 7.74E-05 | rs3769684 | 0.8431 | ↑ | Shin et al |
| CD28 | 2 | 204601910 | rs3181113-G | 5-oxoproline | 0.02233 | 0.0001551 | | 0.8056 | ↑ | Suhre et al |
| CD28 | 2 | 204582623 | rs4673259-T | 5-oxoproline | 0.02194 | 0.0002423 | | 0.8273 | ↑ | Suhre et al |
| CD28 | 2 | 204648661 | rs7599230-T | 5-oxoproline | 0.02156 | 0.0005041 | | 0.5072 | ↑ | Suhre et al |
| CD28 | 2 | 204601910 | rs3181113-G | gamma-glutamylglutamine | 0.02539 | 5.33E-04 | | 0.8056 | ↑ | Suhre et al |

**Table S28. Drug targets among genes encoded by significant GWAS loci.** PNH: paroxysmal nocturnal hemoglobinuria; AMD: age-related macular degeneration; AIHA: autoimmune hemolytic anemia; CAD: cold agglutinin disease; C3G: C3 glomerulopathy; DDD: Dense Deposit Disease, ABOi: ABO incompatible; MAb: monoclonal antibody; COPD: chronic obstructive pulmonary disease; HSC: hemopoietic stem cell; CKD: chronic kidney disease; ADPKD: autosomal dominant polycystic kidney disease; T2D: type 2 diabetes; T1D: type 1 diabetes; FSGS: focal segmental glomerulosclerosis; MN: membranous nephropathy; AAV: ANCA-associated vasculitis, GvHD: graft versus host disease. ^ candidate effector genes prioritized in Figure 5.

| Gene | Target | Drug Name | Disease | Study Phase | Mechanism of Action |
|---|---|---|---|---|---|
| ***CFH^*** | CD21/Factor H | TT-30 | PNH | Clinical Trial (Phase I) | links C3d-binding domain of human complement receptor 2 (CD21) with the complement regulatory domain of CFH, inhibiting complement cascade |
| | C3 | APL-2 | AMD | Clinical Trial (Phase III) | binds C3 and inhibits its cleavage into C3a and C3b, inhibiting complement cascade |
| | | APL-2 | PNH | Clinical Trial (Phase I) | |
| | | APL-2 | Glomerulopathies, AIHA, CAD | Clinical Trial (Phase II) | |
| | C3 | POT-4 | AMD | Clinical Trial (Phase II)* | binds C3 and inhibits its cleavage into C3a and C3b, inhibiting complement cascade |
| | C3 | AMY-101 | C3G, ABOi transplant, PNH, AMD, periodontitis** | Clinical Trial (Phase I) | binds C3 and inhibits its cleavage into C3a and C3b, inhibiting complement cascade |
| | C5 | Eculizumab | aHUS, PNH, C3GN, DDD | Approved | Binds and inactivates C5, inhibiting terminal complement pathway |
| | C5 | Ravilizumab | aHUS, PNH, C3GN, DDD | Approved | Binds and inactivates C5, inhibiting terminal complement pathway |
| | C5 | Cemdisiran | PNH, IgAN | Clinical Trials (Phase I/II) | |
| | C3/CR3 | Imprime PGG immunotherapeutic | Solid tumors, non-Hodgkin's lymphoma | Clinical Trials (Phase II/III) | binds to an alternate site on the neutrophil complement receptor 3 (CR3), priming neutrophils to become cytotoxic when binding to complement on tumor cells via CR3 |
| | Factor B | IONIS-FB-LRx | PNH, IgAN | Clinical Trials (Phase I/II) | Antisense Inhibitor of Complement Factor B |
| | Factor B | Iptacopan (LNP023) | PNH, C3GN, DD, aHUS, IgAN, MN | Clinical Trials (Phase II/III) | Oral inhibitor of Factor B |
| | Factor D | Danicopan, ALXN2050 | PNH, C3GN, DDD, MPGN | Clinical Trials (Phase I/II/III) | Oral inhibitors of Factor D |
| ***TNFSF4^*** | OX40L | RO4989991 (Oxelumab) | Allergic Asthma, allergis rhinitis | Clinical Trial (Phase II) | anti-OX40L mAb, inhibits OX40/OX40L signaling, inhibiting T cell activation*** |
| | OX40 | KHK4083/ISB-830 | Atopic dermatitis, Ulcerative colitis. lupus | Clinical Trial (Phase II/I) | anti-OX40 mAb, inhibits OX40/OX40L signaling, inhibiting T cell activation*** |
| | OX40 | RG7888 (Vonlerolizumab), MEDI0562 | Solid tumors | Clinical Trial (Phase I) | anti-OX40 mAb which acts as an agonist, activating T cell response and anti-tumor immunity |
| ***FASLG*** | CD95 (FAS) | DE-098 | Rheumatoid arthritis | Clinical Trial (Phase II)* | induces Fas antigen-mediated apoptosis |
| ***CD28^*** | CD28 | Belatacept | Kidney transplant rejection | Approved | Inhibitory activity on T lymphocytes |
| | CD80/CD86 Receptors | Abatacept | Heart transplant rejection | NDA filed | Inhibition of autoimmune T-Cell activation |
| | | | Rheumatoid arthritis | Approved | |
| | CD28/ICOS | ALPN-101 | Inflammatory Bowel Disease | Clinical Trial (Phase I) | Dual inhibitor of the CD28 and ICOS T cell costimulatory pathways. |
| ***CTLA4*** | CTLA4 | Ipilimumab | Solid tumors (metastatic melanoma) | Approved | anti-CTLA4 mAb, inhibits CTLA4 inhibitory functions and consequently activates cytotoxic T lymphocyte response against tumoral cells |
| ***REL^*** | C-Rel | IT-603/IT-901 | Graft Versus Host Disease, transplant Rejection | Pre-clinical | Not Available |
| ***CXCL8 (IL8)*** | IL-8 | ABX-IL8 | Melanoma, COPD | Clinical Trial (Phase II) | anti-IL-8 antibody, neutralizes IL-8-dependent human neutrophil activation |
| | | Troxipide | Gastric Ulcers, Gastritis | Approved | inhibit IL-8-induced migration of inflammatory cells, inhibiting neutrophil mediated inflammation and oxidative stress. |
| | IL-8 receptor B | Clotrimazole | Fungal infections | Approved | Suppression of IL-8 expression, inhibits intestinal inflammation |
| ***PF4 (CXCL4)*** | Heparin | Heparin | Thrombosis | Approved | Activation of Anti-thrombin III and inhibition of cascade coagulation (heparin binds to CXCL-PF4) |
| | CXCR4 | Plerixafor | Non Hodgkin's lymphoma, multiple myeloma before HSC transplantation | Approved | HSC mobilizer, inhibition of CXCR4 chemokine receptors on CD34+ cells, recruiting them in the peripheral blood for collection before HSC transplantation |
| ***CCR6^*** | CCR6 | PF-07054894 | Inflammatory Bowel Disease | Clinical Trial (Phase I) | CCR6 antagonist, prevents recruitment of CCR6+, IL17-producing cells to the affected tissues |
| ***LYN^*** | LYN | Bafetinib | Chronic lymphocytic leukemia, metastatic hormone refractory prostate cancer, glioma and brain metastasis | Clinical Trial (Phase II/I) | Inhibition of Lyn tyrosine kinase, decreases cellular proliferation and induce apoptosis, osteoclasts inhibitor |
| | SRC | Dasatinib | Chronic myelogenous leukemia | Approved | Src family tyrosine kinase inhibitor |
| | SRC | Bosutinib | Philadelphia chromosome-positive chronic myelogenous leukemia | Approved | Inhibition of Src and Abl tyrosine kinases, minimal inhibitory activity against c-KIT or platelet-derived growth factor receptor |
| ***TNFSF8 (CD30L)*** | CD30 (TNFRSF8) | Brentuximab vedotin | Hodgkin's lymphoma | Approved | anti-CD30 mAb, inhibits CD30/CD30L signaling inducing cell cycle arrest and apoptosis of the tumor cells |
| ***TNFSF15^ (TL1)*** | *TNFSF15* | PF-06480605, PR-200 | Inflammatory Bowel Disease | Clinical Trial (Phase II/I) | anti-TL1A mAb , it binds to TL1A, neutralizing TL1A-DR3 binding and signaling.. |
| ***PPIF*** | Cyclophilin D | Not Available | Not Available | Pre-clinical | NA (note: PPIF encodes cyclophilin D, which is suppressed by Cyclosporine A) |
| ***MAP3K11*** | MAP3K11 | URMC-099 | Not Available | Pre-clinical | Demonstrates anti-inflammatory and neuroprotective effects in mouse |
| | SYK | Fostamatinib | Immune Thrombocytopenic Purpura (ITP) | Approved | Syk kinase inhibitor, inhibits signal transduction by Fcγ receptors involved in the antibody-mediated destruction of platelets by immune cells in chronic ITP |
| ***RELA^ (p65)*** | Nrf2 pathway | Tecfidera | Multiple sclerosis | Approved | Up-regulates the Nuclear factor (erythroid-derived 2)-like 2 (Nrf2) pathway |
| | NF-kB | Bardoxolone methyl | IgAN, CKD, Alport syndrome, ADPKD, Pulmonary hypertension, Diabetic kidney disease, T2D | Clinical Trial (Phase III) | Inhibits the activity of nuclear factor kappa-B (NF-kB) activated by tumor necrosis factor (TNF) and other inflammatory agents |
| | | Bardoxolone methyl | T1D-associated CKD, FSGS | Clinical Trial (Phase II) | |
| | | Bardoxolone methyl | Obesity, solid tumors, lymphoid malignancies | Clinical Trial (Phase I) | |
| | NF-kB | Astaxanthin | Investigated for use/treatment in lymphoma (unspecified), multiple myeloma, and solid tumors. | Pre-clinical | Mediates anti-oxidant and anti-inflammatory actions |
| | NF-kB | SC-236 | cancer therapy, lower back pain, and inflammation | Pre-clinical | Suppresses the nuclear translocation of RelA/p65, inhibiting NF-kB signaling |
| ***KAT5*** | Histone acetyltransferase KAT5 | Coenzyme A | Hyperlipoproteinemia | Clinical Trial (Phase II/III) | Acts as a liposoluble antioxidant that can participate in redox reactions |
| ***PRSS8*** | protease, serine, 8 | B3C | Not Available | Pre-clinical | Not Available |
| ***FUS*** | Tumor suppressor candidate 2 | Fus-1 gene therapy | Non-small cell lung cancer | Clinical Trial (Phase I/II) | Expression of FUS1 protein is absent or reduced in the majority of lung cancers |
| ***KAT8*** | KAT8 | Naringenin | Hepatitis C, Cardiovascular diseases | Clinical Trial (Phase I) | Promotes carbohydrate metabolism, increases antioxidant defenses, scavenges reactive oxygen species, modulates immune system activity |
| ***TNFSF12*** | TWEAK (TNFSF12) | BIIB 023 | Rheumatoid arthritis, Lupus nephritis | Clinical Trial (Phase II)* | anti-TWEAK mAb, inhibits TWEAK/FN14 pathway, attenuating inflammation |
| ***TNFSF13^*** | TACI | Atacicept | Lymphoma; graft rejection; ITP; RA; Multiple sclerosis | Clinical Trial (Phase II/III) | Blocks binding of BAFF (BLYS) and APRIL to TACI, Inhibits B-cell development & survival, reduces autoimmune response |
| | | | IgA Nephropathy | Clinical Trial (Phase II) | |
| | APRIL | BION1301 | IgA Nephropathy | Clinical Trial (Phase II) | Specific anti-APRIL monoclonal antibody |
| | APRIL | VIS649 | IgA Nephropathy | Clinical Trial (Phase II) | Specific anti-APRIL monoclonal antibody |
| ***TNFRSF13B^*** | TACI | Atacicept | Lymphoma; graft rejection; ITP; RA, Multiple sclerosis | Clinical Trial (Phase II/III) | Blocks binding of BAFF (BLYS) and APRIL to TACI |
| | BAFF | Belimumab | Systemic lupus erythematosus, Lupus nephritis | Approved | Selectively binds to soluble B lymphocyte stimulator (BLyS), also known as BAFF, inhibiting its binding to B cells and their activation |
| | | | MN, AAV, COPD/Emphysema | Clinical Trial (Phase II) | |
| | | | kidney transplant, GvHD | Clinical Trial (Phase I) | |
| | BAFF | Blisibimod | Systemic lupus erythematosus, IgA nephropathy | Clinical Trial (Phase II-III) | Fusion protein BAFF binding domains with Fc receptor, inhibits BAFF |
| ***FCAR^*** | CD89 (FCAR) | MDX-214 | Cancer | Clinial Trial (Phase I/II)* | anti-CD89 mAb, it inhibits tumor cell growth by mediating antibody-dependent cellular cytotoxicity |
| ***LIF^*** | LIFR | Emfilermin (recombinant human LIF) | Infertility | Clinical Trial (Phase I/II)* | Facilitates embryonic development and implantation |
| ***NF2*** | NF2 gene promoter | Not Available | Not Available | Pre-clinical | Not Available |

* Clinical Trial has been discontinued; ** Potential target diseases; *** These molecules are not predicted to be effective in IgAN based on eQTL effects

**Table S29. Clinical correlations of genetic risk scores and individual risk alleles.** Individual risk loci and risk score models were tested for associations with clinical parameters at diagnosis. The association analysis was performed under additive genotype coding using logistic regression for binary outcomes, linear regression for continuous outcomes, and Cox proportional hazards model for survival outcomes. All genetic risk scores were standard normalized to mean 0 and variance of 1 before association testing, thus the depicted effect sizes are expressed per one standard deviation of the score. Significant associations after Bonferroni correction highlighted in bold.

| Locus | SNP | Risk Allele | Age at biopsy* (N=5,089) | | | eGFR at biopsy** (N=4,396) | | | Proteinuria at biopsy** (N=3,038) | | | Microhematuria** (N=2,627) | | | HTN** (N=6,547) | | | Gross Hematuria** (N=6,547) | | | Lifetime risk of kidney failure* (N=2,879, 913 events) | |
|---|---|---|---|---|---|---|---|---|---|---|---|---|---|---|---|---|---|---|---|---|---|---|
| | | | Beta | SE | P value | Beta | SE | P value | Beta | SE | P value | Beta | SE | P value | Beta | SE | P value | Beta | SE | P value | HR (CI 95%) | P value |
| FCRL | *rs849815* | A | 0.56 | 2.45 | 0.82 | -0.07 | 0.12 | 0.54 | 0.09 | 0.14 | 0.52 | 0.13 | 0.89 | 0.89 | -0.17 | 0.49 | 0.74 | 0.71 | 0.64 | 0.27 | 1.28 (0.52-3.11) | 0.59 |
| TNFSF4 | *rs4916312* | A | -1.81 | 2.51 | 0.47 | -0.52 | 0.13 | 5.0E-05 | 0.28 | 0.14 | 0.047 | 1.98 | 1.08 | 0.07 | 0.88 | 0.52 | 0.09 | -0.27 | 0.57 | 0.64 | 1.37 (0.67-2.82) | 0.39 |
| CFH | *rs12029571* | A | 0.72 | 2.52 | 0.77 | 0.04 | 0.12 | 0.74 | -0.28 | 0.15 | 0.06 | -1.19 | 1.03 | 0.25 | -0.14 | 0.51 | 0.78 | 0.59 | 0.62 | 0.34 | 0.71 (0.28-1.79) | 0.47 |
| CFH | *rs6677604* | G | -1.56 | 2.22 | 0.48 | 0.05 | 0.11 | 0.63 | -0.36 | 0.13 | 0.01 | 1.23 | 0.89 | 0.17 | 0.02 | 0.46 | 0.96 | 0.44 | 0.52 | 0.40 | 1.36 (0.69-2.71) | 0.38 |
| REL | *rs842638* | T | 1.01 | 1.99 | 0.61 | -0.02 | 0.10 | 0.81 | -0.16 | 0.11 | 0.16 | -0.52 | 0.81 | 0.52 | -0.15 | 0.41 | 0.70 | -0.39 | 0.46 | 0.40 | 1.08 (0.58-2.01) | 0.81 |
| CD28 | *rs3769684* | T | -3.93 | 1.93 | 0.04 | 0.08 | 0.09 | 0.37 | 0.10 | 0.11 | 0.37 | -0.61 | 0.73 | 0.40 | 0.22 | 0.39 | 0.56 | -0.31 | 0.51 | 0.54 | 1.08 (0.57-2.02) | 0.82 |
| PF4V1/CXCL8 | *rs6828610* | G | -0.47 | 2.48 | 0.85 | -0.03 | 0.12 | 0.82 | -0.27 | 0.14 | 0.06 | -0.40 | 0.97 | 0.68 | 0.46 | 0.51 | 0.37 | -0.95 | 0.64 | 0.14 | 1.27 (0.52-3.10) | 0.60 |
| IRF4 | *rs12201499* | C | -1.09 | 2.08 | 0.60 | 0.03 | 0.10 | 0.77 | -0.04 | 0.12 | 0.72 | 0.50 | 0.79 | 0.53 | 0.33 | 0.43 | 0.45 | 1.07 | 0.53 | 0.046 | 1.06 (0.50-2.23) | 0.89 |
| LY86 | *rs12530084* | C | 0.36 | 2.26 | 0.87 | -0.13 | 0.11 | 0.25 | 0.11 | 0.13 | 0.39 | 0.35 | 0.89 | 0.69 | 0.31 | 0.46 | 0.51 | 0.06 | 0.57 | 0.91 | 1.16 (0.52-2.60) | 0.72 |
| HLA | *rs9268557* | C | 0.37 | 1.35 | 0.79 | 0.00 | 0.07 | 0.97 | -0.03 | 0.08 | 0.72 | -0.14 | 0.52 | 0.79 | 0.00 | 0.27 | 0.99 | 0.39 | 0.34 | 0.25 | 1.06 (0.68-1.66) | 0.79 |
| HLA | *rs9272105* | A | -2.17 | 1.23 | 0.08 | -0.01 | 0.06 | 0.88 | -0.03 | 0.07 | 0.65 | -0.76 | 0.46 | 0.10 | 0.43 | 0.25 | 0.09 | -0.29 | 0.30 | 0.33 | 0.97 (0.63-1.48) | 0.88 |
| HLA | *rs9275355* | C | -4.32 | 1.41 | 0.002 | -0.06 | 0.07 | 0.40 | -0.01 | 0.09 | 0.89 | 0.37 | 0.78 | 0.64 | -0.17 | 0.29 | 0.57 | -0.01 | 0.30 | 0.97 | 1.32 (0.87-2.00) | 0.20 |
| HLA | *rs9275596* | T | -2.70 | 1.29 | 0.04 | 0.04 | 0.06 | 0.51 | -0.05 | 0.07 | 0.53 | 0.25 | 0.49 | 0.61 | 0.12 | 0.27 | 0.65 | 0.08 | 0.32 | 0.81 | 0.98 (0.66-1.44) | 0.90 |
| HLA | *rs3128927* | C | -3.85 | 1.71 | 0.02 | 0.08 | 0.09 | 0.32 | 0.05 | 0.10 | 0.65 | 1.79 | 0.65 | 0.01 | -0.58 | 0.35 | 0.10 | 0.20 | 0.41 | 0.62 | 1.04 (0.59-1.84) | 0.90 |
| DEFA | *rs2075836* | T | -3.14 | 1.67 | 0.06 | -0.02 | 0.08 | 0.84 | 0.06 | 0.10 | 0.57 | 0.62 | 0.64 | 0.33 | -0.16 | 0.33 | 0.64 | -0.08 | 0.41 | 0.84 | 1.14 (0.62-2.07) | 0.67 |
| LYN | *rs75413466* | A | 1.46 | 1.82 | 0.42 | -0.12 | 0.09 | 0.19 | 0.01 | 0.12 | 0.91 | 0.56 | 0.87 | 0.52 | 0.40 | 0.37 | 0.28 | 0.13 | 0.43 | 0.76 | 1.14 (0.60-2.16) | 0.68 |
| ANXA13 | *rs34354351* | T | -0.73 | 2.15 | 0.73 | -0.04 | 0.10 | 0.71 | -0.11 | 0.12 | 0.36 | 1.78 | 0.81 | 0.03 | -0.15 | 0.43 | 0.73 | 0.88 | 0.55 | 0.11 | 0.77 (0.35-1.71) | 0.52 |
| TNFSF15/TNFSF8 | *rs13300483* | T | 1.05 | 2.29 | 0.65 | -0.02 | 0.11 | 0.83 | -0.14 | 0.13 | 0.30 | 0.62 | 0.95 | 0.51 | -0.06 | 0.47 | 0.90 | 0.86 | 0.55 | 0.12 | 1.12 (0.50-2.52) | 0.78 |
| CARD9 | *rs4077515* | T | -3.85 | 2.26 | 0.09 | 0.07 | 0.11 | 0.50 | -0.01 | 0.13 | 0.94 | -1.61 | 0.84 | 0.055 | 0.42 | 0.45 | 0.35 | 0.64 | 0.59 | 0.28 | 0.77 (0.33-1.81) | 0.56 |
| REEP3 | *rs57917667* | G | 3.42 | 2.29 | 0.13 | 0.06 | 0.11 | 0.59 | 0.15 | 0.13 | 0.26 | 0.01 | 0.88 | 0.995 | -0.59 | 0.45 | 0.19 | 0.24 | 0.61 | 0.70 | 0.91 (0.33-2.57) | 0.87 |
| ZMIZ1 | *rs1108618* | A | -0.28 | 2.06 | 0.89 | -0.22 | 0.10 | 0.03 | -0.04 | 0.12 | 0.76 | 1.17 | 0.82 | 0.15 | 0.21 | 0.41 | 0.61 | -0.02 | 0.50 | 0.96 | 1.39 (0.66-2.94) | 0.38 |
| RELA | *rs10896045* | A | 0.30 | 1.61 | 0.85 | -0.16 | 0.08 | 0.045 | 0.16 | 0.09 | 0.08 | 0.30 | 0.65 | 0.64 | 0.16 | 0.33 | 0.62 | 0.00 | 0.39 | 0.99 | 1.20 (0.69-2.08) | 0.52 |
| ETS1 | *rs7121743* | C | -2.98 | 2.42 | 0.22 | -0.19 | 0.12 | 0.11 | 0.03 | 0.14 | 0.83 | 0.50 | 0.97 | 0.61 | 0.79 | 0.49 | 0.11 | 0.85 | 0.61 | 0.16 | 1.47 (0.65-3.32) | 0.36 |
| IGH | *rs751081288* | A | 1.07 | 1.76 | 0.54 | -0.04 | 0.09 | 0.67 | 0.12 | 0.10 | 0.21 | 1.31 | 0.67 | 0.049 | -0.43 | 0.36 | 0.23 | 0.09 | 0.41 | 0.83 | 0.88 (0.48-1.62) | 0.69 |
| ITGAM | *rs11150612* | A | 0.61 | 1.84 | 0.74 | -0.08 | 0.09 | 0.36 | -0.03 | 0.11 | 0.76 | 0.84 | 0.73 | 0.25 | 0.38 | 0.38 | 0.31 | 0.49 | 0.45 | 0.28 | 1.11 (0.63-1.94) | 0.72 |
| IRF8 | *rs1879210* | T | 2.98 | 2.32 | 0.20 | -0.07 | 0.12 | 0.52 | 0.17 | 0.13 | 0.20 | -0.24 | 0.95 | 0.80 | -0.37 | 0.48 | 0.43 | 0.52 | 0.55 | 0.35 | 1.02 (0.49-2.15) | 0.95 |
| TNFSF13 | *rs3803800* | A | -1.34 | 2.05 | 0.51 | -0.09 | 0.10 | 0.37 | 0.00 | 0.12 | 0.99 | 1.22 | 0.82 | 0.14 | 0.55 | 0.42 | 0.19 | -0.53 | 0.50 | 0.29 | 1.22 (0.58-2.57) | 0.61 |
| TNFRSF13B | *rs57382045* | A | -4.12 | 2.03 | 0.04 | -0.19 | 0.10 | 0.06 | 0.00 | 0.12 | 0.97 | 1.41 | 0.84 | 0.09 | 0.20 | 0.41 | 0.62 | 0.49 | 0.51 | 0.34 | 1.53 (0.71-3.30) | 0.28 |
| FCAR | *rs1865097* | A | -1.75 | 2.35 | 0.46 | 0.24 | 0.12 | 0.04 | 0.32 | 0.14 | 0.02 | 1.20 | 0.95 | 0.21 | -0.04 | 0.48 | 0.93 | -0.80 | 0.57 | 0.16 | 0.79 (0.34-1.82) | 0.58 |
| HORMAD2/LIF | *rs4823074* | G | -2.91 | 1.81 | 0.11 | 0.00 | 0.09 | 0.96 | 0.07 | 0.10 | 0.47 | -0.06 | 0.71 | 0.93 | 0.16 | 0.37 | 0.66 | 0.00 | 0.44 | 0.995 | 1.44 (0.80-2.62) | 0.23 |
| 15-SNP Genetic Risk Score | | | **-0.03** | **0.01** | **9.7E-05** | 0.00 | 0.00 | 0.80 | 0.00 | 0.00 | 0.15 | 0.00 | 0.00 | 0.45 | 0.00 | 0.00 | 0.26 | 0.00 | 0.00 | 0.06 | 1.00 (0.99-1.00) | 0.37 |
| 30-SNP Genetic Risk Score | | | **-0.81** | **0.19** | **2.6E-05** | -0.02 | 0.01 | 3.8E-02 | 0.00 | 0.01 | 0.95 | 0.21 | 0.07 | 6.0E-03 | 0.05 | 0.04 | 0.24 | 0.07 | 0.05 | 0.17 | **1.12 (1.05-1.20)** | **6.4E-04** |
| 77-SNP Genetic Risk Score | | | **-0.82** | **0.19** | **2.3E-05** | -0.02 | 0.01 | 4.9E-02 | 0.00 | 0.01 | 0.78 | 0.18 | 0.08 | 2.0E-02 | 0.06 | 0.04 | 0.10 | 0.09 | 0.05 | 0.053 | **1.15 (1.08-1.22)** | **2.5E-05** |
| Genome-wide Polygenic Score | | | **-0.81** | **0.20** | **5.5E-05** | -0.02 | 0.01 | 0.12 | 0.00 | 0.01 | 0.77 | 0.09 | 0.08 | 0.26 | -0.03 | 0.04 | 0.49 | 0.08 | 0.05 | 0.07 | **1.17 (1.09-1.24)** | **3.3E-06** |

* Adjusted for Sex, Site, and Race/Ethnicity; ** Adjusted for Age, Sex, Site, and Race/Ethnicity

**Table S30. Meta-PheWAS of the genome-wide polygenic risk score (GPS) for IgAN with and without HLA.** Only the top associations meeting the Bonferroni-corrected significance level in either analysis are listed; the phenotypes significant in both analyses are highlighted in grey. Before association testing, GPS with and without HLA were standard normalized (mean 0 and variance of 1) thus the depicted effect sizes are expressed per one standard deviation of a GPS. OR: odds ratio; SE: standard error; P: *P*-value.

| Phenotype | Description | Group | GPS with HLA | | | GPS without HLA | | | No. of cases | No. of controls |
|---|---|---|---|---|---|---|---|---|---|---|
| | | | OR | SE | P | OR | SE | P | | |
| 557.1 | Celiac disease | digestive | 0.601 | 0.020 | 4.16E-148 | 0.892 | 0.020 | 1.89E-08 | 2770 | 388104 |
| 714 | Rheumatoid arthritis and other inflammatory polyarthropathies | musculoskeletal | 1.147 | 0.010 | 1.10E-39 | 1.061 | 0.011 | 9.50E-08 | 10652 | 465415 |
| 714.1 | Rheumatoid arthritis | musculoskeletal | 1.155 | 0.011 | 2.82E-39 | 1.067 | 0.012 | 2.94E-08 | 9365 | 465415 |
| 335 | Multiple sclerosis | neurological | 0.814 | 0.022 | 2.81E-21 | 0.959 | 0.023 | 6.77E-02 | 2309 | 463423 |
| 593 | Hematuria | genitourinary | 1.064 | 0.007 | 7.26E-21 | 1.038 | 0.007 | 9.32E-08 | 27491 | 442554 |
| 244.4 | Hypothyroidism NOS | endocrine/metabolic | 1.053 | 0.006 | 2.61E-16 | 1.034 | 0.007 | 6.48E-07 | 31035 | 463751 |
| 244 | Hypothyroidism | endocrine/metabolic | 1.050 | 0.006 | 1.99E-15 | 1.032 | 0.007 | 1.10E-06 | 32836 | 463751 |
| 477 | Epistaxis or throat hemorrhage | respiratory | 1.092 | 0.015 | 2.55E-09 | 1.070 | 0.016 | 1.30E-05 | 5277 | 446984 |
| 695.4 | Lupus (localized and systemic) | dermatologic | 0.879 | 0.025 | 2.13E-07 | 1.001 | 0.027 | 9.57E-01 | 2000 | 474880 |
| 250.1 | Type 1 diabetes | endocrine/metabolic | 1.061 | 0.012 | 1.48E-06 | 1.041 | 0.013 | 2.26E-03 | 7915 | 450116 |
| 495 | Asthma | respiratory | 1.024 | 0.005 | 1.50E-06 | 1.026 | 0.005 | 5.99E-07 | 51118 | 436029 |
| 695.42 | Systemic lupus erythematosus | dermatologic | 0.883 | 0.026 | 2.10E-06 | 1.005 | 0.029 | 8.64E-01 | 1788 | 474880 |
| 717 | Polymyalgia Rheumatica | musculoskeletal | 1.095 | 0.020 | 4.02E-06 | 1.028 | 0.021 | 1.84E-01 | 2822 | 510063 |
| 250 | Diabetes mellitus | endocrine/metabolic | 1.025 | 0.005 | 4.41E-06 | 1.024 | 0.006 | 2.45E-05 | 48837 | 450116 |
| 695.41 | Cutaneous lupus erythematosus | dermatologic | 0.814 | 0.045 | 5.43E-06 | 0.923 | 0.050 | 1.08E-01 | 584 | 474880 |
| 401.1 | Essential hypertension | circulatory system | 1.016 | 0.003 | 8.37E-06 | 1.009 | 0.004 | 1.20E-02 | 149236 | 354167 |
| 401 | Hypertension | circulatory system | 1.015 | 0.003 | 9.96E-06 | 1.009 | 0.004 | 1.42E-02 | 150452 | 354167 |
| 697 | Sarcoidosis | dermatologic | 0.887 | 0.028 | 2.10E-05 | 0.946 | 0.030 | 6.54E-02 | 1501 | 477327 |

### SUPPLEMENTAL NOTES

#### Discovery Cohorts

##### Italian, French, British, US, and Beijing-I Cohorts:

The composition, genome-wide genotyping, and genotype quality control (QC) of these cohorts have been published previously[1]. As part of this study, the genotype data was re-imputed using the latest ancestry-matched reference panels of 1000 Genome Project (Phase 3). The numbers of common and high-quality imputed markers ($R^2$>0.8, MAF>0.01) used in downstream analyses were as follows: 6,898,974 (French cohort), 7,058,018 (Italian cohort), 7,241,153 (US cohort), 4,344,488 (UK cohort) and 5,979,671 (Beijing-I cohort).

##### Turkish Cohort:

This cohort consists of 525 individuals (133 biopsy-diagnosed cases and 392 healthy controls) recruited at the Istanbul Faculty of Medicine, Istanbul University, Istanbul, Turkey. All cases had a histology-proven diagnosis of IgAN. The genotyping was performed using the Illumina Multi-ethnic Global Ancestry Array (MEGA) at the University of Michigan genotyping facility. Genotype calls were performed in Illumina Genome Studio software. All SNPs were called on the forward strand. Standard quality filters included per-sample genotype call-rates >95%, per-SNP genotype call rates >95% and MAF >0.01. Controls were also filtered by HWE test p-value $1x10^{-4}$. The duplicated and cryptic relatedness was assessed using the software KING software (http://people.virginia.edu/~wc9c/KING/). We excluded individuals based on a pairwise kinship coefficient >0.354 (third-degree relatives). Sex of each individual was imputed based on the analysis of sex chromosome markers and individuals with mismatched gender were excluded. Autosomal heterozygosity was calculated using KING and individuals who deviated ±3 SD from the sample's heterozygosity rate mean were excluded. Ancestry was evaluated using Principal Component Analysis (PCA)[5], we selected a subset of 126,864 high quality and independent (r2 <0.05) markers. PCA produced four significant PCs and adequate matching between cases and controls within the cohort. After QC, the final dataset was comprised of 439 individuals (115 cases and 324 controls) and 715,590 makers genotyped with an overall genotyping rate of 99.9%. We carried out imputation analysis as described below. A total of 7,543,852 high-quality common markers were imputed and used in downstream analysis.

##### European Cohort I:

All individuals were self-reported White/Caucasians of European ancestry. The following institutions contributed to this cohort (only centers contributing >10 cases to this cohort are listed): Columbia University, New York, USA; University Hospitals in Leuven, Belgium; University of Aachen, Germany; Jean Monnet University, Saint Etienne, France; Poznan, Warsaw, Wroclaw, Gdansk, and Bialystok Universities of Medical Sciences in Poland; Charles University, Prague, Czech Republic; Karolinska and Danderyd University Hospitals, Stockholm, Sweden; Zagreb Medical School, Zagreb, Croatia; University of Utah, Salt Lake City, USA; The Aristotle University of Thessaloniki, Greece; University of Turin, Italy; Regina Margherita Children's Hospital in Turin, Italy; Universities of Messina, Italy; University of Pavia, Italy, University of Modena and Reggio Emilia, Italy; Gaslini Institute, Genoa, Italy; Nephrology and Dialysis A.R.N.A.S. Civico and Benfratellio, Palermo, Italy; VU University Medical Center, Amsterdam, the Netherlands; and the Hospital Universitario Puerta del Hierro Majadahonda, Spain. All cases were ascertained based on a biopsy-proven diagnosis of IgAN. In total, 1,072 cases and 1,536 ancestry and geography-matched controls were genotyped using the Illumina Multi-ethnic Global Ancestry Array (MEGA) at the University of Michigan genotyping facility. The analysis of intensity clusters and genotype calls were performed using the Illumina Genome Studio software and all SNPs were called on the forward strand. The same standard QC as in the above cohorts were performed. For PCA, we selected a subset of 122,697 high-quality independent markers, resulting in seven significant PCs. The PCA identified and excluded six ancestry outliers, resulting in a total of 920 cases and 1,536 controls. After imputation a total of 7,867,380 high-quality markers were used in association analysis.

##### European Cohort II:

This cohort consisted of 976 individuals (503 biopsy-diagnosed cases and 473 controls) recruited among self-reported White/Caucasian individuals of European ancestry. The healthy controls were recruited from the same geographic regions as the cases. This cohort was built based on cases and controls recruited by the following institutions (only the centers that contributed >10 cases to the cohort are listed): Columbia University, New York, USA; University of Alabama at Birmingham, USA; University of Toronto, Canada; Karolinska and Danderyd University Hospitals, Stockholm, Sweden; Charles University, Czech Republic; University of Brescia, Italy; University of Milan, Italy; University of Verona, Italy; Poznan, Warsaw, and Gdansk Universities of Medical Sciences, Poland; Bern University,

Switzerland; University of Istanbul, Turkey. The genotyping was performed at the University of Michigan genotyping facility using the Illumina Multi-ethnic Global Ancestry Array (MEGA). Genotype calls were performed in Illumina Genome Studio software. All SNPs were called on the forward strand. The same standard genotype QC was performed as above. For PCA, we selected a subset of 126,479 high-quality independent makers. The PCA of the final dataset resulted in five significant PCs and demonstrated minimal population stratification and a successful matching between cases and controls within the cohort. In total 781,848 SNPs passed all the QC filters in the remainder 754 individuals (397 cases and 357 controls) with an overall genotyping rate of 99.9%. After imputation using European samples from 1000 Genome Project as a reference, a total of 7,718,643 high-quality markers were used in downstream analyses.

**Argentinian Cohort:**

This cohort was recruited at the University of Buenos Aires, Buenos Aires, Argentina, and consisted of 198 individuals (101 biopsy-diagnosed cases and 97 healthy controls) all self-reported Argentinian Whites. The genotyping was performed at the University of Michigan genotyping facility, using the Illumina Multi-ethnic Global Ancestry Array (MEGA). The same genotype processing steps and standard QC analyses were performed as above. The PCA based on 125,264 independent markers demonstrated excellent genetic matching between cases and controls, resulting in four significant PCs of ancestry. After QC, the final dataset consisted of 194 individuals (100 cases and 94 controls) and 750,842 makers genotyped with an overall genotyping rate of 99.9%. Imputation was carried out as described below. A total of 7,739,980 high-quality imputed markers were used in downstream analysis.

**Japanese Cohort:**

This cohort consisted of 807 individuals (427 biopsy-diagnosed cases and 380 controls) recruited among self-reported Japanese. The participants were recruited by the Division of Nephrology, Juntendo University Faculty of Medicine in Tokyo, Japan as well as by the Division of Nephrology in Niigata University Graduate School of Medical and Dental Science, Niigata, Japan. The DNA was genotyped using the Illumina Multi-ethnic Global Ancestry Array (MEGA). The same genotype processing steps and standard QC analyses were performed as above. The PCA using an independent set of high-quality markers resulted in two significant PCs and demonstrated minimal population stratification and a successful matching between cases and controls within the cohort. In total 590,245 SNPs passed all the QC filters in the remainder 776 individuals (414 cases and 362 controls) with an overall genotyping rate of 99.9%. After imputation a total of 6,027,358 high-quality markers were included in downstream analysis.

**Shanghai Cohort:**

This cohort consisted of 1,413 individuals (690 biopsy-diagnosed cases and 723 controls) all self-reported as Han Chinese from the Shanghai area recruited from the clinics of the Department of Nephrology of Ruijin Hospital, Shanghai Jiao Tong University School of Medicine, Shanghai, China. The genotyping was performed using the Illumina Multi-ethnic Global Ancestry Array (MEGA). The same genotype processing steps and standard QC analyses were performed as above. The PCA using high quality independent markers demonstrated excellent genetic mismatch between cases and controls, resulting in three significant PCs of ancestry. After QC, the final dataset consisted of 1,345 individuals (656 cases and 689 controls) and 593,475 markers genotyped with an overall genotyping rate of 99.9%. Imputation was carried out as described below. A total of 6,056,327 high-quality imputed markers were used in downstream analysis.

**Korean Cohort:**

The cases and controls were recruited from Seoul National University Hospital, Seoul, South Korea. and consisted of 1,463 individuals (750 biopsy-diagnosed cases and 713 healthy controls) all self-identified as Korean. The genotyping was performed using the Illumina Multi-ethnic Global Ancestry Array (MEGA). Genotype calls were performed in Illumina Genome Studio software. All SNPs were called on the forward strand. Same standard QC as in the above cohorts was performed. No individual was excluded based on PCA, heterozygosity, or gender mismatch. The final PCA demonstrated excellent genetic matching between cases and controls, resulting in only one significant PC of ancestry. After QC, the final dataset consisted of 1,443 individuals (735 cases and 708 controls) and 598,716 makers genotyped with an overall genotyping rate of 99.9%. Imputation was carried out as described below. A total of 5,942,775 high-quality imputed markers were used in downstream analysis.

**Beijing Cohort II:**

This cohort was recruited by the Renal Division in the Department of Medicine of Peking University First Hospital, Beijing, China. The cohort consisted of 4,623 individuals (500 biopsy-proven diagnosed cases and 4,123 controls), all self-reported as of Han Chinese ancestry. The genotyping was performed using the Illumina Infinium OmniZhongHua-8 v1.3 array. Genotype calls were performed in Illumina Genome Studio software. All SNPs were called on the forward strand. Same standard QC as in the above cohorts was performed. PCA on a subset of 46,958 high quality independent makers resulted in two significant PCs and demonstrated minimal population stratification and a successful matching between cases and controls within the cohort. In total 727,733 SNPs passed all the QC filters in the remainder 4,151 individuals (479 cases and 3,672 controls) with an overall genotyping rate of 99.9%. After imputation using 1000 Genomes Phase 3 v5 as reference panel, a total of 7,797,150 high-quality markers were used in downstream analysis.

**Beijing Cohort III:**

This cohort was recruited by the Renal Division in the Department of Medicine of Peking University First Hospital, Beijing, China. The cohort consisted of 3,152 individuals (1,230 biopsy-proven diagnosed cases and 1,922 controls), all self-reported as Han Chinese. The genotyping was performed using the Illumina Infinium Global Screening Array-24 v1.0 (GSA) BeadChip. Genotype calls were performed in Illumina Genome Studio software. All SNPs were called on the forward strand. Same standard QC as in the above cohorts was performed. For PCA, we selected a subset of 59,525 high quality independent makers. The PCA of the final dataset resulted in two significant PCs and demonstrated minimal population stratification and a successful matching between cases and controls within the cohort. In total 362,250 SNPs passed all the QC filters in the remainder 2,352 individuals (1,112 cases and 1,240 controls) with an overall genotyping rate of 99.9%. After imputation using 1000 Genomes Phase 3 v5 as reference panel, a total of 6,019,668 high quality markers were used in downstream analysis.

**Immunochip Cohorts:**

The Immunochip Consortium was formed to design a custom Illumina Infinium array that leveraged the genetic overlap of susceptibility loci identified across a range of autoimmune diseases. Briefly, the chip contains 196,524 SNPs designed to perform both replication of suggestive findings from autoimmune and inflammatory disease GWAS and fine-mapping of the established loci for autoimmune diseases[6,7]. Using this chip, we generated high-quality genotypes for 2,201 cases with a kidney biopsy diagnosis of IgAN referred for genetic studies across our network of collaborating European nephrology centers, including in Germany, France, Italy, Poland, Croatia, Hungary, Czech Republic, Turkey, and the UK. For controls, we used a total of 11,038 previously genotyped European population controls previously genotyped by the Immunochip consortium[6]. To avoid any potential genotype artifacts, genotype calls were performed jointly for all cases and controls from primary idat files in a single Illumina Genome Studio project. All SNPs were called on the forward strand. The genotype QC filters included per-sample genotype call-rates >95%, per-SNP genotype call-rates >98%, and MAF >0.01. We have also performed cryptic relatedness analysis based on genotype data and excluded any duplicate case and control within the Immunochip cohort and across all other European cohorts used in this study. Based on iterative PCA and clustering analyses, we subdivided our case-control cohorts into three separate cohorts broadly mapping to North Europe (NE), South Europe (SE) and Central-West Europe (CWE). The controls were matched genetically to the cases, and ancestry outliers were eliminated. The final PCA using 20,620, 20,863 and 20,505 high-quality independent markers for NE, SE and CWE cohorts, respectively, demonstrated excellent genetic matching between cases and controls, resulting in three significant PCs for the NE cohort, five significant PCs for the SE cohort and seven significant PCs for the CWE cohort. After final QC and removal of outliers, the three analysis datasets consisted of the following: the NE cohort with 7,624 individuals (527 cases and 7,097 controls) and 128,307 genotyped markers with overall genotyping rate of 99.9%; the SE cohort with 3,219 individuals (514 cases and 2,705 controls) and 127,837 genotyped markers with overall genotyping rate of 99.9%; and the CWE cohort of 2,202 individuals (966 cases and 1,236 controls) and 128,470 genotyped markers with overall genotyping rate of 99.8%.

**GCKD Testing Cohort**

This cohort was used for independent testing the performance of genome-wide polygenic score (GPS) optimized based on the discovery GWAS. The GCKD Study has previously been described in detail[3,4]. For GPS testing, we constructed a GCKD kidney-biopsy sub-cohort composed on 314 chronic kidney disease (CKD) patients with a biopsy-diagnosed IgAN cases and 663 patients with a biopsy-diagnosis of non-IgAN CKD to serve as controls. All participants were genotyped using the Illumina Omni2.5Exome array and imputed using the HRC version r1.1 reference panel. The best-performing GPS based on the discovery study explained 7.3% of variance in IgAN risk ($P$=3.1x$10^{-12}$) with AUROC of 0.65 (95% CI: 0.61-0.68).
